# Photobiomodulation for Cognitive Dysfunction (Brain Fog) in Post-COVID-19 Condition: A Randomized Sham-Controlled Pilot Trial

**DOI:** 10.1101/2025.07.24.25331961

**Authors:** Lew Lim, Nazanin Hosseinkhah, Mark Van Buskirk, Kevin Oei, Andrea Berk, Abhiram Pushparaj, Janine Liburd, Zara Abbaspour, Jonathan Rubine, David Jackson, Reza Zomorrodi

**Affiliations:** Vielight Inc., 471 Jarvis St., Toronto, ON M4Y 2G8, Canada; Vielight Inc., Toronto, ON, Canada; Data Reduction LLC, Chester, NJ, USA; Ascada Research, Fullerton, CA, USA; Andrea Berk Consulting & Management Services, Vaughan, ON, Canada; +ROI Regulatory Advisory, Grimsby, ON, Canada; MKR Clinical Research Consultants, Boynton Beach, FL, USA; Ivy Brain Tumor Center, Phoenix, AZ, USA; Centre for Addiction and Mental Health, University of Toronto, Toronto, ON, Canada

## Abstract

Post-COVID-19 condition (PCC) affects millions globally, with cognitive dysfunction (“brain fog”) impairing daily functioning in up to 88% of patients. No effective treatments exist for PCC-related cognitive impairment. We conducted a randomized, double-blind, sham-controlled pilot clinical trial to evaluate the efficacy of home-based photobiomodulation (PBM) using the Vielight Neuro RX Gamma device in 43 adults with PCC. Participants received 8 weeks of daily 20-minute PBM or sham treatment, targeting the default mode network. The primary outcome was change in cognitive performance (Creyos battery) at Day 56. Active PBM showed greater improvement in composite cognitive scores (p=0.088), with significant gains in participants under 45 years (p=0.028). Attention tasks improved consistently across groups. PBM was safe, with high compliance and no serious adverse events. These findings suggest PBM’s potential as a non-invasive intervention for PCC cognitive impairment, warranting larger trials to confirm efficacy.

## Introduction

Post-COVID-19 condition (PCC), or long COVID, affects millions globally, with cognitive dysfunction (“brain fog”) reported in up to 88% of patients, impairing attention, memory, and executive function.^1, 2, 3^ This debilitating symptom disrupts daily functioning, yet no effective treatments exist.^4, 5^

PCC brain fog is believed to arise from neuroinflammation, blood-brain barrier compromise, and disrupted default mode network (DMN) connectivity, compounded by mitochondrial dysfunction.^6, 7, 8, 9^ The neuroinflammation, driven by cytokine storms and microglial activation, compromises the blood-brain barrier, leading to cortical thinning and reduced DMN connectivity, impairing attentional and memory networks.^8, 9, 10, 11^ These collective neurological disruptions contribute to persistent cognitive deficits.^12^

No pharmacologic treatments target PCC cognitive symptoms, and while cognitive rehabilitation is under study, evidence-based therapies are urgently needed.^4, 13^

Photobiomodulation (PBM), or low-level light therapy, is a promising non-invasive strategy. PBM delivers near-infrared light (600–1100 nm) to modulate biological activity, enhancing mitochondrial function, increasing ATP production, reducing oxidative stress, and modulating inflammation.^14^ PBM delivering 40 Hz gamma oscillations could reduce amyloid burden and modify microglial activity for neuroprotection, and targeting the DMN may aid the recovery of impaired cognition.^15, 16^ An open-label study suggests transcranial PBM (tPBM) improves cognitive function in PCC. ^17^ When combining intranasal PBM with tPBM (itPBM) the modality could be more adapted to address complex and multifactorial pathophysiological mechanisms as demonstrated in traumatic brain injury studies.^18^

A prior trial of the Vielight RX Plus for acute COVID-19 showed that PBM targeting the upper respiratory system accelerated the patients’ recovery with clearer thinking. This experience prompted a targeted approach with the Vielight Neuro RX Gamma embodied as an itPBM modality, to deliver 810 nm near-infrared light at 40 Hz to DMN hubs.^19^

We conducted a randomized, double-blind, sham-controlled pilot clinical trial to evaluate the efficacy of home-based itPBM in adults with PCC-related cognitive dysfunction. The primary aim was to assess improvements in cognitive performance, with secondary outcomes including fatigue, quality of life, and safety.

## Methods

### Study Design

This prospective, randomized, double-blind, sham-controlled pilot clinical trial (**Supplementary 1: Protocol**), reported per CONSORT guidelines, evaluated the efficacy of itPBM for cognitive impairment in PCC. The home-based study was conducted remotely using an electronic data capture (EDC) system, with all assessments performed online. The protocol received IRB approval, followed ethical guidelines, and was registered at ClinicalTrials.gov (NCT05857124).

Participants were randomized 1:1 to active PBM (Vielight Neuro RX Gamma v2) or sham equivalent for 8 weeks (56 days), followed by a 4-week observation period. Blinding was maintained for participants, investigators, and assessors.

### Participants

Adults aged 18–65 meeting WHO PCC criteria with cognitive symptoms ≥12 weeks post-COVID were eligible.^1^ WHO criteria were operationalized via physician diagnosis of cognitive dysfunction and documented SARS-CoV-2 infection (positive PCR/antigen test or clinical diagnosis). English proficiency and internet access were required. Exclusion criteria included neurological or psychiatric conditions (e.g., dementia, uncontrolled depression), significant brain injury, unstable illness, pregnancy, photosensitivity, seizure risk, or use of cognition-impairing substances (**Supplementary 1: Protocol**).

Sex was self-reported as male or female, and race/ethnicity was self-reported using U.S. Census categories to assess representativeness. Enrollment occurred from April 2023 to September 2024. All procedures, including screening and device shipment, were remotely managed. Electronic informed consent was obtained through Castor eConsent.

### Randomization and Blinding

Randomization used computer-generated treatment assignment, stratified by age (<45 vs ≥45 years). Active and sham devices were identical visually with similar auditory cues and identified by randomization codes. Study personnel and participants were blinded to device allocation.

### Intervention

The active device delivered 810 nm near-infrared light at 40 Hz transcranially and intranasally via LEDs targeting DMN hubs (**Figs. 1a, 1b; Supplementary 2: PBM specifications**). The Vielight Neuro RX Gamma uses six transcranial and one intranasal LED to optimize cerebral perfusion and neural oscillation modulation, with the 40 Hz pulse rate chosen to enhance gamma entrainment.^20^ Sessions lasted 20 minutes daily, six days per week for 8 weeks. Participants were trained using a 10-minute video tutorial and written manual to ensure consistent home use. Participants recorded session completion and were asked to avoid other treatments or maintain stable medication doses. Sham devices mimicked operation without light delivery, using internal sensors to disable output on scalp contact. Compliance was monitored via daily electronic diaries and device usage logs, with reminders sent for missed sessions.

**Fig. 1a.**
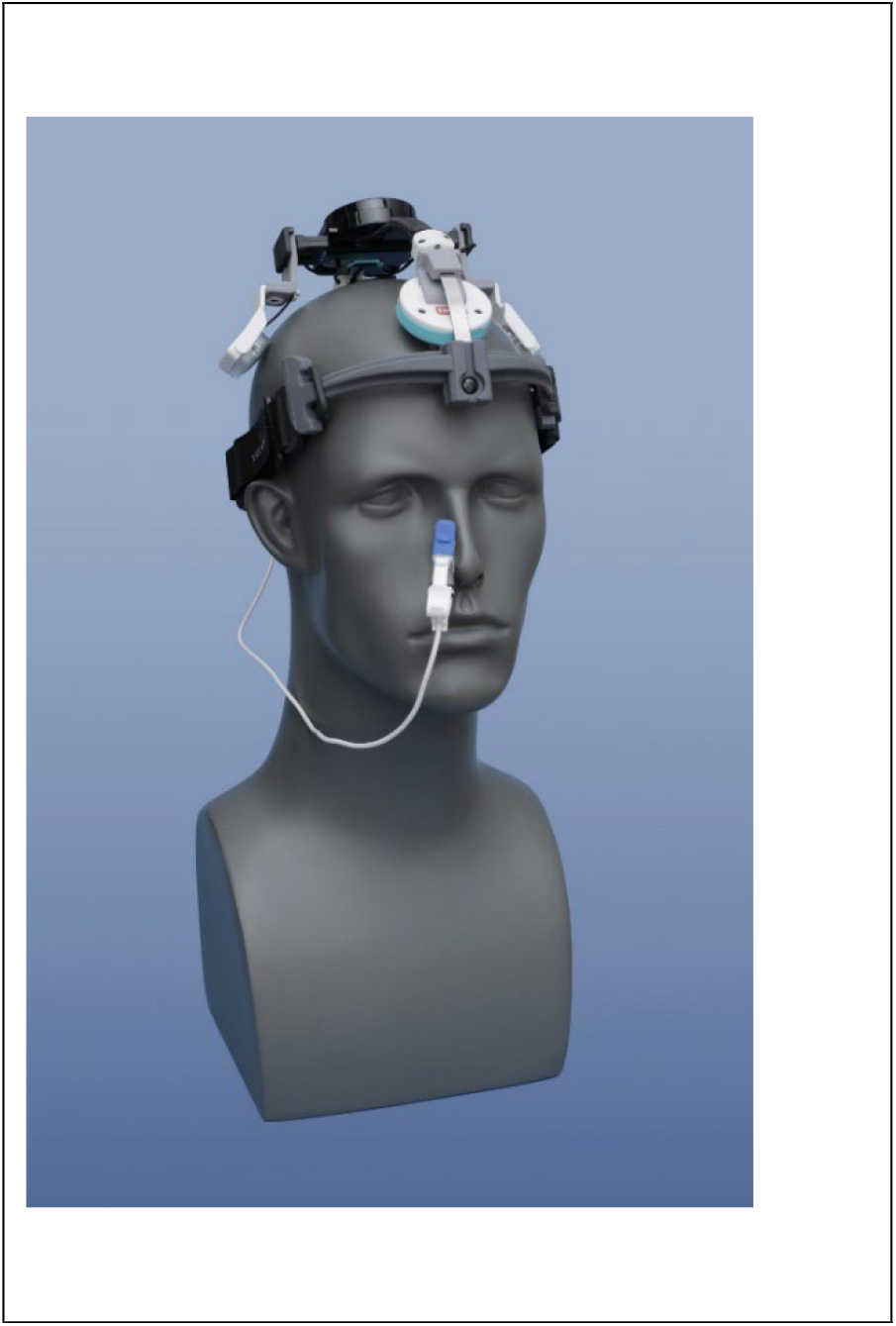
The intervention PBM device, Vielight Neuro RX Gamma.

**Fig. 1b.**
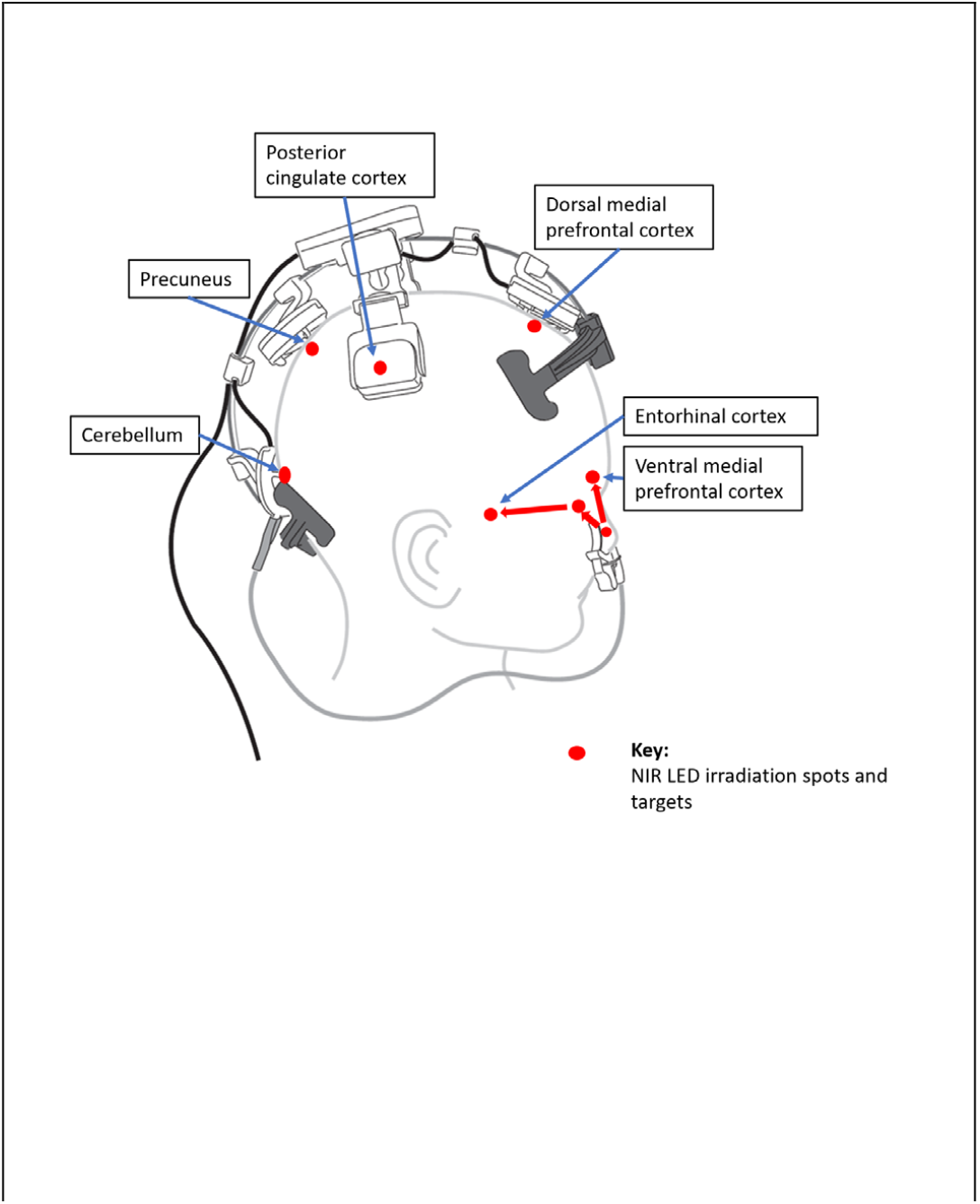
The near infrared (NIR) LEDs target the hubs of the default mode network (DMN) and cerebellum.

### Outcome Measures

#### Primary Outcome

The primary efficacy endpoint was the composite change in cognitive performance from baseline to Day 56, measured from the mean of seven tasks from the Creyos Research cognitive battery (formerly Cambridge Brain Sciences). These tasks assessed key cognitive domains: short-term memory (Monkey Ladder, Token Search), executive function and reasoning (Spatial Planning, Rotations), attention (Feature Match), and short-term memory (Paired Associates, with Polygons contributing aspects of visual memory). Online testing with the Creyos battery demonstrated comparable strength to comprehensive in-person assessment of cognitive performance.^21^ **Table 1** sets out the purposes of each task. Each task generated quantitative scores (higher = better), and a global composite score was calculated by averaging Z-scores across all Creyos tasks at each time point. Brain fog in long COVID was operationalized as impairment across these domains, with improvements in the composite score interpreted as cognitive recovery.

**Table 1.**
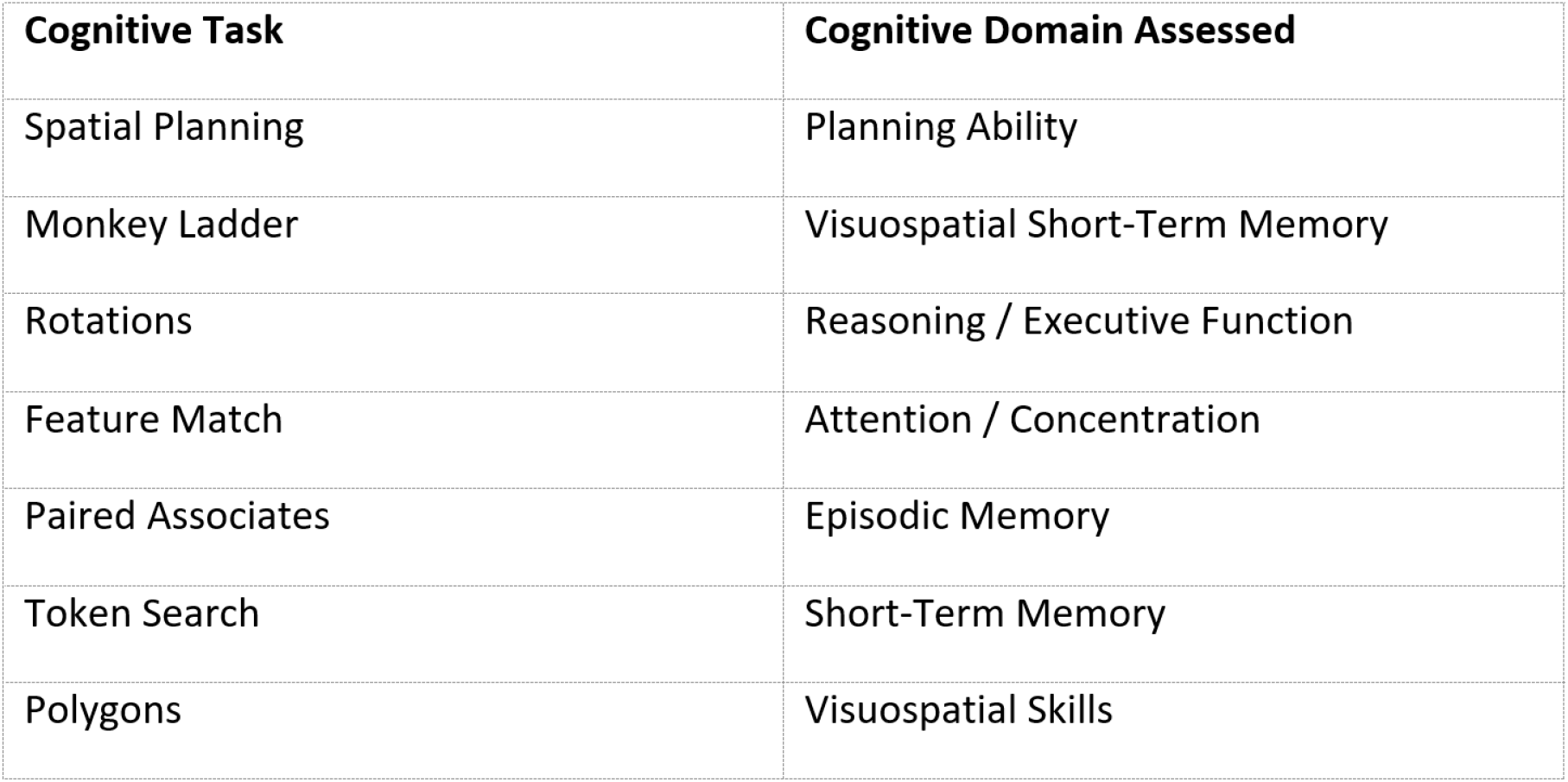
Key Cognitive Tasks – Components of Primary Endpoints.

#### Secondary Outcomes

- Additional Creyos tasks (Grammatical Reasoning, Spatial Span, Digit Span, Odd One Out, Double Trouble) captured broader cognitive function.
- Quality of Life: The EQ-5D-5L assessed mobility, self-care, usual activities, pain/discomfort, and anxiety/depression. A visual analog scale in response to ‘We would like to know how good or bad your health is today’ is recorded on a scale of 0 to 100.
- Fatigue: The Fatigue Assessment Scale (FAS) evaluated physical and cognitive fatigue (10 items, lower scores indicate less fatigue). The FAS is validated in chronic illness populations. ^22^
- Cognitive Symptoms: The PDQ-20 assessed perceived difficulties in memory, attention, planning, and organization.
- Symptom Burden: The modified Symptom Burden Questionnaire (MSBQ) captured PCC symptoms (e.g., fatigue, memory problems).

Safety: Adverse events (AEs) were self-reported in diaries and categorized per MedDRA.

### Data Collection

All data were collected electronically via Castor secure portal at baseline, Days 14, 28, 56, and 84. Participants maintained daily diaries for device usage, AEs, and health changes. Study staff sent reminders to ensure compliance.

**Fig. 2** presents the schedule of interventions and assessments. All entries were time-stamped and monitored for completion by the study team.

**Fig. 2.**
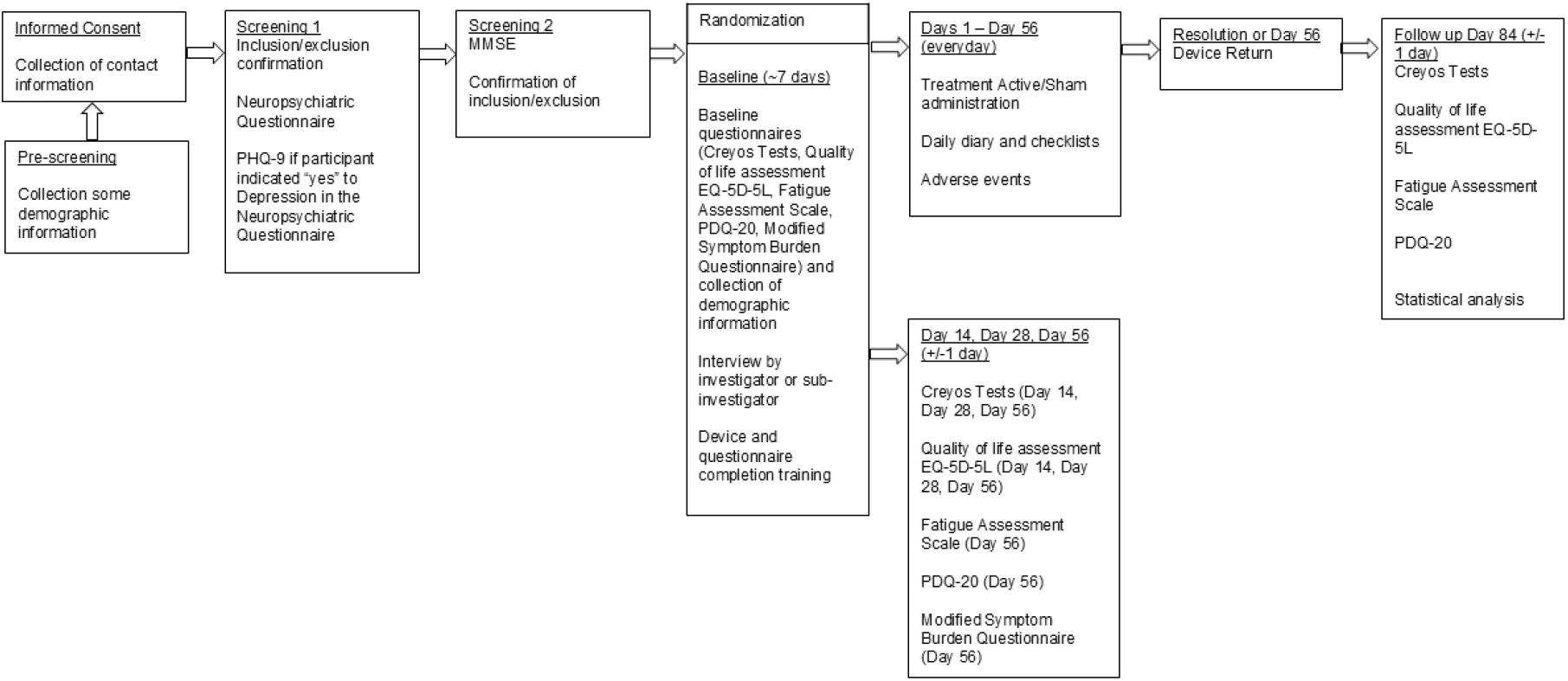
Study timeline depicting enrollment, PBM/sham administration, interim assessments, and follow-up.

### Statistical Analysis

This pilot study targeted 36 participants but enrolled 43 to assess preliminary efficacy, safety, and feasibility, ensuring adequate power for exploratory analyses. The Safety Population, equivalent to Intent-to-Treat (ITT) included all randomized participants who received the device. The Per-Protocol (PP) Population included those with valid Creyos data at baseline and adhered to the protocol, excluding major protocol deviations.

Primary outcomes were analyzed via mixed-model repeated measures (MMRM) with fixed effects for group, visit, group-by-visit interaction, age stratum, and baseline score covariate in the PP population. No adjustments for multiple comparisons were made for secondary outcomes and timepoints due to the exploratory nature. Effect sizes (Cohen’s d) were calculated for the primary endpoint. Secondary outcomes used ANCOVA for change from baseline to Day 56. Analyses used SAS v9.4 and R v4.1. A p-value < 0.05 is interpreted as statistically significant for hypothesis generation.

## Results

### Participant Disposition and Baseline Characteristics

Of 90 screened individuals, 47 were excluded (10 failed cognitive dysfunction criteria, 29 lost to follow-up, 8 withdrew). Forty-three participants were randomized: 23 to active PBM and 20 to sham. Thirty-three (76.7%) were female. Two active group participants were excluded from the PP analysis (one for incomplete Day 56 assessments, one for non-compliance), resulting in 41 PP participants (21 active, 20 sham). Ninety-eight percent completed Day 84 follow-up (**Fig. 3**).

**Fig. 3.**
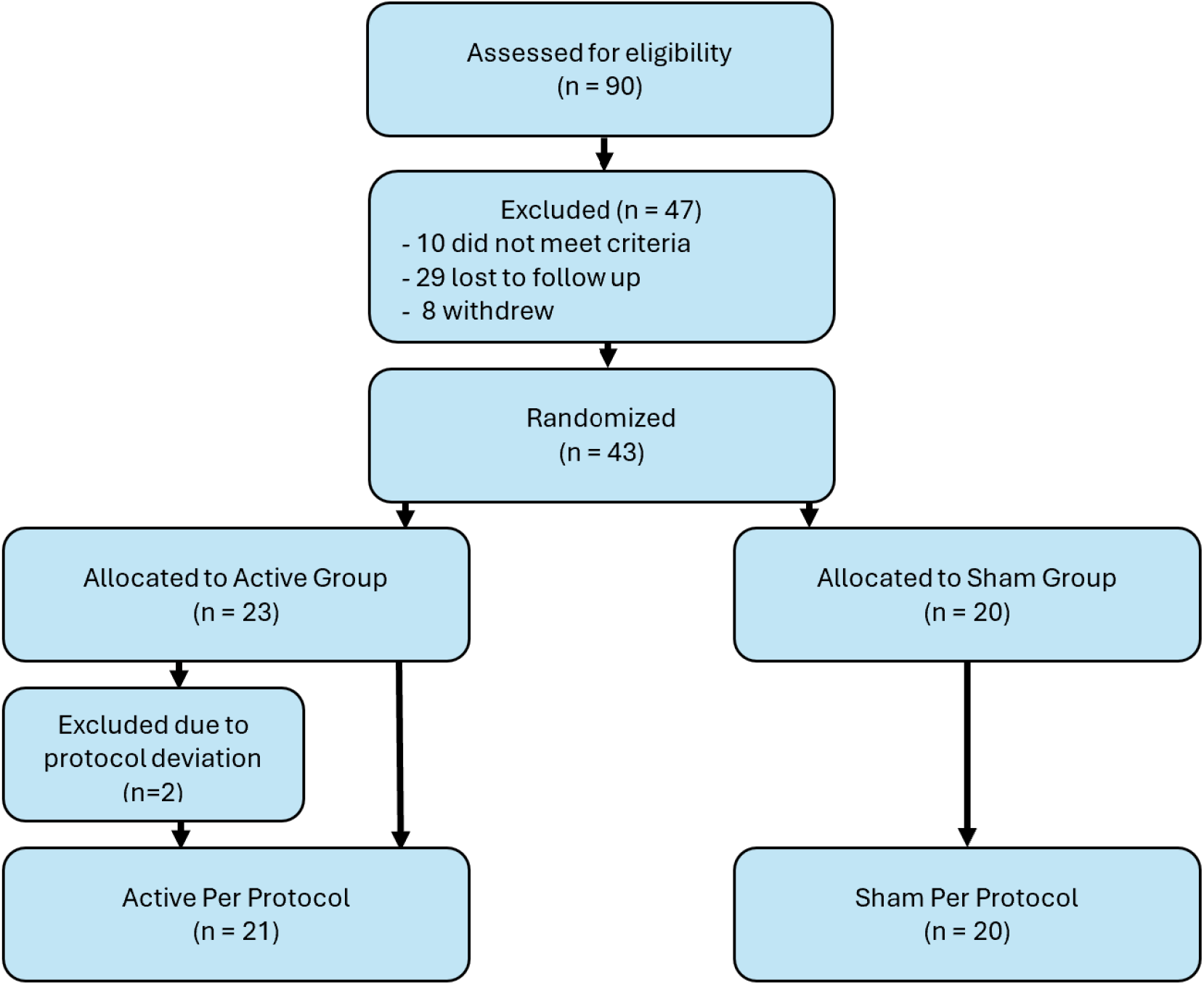
CONSORT flow diagram detailing recruitment, allocation, follow-up, and analysis. It illustrates the screening of 90 individuals, exclusion of 47 before randomization, and the flow through to final efficacy analysis with 41 participants. No sham participants were lost to follow-up; two active participants were excluded due to protocol deviations.

Table 2 summarizes baseline characteristics. Mean age was 40.6 years (SD 10.6), with 76.7% female and 93% White, non-Hispanic. The active/sham groups were each balanced for age, sex, and BMI. Sex differences were not considered in the endpoint analyses, justified by their balance at baseline (**Supplementary 3 table 1**).

**Table 2.** Demographics and baseline characteristics (Safety Population, n=43). Values are n (%) or mean ± SD. No significant differences between active and sham population within the age groups. BMI = Body Mass Index.

| Characteristic | Active PBM (n = 23) | Sham (n = 20) | Total (N = 43) | p-value (between groups) |
| --- | --- | --- | --- | --- |
| Sex – Female | 17 (73.9%) | 16 (80.0%) | 33 (76.7%) | 0.641 |
| Sex – Male | 6 (26.1%) | 4 (20.0%) | 10 (23.3%) | — |
| **Age (years)** | 39.2 ± 10.9 | 42.2 ± 10.3 | 40.6 ± 10.6 | 0.365 |
| Age < 45 years | 17 (73.9%) | 14 (70.0%) | 31 (72.1%) | 0.778 |
| Age ≥ 45 years | 6 (26.1%) | 6 (30.0%) | 12 (27.9%) | — |
| Race – White | 22 (95.7%) | 18 (90.0%) | 40 (93.0%) | 0.557 |
| Race – Black | 1 (4.3%) | 1 (5.0%) | 2 (4.7%) | — |
| Race – Other<sup>+</sup> | 0 (0%) | 1 (5.0%) | 1 (2.3%) | — |
| Ethnicity – Hispanic/Latino | 1 (4.3%) | 2 (10.0%) | 3 (7.0%) | 0.473 |
| Ethnicity – Non-Hispanic | 22 (95.7%) | 18 (90.0%) | 40 (93.0%) | — |
| BMI (kg/m<sup>2</sup>) | 27.1 ± 5.3 | 26.4 ± 4.8 | 26.8 ± 5.1 | 0.684 |
| Current smoker | 0 (0%) | 0 (0%) | 0 (0%) | — |
| Former smoker | 8 (34.8%) | 2 (10.0%) | 10 (23.3%) | 0.082* |

<sup>+</sup> “Other” race category includes participants who identified with a race not listed (e.g., multi-racial).
\* Fisher’s exact test for former smoker status.

### Cognitive Outcomes (Primary Endpoint)

All participants exhibited some degree of cognitive impairment at baseline relative to normative data (by inclusion criteria). Over the 8-week treatment period, both age groups showed improvement in cognitive test scores over their baselines, but improvements were generally greater in the active PBM group, and they were statistically significant for the <45 years of age stratum. **Fig. 4a** presents the primary summary composite score of active minus sham in least square (LS) mean, as the targeted cognitive change from baseline for Day 56, defined as the primary endpoint. The detailed results dataset is shown in **Supplementary 3 tables 4, 4.1, 4.2 and 5, 5.1, 5.2.**

**Fig. 4a.**
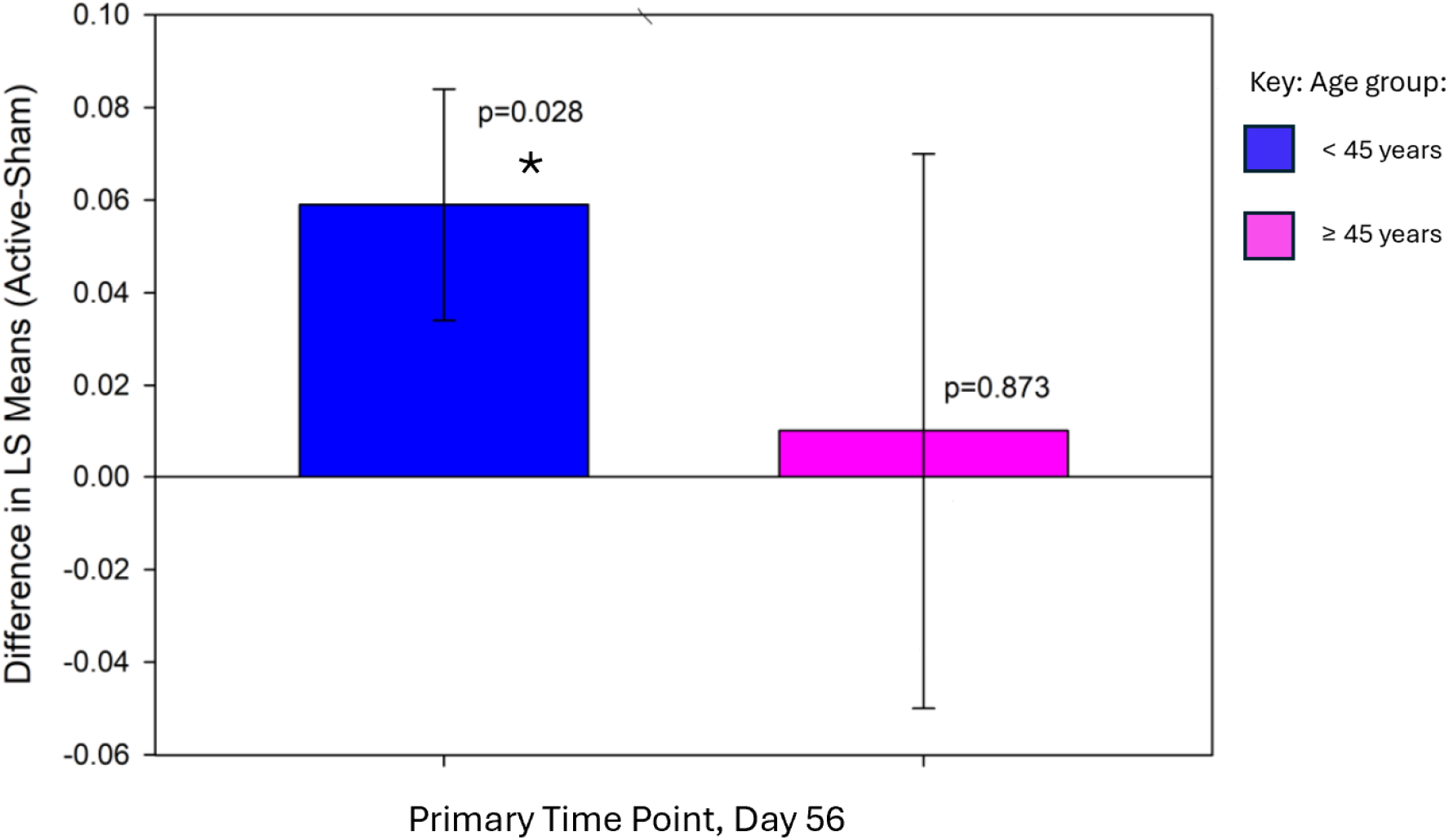
**Primary endpoint by age stratum**, showing the change in the least squares (LS) mean difference in composite cognitive score between active and sham groups by age group on Day 56. Each bar or column represents the LS mean difference, includes standard error bars and p values

**Fig. 4b** illustrates the mean composite cognitive performance of active minus sham over the time points by age in LS mean. It is notable that after the evaluation period of 56 days, both the <45 and ≥ 45-year-old populations were trending to continued improvement.

**Fig. 4b.**
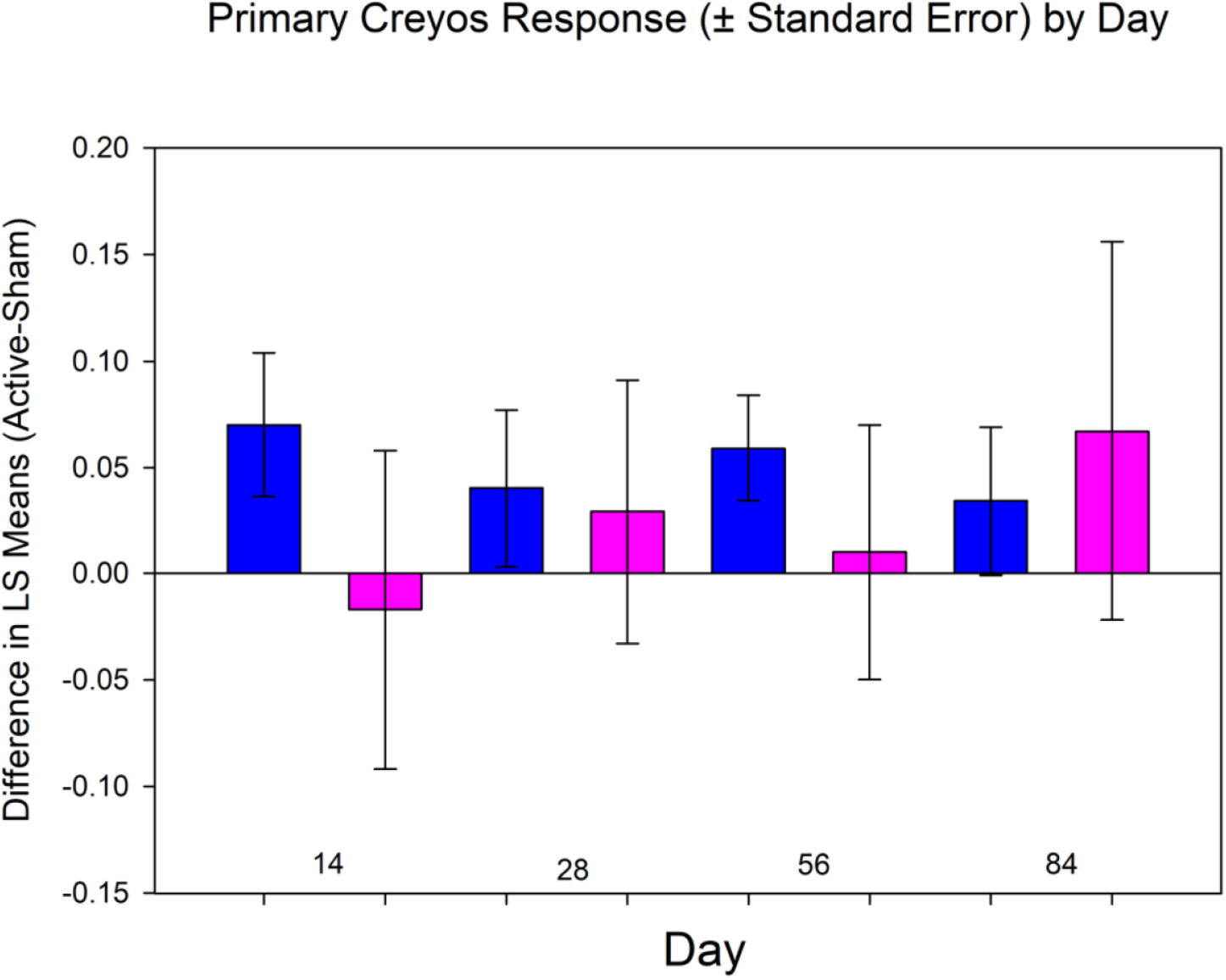
**Change in Composite Cognitive Score** from Baseline (Active-Sham) by Age group in LS mean. The columns display the change from baseline in composite cognitive score by treatment group and age stratum across the time points (Days 14, 28, 56, 84) with standard error bars. Statistical significance (*) of p=0.0280 on day 56 was demonstrated by the <45-year-old age stratum. The remaining timepoints showed the trend although not statistically significance.

By the end of treatment (Day 56), the composite cognitive score, reflecting overall performance across the seven core cognitive tasks, improved more in the active PBM group (mean change = 0.050) than in the sham group (mean change = 0.007), though the difference between the active and sham groups of 0.043 (95% CI: –0.007 to 0.092) did not reach statistical significance (p = 0.088).

The pre-specified subgroup analysis by age revealed a clearer pattern. Among participants under 45 years (n = 31), the active PBM group demonstrated a greater improvement in the composite score (mean = 0.082) than the sham group (mean = 0.023) at Day 56, yielding a statistically significant difference of 0.059 (SE = 0.025; 95% CI: 0.007 to 0.111; p = 0.028). In participants aged 45 and older, the mean change in the active group (0.026) exceeded that of the sham group (0.016), but the difference (0.010) was not statistically significant.

Effect size analysis (Cohen’s d) for the primary endpoint revealed a small effect overall (d = 0.280), a medium effect in participants under 45 years (d = 0.438), and a negligible effect in those 45 years and older (d = 0.048). These findings reinforce the age-related differences observed in the main analyses.

In analyzing the secondary component cognitive tasks, improvements were observed across most tasks in the active group illustrated by **Fig. 4c**.

**Fig. 4c.**
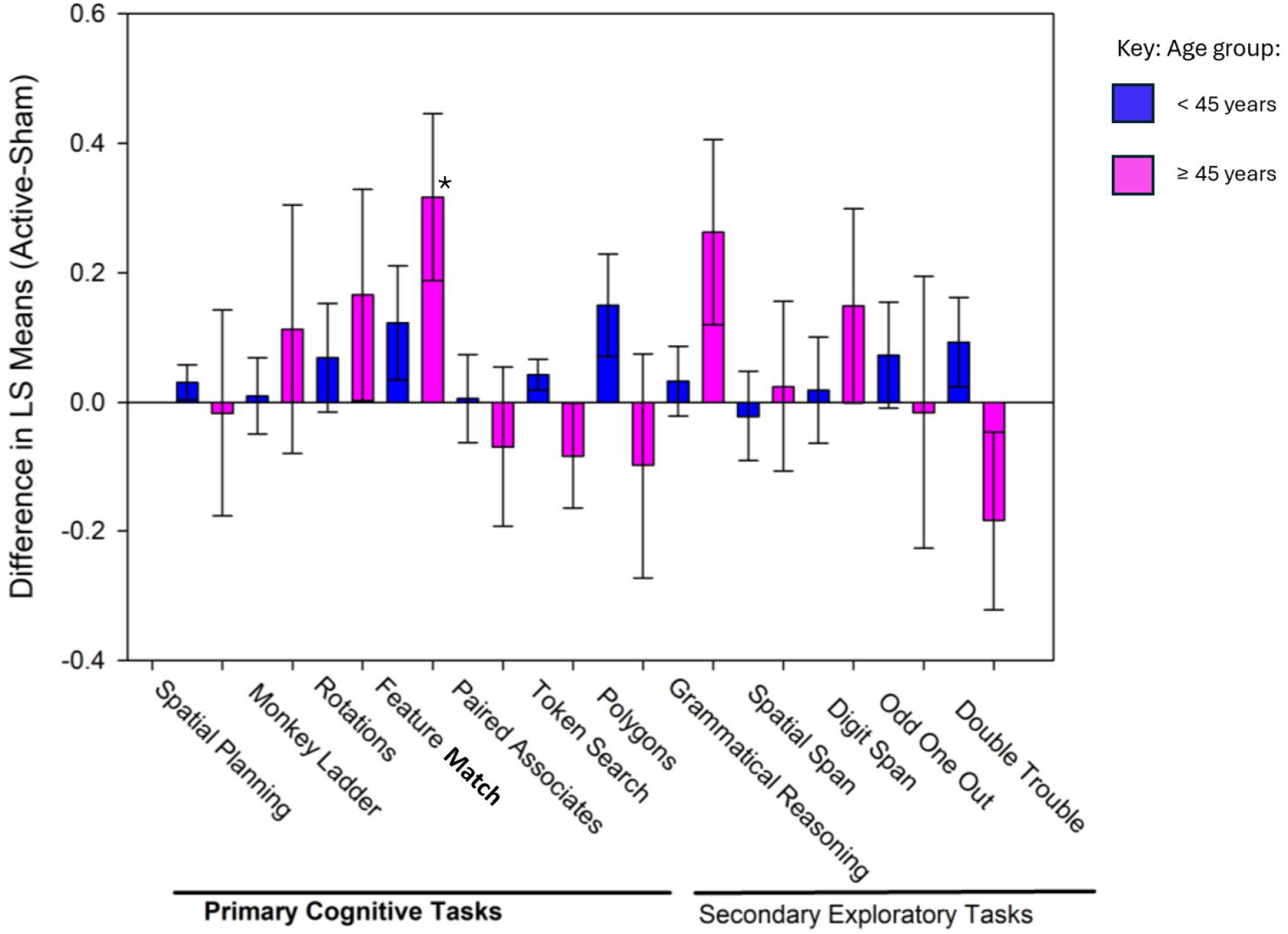
Component Cognitive Task Outcomes by Age Group at Day 56. The bar chart displays the LS mean differences (Active – Sham) for each component cognitive task at Day 56, grouped by age, the blue columns for participants under 45 years and the magenta-colored columns for those 45 years and older. Error bars represent the standard error of the LS mean difference. The Feature Match score of the ≥45 years old showed statistical significance (*) with p=0.034.

When analyzed individually, six of the seven component tasks favored active PBM on Day 56, though only one—Feature Match (a test of visual attention and pattern recognition)—showed statistically significant in active-sham group differences.^23^ For the all-age-groups sample, significant improvements in Feature Match accuracy for the active group compared to sham were observed at Day 28 (p = 0.019) and Day 84 (p = 0.017). In the ≥45 age group, active PBM outperformed sham at Day 28 (p = 0.023) and Day 56 (p = 0.034). In the <45 age group, performance on Feature Match consistently favored the active group at all timepoints, reaching significance at Day 84. See details in **Supplement 3 tables 5, 5**,**1, 5.2**.

In the older age group (≥45 years), improvements in tasks such as Feature Match, Rotations (mental rotation), and Monkey Ladder (visuospatial working memory) were offset by lesser or negative changes in Paired Associates (episodic memory), Token Search (working memory/strategy), Spatial Planning (planning), and Polygons (visuospatial skills), contributing to a non-significant composite effect.

In summary, active PBM was associated with consistent cognitive improvements, especially in younger participants. The strongest and most consistent gains were seen in attentional performance (Feature Match), with multiple significant differences over sham. While the composite score improvement in the full sample did not reach statistical significance, the results suggest a meaningful cognitive signal— particularly in younger individuals—which warrants further exploration in larger studies. **Table 3** highlights the primary cognitive outcomes.

**Table 3.** Primary cognitive outcome results at Day 56 (change from baseline) in the Per-Protocol population.

| **Cognitive Outcome** | **Active PBM vs Sham: Outcome at Day 56** | **p-value** |
| --- | --- | --- |
| **Composite Cognitive Score** (all ages) | Greater improvement in Active (mean increase vs baseline higher than Sham) | 0.088 (n.s.) |
| **Composite Cognitive Score** (<45 subset) | Active significantly > Sham (notable improvement in Active group) | 0.028 * |
| **Feature Match task** (attention) | Active significantly > Sham (better performance by Day 28 onward) | < 0.05 † |
| Other individual cognitive tasks (6 tasks) | No significant group differences (trends favoring Active on some tasks) | n.s. |

\*Statistically significant difference favoring Active PBM in participants <45 years old.
†Statistically significant difference favoring Active PBM in the overall group for the Feature Match task at Day 28 (p=0.019), Day 56 (p=0.133), Day 84 (p=0.017).

### Secondary Outcomes

#### Quality of Life (EQ-5D-5L)

Both groups reported modest decreases with less negative impact in sham PBM on overall quality of life, with no significant differences in EQ-5D index at Day 56. The <45 sham PBM group showed significantly greater improvement in the mobility domain at Day 56 (p = 0.007), and this advantage emerged as early as Day 14. The VAS measure improved in both groups but was not statistically significant at Day 56. In summary, beyond mobility, PBM did not offer superior quality of life benefits during the treatment period.

#### Fatigue (Fatigue Assessment Scale)

Fatigue Assessment Scale (FAS) sum of response aligned to questions 1 to 10 favored sham treatment. Combining all age groups at Day 56 was statistically significant favoring sham in “I am bothered by fatigue” (p = 0.038), “and “I get tired very quickly” (p=0.037). Specifically for the older ≥45 group, sham was favored in “I have enough energy for everyday life” (p-0.029) and “I have problems to think clearly” (p=0.020).

#### Cognitive Symptom Questionnaire (PDQ-20)

Perceived cognitive difficulties (PDQ-20) improved similarly in both groups without statistical significance in most of the scores at Day 56. An exception was shown by younger PBM participants reporting significantly less difficulty in remembering names at Day 56 (p = 0.048). Among older participants, sham users reported greater improvements at Day 84 for trouble concentrating (q 9) and forgetting to take medication (q 19).

#### Symptom Burden (MSBQ)

Both groups reported slight decreases in total symptom burden, though no statistical significance between-group differences were observed. However, the sham group, particularly in the older group, reported greater improvement across key symptoms, including fatigue and cognitive issues. By Day 84, symptom scores converged somewhat as PBM participants continued to improve post-treatment.

Table 4 provides a summary of key secondary outcome results at the primary endpoint (Day 56).

**Table 4.** Summary of secondary outcome results at Day 56 (change from baseline).

| **Outcome Measure** | **Between-Group Comparison (Active PBM vs Sham)** | **p-value** |
| --- | --- | --- |
| **EQ-5D-5L Mobility (function)** | Active PBM improved more (greater reduction in mobility limitations) | 0.007 |
| EQ-5D-5L Other Domains (Self-care, etc.) | Favoring active PBM but not significant | > 0.05 (NS) |
| EQ-5D VAS (overall health) | No significant difference (trend: sham higher VAS) | 0.10 (NS) |
| Fatigue (FAS total score) | No significant difference (fatigue improved in both groups) | 0.67 (NS) |
| **FAS key items (fatigue perception)** | Sham better on some (less "bothered by fatigue") | < 0.05 on select items |
| Cognitive symptoms (PDQ-20 total) | No significant difference overall | 0.75 (NS) |
| **PDQ-20 key items (memory, attention)** | Mixed: Active better on some (younger memory), Sham better on others (older concentration) | < 0.05 on select items |

| Outcome Measure | Between-Group Comparison (Active PBM vs Sham) | p-value |
| --- | --- | --- |
| Symptom Burden (MSBQ total) | No significant difference | > 0.05 (NS) |
| MSBQ specific symptoms | No significant differences (trend toward sham reporting less fatigue, cognitive issues and 'Memory' in Sham was superior at Day 56 (p=0.011) in older adults) | > 0.05 (NS) |

NS = not statistically significant. Bolded outcomes highlight where a statistically significant difference was observed in favor of one group.

#### Adverse Events and Tolerability

No serious adverse events (AEs) occurred. Common AEs included mild headache (7/43, 16.3%) and skin irritation (6/43, 14.0%), resolving without intervention (**Supplementary 3, Table 9**). One active participant reported nasal discomfort, alleviated with saline gel. Treatment adherence was high (median 45/48 sessions), with 90% achieving >80% compliance.

## Discussion

This pilot randomized controlled trial provides encouraging evidence that home-based intranasal and transcranial photobiomodulation (itPBM) is a safe and feasible intervention for cognitive impairment in long COVID. Active PBM improved composite cognitive scores more than sham (p=0.088), with significant gains in participants <45 years (p=0.028), likely due to enhanced neuroplasticity, as 40 Hz gamma oscillations strengthen synaptic connectivity in neural networks.^24^ Older adults may require higher PBM doses to achieve similar effects.^25^

Significant improvements in the Feature Match task (attention) across groups align with PBM’s modulation of gamma oscillations, known to enhance attentional processing.^18^ Younger participants’ greater responsiveness may reflect higher baseline neuroplasticity, enabling more robust neural repair.^**Error! Bookmark not defined**.^ PBM’s anti-inflammatory and mitochondrial-enhancing effects likely mitigate PCC’s neuropathophysiology, restoring cortical function.^26, 27^

Secondary outcomes were mixed, with sham PBM improving mobility (p=0.007) particularly for the younger participants. Through assessments with FAS, fatigue generally favored sham. The lifting of brain fog may increase the awareness of fatigue.^28^ PCC patients have limited energy reserves. If more of this energy is directed toward cognitive improvement, it may temporarily reduce physical energy, leading to increased fatigue and reduced mobility.

Through the PDQ-20 assessments, the younger participants remembered names better, but the older participants were more forgetful after the treatment stopped. It may suggest that the older population may have experienced neurodegenerative characteristics in long COVID that were previously arrested with itPBM and resumed after treatment stopped.^29^ A similar phenomenon was observed in prior itPBM studies of neurodegenerative diseases covering dementia and chronic traumatic encephalopathy (CTE).^30, 31^

Compared to cognitive rehabilitation’s mixed efficacy or unproven pharmacological treatments, PBM offers a non-invasive, safe alternative, though efficacy is more predictable in younger adults.

### Clinical Implications

Cognitive gains, particularly in attention, could enhance daily functioning and work productivity for PCC patients. Continuous treatment is suggested for the older patients to avoid some potential post-treatment memory decline. PBM’s home-based delivery offers a scalable, cost-effective intervention, potentially integrating with cognitive rehabilitation to enhance functional recovery.

### Limitations

The small sample size limited statistical power to detect subtle effects. The 8-week duration may not capture long-term recovery. Remote assessments risked variability from environmental distractions. The predominantly White (93%) and female (76.7%) cohort limits generalizability; diverse recruitment is needed in future trials. Lack of sex/gender-based analyses, due to sample size, is a further limitation to generalizability. Absence of biomarkers (e.g., EEG, inflammatory markers) restricted mechanistic insights.

### Future Directions

Larger trials should explore alternative PBM parameters (e.g., higher dose) for older adults and incorporate EEG and inflammatory biomarkers to elucidate mechanisms. Diverse populations and sex/gender analyses are critical to ensure equitable outcomes, addressing the current study’s demographic limitations.

## Research in Context

### Evidence before this study

We searched PubMed and Google Scholar for studies on photobiomodulation (PBM) for cognitive impairment in post-COVID-19 condition (PCC) from January 2020 to March 2025, using terms “photobiomodulation,” “long COVID,” and “cognitive dysfunction.” No randomized controlled trials were found, but studies suggested PBM improved cognitive function in conditions like PCC. PBM’s improvement of other neurological conditions, such as traumatic brain injury and dementia suggest that it could be favorable to brain fog in PCC with its similar underlying pathophysiology. Examples in preclinical data indicated PBM enhances mitochondrial function and reduces neuroinflammation, aligning with PCC’s pathophysiology.

### Added value of this study

This pilot randomized, double-blind, sham-controlled trial provides the first controlled evidence of PBM’s efficacy for PCC cognitive impairment, showing significant improvements in attention and composite cognitive scores in younger adults (<45 years).

Implications of all the available evidence: PBM is a safe, feasible intervention with potential to alleviate PCC cognitive dysfunction, particularly in younger patients. Larger trials are needed to confirm efficacy, optimize parameters for broader populations, and explore its role in global health systems.

## Supporting information

Supplementary 1: Protocol

Supplementary 2: PBM Specifications

Supplementary 3: Detailed Results Dataset

## Data Availability

All data produced in the present study are available upon reasonable request to the authors.

## Declarations

### Authors’ Contributions

LL, NH, MVB, AP, JL, ZA, AP designed the study and wrote the protocol. KO and JR were the clinical investigators, AB managed data collection and participant follow-up, MVB accessed, verified the data and performed statistical analyses. JD was the medical monitor. RZ provided supervision on the study quality, objectivity, and contributed to statistical analyses. LL wrote the main manuscript. All authors contributed to data interpretation and approved the final version.

### Data Sharing Statement

Deidentified participant data and a data dictionary will be available upon request after publication, subject to approval of a proposal and a signed data access agreement. Data will be accessible via the corresponding author for 5 years post-publication. The study protocol is included as Supplementary 1.

## Acknowledgments

None.

