## Supplementary 1: Protocol for "Photobiomodulation for Cognitive Dysfunction (Brain Fog) in Post-COVID-19 Condition: A Randomized Sham-Controlled Pilot Trial"

#### VIELIGHT NEURO RX GAMMA FOR POST COVID-19

#### A Pilot Study Evaluating the Efficacy of the Vielight Neuro RX Gamma in the Treatment of post COVID-19 Cognitive Impairment

---

Protocol No:

Revision date: 2023-08-16 V2.0

Study Products: Vielight Neuro RX Gamma

SPONSOR: Vielight Inc.
Contact: Lew Lim, PhD, MBA
346A Jarvis St.
Toronto, Ontario
M4Y 2G6, Canada.
1-855-875-6841


***CONFIDENTIAL:*** *This protocol, its contents, and the information relating to it are the property of Vielight Inc. All information is to be kept confidential.*

**REVISION HISTORY**

2023-03-07 Initial protocol drafted by Vielight Research Team

2023-08-15 Protocol revised to eliminate the MMSE screening tool and update the inclusion criteria based on physician's diagnosis, and to add the Creyos test on the screening

**Summary of changes, Protocol version V2.0**

| **Item (Section)** | **Original Text** | **Revised Text** | **Rational for Change** |
|---|---|---|---|
| Table of contents | | | Updated |
| 1.0 Protocol Summary and Section 6.2 | Mini Mental State Examination (MMSE) score of <27 | ~~Mini Mental State Examination (MMSE) score of <27~~<br><br>Meet physician's diagnosis of Cognitive Dysfunction. | To update the inclusion criteria using physician's diagnosis |
| 1.0 Protocol Summary | The primary efficacy endpoint | The z-scores and percentiles relative to an age (decile) adjusted | |

©2023 Vielight Inc.
All rights reserved

BRANY
IRB APPROVED
08/24/2023

| | | | |
|---|---|---|---|
| | is the change from baseline to Day 56 in the combined scores of the 7 Creyos tests (above). | reference population will be performed.<br>. | |
| Throughout the whole text | | | Wording and grammatical update |
| 1.0 Protocol Summary | | PD subjects will exclude those with significant protocol deviations with potential effect on efficacy as determined prior to unblinding. | |
| 1.0 Protocol Summary | Creyos scores of the 5 remaining tests (i.e. Grammatical Reasoning, Spatial Span, Digit Span, Odd One Out, Double Trouble) and changes from baseline for Sham and Active treatment devices will be compared at each testing timepoint. | The change from baseline in Creyos scores of the primary efficacy endpoint (7 tests) and secondary Creyos items (5 remaining tests) will be compared between treatments at each follow-up interval through Day 56 (i.e., Grammatical Reasoning, Spatial Span, Digit Span, Odd One Out, Double Trouble) and changes from baseline for Sham and Active treatment devices will be compared at each testing timepoint. The Creyos questionnaire is administered at baseline, 14, 28, 56 and 84. | Secondary endpoint was updated for clarity |

©2023 Vielight Inc.
All rights reserved

BRANY
IRB APPROVED
08/24/2023

| | | | |
|---|---|---|---|
| Section 2.1 | | Brain fog is defined in Section 2.2.1. | Added text for clarity |
| Section 4 and 5.5.1 | | Figures 7 and 8 where updated | To eliminate the MMSE test and add Creyos test at screening |
| Section 5.3 | | The Creyos online test will be sent to participants at the screening. We have the ability to resend the questionnaires through the EDC as reminders but will need to call or email subjects to remind them to complete Creyos. | Added text for clarity on the process |
| Section 5.4 | Study staff will complete a randomization using SAS (SAS, Inc., Cary NC) software. Participants will be stratified by age (>=45 years, < 45 years) at baseline and randomly allocated to the Treatment group or Sham group (1:1) which aims to reduce the differences in the two groups | Study staff will complete a randomization list that was prepared using SAS (SAS, Inc., Cary NC) software. Participants will be stratified by age (>=45 years, < 45 years) at baseline and randomly allocated to the Treatment group or Sham group (1:1) which aims to reduce the differences in the two groups based on baseline factors. | Added text for clarity |

| | | | |
|---|---|---|---|
| | based on baseline factors by equal distribution within strata to the device and sham device. | | |
| Section 5.7 | | It will be recommended that the participants take the Creyos tests at a constant time of the day preferably starting in the afternoon.<br><br>In the rare event that the study device is not working properly, a replacement treatment device will be provided. In this instance, the study blind would be maintained by communication of the replacement device serial number to the study personnel. | Added text for clarity on the process |
| Section 8 | The pharmacodynamic population (PD) includes safety subjects with a CBS<br>measurement at baseline and at least one post-baseline measurement. | The pharmacodynamic population (PD) includes safety subjects with a Creyos measurement at baseline and at least one post-baseline measurement with no significant protocol deviations as determined prior to unblinding. | Added text for clarity |

©2023 Vielight Inc.
All rights reserved

BRANY
IRB APPROVED
08/24/2023

| | | | |
|---|---|---|---|
| Section 15 | **Mini Mental State Examination (MMSE)** - is a questionnaire that is used extensively in clinical and research settings to measure cognitive impairment. | | Deleted the MMSE definition and test from Appendix 1 |

**Table of Contents**

1.0 Protocol Summary 11

2.0 Introduction 18

2.1 Post COVID-19 Pathology and Study Focus 18

2.2 Investigational Device: The Vielight Neuro RX Gamma 20

2.2.1 Design Principles 20

2.2.2 Device Description 22

A. System Architecture 24

24

B. Headset with built-in controller 24

C. Nasal Applicator 24

D. Power Supply 25

E. Device Specifications 25

2.2.3 Device Placement 25

2.2.4 Procedure 26

2.2.5 Regulatory Status 26

2.3 Mechanisms of Action of the Vielight Neuro RX Gamma 26

2.3.1 The Systemic Effect of Irradiating the Free-Floating Mitochondria 27

2.3.2 Anti-inflammatory effect of PBM 27

2.4 Clinical Data and Summary of Related Evidence to Date 27

2.4.1 Support recovery of damaged cells 28

©2023 Vielight Inc.
All rights reserved

BRANY
IRB APPROVED
08/24/2023

2.4.2 Management of inflammation 28
2.4.3 Vielight devices have produced evidence of tissue responses 28
2.4.4 Track record of efficacy in other clinical studies 28
3.0 Study Objective and Rationale 28
3.1 Study Objective 28
3.2 Study Design Rationale 28
4.0 Overview of Study Design 29
5.0 Study Procedures 29
6.0 Eligibility Criteria and Recruitment 35
6.1 Recruitment Strategy 35
6.2 Inclusion Criteria 35
6.3 Exclusion Criteria 35
7.0 Study Endpoints 36
7.1 Primary Endpoint 36
7.2 Secondary Endpoints 37
8 Statistical Methods 38
8.1 Randomization and Stratification 39
8.2 Blinding 39
8.3 Efficacy Endpoint Analysis 39
8.3.1 Primary Endpoint 39
8.3.2 Secondary Endpoints 40
8.3.2.1 EQ-5D-5L Quality of Life 40
8.3.2.2 FAS 40
8.3.2.3 PDQ-20 40
8.3.2.4 Secondary Creyos item 40
8.3.2.5 SBQ 41
8.3.3 Compliance 41
8.3.4 Exploratory Endpoints 41
8.4 Assessment of Device Safety: Incidence of device attributable adverse events and usability 41
8.5 Subgroup Analysis 42
8.6 Interim Analysis 42
9 CRO and Data Collection Methods 42
9.1 Study Duration 42

©2023 Vielight Inc.
All rights reserved

BRANY
IRB APPROVED
08/24/2023

10 Ethical Considerations 42
10.1 Consent Process 42
10.2 Institutional Review Board (IRB) and Research Ethics Board (REB) Approvals 43
10.3 Data Protection 43
10.4 Study Suspension or Early Termination 43
11 Investigational Devices 44
12 Safety and Adverse Events 44
12.1 Definition of Adverse Events 44
12.2 Definition of Device Incident 45
12.3 Reporting Adverse Events and Device Incidents 46
12.4 Adverse Event Follow-up 46
12.5 Anticipated Adverse Events 46
12.6 Protocol Deviations 46
13 Study Management 47
13.1 Sponsor Overall Responsibility 47
13.2 Investigator Responsibilities 47
13.3 Required Documents from the Investigators 48
13.4 Investigator Records 48
13.5 Specific Sponsor Responsibilities 49
13.6 Sponsor Records 50
13.7 Study Monitoring Plan 50
13.8 Medical Monitoring 50
13.9 Ethical Considerations 50
13.10 Protection of Subject Confidentiality 51
13.11 Quality Assurance and Supervision by Authorities and Privacy 51
13.12 Final Report 51
13.13 Information Confidentiality 51
13.14 Trial Registration 52
14 Study Close-out 52
14.1 Timeline of Close-out 52
14.2 Record Storage and Retention 52
14.3 Discontinuation of Study 52
15 Definitions and Acronyms 52
16 References 55

©2023 Vielight Inc.
All rights reserved

BRANY
IRB APPROVED
08/24/2023

Appendix 1 59
Appendix 2 61
Appendix 3 61
Appendix 4 63
Appendix 5 64
Appendix 6 67

#### 1.0 Protocol Summary

| | |
|---|---|
| Study Title | **A Pilot Study Evaluating the Efficacy of the Vielight Neuro RX Gamma in the Treatment of Post COVID-19 Cognitive Impairment** |
| Methodology | Double-blind, Sham-controlled, Prospective, parallel randomized clinical trial |
| Study Duration | Duration for each participant is approximately 3 months. Duration of trial (from first subject enrolled to completion of final participant) is expected to be approximately 12 months. |
| Study Site(s) | Participants are monitored remotely by the Contract Research Organization (CRO) and Study Investigators |

©2023 Vielight Inc.
All rights reserved

BRANY
IRB APPROVED
08/24/2023

| | |
|---|---|
| Number of Subjects | 36 enrolled subjects (18 active Vielight Neuro RX Gamma; 18 sham) |
| Major Inclusion Criteria | ● Age 18-65<br>● Meets WHO-defined post-COVID-19 condition (WHO definition: 'Post COVID-19 condition occurs in individuals with a history of probable or confirmed SARS-CoV-2 infection, usually 3 months from the onset of COVID-19 with symptoms that last for at least 2 months and cannot be explained by an alternative diagnosis. Common symptoms include fatigue, shortness of breath, cognitive dysfunction but also others (more information is found here)* and generally have an impact on everyday functioning. Symptoms may be new onset following initial recovery from an acute COVID-19 episode or persist from the initial illness. Symptoms may also fluctuate or relapse over time. To ensure the above criteria is met, participants will only be included in the study if they meet all eligibility criteria more than 12 weeks from the onset of their acute Covid-19 symptoms or positive PCR/antigen test.<br>● Meet physician's diagnosis of Cognitive Dysfunction.Documented history of SARS-CoV-2 infection (positive PCR/antigen test during acute illness OR clinical diagnosis by physician during or after the acute illness).<br>●<br>● Ability to provide informed consent.<br>● Ability to read and communicate in English |

| | |
|---|---|
| Major Exclusion Criteria | • Current symptoms are explained by a psychiatric or neurological disorder (e.g., major depressive disorder or bipolar disorder).<br>• Having or history of any major neurological or psychiatric illness, and such conditions that may cause cognitive impairment, or symptoms like those seen in post-COVID-19 condition (e.g., mild, or major neurocognitive disorder, lifetime psychotic episodes, bipolar disorder, active suicidal ideation, and homicidal ideation, or diagnosis of chronic fatigue syndrome [CFS]). On the Neuropsychological Questionnaire at screening, If the answer to Delusions, Hallucinations, Bipolar disorder, Motor Disturbance, Suicidal thoughts question is “Yes”, it is an exclusion. If Depression was “Yes”, PHQ-9 questionnaire will appear. If the total score on PHQ-9 is above or equal to 15, it is an exclusion.<br>• Inability to follow study procedures.<br>• Physical, cognitive, or language impairments sufficient to adversely affect data derived from cognitive assessments.<br>• History of mild traumatic brain injury (TBI) within the past 6 months, or a lifetime history of moderate or severe TBI (e.g., loss of consciousness for >30 minutes or GCS=<13).<br>• Current unstable medical condition or significant disease that may affect efficacy or safety assessments, or any other reason which, in the investigator’s opinion, may preclude the subject’s participation for the full duration of the trial (e.g., uncontrolled diabetes mellitus or hypertension).<br>• Pregnant and/or breastfeeding.<br>• Participants who are taking medication should be on stable therapy or been taken at a stable dose for at least 4 weeks prior to study entry.<br>• Any medication, in the opinion of the investigator, may affect cognitive function and may affect efficacy or safety assessments<br>• Photosensitivity reactions to sunlight or visible light (polymorphous light eruption, solar urticaria, persistent light reactivity).<br>• History of recurrent epistaxis within the last 24 weeks or currently taking major anti-coagulants (including warfarin, low molecular weight heparin)<br>• Increased skin sensitivity at the treatment site including active herpes simplex in the treatment area, history of keloid formation, or history of retinoid use in the past month. |

©2023 Vielight Inc.
All rights reserved

BRANY
IRB APPROVED
08/24/2023

| | |
|---|---|
| Treatment | Vielight Neuro RX Gamma administered for 20 minutes once a day, for the first 6 of 7 days per week for 8 consecutive weeks (56 days). Banded near infrared LED module on the head and intranasal applicator inside the left or right nostril for the study duration |
| Study Design and Measures | ● Post COVID-19 participants will be pre-screened and consented through an electronic informed consent form (eConsent) via the electronic data capture system (EDC).<br>● Pre-existing health conditions, medication use, cognitive conditions, Fitzpatrick scale, race/ ethnicity, sex, and age are recorded during the screening period after informed consent. Eligible Participants will be randomized to sham or active Vielight Neuro RX Gamma (Treatment) Groups.<br>● Participants will complete an electronic shipping address form via the EDC during the screening period. Devices will be express couriered to randomized participants.<br>● Participants will complete the self-report measure of cognitive symptoms at baseline, Days 14, 28, 56 (end of treatment) and 84 (follow-up) via the EDC.<br>● Participants will use a daily diary via the EDC to record:<br>1) treatment administration (compliance)<br>2) complications related to the use of the device (e.g., nasal irritation)<br>3) technical complications (e.g., device not turning on).<br>4) change in medication or concomitant treatment<br>5) emergent medical condition requiring medical attention (safety)<br>6) hospitalization (safety)<br>7) adverse events |
| Primary Endpoint | Primary efficacy objectives include tests of working memory, executive function, attention, and processing speed. The primary efficacy endpoint is the change from baseline to Day 56 in the combined results of 7 Creyos items: Spatial Planning, Monkey Ladder, Rotations, Feature Match, Paired Associates, Token Search, and Polygons. The z-scores and percentiles relative to an age (decile) adjusted reference population will be performed.<br>. |

©2023 Vielight Inc.
All rights reserved

BRANY
IRB APPROVED
08/24/2023

| | |
|---|---|
| Secondary Endpoints | 1. The change from baseline in Creyos scores of the primary efficacy endpoint (7 tests) and secondary Creyos items (5 remaining tests) will be compared between treatments at each follow-up interval through Day 56 (i.e., Grammatical Reasoning, Spatial Span, Digit Span, Odd One Out, Double Trouble) and changes from baseline for Sham and Active treatment devices will be compared at each testing timepoint. The Creyos questionnaire is administered at baseline, 14, 28, 56 and 84.<br><br>2. EQ-5D-5L Quality of Life<br><br>The EQ-5D-5L questionnaire consists of 5 questions regarding mobility, self-care, usual activities, pain/discomfort, and anxiety/depression. Ordinal responses to each question are on a 5-point integer scale ranging from 1 (no problem) to 5 (extreme problem). A visual analog scale (VAS) ranging from 0 to 100, with 100 meaning the best health that you can imagine and 0 meaning the worst health that you can imagine. The questionnaire is administered at 0, 14, 28, 56, and 84 Days.<br><br>3. Fatigue Assessment Scale (FAS)<br><br>The FAS consist of 10 questions regarding memory and brain functioning. The maximum score is 50. The questionnaire is administered at 0, 56 and 84 Days.<br><br>4. Perceived Deficits Questionnaire (PDQ-20)<br><br>The perceived deficits questionnaire – 20 item version (PDQ-20) has 20 questions, and each is rated as never (0), rarely (1), sometimes (2), often (3), and almost always 4). The questions describe situations encountering problems with memory, attention and concentration occurring in the past week. The questionnaire is administered at 0, 56, and 84 Days. |

©2023 Vielight Inc.
All rights reserved

BRANY
IRB APPROVED
08/24/2023

| | |
|---|---|
| | 5. Compliance and Technical Complications<br><br>Daily diary checklists for the device include treatment administration, compliance, and technical complications. Compliance refers to wearing the device as prescribed for the recommended duration.<br><br>6. Modified Symptom Burden Questionnaire<br>Questions to capture if participants have the following symptoms:<br><br>Breathing<br><br>Pain<br><br>Fatigue<br><br>Memory, Thinking and Communication<br><br>Sleep<br><br>Ears, Nose and Throat<br><br>Stomach and Digestion<br><br>Other Symptoms<br>The questionnaire is administered at 0 and 56 Days.<br><br>7. Exploratory Endpoints<br><br>Statistical analysis of the Creyos changes between treatment groups may warrant further exploration of the primary endpoint model.<br><br>Subgroup analysis of the primary efficacy endpoint by the pre-specified age cohorts will be undertaken. The influence of age as a continuous variable will be explored in the statistical model.<br><br>The principal component analysis (PCA) analysis is exploratory in nature, and this approach is utilized to identify influential Creyos items from the original 12 items that explain the most of variability in the data. |

©2023 Vielight Inc.
All rights reserved

BRANY
IRB APPROVED
08/24/2023

| | |
|---|---|
| Statistical Methodology | The current study size will be informative as a pilot study conducted to evaluate changes in Creyos primary items and is not based on formal power calculations. A sample size of 36 enrolled subjects (18 active, 18 sham) will provide estimates of the mean and standard deviation for each treatment group, mean changes from baseline, and the mean difference between treatment groups in the change from baseline. The observed magnitude of treatment effect will inform future studies.<br><br>Descriptive statistics (n, mean, median, standard deviation, interquartile range (IQR), minimum, maximum, and interquartile range) will summarize continuous variables, and frequency distributions and percentages will summarize categorical variables.<br><br>The safety population (Safety) includes all participants who are randomized and receive the study device. The pharmacodynamic population (PD) includes safety subjects with a Creyos measurement at baseline and at least one post-baseline measurement. PD subjects will exclude those with significant protocol deviations with potential effect on efficacy as determined prior to unblinding.<br><br>Daily diary checklists for the device include treatment administration, compliance, and technical complications. Safety reporting includes changes in medication, adverse events, and hospital admissions. Adverse events will be classified according to the Medical Dictionary for Regulatory Affairs (MedDRA).<br><br>The primary endpoint is the mean percentile, z-score and observed Creyos cognitive tests at Day 56 from the 7 primary efficacy items in the PD population. Analysis of variance methods are used to test for treatment differences in the change from baseline to Day 56 (end of treatment) in the average of the 7 items in the primary set.<br><br>Adverse events will be classified by MedDRA as preferred terms within system organ class (SOC) and presented as treatment group frequencies from baseline through Day 56, from Day 56 to Day 84<br><br>Exploratory efficacy analyses utilize principal components analysis.<br><br>SAS (SAS Institute, Cary, NC, USA) will be used for statistical analysis. |

©2023 Vielight Inc.
All rights reserved

BRANY
IRB APPROVED
08/24/2023

### 2.0 Introduction

#### 2.1 Post COVID-19 Pathology and Study Focus

Some people who have been infected with the virus that causes COVID-19 can experience long-term effects from their infection, known as post COVID-19 conditions (PCC) or long COVID[1]. The medical circles often describe it as post-acute sequelae of Covid-19 (PASC). People with post-COVID conditions can have a wide range of symptoms that can last more than four weeks or even months after infection. Sometimes the symptoms can even go away or come back again. The Centers for Disease and Prevention (CDC) listed a constellation of 19 symptoms related to post COVID-19[1].

Based on Vielight's strength in neurological and cognition research, attention would be narrowed to the neurological aspect. CDC published findings on Post COVID-19 showed elevated risk ratios that include neurological conditions as significant[2]. See Figure 1.

©2023 Vielight Inc.
All rights reserved

BRANY
IRB APPROVED
08/24/2023

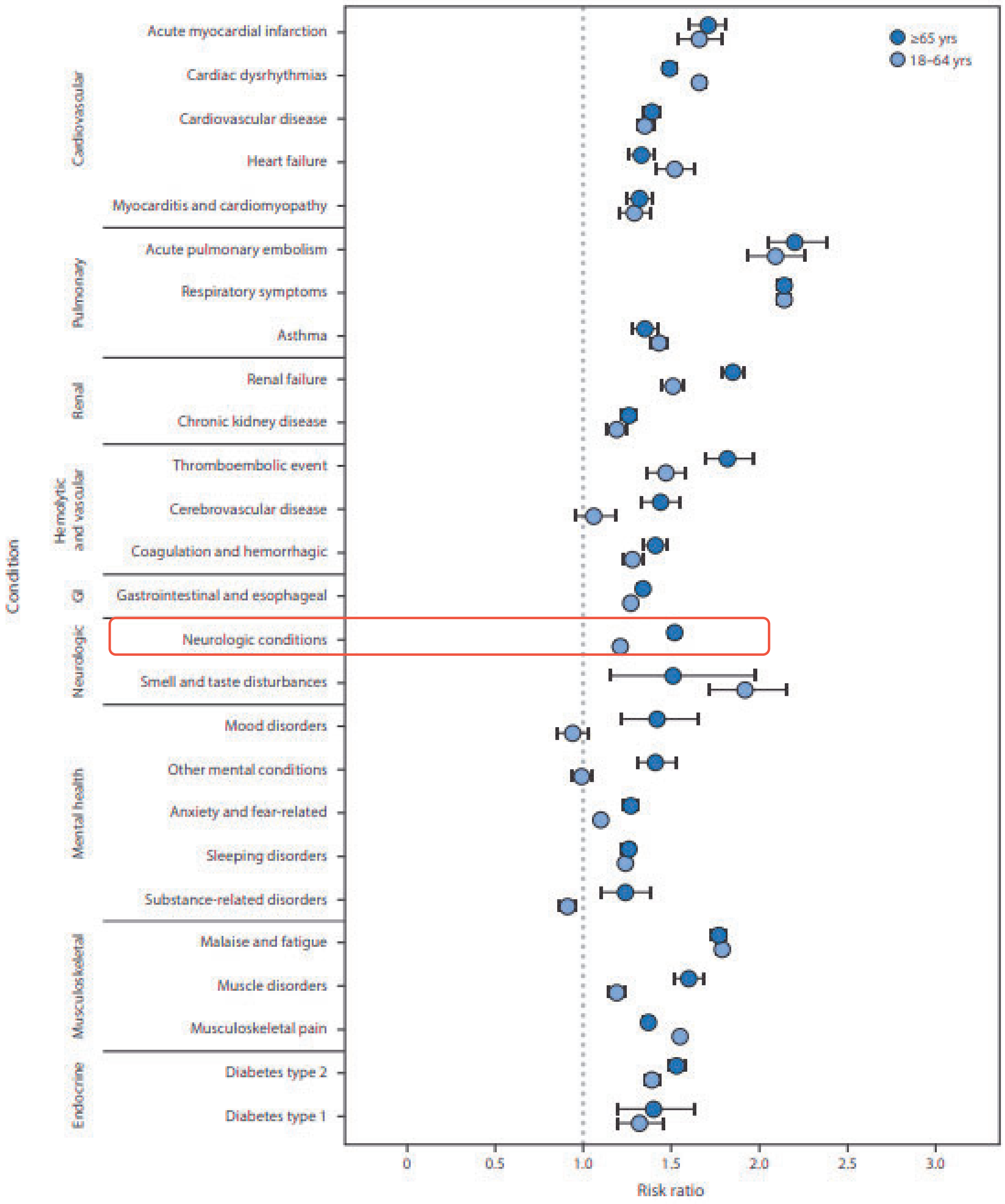

**Figure 1.** Risk ratios* for developing post COVID-19 conditions among adults aged 18–64 years and ≥65 years — United States, March 2020–November 2021

It has also been estimated that after 7 months after post-acute infection, more than 80% of the cohort with Post COVID-19 have brain fog[2] – see Figure 2.

©2023 Vielight Inc.
All rights reserved

BRANY
IRB APPROVED
08/24/2023

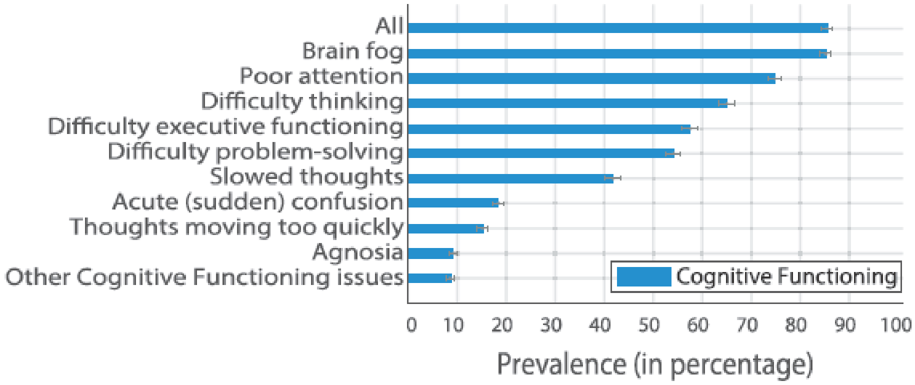

**Figure 2.** Cognitive Functioning Symptoms prevalence estimates Reproduced from Davis HE et al (available through creative commons attribution)[3]. Each bar represents the percentage of respondents who experienced that symptom. Error bars are bootstrap 95% confidence intervals.

In research, brain fog is prominent among the most reported neurological symptoms which also include, numbness, tingling, headache, dizziness, blurred vision, tinnitus, and fatigue that last more than a year post-infection[4.] Brain fog is defined in Section 2.2.1.

Combining the high prevalence of brain fog as a post COVID-19 symptom, and the success in cases of treating cognitive impairment with Photobiomodulation[5], Vielight Inc. designed this clinical pilot trial to assess the Vielight Neuro RX Gamma device on Brain Fog as endpoint targets.

#### 2.2 Investigational Device: The Vielight Neuro RX Gamma

##### 2.2.1 Design Principles

The Vielight Neuro RX Gamma (Figure 3) is a home-use photobiomodulation (PBM) device designed to deliver near-infrared (810 nm) light (or photons) to the brain/scalp and nasal tissues.

For this experimental trial, we will follow the definition that “brain fog” is a usually temporary state of diminished mental capacity marked by inability to concentrate or to think or reason clearly[6]. These are symptoms that are also identifiable with cases of “chemo brain” (cognitive impairment as the result of cancer therapies)[7, 8] and Alzheimer’s disease (which includes mid cognitive impairment)[9].

Brain fog has also been associated with myalgic encephalomyelitis/chronic fatigue syndrome (ME/CFS)[4], which is amongst the most prevalent symptom in post COVID-19. The pathogenesis of brain fog could be attributed to chronic low-grade inflammation[4] and abnormal biomarkers of inflammation[10]. The characteristics of chemo brain and inflammation could in turn be attributable to the overactivity of microglia[11].

Magnetic Resonance Imaging (MRI) has also shown that post COVID-19 subjects have abnormal brain structures noticeably in the limbic system. They are connected to the olfactory bulb, which could be the first altered structure in the infection that leads to reduced brain network connectivity and brain fog[12].

©2023 Vielight Inc.
All rights reserved
BRANY
IRB APPROVED
08/24/2023

The brain appears to compensate for deficits from olfactory dysfunction in post COVID-19 with higher intra-network connectivity in the default mode network (DMN)[13]. This suggests that additional support for DMN network functions would be helpful.

Other imaging research has shown that other than the DMN, there are higher connectivity between cerebellum, sensorimotor and visual networks, indicating post COVID-19 patients spend abnormally higher time in specific brain states compared to healthy controls[14]. There is also reduced functional connectivity within several layers of the vermal lobules in the cerebellum[15]. These lobules could be involved in cognition and emotion processing[16].

Brain fog in post COVID-19 has drawn similarities with "chemo-brain" (post-cancer therapy effects) explained by elevated levels of chemokines causing inflammatory microglia reactivity, particularly affecting the hippocampus[17]. NIR PBM has been shown to have the potential to reduce inflammatory markers relevant to COVID-19[18, 19], and when pulsed at 40 Hz can activate the non-inflammatory M2-genotype microglia to remove markers of Alzheimer disease, such as beta-amyloid and possibly tau deposits[20, 21].

On the assumption that red and NIR irradiation with the correct set of parameters correlate with desired outcomes, the location of the LEDs in the proper locations could be a consideration.

The Vielight Neuro RX Gamma (Figure 3) has the attributes to direct 810 nm NIR light to the desired anatomical regions:

1. Connect to the limbic system via the olfactory bulb.
2. LEDs to deliver NIR to the nodes of the DMN, the cerebellum and visual cortex to regularize their functional connectivity.
3. 40 Hz to activate M2 microglia to remove unwanted tau and beta-amyloid deposits, while reducing the risk of inflammation.
4. An LED on the confluences of sinuses to facilitate lymphatic drainage of beta-amyloid deposits – the same position as the primary visual cortex or $O_z$ according to the 10-20 EEG system.

In addition, the portability of the device is amenable for home-use, and the largely invisible 810 nm NIR allows for the potential of a sham/placebo device in a randomized clinical trial.

©2023 Vielight Inc.
All rights reserved

BRANY
IRB APPROVED
08/24/2023

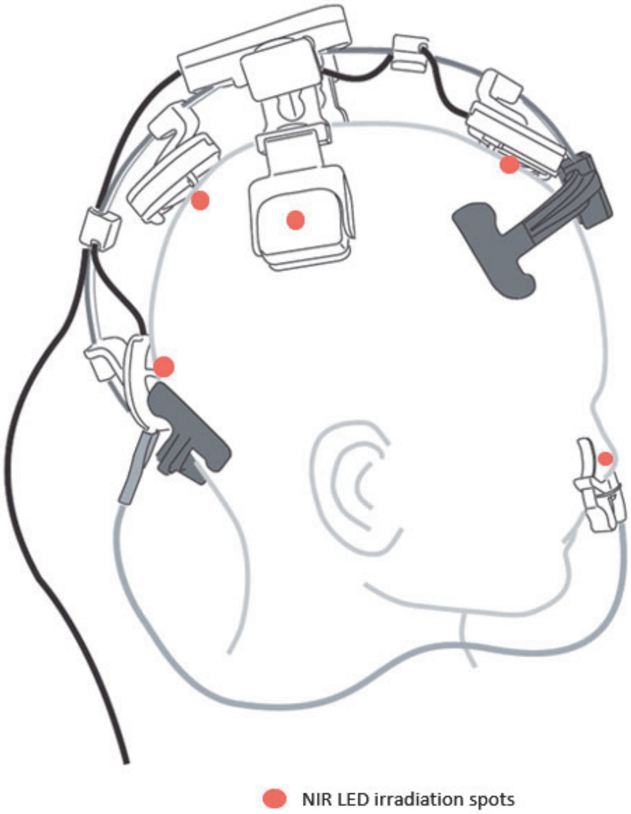

**Figure 3**: The Vielight Neuro RX Gamma with head LED and intranasal LED

##### 2.2.2 Device Description

The device is based on the science of photobiomodulation (PBM) and uses 6 light emitting diodes (LED) at a near-infrared (NIR) wavelength of 810 nm (Figure 4).

©2023 Vielight Inc.
All rights reserved

BRANY
IRB APPROVED
08/24/2023

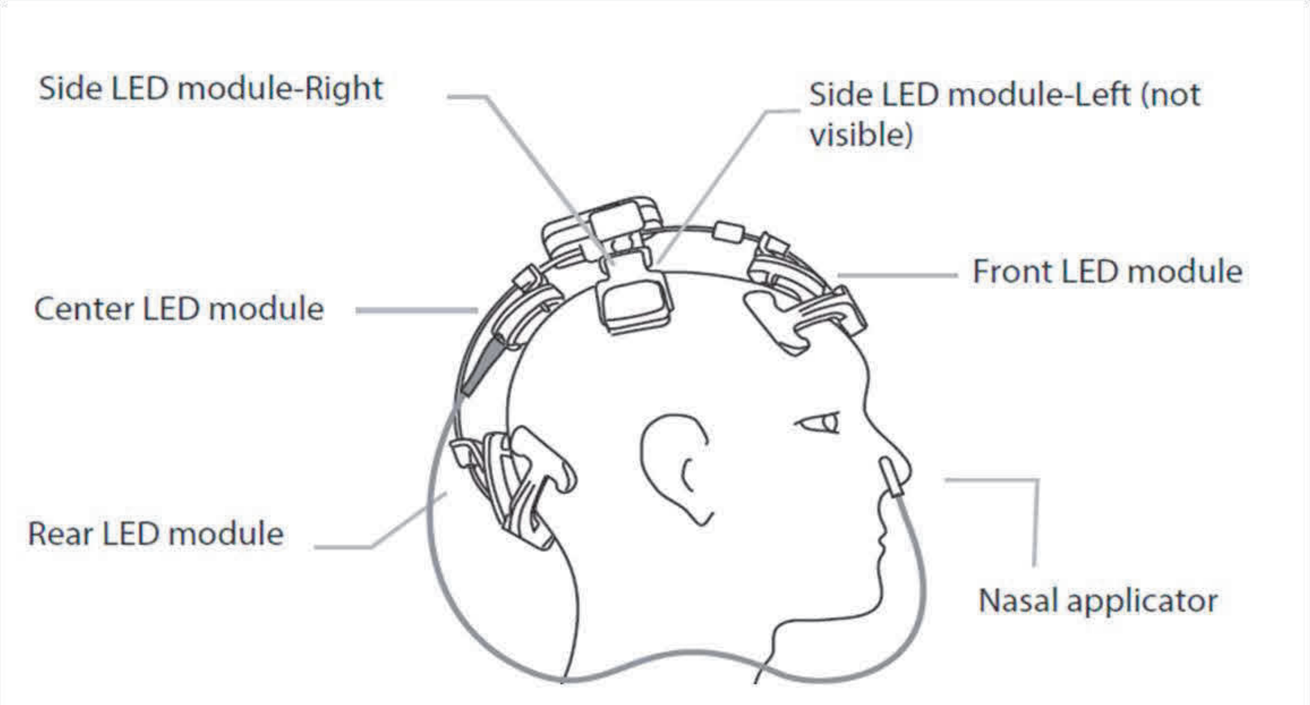

**Figure 4**: Diagram of the LEDs of the Neuro RX Gamma (v2)

Diodes are placed on the skull, held in position by stainless steel band, as well as intranasally. These LEDs target the hippocampal area, posterior cingulate cortex, medial prefrontal cortex and precuneus – the nodes of the default mode network (DMN), as well as the primary visual cortex and cerebellum. The power density of each transcranial LED diode is: 75 $mW/cm^2$ for the front LED, 100 $mW/cm^2$ each for the left and right-side LEDs and the center LED, and 50$mW/cm^2$ for the rear LED. The power density of the single intranasal LED diode is 25 $mW/cm^2$. All diodes are pulsed at a rate of 40Hz, 50% duty cycle, each with a beam spot size of 1 $cm^2$. The intranasal and front LEDs are anti-phase to the side LEDs (L&R) and rear LED. No significant heat is generated.

The sham is indistinguishable from the intervention to the participants. It has an optical sensor which prompts the device to switch off after the headset is in contact with the scalp for 3 seconds. If the device is switched on but not worn, it sounds out an alarm with regular beeps (one beep per second) until it is either placed on the head or switched off. If functioning properly, the device will sound out one beep every 10 seconds. The beep can be switched on and off by press and hold the start button for 3 seconds.

Both intervention and sham group participants will be treated in 20-minute sessions once daily for 6 consecutive days each week. Participants will have one day off treatment before resuming treatment for another 6 consecutive days. Participants will be taught to use the device and maintain a daily diary.

The Vielight Neuro RX Gamma device is designed to be portable and wearable. It can be used by the participants at home after training on how to use the device by trained Device Operators (i.e., study investigator or study team member).

©2023 Vielight Inc.
All rights reserved

BRANY
IRB APPROVED
08/24/2023

#### A. System Architecture

The Neuro RX Gamma device consists of a nasal applicator and a headset with built-in controller (Figure 5).

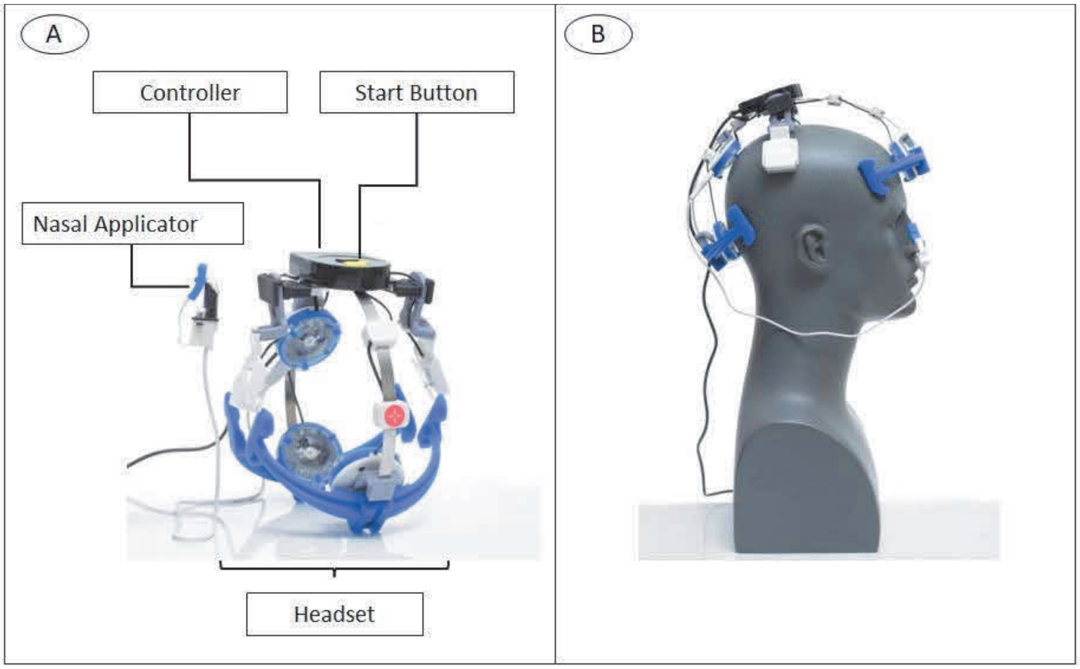

**Figure 5**. Vielight Neuro RX Gamma (v2). (A) Components of the device – the nasal applicator and headset with built-in controller are identified. (B) The device is shown on a mannequin head to demonstrate the placement of the six LEDs.

#### B. Headset with built-in controller

The headset is a wearable applicator consisting of 5 LEDs. A red star symbol identifies the front of the headset and helps the user orientate the headset on the head. Supports at the front and rear of the headset secure the device on the head. The front LED is placed centrally on the forehead at the hairline to target the medial prefrontal cortex. The center LED is placed over the precuneus, and the left and right-side LED over the left and right angular gyri to target the posterior cingulate cortex. The rear LED is placed over the primary visual cortex and cerebellum.

The built-in controller houses electronics for controlling the Neuro RX Gamma. To initiate a session, the USB-A plug (black cable) from the headset is connected to the medical grade power adapter which is then plugged into an electrical outlet. A steady green light at the power indicator confirms the flow of electric current to the device. Pressing the start button initiates a software controlled 20-minute therapy session which can be discontinued by pressing the start button a second time. The power indicator LED flashes green (visible by an observer) when therapy is being delivered. The circuitry in the controller manages the other LEDs with the specified parameters.

#### C. Nasal Applicator

©2023 Vielight Inc.
All rights reserved
BRANY
IRB APPROVED
08/24/2023

The nasal applicator plugs into the center LED. The other end of the nasal applicator consists of a single LED that is placed in the nostril and clipped into place. The nasal LED has an 810 nm wavelength and has an output power of 25mW delivered to a beam spot size of 1cm2.

##### D. Power Supply

A medical grade, certified DC supply is used to power the device.

##### E. Device Specifications

LED classification according to IEC 62471 Power Supply: 7.5 VDC, 1Amp

Maximum Power Output Density to Nasal Mucosa: 25 mW/cm2 @ 50% duty cycle

Total Energy Delivered to Nasal Mucosa: 15J.

Total Energy Delivered to Scalp (Headset): 255J.

Maximum Power Output Density to Scalp-Front LED: 75mW/cm2 @ 50% duty cycle = 45J.

Maximum Power Output Density to Scalp-Side (L&R) and Center LEDs: 100 mW/cm2 @ 50% duty cycle = 180J.

Maximum Power Output Density to Rear LED: 50mW/cm2 @ 50% duty cycle = 30J.

##### 2.2.3 Device Placement

The intranasal applicator is inserted into the nostril as far as is comfortable (Figure 6A). The small blue sleeve is added to the intranasal applicator clip to provide added grip if needed during treatment.

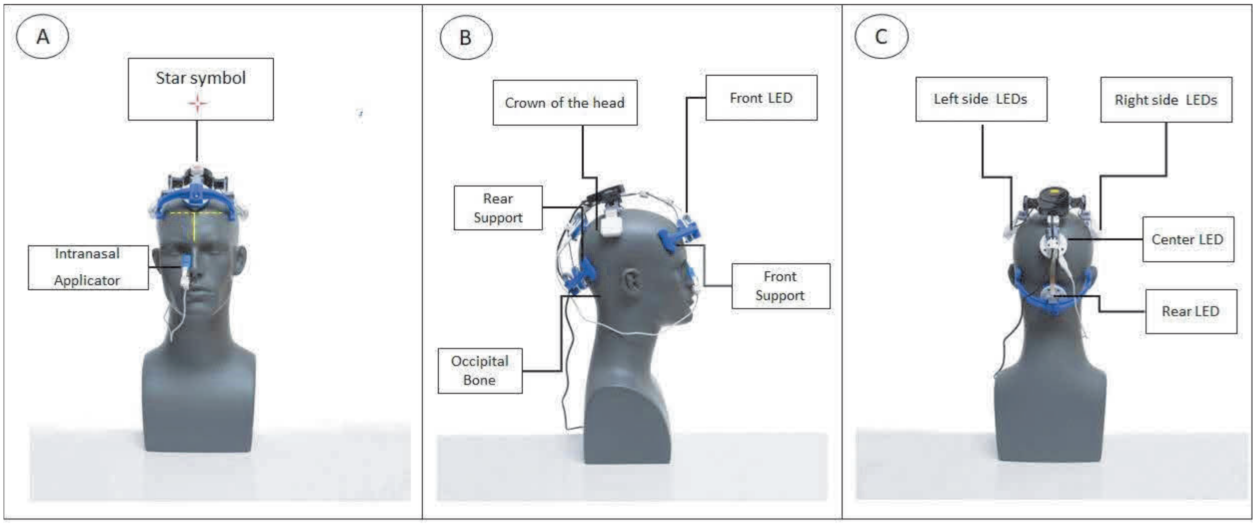

**Figure 6**. Placement of the Neuro RX Gamma (v2). (A) Orientation of the device on the head with the star symbol at the front. (B) Positioning of the Front LED and identification of markers to help guide placement of the center and rear LEDs. (C) Positioning of the Side, Center, and Rear LEDs.

©2023 Vielight Inc.
All rights reserved

The headset is placed on the head so that the red star symbol is at the front of the head (Figure 6A). Supports at the front and rear of the headset secure the device on the head (Figure 6B). The front LED is positioned midline of the face centered just above the natural hairline (Figure 6A&B). The center LED is placed on the crown of the head (between the top and flatter back portion of the head) (Figure 6B&C). The side LEDs (L&R) will automatically fall into the correct positions (Figure 6C). The rear LED is positioned at the back of the head where the head and neck connect (occipital bone) (Figure 6B&C). If the treatment session is initiated prior to proper placement of the headset, the device controller will beep continuously until the headset is correctly positioned.

###### 2.2.4 Procedure

Once the headset and nasal applicator have been positioned, the device must be connected to an electrical outlet to initiate the session. The black cable from the headset is plugged into a medical grade power adapter which is plugged into an electrical outlet. The power indicator shows a steady green light confirming the flow of electric current to the device. The operator initiates the treatment by pressing the start button on the controller (Figure 5A). The device proceeds to deliver light to each LED at 40 Hz with a 50% duty cycle for 20 minutes and stops automatically. During the 20-minute treatment the power indicator LED on the controller flashes green. The operator is notified that the treatment is finished when the power indicator LED on the controller stops flashing and the device beeps 2 times.

###### 2.2.5 Regulatory Status

The Vielight Neuro RX Gamma has been available in its current commercialized form as the "Vielight Neuro Gamma": a low-risk general wellness device with no medical claims attached. It has the status of a Class II medical device registered with the United States Food and Drug Administration (FDA) but has not yet been available commercially under this status.

##### 2.3 Mechanisms of Action of the Vielight Neuro RX Gamma

The fundamental mechanisms of PBM are based on the absorption of photons by the mitochondria to modulate cellular functions. The Vielight Neuro RX Gamma delivers light of specific wavelengths (810 nm), power and duration to the brain/nasal cavity to achieve this. The biological process involves numerous interacting mechanisms that modulate bodily functions[22]. One result of PBM is the benefits it could offer the post COVID-19 (long COVID) population[23, 24].

The Vielight Neuro RX Gamma emitting NIR might reduce inflammatory markers relevant to COVID-19[25, 26], and since it pulses at 40 Hz can activate the non-inflammatory M2-genotype microglia to remove markers of Alzheimer disease, such as beta-amyloid and possibly tau deposits[27, 28]. Using Vielight Neuro RX Gamma, the same activation of non-inflammatory markers might occur with post COVID-19 (long-COVID) patient population as well as the reduction in the brain fog. The positioning of Vielight Neuro RX Gamma over the head is shown in Figure 4.

©2023 Vielight Inc.
All rights reserved

BRANY
IRB APPROVED
08/24/2023

##### 2.3.1 The Systemic Effect of Irradiating the Free-Floating Mitochondria

The intranasal applicator could enhance the systemic effect of PBM through irradiating the free-floating mitochondria. The nasal cavity has been chosen to position an LED for access to the dense local blood capillary networks which are shielded by a very thin light-permeable membrane in the nasal mucosa. This makes it relatively easy for light from the nasal LED to reach the blood circulatory system. PBM has a body-wide systemic effect that could be mediated by free-floating mitochondria[29].

##### 2.3.2 Anti-inflammatory effect of PBM

Photobiomodulation (PBM), also known as Low Level Light Therapy (LLLT), was discovered by Ender Mester while he was trying to use a ruby laser to treat experimental tumors in Syrian hamsters[30]. Although, his laser power was not sufficient to treat the tumor, he observed that the laser treated group experienced an accelerated healing of wounds[31].

The fundamental mechanism thought to underly PBM is based on its effect on Cytochrome C Oxidase (CCO). CCO is the fourth unit of the mitochondrial respiratory chain. The absorption of photons by CCO causes nitric oxide photodissociation[32]. This event leads to increase in electron transport activity, an increase in adenosine triphosphate (ATP), the energy source of cells. Coincidently there is an increase in production of reactive oxygen species (ROS) by the mitochondria and the release of $Ca2+$ as versatile second messengers. In turn, this leads to the activation of transcription factors and signaling mediators (i.e., NF-κB), producing long-lasting cellular effects[22].

ROS play a central role in the progression of inflammation. During inflammatory conditions, ROS production causes an increase in the migration of inflammatory cells to the damaged tissue. ROS production is increased by polymorphonuclear neutrophils (PMNs) at the location of inflammation, in turn producing disfunction of endothelial cells and additional tissue injury. This migration results in more injury in inflamed tissue, pain and postpones healing process[33]. While PBM (wavelengths greater than 500-nm) increases the production of ROS in normal cells[34], it reduces markers of oxidative stress in stressed and damaged cells[22], thereby promoting faster healing.

In addition, PBM can suppress inflammation by reduction in PGE2- levels and inhibition of cyclooxygenase-2 (COX-2) in cell cultures[35]. In short, the appropriate dose of PBM directed to the injured tissue could positively control pain, reduce inflammation, and expedite healing. In addition, PBM can suppress inflammation by reduction in PGE2- levels and inhibition of cyclooxygenase-2 (COX-2) in cell cultures[35]. In studies, PBM has been shown to promote inflammation through NF-$_{K}$B proteins in normal cells. However, in the presence of excessive inflammatory markers, PBM has been shown to behave differently. It has been shown to be anti-inflammatory[36]. The anti-inflammatory characteristic of PBM is expected to calm a potential cytokine storm. In short, the appropriate dose of PBM directed to the injured tissue could positively control pain, reduce inflammation, and expedite healing.

#### 2.4 Clinical Data and Summary of Related Evidence to Date

Previously, Vielight Inc. conducted a randomized clinical trial on COVID-19 positive participants. Vielight Inc. using the lessons learned to offer a device-based option to treat post COVID-19. The device used in the study was the Vielight RX Plus, which delivered red and near infrared

©2023 Vielight Inc.
All rights reserved

BRANY
IRB APPROVED
08/24/2023

light (a process called PBM) intranasally and to the thymus gland and the lungs. The symptoms that responded positively in the RCT with statistical significance were sinus pain, chest congestion, body aches, think clearly, ear discomfort, sinus drainage, headache, coughing up stuff and sneezing. A large number of subjects affected involved “think clearly” or brain fog.

###### 2.4.1 Support recovery of damaged cells

PBM has established mechanisms in the production of growth factors to promote cellular regeneration[22].

###### 2.4.2 Management of inflammation

PBM has been shown to control inflammation generated through the activities of the cytokines mitigating the risk of a cytokine storm[36].

###### 2.4.3 Vielight devices have produced evidence of tissue responses

Vielight devices have demonstrated in peer-reviewed literature that tissues such as those in the brain respond to NIR. This is measured with electroencephalography (EEG)[37] and functional magnetic resonance imaging (fMRI)[38]. Numerous other studies have shown that different tissues have positive responses to PBM[22], which can potentially be related to a device such as the Vielight Neuro RX Gamma.

###### 2.4.4 Track record of efficacy in other clinical studies

PBM with Vielight devices were used in other clinical studies, producing positive clinical outcomes. These include positive clinical outcomes for dementia[38, 39] and traumatic brain injury[40]. These devices share the same platform as the Vielight Neuro RX Gamma proposed in this post COVID-19 study.

#### 3.0 Study Objective and Rationale

##### 3.1 Study Objective

The objective of this pilot study is to evaluate the efficacy of the Vielight Neuro RX Gamma in the treatment of post COVID-19 cognitive impairment. Comparisons of active and sham devices from this double-blind study will provide a measure of improvement based on the cognitive impairment instruments. Safety has been established previously. Adherence to protocol-specified instructions of use and compliance in wearing the device will be evaluated.

##### 3.2 Study Design Rationale

The study design is aimed at obtaining data to assess the efficacy of the Vielight Neuro RX Gamma in improving cognitive impairment in post COVID-19 population while recognizing the safety needs. With a single site and no in-person visits, this study is set up to occur virtually and based on self-report measures and online cognitive assessment tools.

©2023 Vielight Inc.
All rights reserved
BRANY
IRB APPROVED
08/24/2023

#### 4.0 Overview of Study Design

The study will be managed by an independent clinical research organization (CRO), supporting a Principal Investigator (PI) in the United States. Vielight Inc will supply the Vielight Neuro RX Gamma devices free of charge and will sponsor the study. The study is not dependent on the success of achieving external financial support.

A diagram of study flow is shown in Figure 7.

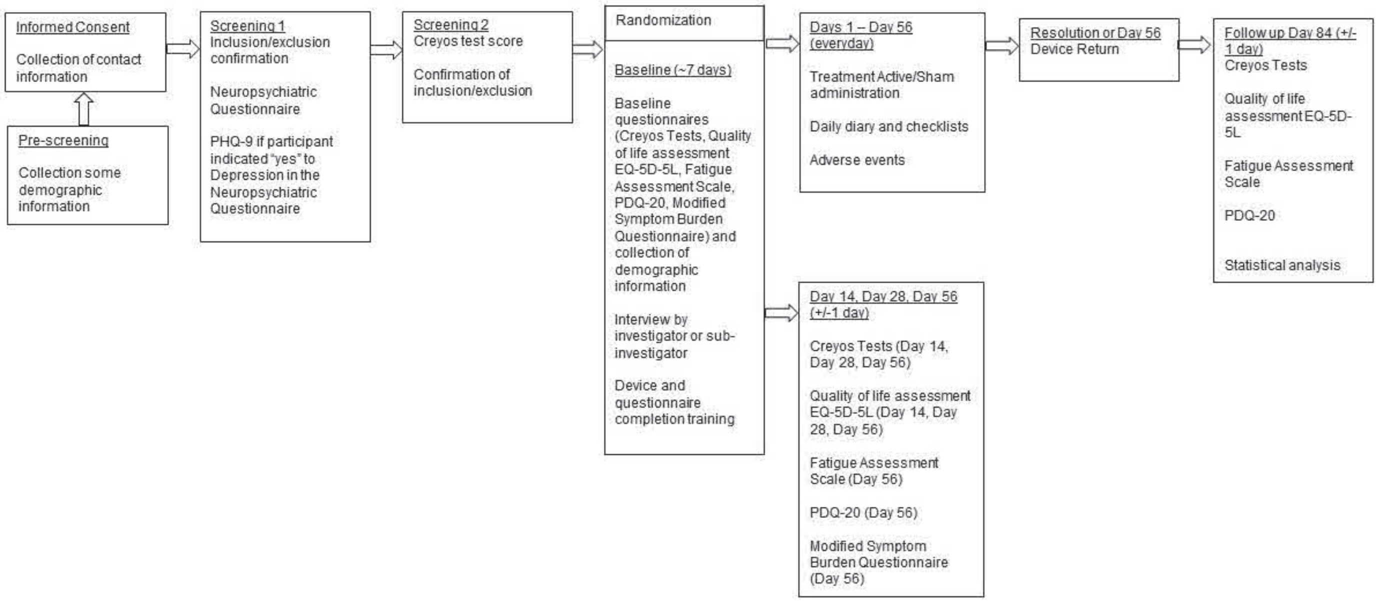

**Figure 7**. Study Flow Chart

#### 5.0 Study Procedures

The study information will be available on the study website. Potential participants will have access to the study landing page in which they provide their email address.

##### 5.1 Pre-Screening

Potential participants will receive an email to complete a screening form if they are interested in participating. The Uniform Resource Locator (URL) for the study website will be included on all recruitment methods. The pre-screening form will capture the following information:

1. Participant age range (18-30, 31-45, 46-65)
2. If they experience "brain fog" after being infected with COVID.
3. Daily access to an electronic device (laptop, desktop, tablet, cell phone) and internet (Yes or No).
4. If they have a physician who can be reached by the study team to verify their medical records.
5. If they can participate in a study that lasts for 12 weeks (about 38 hours time allocation)?
6. If they are pregnant.

The EDC will generate for each individual a unique Screening ID number and generate a Screening log. The Screening log will include email addresses information. The Screening log will

©2023 Vielight Inc.
All rights reserved

BRANY
IRB APPROVED
08/24/2023

have access restricted to only study staff, ensuring confidentiality is strictly maintained. The preliminary eligibility of potential participants is assessed by study staff at this level.

##### 5.2 Electronic Informed Consent Form and Emergency Contact

Once initial eligibility is assessed at pre-screening, potential participants will receive an email link to the informed consent form module (eConsent module within the EDC) which will include review of study procedures, overview of the medical device, their rights and responsibilities and all required aspects of the ICF. Potential participants will have access to the study team email and phone number to contact study staff if they have any questions regarding the device and/or study procedures. Study staff will contact the potential participant to discuss any questions and will document that all participant questions have been answered.

A difference platform is used for eConsent, where participants will create an account and the electronic informed consent form will be completed prior to completing study procedures. The ICF used under this study protocol will be consistent with applicable elements of ISO 14155: 2011 Good Clinical Practice Guidelines and the and 21 CFR Part 50 Protection of Human Subjects.

Subjects who have signed the ICF will be given a unique subject number.

After the ICF is reviewed and countersigned by the study investigator/ study staff, a record is created in the Real Study in EDC and a record ID is assigned to the eligible participant that shall be used to identify the subject on all study related documents.

After the eConsent is signed by the Participants, a separate email will be sent with a link to an EDC form to capture Participant's mailing address, emergency contact information (This information will be used to reach out to the emergency contact in the event that the subject stops responding to questionnaires and study staff are unable to contact the participant to conduct a safety assessment) and their doctor's contact information.

Study information, training materials and ICF will reiterate that the participant is responsible for their health, that Vielight Inc., and study staff are not monitoring their health, and that they are instructed to follow local public health guidance for seeking medical assistance and care.

##### 5.3 Screening Period

Upon providing informed consent, participants will receive a link from the EDC where they are screened for behavioural issues using a Neuropsychological questionnaire. Participants would fail the screening if they answered yes to any of the questions (i.e., Delusions, Hallucinations, Bipolar disorder, Motor Disturbance, Suicidal thoughts) except Depression/Dysphoria. If Depression was "yes", PHQ-9 questionnaire will appear. If the total score on PHQ-9 is above or equal to 15, it is an exclusion. Successful completion of the Neuropsychological questionnaire is followed by Medical history and concomitant medications usage, as well as the date the participant tested COVID-19 positive. Eligibility will be reviewed by study team members and verified with participant's doctor to confirm inclusion/exclusion criteria prior to randomization. The Creyos online test will be sent to participants at the screening. We have the ability to resend the questionnaires through the EDC as reminders but will need to call or email subjects to remind them to complete Creyos.

©2023 Vielight Inc.
All rights reserved

BRANY
IRB APPROVED
08/24/2023

#### 5.4 Registration and Randomization

This is a Double-blind, Sham-controlled, Prospective, parallel, randomized study. Subjects who provide informed consent and pass the screening will be registered on the enrollment log via the EDC with a unique Study ID.

Study staff will complete a randomization list that was prepared using SAS (SAS, Inc., Cary NC) software. Participants will be stratified by age (>=45 years, < 45 years) at baseline and randomly allocated to the Treatment group or Sham group (1:1) which aims to reduce the differences in the two groups based on baseline factors.

The study staff will contact participants, letting them know that the participant has been randomized, and inform them about the device shipment. At the same time the participant is asked to schedule the training/interview with the sub-investigator for a time after the participant has received the device.

#### 5.5 Shipment of Devices

Vielight Neuro RX Gamma device will be couriered to participants within 24 to 48 hours of randomization. Participants will be provided with a shipment notification, tracking information and will confirm receipt of the device via the EDC.

##### 5.5.1 Device Training

Device training for the Vielight Neuro RX Gamma device will be provided by the study staff with participants via a telemedicine platform call prior to initiating intervention procedures. On the same call, baseline information will be reviewed. Instructional materials will also be included with each device shipment. There will also be instructions for how to return the device at the end of the study. Participants are asked to fill a daily diary and records any issues with the device. They also have access to the study phone number and email address to inquire about how to use the device and deal with potential issues.

Figure 8 shows participants tasks throughout the study. The following section will explain each activity after participant's successful screening phase.

©2023 Vielight Inc.
All rights reserved

BRANY
IRB APPROVED
08/24/2023

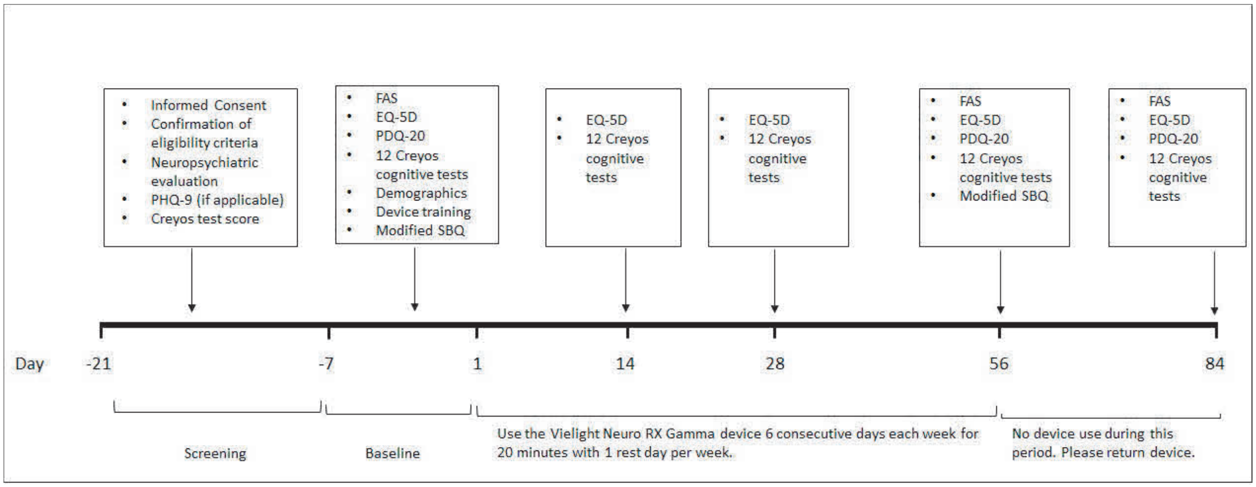

**Figure 8**. Diagram showing participant tasks throughout the study.

##### 5.6 Baseline (Day 0)

All participants will complete the following baseline forms via EDC:

- FAS
- EQ-5D
- PDQ-20
- Modified Symptom Burden Questionnaire
- Race/ ethnicity questionnaire
- Fitzpatrick scale (It is a scale to capture skin tone in addition to the ethnicity and race self-report)

A full set of 12 Creyos cognitive tests will be sent to participants subsequently. CRO will pre-register the participants and will create customized Creyos links for each participant, to be completed at each time point. The links to Creyos cognitive tests will be sent to participants via email.

##### 5.7 Days 1 to Day 56

Participants will be instructed to complete the 12 Creyos cognition tests at Day 1, Day 14, Day 28, and Day 56 at around the same time of day (exact time and date recorded) to have consistency of readings. A window of 24 hours is allowed for participants to complete the 12 Creyos cognition tests, but the tests should be done in one sitting and will expire if inactive after 1 hour. It will be recommended that the participants take the Creyos tests at a constant time of the day preferably starting in the afternoon. Furthermore, participants will be instructed to complete a daily diary to capture adverse events and device issues.

On Day 14 and Day 28, participants will also receive EQ-5D questionnaire via EDC. On Day 56, participants will receive EQ-5D, FAS, PDQ-20, and Modified Symptom Burden Questionnaire via EDC.

©2023 Vielight Inc.
All rights reserved
BRANY
IRB APPROVED
08/24/2023

The primary endpoint for the subject is completed at Day 56 at the end of study device administration, or when there is discontinuation because of emergency hospitalization, death, or a voluntary withdrawal by the subject.

During the study, research staff will be available during normal business hours to provide any requested assistance. A contact number (24 hour) will be provided to the participant for any safety events.

A daily diary will be sent to participants every day from Day 1 to Day 56 through the EDC. Participants are asked to record their device usage, time of the device use, if they had any adverse events, if they experienced any adverse events due to the device usage, if they have started, or stopped a medication, or had a medication dose changed.

If a participant does not complete the electronic Cognition assessment tool as well as other required questionnaires, the subject will be sent a notification from the EDC. Study staff will also contact them via phone call within 24 hours to confirm that the participant is aware of the study tasks, and whether any safety event(s) have occurred. If the participant is not reachable, study staff will attempt to contact the listed emergency contact to confirm the participants' safety. If a safety event has occurred, study staff will speak with the participant or emergency contact to obtain any details regarding this event.

In the rare event that the study device is not working properly, a replacement treatment device will be provided. In this instance, the study blind would be maintained by communication of the replacement device serial number to the study personnel.

##### 5.8 Final day of device use, Day 56

Day 56 is the final day of device use and final day for daily self-reporting. Participants will be given instructions on how to return devices.

##### 5.9 Between Day 56 and Day 84

During this period, participants will not use the device.

##### 5.10 Final, Day 84

Day 84 is the final follow up day on the electronic Cognition assessment tool and data collection by subject. On Day 84, participants will receive EQ-5D, FAS, and PDQ-20 via EDC.

##### 5.11 Subject Withdrawal

Participants may be withdrawn from the study for any of the following reasons:

1. Subject's choice to withdraw their consent to participate. Any participant has the right to withdraw from the study at any time and for any reasons without prejudice to future medical care by the physician or the institution. This will be considered a screening failure if they withdraw from the study prior to being randomized. Participants who withdraw consent after randomization will have any available data evaluated, up to the time of withdrawal.
2. Investigator's choice to withdraw the subject from the study. This will be considered a screening failure if the Investigator withdraws the subject prior to randomization.
3. The study may also be terminated for either administrative or safety reasons.

©2023 Vielight Inc.
All rights reserved
BRANY
IRB APPROVED
08/24/2023

For participants who withdraw their consent for study participation, the study team will complete the “End of Study CRF” and no additional data will be collected after the date of their discontinuation.

Participants will be instructed to notify the study team if they become pregnant during the study. Participants will be withdrawn if they become pregnant and recorded using the ‘End of Study CRF’.

##### 5.12 Participants data flow from the landing page to eConsent and EDC

The email address of the participant would be captured on the landing page. This email address will flow from the landing page and stored in eConsent Screening study and EDC. A screening ID will be assigned to this participant record in eConsent and a record ID will be assigned in EDC. An email goes out to the participants to fill out the survey. When the participant fills out the survey, the data is stored in EDC and is accessible to all the study team members who have survey access privileges. Once the participant is found eligible, the subject will be presented with ICF to sign. While creating their account in eConsent, their data will be stored in eConsent which will be visible to the study investigators.

A Screening ID is assigned to the record in eConsent and Subject ID is assigned in the Screening Study. After the ICF is reviewed and countersigned by the study team member, a record is created in the Real Study in EDC and a record ID is assigned to the eligible participant. Once the ICF is duly signed a record is created in the real EDC study. A record ID is assigned to the participants and all the data within the real EDC study is visible to the study team members who have survey access privileges. The data is to be stored on the EDC for 25 years.

##### 5.13 Treatment Protocol

The Vielight Neuro RX Gamma is administered for 20 minutes once a day, for 6 days skipping day 7 for 56 days. It is a banded near infrared LED module on the head and an intranasal applicator inside the left or right nostril for the study duration.

#### 6.0 Eligibility Criteria and Recruitment

#### 6.1 Recruitment Strategy

Potential subjects could be recruited utilizing a variety of recruitment methods, including advertisements (e.g., brochures and posters at medical clinics), a study website, social media, and physician referrals.

#### 6.2 Inclusion Criteria

Only subjects with the following characteristics are eligible for study entry:

- Age 18-65
- Meets WHO-defined post-COVID-19 condition (WHO definition: ‘'Post COVID-19 condition occurs in individuals with a history of probable or confirmed SARS-CoV-2 infection, usually 3 months from the onset of COVID-19 with symptoms that last for at least 2 months and cannot be explained by an alternative diagnosis. Common symptoms include fatigue,

©2023 Vielight Inc.
All rights reserved

shortness of breath, cognitive dysfunction but also others (more information is found here) and generally have an impact on everyday functioning. Symptoms may be new onset following initial recovery from an acute COVID-19 episode or persist from the initial illness. Symptoms may also fluctuate or relapse over time. To ensure the above criteria is met, participants will only be included in the study if they meet all eligibility criteria more than 12 weeks from the onset of their acute Covid-19 symptoms or positive PCR/antigen test.

- Meet Physician's diagnosis of Cognitive Dysfunction.
- Documented history of SARS-CoV-2 infection (positive PCR/antigen test during acute illness OR clinical diagnosis by physician during or after the acute illness).
- 
- Ability to provide informed consent.
- Ability to read and communicate in English

#### 6.3 Exclusion Criteria

Subjects with the following characteristics are not eligible for study entry:

- Current symptoms are explained by a psychiatric or neurological disorder (e.g., major depressive disorder or bipolar disorder).
- Having or history of any major neurological or psychiatric illness, and such conditions that may cause cognitive impairment, or symptoms like those seen in post-COVID-19 condition (e.g., mild, or major neurocognitive disorder, Lifetime psychotic episodes, bipolar disorder, active suicidal ideation, and homicidal ideation, or diagnosis of chronic fatigue syndrome [CFS]). On the Neuropsychological Questionnaire at screening, If the answer to Delusions, Hallucinations, Bipolar disorder, Motor Disturbance, Suicidal thoughts question is "Yes", it is an exclusion. If Depression was "Yes", PHQ-9 questionnaire will appear. If the total score on PHQ-9 is above or equal to 15, it is an exclusion.
- Inability to follow study procedures.
- Physical, cognitive, or language impairments sufficient to adversely affect data derived from cognitive assessments.
- History of mild traumatic brain injury (TBI) within the past 6 months, or a lifetime history of moderate or severe TBI (e.g., loss of consciousness for >30 minutes or GCS=<13).
- Current unstable medical condition or significant disease that may affect efficacy or safety assessments, or any other reason which, in the investigator's opinion, may preclude the subject's participation for the full duration of the trial (e.g., uncontrolled diabetes mellitus or hypertension).
- Pregnant and/or breastfeeding.

©2023 Vielight Inc.
All rights reserved

BRANY
IRB APPROVED
08/24/2023

- Participants who are taking medication should be on stable therapy or been taken at a stable dose for at least 4 weeks prior to study entry.
- Any medication, in the opinion of the investigator, may affect cognitive function and may affect efficacy or safety assessments
- Photosensitivity reactions to sunlight or visible light (polymorphous light eruption, solar urticaria, persistent light reactivity).
- History of recurrent epistaxis within the last 24 weeks or currently taking major anti-coagulants (including warfarin, low molecular weight heparin)
- Increased skin sensitivity at the treatment site including active herpes simplex in the treatment area, history of keloid formation, or history of retinoid use in the past month.

#### 7.0 Study Endpoints

##### 7.1 Primary Endpoint

The primary efficacy endpoint is the change from baseline to Day 56 in the combined results of 7 Creyos items: Spatial Planning, Monkey Ladder, Rotations, Feature Match, Paired Associates, Token Search, Polygons.

Creyos tasks for assessing change in neurocognitive functions are taken at different timeframes [Time Frame: at baseline, Day 14, Day 28, Day 56, and Day 84].

##### 7.2 Secondary Endpoints

- Creyos scores – Creyos scores of the 5 remaining tests (i.e., Grammatical Reasoning, Spatial Span, Digit Span, Odd One Out, Double Trouble) and changes from baseline for Sham and Active treatment devices will be compared at each testing timepoint.

The Cryos 12-item test is administered at 0, 14, 28, 56 and 84 days.

- EQ-5D-5L Quality of Life

The EQ-5D-5L questionnaire consists of 5 questions regarding mobility, self-care, usual activities, pain/discomfort, and anxiety/depression. Ordinal responses to each question are on a 5-point integer scale ranging from 1 (no problem) to 5 (extreme problem). A visual analog scale (VAS) ranging from 0 to 100, with 100 meaning the best health that you can imagine and 0 meaning the worst health that you can imagine. The questionnaire is administered at 0, 14, 28, 56 and 84 Days. See Appendix 3.

- Fatigue Assessment Scale (FAS)

The FAS consists of 10 questions regarding memory and brain functioning. The maximum score is 50. The questionnaire is administered at 0, 56 and 84 Days. See Appendix 4.

©2023 Vielight Inc.
All rights reserved

BRANY
IRB APPROVED
08/24/2023

- PDQ-20

The perceived deficits questionnaire – 20 item version (PDQ-20) has 20 questions, and each is rated as never (0), rarely (1), sometimes (2), often (3), and almost always 4). The questions describe situations encountering problems with memory, attention and concentration occurring in the past week. The questionnaire is administered at 0, 56 and 84 days. See Appendix 5.

- Compliance and Technical Complications

Daily diary checklists for the device include treatment administration, compliance, and technical complications. Compliance refers to wearing the device as prescribed for the recommended duration.

- Modified Symptom Burden Questionnaire

Questions to capture if participants have the following symptoms administered on 0 and 56 days. (See Appendix 6):

Breathing

Pain

Fatigue

Memory, Thinking and Communication

Sleep

Ears, Nose and Throat

Stomach and Digestion

Other Symptoms

- Exploratory Endpoints

Statistical analysis of the Creyos changes between treatment groups may warrant further exploration of the primary endpoint model.

Subgroup analysis of the primary efficacy endpoint by the pre-specified age cohorts may be undertaken. The influence of age as a continuous variable will be explored in the statistical model.

The principal component analysis (PCA) analysis is exploratory in nature, and this approach is utilized to identify influential Creyos items from the original 12 items that explain the most of variability in the data.

#### 8 Statistical Methods

The current study size is informative as a pilot study conducted to evaluate changes in Creyos primary items and is not based on formal power calculations. A sample size of 36 subjects (18

©2023 Vielight Inc.
All rights reserved

BRANY
IRB APPROVED
08/24/2023

active, 18 sham) will provide estimates of the mean and standard deviation for each treatment group, mean changes from baseline, and the mean difference between treatment groups in the change from baseline. The observed magnitude of treatment effect will inform future studies.

Descriptive statistics for continuous variables include the number of non-missing observations (n), the mean, standard deviation (SD), interquartile range (IQR), median, minimum, and maximum values. As appropriate, 95% confidence intervals and standard errors are provided. Frequency distributions with percentages based on non-missing categorical variables will be presented.

Analyses and summary statistics will be provided for each specified time point. Additionally, to explore the course of treatment effects over time, tables of summary statistics and/or plots of means (or proportions) with standard error bars will be provided. As appropriate, 95% confidence intervals will be provided.

Statistical significance of the primary efficacy endpoint is tested at two-sided p=0.05. SAS (SAS Institute, Cary NC) will be the main statistical package used for statistical analysis and reporting. Confidence intervals and p-values provide an estimate of the magnitude of treatment effect while recognizing possibly low statistical power. Other statistical software packages may also be used as necessary.

Due to the small sample size, implementation ANOVA models may present model assumption violation or estimate imprecision, in which case a two-sample t-test and/or Wilcoxon rank sum test may be utilized.

Reported here are the patients who were enrolled, and any screen failure data are not presented in this report. The SAP will be finalized following the sponsor review and signoff and prior to the statistical analysis of the study data.

The safety population (Safety) includes all participants who are randomized and receive the study device. The pharmacodynamic population (PD) includes safety subjects with a Creyos measurement at baseline and at least one post-baseline measurement with no significant protocol deviations as determined prior to unblinding.

Daily diary checklists for the device include treatment administration, compliance, and technical complications. Safety reporting includes changes in medication, adverse events, and hospital admissions. Adverse events will be classified according to the Medical Dictionary for Regulatory Affairs (MedDRA).

Summary statistics for the change from Day 56 (end of treatment) to Day 84 (follow-up).

SAS (SAS Institute, Cary, NC, USA) will be used for statistical analysis.

##### 8.1 Randomization and Stratification

Randomization is used to avoid bias in the assignment of subjects to treatment and to increase the likelihood that known and unknown subject characteristics (e.g., demographic and baseline characteristics) are evenly balanced across treatment groups, and to enhance the validity of statistical comparisons between treatment groups.

Stratification prior to randomization is introduced to control sources of variation related to or assumed to be related to the outcome. The balance is accomplished to ensure equal treatment assignment ratio (1:1). The primary endpoint is believed to be influenced by age, with age inversely related to improvement in clinical study report (CSR) results. Subjects are randomized

©2023 Vielight Inc.
All rights reserved
BRANY
IRB APPROVED
08/24/2023

within strata formed by age categories < 45 years of age and >= 45 years. As the numbers enrolled in each stratum is not known in advance of the study or influenced by the stratification process, stratification does not restrict the number of enrolled subjects in both age stratum and ensures treatment group balance within aga strata.

##### 8.2 Blinding

Blinding is intended to limit the occurrence of conscious and unconscious bias in the conduct and interpretation of a clinical trial arising from the influence which the knowledge of treatment may have on the recruitment and allocation of subjects, their subsequent care, the attitudes of subjects to the treatments, the assessment of endpoints, the handling of withdrawals, the exclusion of data from analysis, and so on[41].

This clinical study will follow a double-blinded approach in which the study treatment identity of the sham device and the Vielight Neuro RX Gamma device is masked and unknown to the study subjects and those involved in the conduct of the trial. Any coded treatment name (e.g., A, B) is also masked in this study.

If participant's enrollment is unblinded by accident and the treatment group assignment revealed to the study PI during the conduct of the study, the sub-investigator will follow up with the participant instead of the PI.

##### 8.3 Efficacy Endpoint Analysis

The PD population is used for the efficacy endpoint analysis.

###### 8.3.1 Primary Endpoint

The primary endpoint is the mean percentile, z-score and observed mean Cryos cognitive test score at Day 56 from the 7 primary efficacy items among 12 in the PD population. Analysis of variance methods are used to test for treatment differences in the change from baseline to Day 56 (end of treatment) n the average of 7 items comprising the primary Creyos set. These items include spatial planning, monkey ladder, rotations, feature match, paired associates, token search, polygons. Each item is normalized by subtracting the mean and dividing it by the standard deviation, and this ratio is compared to the standard normal distribution to generate a z-score and a percentile of the distribution.

The primary efficacy endpoint will be by a mixed model repeated measures (MMRM) analysis of covariance. The change in baseline to Day 56 will be compared between treatments by MMRM with model terms for treatment, day, treatment-by-day interaction, age (years) and baseline covariate.

Exploratory analysis of the primary efficacy endpoint will be by principal component analysis (PCA) of the Creyos items as sources of model variability. The analysis process is to form linear combinations of the original observed Creyos items such that the first order component accounts for the largest variance in the data and subsequent components accounting for decreasing variability in the data.

©2023 Vielight Inc.
All rights reserved
BRANY
IRB APPROVED
08/24/2023

##### 8.3.2 Secondary Endpoints

###### 8.3.2.1 EQ-5D-5L Quality of Life

The EQ-5D-5L questionnaire is provided in Appendix 3. The ratings of each of the 5 questions ranges from 1 (best condition) to 5 (worst condition) for mobility, self-care, usual activities, pain/discomfort, and anxiety/depression. The mean score for each question and the average score over the 5 questions will be used for statistical analysis. Treatments will be compared in the change from baseline in scores using a mixed model repeated measures (MMRM) analysis of covariance. The change from baseline at Day 14, Day 28, and Day 56, are utilized for analysis. Mean changes from Day 56 to Day 84 will be presented by treatment group.

###### 8.3.2.2 FAS

The FAS consist of 10 questions regarding memory and brain functioning. The maximum score is 50. Treatments will be compared in the change from baseline in scores using ANCOVA with model terms representing treatment, day, treatment-by-day interaction, age, and baseline covariate.

Mean changes in FAS from Day 56 to Day 84 will be presented.

###### 8.3.2.3 PDQ-20

The PDQ-20 questionnaire is administered at baseline, Day 56, and Day 84. An analysis of covariance will be used to compare treatments in the change from baseline to Day 56in average PDQ-20 score by an analysis of covariance (ANCOVA). Model terms include treatment, age, day, treatment-by-day interaction and baseline score as covariate.

Mean changes in PDG-20 from Day 56 to Day 84 will be presented.

###### 8.3.2.4 Secondary Creyos item

A MMRM analysis of covariance will be used to evaluate the mean of 5 secondary Creyos items from the 12 test items. The secondary Creyos items include grammatic reasoning, spatial span, digit span, odd one out, and double trouble. The dependent variable is the mean score (observed, z-score, percentile) and model terms include treatment, age, day, treatment-by-day interaction and baseline score.
Mean changes in secondary Creyos items from Day 56 to Day 84 will be provided.

###### 8.3.2.5 SBQ

An ANCOVA model will be used to evaluate treatment differences in SBQ from baseline to Day 56. Terms in the model include treatment, age, day, treatment-by-day interaction and baseline SBQ as covariate.

Mean changes in SBQ from Day 56 to Day 84 will be presented.

©2023 Vielight Inc.
All rights reserved

BRANY
IRB APPROVED
08/24/2023

##### 8.3.3 Compliance

Daily diary checklists are collected from study subjects and include treatment administration, compliance, and any technical complications with the device. Compliance will be defined as the percentage of days in which the device was worn for the recommended duration and as per subject instructions. Compliance will be compared between treatment groups using a two-sample t-test.

##### 8.3.4 Exploratory Endpoints

Exploratory efficacy analyses will be conducted to understand the influence of statistically significant explanatory variables (e.g., age) and in subgroups of interest.

For the primary efficacy endpoint, the influence of the individual scores among the 12-items in the Creyos instrument will be undertaken. A principal components analysis will be used to explore the identity and number of factors that contribute the most of model variability using orthogonal contrasts. An Eigen value of at least 1 will be used to identify potentially influential Creyos items. Creyos item factors identified by PCA will be used in a MMRM analysis to compare treatments, similarly to the analysis of the primary efficacy endpoint.

#### 8.4 Assessment of Device Safety: Incidence of device attributable adverse events and usability

The commercial version of the Vielight Neuro RX Gamma has been commercially available since 2016, with no report of any major side effects, other than the occasional report of a warm feeling from the 810 nm LED targeted for placement over head. It has the same controller and driver platforms as the Vielight Neuro RX Gamma which has been tested for safety and approved for a clinical trial by Health Canada. The Vielight Neuro RX Gamma version 2 being used in this study has a 5th LED on the headset but likely poses little additional safety risk as several customers have simultaneously used the Vielight Neuro RX Gamma with our Vielight RX Plus device to mimic a similar setup as the Vielight Neuro RX Gamma version 2 without any added incidents. Unrelated to safety, the other factor that may arise is a malfunction of the device, in which case the CRO will liaise with the Sponsor who will take corrective action. Notwithstanding the anticipated absence of major issues, a mechanism is in place for the subjects to report an adverse event to the CRO, who will document it and report to the Sponsor for further corrective action.

A device-related event is determined by the "relatedness score" for Adverse Events. A relatedness score of "Possible" or "Probably" or "Definite" or "not related" indicate the extent that the Adverse Event is related to the use of the device. Adverse Events receiving a relatedness score of "Unrelated" or "Unlikely" are considered unrelated to the device use and are not included. All safety endpoints will be adjudicated by one of the study administrators.

A statistical comparison of the incidence rates will be conducted. Additionally, an evaluation of any events due to the general nature of the device (i.e., headset discomfort, nasal irritation) and not specific to the delivery of the NIR energy will be qualitatively evaluated.

#### 8.5 Subgroup Analysis

The primary efficacy endpoint may be explored within age strata (< 45, >= 45 years).

©2023 Vielight Inc.
All rights reserved

BRANY
IRB APPROVED
08/24/2023

##### 8.6 Interim Analysis

No interim analysis of the data is planned.

#### 9 CRO and Data Collection Methods

This study is conducted remotely, via an EDC with oversight from a Contract Research Organization (CRO). The PI, sub-investigators and study staff will conduct study procedures and responsibilities remotely. All study procedures will be completed by participants online.

##### 9.1 Study Duration

Recruitment rate of subjects is to be determined based on availability. The duration of the study is approximately 3 months (84 Days) for each subject, from first use of the device. Duration of trial from first subject enrolled to completion of final participant is expected to be approximately 12 months.

#### 10 Ethical Considerations

##### 10.1 Consent Process

Consent shall be obtained in accordance with World Medical Association Declaration of Helsinki – Ethical Principles for Medical Research Involving Human Participants, and 21 CFR Part 50 Protection of Human Subjects. Potential subjects must be informed as to the purpose of the study and the potential risks and benefits known or that can be reasonably predicted or expected as described in the written consent form. The subject shall have sufficient opportunity to consider participation in the study; consent forms shall be written in English. A subject cannot be led to believe that they are waiving their rights as a subject or the liability of the sponsor or investigator. Subjects are then invited to sign and date the consent form, indicating their consent for enrollment. (Investigators may not date the consent form on the subject's behalf.)

##### 10.2 Institutional Review Board (IRB) and Research Ethics Board (REB) Approvals

The Sponsor will apply for an approval by an IRB.

##### 10.3 Data Protection

A Subject Identification Log shall link the data collected during the study to source documents such as medical records. The Subject ID Log shall be kept in a secure and confidential manner with access restricted to study staff.

All information collected during this study will be kept confidential, except the Sponsor's representatives and regulatory authorities will have access to this information.

The Sponsor's representative and/or regulatory authorities will have access to relevant medical information (i.e., COVID-19 positive test report) for purposes for source data verification, including medical information for monitoring and reporting of safety events.

No information which could identify a subject will be used in reports or publications. Subjects will remain de-identified for data analysis.

©2023 Vielight Inc.
All rights reserved

BRANY
IRB APPROVED
08/24/2023

The source documents will be made available for future review with authorization from the Sponsor, subject to privacy laws.

The EDC is privacy compliant in the U.S. (HIPAA).

##### 10.4 Study Suspension or Early Termination

The study can be discontinued at the discretion of the study Sponsor for reasons including, but not limited to, the following:

- Obtaining new scientific knowledge that shows that the study is no longer valid or necessary.
- Insufficient recruitment of subjects
- Unanticipated adverse device effect (UADE) presenting an unreasonable risk to subjects.
- Persistent non-compliance with the protocol
- Persistent non-compliance with regulatory requirements
- Consumption of alcohol within 8 hours of cognitive assessments.

As defined in 21 CFR 813.3, an unanticipated adverse device effect means any serious adverse effect on health or safety or any life-threatening problem or death caused by, or associated with, a device, if that effect, problem, or death was not previously identified in nature, severity, or degree of incidence in the investigational plan or application (including a supplementary plan or application), or any other unanticipated serious problem associated with a device that relates to the rights, safety, or welfare of subjects.

As per 21 CFR 812.150, if Sponsor determines that an unanticipated adverse device effect presents an unreasonable risk to subjects, all investigations or parts of investigations presenting that risk shall be terminated, as soon as possible. Termination shall occur not later than 5 working days after Sponsor makes this determination and not later than 15 working days after the sponsor first received notice of the effect. The study shall not be resumed without FDA and IRB approval.

If the study is discontinued or suspended, the Sponsor shall promptly inform the PI, CRO and participants of the termination or suspension and the reason(s) for this. The IRB will also be informed promptly and provided with the reason(s) for the termination or suspension by the Sponsor or by the clinical investigator/investigation center(s). Regulatory authorities and the personal physicians of the subjects may also need to be informed if deemed necessary.

#### 11 Investigational Devices

Device Inventory Log records, the serial number, date received, date used, and subject ID, will be maintained by the CRO. Devices will be shipped to the subjects through courier services. Once the treatment phase is over (i.e. right after Day 56), the subjects will be asked to return the device using a pre-print shipping label.

A. Labeling: All products will include labeling on them as well as shelf carton. All products will include Instructions for Use included in the shelf carton.

B. Storage: All products at the CRO will be stored at room temperature in a secure location.

©2023 Vielight Inc.
All rights reserved

BRANY
IRB APPROVED
08/24/2023

C. Return: If the Device is associated with a device related adverse event, malfunction or failure, the Device should be returned to Vielight Inc. for appropriate action or destroyed as instructed by the Sponsor.

### 12 Safety and Adverse Events

#### 12.1 Definition of Adverse Events

An adverse event is any unfavourable and unintended sign, including an abnormal laboratory finding, symptom, or disease temporarily associated with the use of a medical treatment that may or may not be considered related to the medical treatment or procedure.

Adverse events (AEs) may occur after enrolment but prior to the start of treatment or during the treatment period. Adverse events occurring prior to the start of treatment, while not directly associated with the use of the Vielight Neuro RX Gamma, will be documented in the subject's medical record but will not count as a study device-related event. Participants will be asked to report any adverse events on a daily basis through their daily diary. If the subjects are experiencing a device related adverse event, they will be instructed to contact research staff. Participants will be instructed to follow their local public health guidance on seeking medical attention. It is the participants' responsibility to monitor their own health and to seek medical attention per their local public health guidance and instructions from their local health care provider.

Investigators will record characteristics of each adverse event on an Adverse Event CRF. Each adverse event will be judged by the Investigator as to its relationship and level of relatedness to the investigational device. Relatedness will be scored consistent with CTCAE v4.03 guidelines. CTCAE v4.03 is the National Cancer Institute (USA) Common Terminology Criteria for Adverse Effects version 4.03 (published June 14, 2010) is a descriptive terminology which can be used for Adverse Event (AE) reporting.

- Unrelated – the AE is clearly not related to the investigational agent(s),
- Unlikely – the AE is doubtfully related to the investigational agent(s),
- Possible – the AE may be related to the investigational agent(s),
- Probable – the AE is likely related to the investigational agent(s),
- Definite – the AE is clearly related to the investigational agent(s).

In addition, the Investigator will identify the date of onset, severity, and duration. Severity is determined from the grades presented in CTCAE v4.03. The CTCAE v4.03 displays Grades 1 through 5 with unique clinical descriptions of severity for each AE based on this general guideline:

- Grade 1 – Mild AE
- Grade 2 – Moderate AE
- Grade 3 – Severe AE.
- Grade 4 – Life-threatening or disabling AE.
- Grade 5 – Death related to AE.

©2023 Vielight Inc.
All rights reserved

BRANY
IRB APPROVED
08/24/2023

An AE is defined as being serious (SAE) if it results in death or meets the definition of serious injury, as defined in Title 21 of the Code of Federal Regulations, section 803.3 (21 CFR 803.3), which means an injury or illness that:

i. Is life-threatening,
ii. Results in permanent impairment of a body function or permanent damage to a body structure; or
iii. Necessitates medical or surgical intervention to preclude permanent impairment of a body function or permanent damage to a body structure. Permanent means irreversible impairment or damage to a body structure or function, excluding trivial impairment or damage.

As defined in 21 CFR 803.3, caused, or contributed means that the adverse device effect was or may have been attributed to the study device, or may have been a factor in in the event, including events occurring because of:

- Failure,
- Malfunction,
- Improper or inadequate design,
- Manufacture,
- Labeling, or
- User error.

All adverse events will be monitored until they are adequately resolved or explained.

##### 12.2 Definition of Device Incident

An incident according to Good Clinical Practice

(http://www.fda.gov/RegulatoryInformation/Guidances/ucm122046.htm) is defined as an event that:

(a) is related to a failure of the device or a deterioration in its effectiveness, or any inadequacy in its labeling or in its directions for use; and

(b) has led to the death or a serious deterioration in the state of health of a subject, user, or other person, or could do so were it to recur.

##### 12.3 Reporting Adverse Events and Device Incidents

All Serious Adverse Events (SAEs, see Definitions) must be reported to the study Sponsor immediately, not to exceed 24 hours after the investigator first learns of the event. The completed adverse event investigation form shall be faxed or emailed to the Sponsor within 24 hours together with a cover letter describing the event and detailing the medical history, concomitant medication, and an assessment of compliance with therapy.

All adverse events, serious or non-serious adverse events will be reported in the daily diary.

All SAEs need to be followed until the event is resolved (with or without sequelae). The PI will decide if more follow up information is needed in case the event is not resolved at study completion. In case of death, all possible information that is available, including the possible relationship to the device, should be provided.

The investigator must submit to the Sponsor any unanticipated adverse device incident (see Definitions) within 24 hours after the investigator first learns of the effect. The investigator must

©2023 Vielight Inc.
All rights reserved

BRANY
IRB APPROVED
08/24/2023

also report the unanticipated adverse device effect to the IRB within its pre-specified timeline. As per 21 CFR 812.150, this incident reporting shall not be later than 10 working days after the investigator first learns of the effect. The Investigator will report all the above to the reviewing IRB (as applicable) according to their reporting requirements. Reports must identify subjects using the study's unique identifier to protect subject's confidentiality.

The Sponsor will be responsible for submitting incident information to FDA within the specified timeframe. As per 21 CFR 812.46, Sponsor shall immediately conduct an evaluation of any unanticipated adverse device effect. Sponsor shall report the results of such evaluation to FDA and to the reviewing IRB and participating investigators within 10 working days after receiving notice of the effect. Thereafter the sponsor shall submit such additional reports concerning the effect as FDA requests.

##### 12.4 Adverse Event Follow-up

The investigator shall continue to monitor any unanticipated, device related adverse event until it is resolved or up to one month following the end of the study.

##### 12.5 Anticipated Adverse Events

There are no adverse events anticipated to be device attributable with use of the Vielight Neuro RX Gamma. The device predecessor (Vielight Neuro Gamma) has been currently sold as a general wellness device worldwide with no major adverse events having been reported to date. It satisfies the criteria of an unregulated low risk device defined by the FDA policy, "General Wellness: Policy for Low Risk Devices".

##### 12.6 Protocol Deviations

A protocol deviation is defined as any study action taken by the PI to conflict with the Study Protocol. Investigators must make every effort to follow the protocol except where necessary to protect the life or physical well-being of a subject in an emergency. All deviations from the protocol will be reported on the appropriate Deviations Log.

Deviations must be reported to the Sponsor within a reasonable time frame. Subject specific deviations will be reported on the Deviations Log. Non-subject specific deviations, (e.g., unauthorized use of an investigational device outside the study, etc.), will be reported to the PI. Investigators will also adhere to procedures for reporting study deviations to the IRB if applicable.

Regulations require that Investigators maintain accurate, complete, and current records, including documents showing the dates of and reasons for each deviation from the protocol.

For participants who are noncompliant with follow up visits, every effort will be made to collect the data requested by this protocol through the follow up period. All data collected up until the period the subject has withdrawn from the study will be used for this study. Participants will be defined as non-compliant if they miss 4 consecutive days of treatment.

#### 13 Study Management

##### 13.1 Sponsor Overall Responsibility

As the study sponsor, Vielight Inc. has the overall responsibility for the conduct of the study according to Good Clinical Practice Consolidated Guidance, ICH, 1997), ISO 14155: Part 1 and

©2023 Vielight Inc.
All rights reserved
BRANY
IRB APPROVED
08/24/2023

2, the Declaration of Helsinki, Medical Device Directive, Annex X, conditions imposed by the reviewing IRB/REB, FDA 21 CFR Part 812, 21 CFR Part 50, 21 CFR Part 56 and all applicable local regulatory requirements. For this study, Vielight Inc. will have certain direct responsibilities and will delegate other responsibilities to appropriate consultants and contract research organizations (CROs). Together, Vielight Inc., consultants and CROs will ensure that the study is conducted according to all applicable regulations. All personnel to participate in the conduct of this clinical trial will be qualified by training, education and/or experience to perform his or her respective tasks.

NOTE: A complete list of participating investigators will be maintained and will be available upon request.

##### 13.2 Investigator Responsibilities

The Investigator shall be responsible for the day-to-day conduct of the investigation as well as for ensuring that the investigation is conducted according to all signed agreements, applicable elements of ISO 14155, the Clinical Investigational Plan, applicable FDA regulations, the principles that have their origin in the Declaration of Helsinki. The investigator is also responsible for protecting the rights, safety, and welfare of subjects under the investigator's care and for obtaining informed consent in accordance with FDA's regulations 21 CFR Part 50 Protection of Human Subjects and 21 CFR Part 812 Investigational Device Exemptions. Each Investigator must sign the Investigator Agreement and Financial Disclosure prior to subject enrolment. No investigator will be added to the investigation until a signed Investigator Agreement is provided.

Responsibilities of the Investigator include, but are not limited to:

1. Submit proposed amendments to the protocol and informed consent to the IRB and await approval unless the change reduces the risk to subjects.
2. Ensuring that all personnel assisting with the clinical trial are adequately informed and understand their trial-related duties and functions.
3. Obtain informed consents of subjects.
4. Permit monitor to inspect the records.
5. Maintain medical histories of subjects.
6. Enroll subjects, execute the study, transcribe data from source documents to case report forms, and conduct study in accordance with protocol.
7. Submit annual progress reports, final reports, and Adverse Event reports to the IRB and to Sponsor.
8. Notify the reviewing IRB and the manufacturer (Vielight Inc.) of any unanticipated adverse device effect occurring during an investigation as soon as possible, but in no event later than 10 working days after the investigator first learns of the effect, in accordance with 21CFR812.150.
9. Disclose to Vielight Inc. sufficient accurate financial information to allow them to submit completed and accurate certification and disclosure statements under US 21 CFR54. The investigator shall promptly update this information if any relevant changes occur during the investigation and for 1 year following completion of the study.

©2023 Vielight Inc.
All rights reserved
BRANY
IRB APPROVED
08/24/2023

The Investigator will allow direct access to source data/documents for trial related monitoring, audit, IRB review and regulatory inspection. Also, the investigator will allow auditing of their clinical investigational procedure(s).

##### 13.3 Required Documents from the Investigators

At a minimum, the following documents will be provided by the Investigators to the study sponsor:

- Signed Investigator Agreement
- Current Curriculum Vitae
- Completed and accurate certification and disclosure statements under US 21 CFR54
- Any other relevant documents requested by the study sponsor, IRB, or other regulatory authority.

The study procedures will not be initiated until all the above listed documents have been provided to the study sponsor.

##### 13.4 Investigator Records

The Investigator is responsible for maintaining medical and study records for every subject participating in the clinical study (including information maintained electronically). The Investigator will also maintain original source documents from which study-related data are derived, which include, but are not limited to:

- all correspondence including required reports with another investigator, an IRB, the sponsor, FDA, or any other competent authority.
- records of each subject's case history and exposure to the device which must include,
- signed and dated consent forms
- condition of each subject upon entering the study
- relevant previous medical history
- observations of relevant adverse device effects (anticipated or unanticipated)
- adverse events reporting and follow-up of the adverse events.
- case report forms
- subjects' condition upon completion of or withdrawal from the study
- any other supporting data
- the protocol and documentation (date and reason) for each deviation from the protocol.
- any other records that FDA or other regulatory body requires to be maintained by regulation or by specific requirement for a category of investigation or a particular investigation.
- The Investigator must ensure that all subject records are stored for at least 2 years after the end of the clinical study or the records are no longer required to support a regulatory approval, whichever date is later, per 21 CFR 812.140(d). To avoid error, the CRO should contact Vielight Inc. prior to the destruction of study records to ensure that they no longer need to be retained. In addition, Vielight Inc. should be contacted if the Investigator plans to leave the study so that arrangements can be made for the handling or transfer of study records. Records must be retained in designated administrative files at each Investigational Site and/or retained by the study sponsor.
    - Trial protocol and all amendments

©2023 Vielight Inc.
All rights reserved
BRANY
IRB APPROVED
08/24/2023

- Ethics Committee Approval Letter(s) and approved informed consent(s) (including any revisions)
- Device Instructions for Use
- CVs of all investigators
- Site Subject Log
- Monitoring reports
- Site Authorized Personnel Signature List
- Reports (includes Adverse Event reports and final reports from investigator and sponsor)

##### 13.5 Specific Sponsor Responsibilities

Vielight Inc. is the Sponsor of this study. Vielight Inc.'s responsibilities include but are not limited to:

1. Selecting investigators that are qualified by training and education (qualifications will be documented by curriculum vitae).
2. Providing investigators with the information necessary to conduct the investigation properly.
3. Providing appropriate training to each the CRO and all study personnel (monitors), as necessary.
4. Documenting training where appropriate.
5. Selecting monitors qualified by training and experience to monitor the investigational study in accordance with FDA regulations or train monitors if necessary.
6. Ensuring that the IRB approval is obtained.
7. Ensuring that any reviewing IRB or regulatory authority are informed of significant new information.
8. Providing the devices to Sub-I
9. Report and investigate unanticipated, device related Adverse Events.
10. Obtaining signed Investigator Agreement for each investigator prior to their participation in the study.
11. Obtaining sufficient and accurate financial disclosure information
12. Reporting per local regulations.
13. Retain records for at least 2 years following completion of this study.

##### 13.6 Sponsor Records

The Sponsor shall maintain the following accurate, complete, and current records relating to the investigation:

- All correspondence including required reports.
- Signed investigator’s agreements including financial disclosure information.
- Records concerning adverse device effects and complaints.

##### 13.7 Study Monitoring Plan

The study will be monitored to ensure that applicable regulations are followed. Written procedures will be established in a monitor plan to ensure the quality of the study and to ensure that each person involved in the monitoring process carries out the required duties. The sponsors shall designate or assign monitors to this clinical study.

Source-documents will be reviewed to verify CRFs and database information for completion and accuracy. CRF findings of non-compliance or required modifications shall be reviewed with the

Investigator(s). The monitor will report to the sponsor any non-compliance with the signed Investigator's Agreement, conditions imposed by the IRB or regulatory authorities. The sponsor shall then either secure compliance or terminate the investigator's participation in the investigation.

An initial review of study progress and compliance will be conducted after the first 5 participants have completed baseline procedures. Next visit will take place after 25 participants. At this time the monitor will review study progress and compliance with the protocol, including but not limited to, review of rate of recruitment, number of screen failures, query resolution, and review of ISF/TMF documents.

A final monitoring assessment will be conducted once all participants (i.e., 36) have completed the study procedures for study-close out. Monitoring visits may occur at other timepoints at the Sponsor's discretion. At each monitoring visit the CRO will be available during monitor's visit for query resolution and any other requested study material for review.

##### 13.8 Medical Monitoring

An independent Medical Monitor will review safety reports biweekly. The reports will contain the number and type of adverse events, and all the serious adverse events classified as probably or definitely related to enrolment in the trial. The Medical monitor will review all SAEs within 48 hours of their report. The Medical Monitor will have the ability to request additional safety analyses and make recommendations about the safe conduct of the trial. Within 7 business days of the review date, the medical monitor will communicate any findings or recommendations directly with Principal Investigator**.** The Medical Monitor can recommend stopping the investigation due to safety reasons.

Participants will be instructed to seek medical attention per local public health guidance and according to their healthcare practitioner.

##### 13.9 Ethical Considerations

The rights, safety and well-being of clinical investigation subjects shall be protected consistent with the ethical principles laid down in the Declaration of Helsinki. This shall be understood, observed, and applied at every step in this clinical investigation.

It is expected that all parties will share in the responsibility for ethical conduct in accordance with their respective roles in the investigation. The Sponsor and the Investigator(s) shall avoid improper influence or inducement of the subject, monitor, the clinical investigator(s), or other parties participating in or contributing to the clinical investigation.

##### 13.10 Protection of Subject Confidentiality

At all times throughout the clinical investigation, confidentiality will be observed by all parties involved. All data shall be secured against unauthorized access. Privacy and confidentiality of information about each subject shall be preserved in study reports and in any publication. Each subject participating in this study will be assigned a unique identifier. All CRFs will be tracked, evaluated, and stored using only this unique identifier.

The EDC will be overseen by the CRO and will maintain a confidential subject list identifying all enrolled subjects (Enrollment Log). This list will contain the assigned subject's unique identifier and name. The EDC is Health Insurance Portability and Accountability Act (HIPAA) compliant.

©2023 Vielight Inc.
All rights reserved
BRANY
IRB APPROVED
08/24/2023

Monitors and auditors will have access to the subject list and other personally identifying information of subjects to ensure that data reported in the CRF corresponds to the person who signed the ICF, and the information contained in the original source documents. Such personal identifying information may include, but is not limited to date of birth, sex, race, and COVID-19 test report.

Any source documents copied for monitoring purposes by the Sponsor will be identified by using the assigned subject's unique identifier to protect subject confidentiality.

##### 13.11 Quality Assurance and Supervision by Authorities and Privacy

All documents and data shall be produced and maintained in such a way to assure control of documents and data to protect the subject's privacy as far as reasonably practicable. The EDC technology platform is privacy compliant with the U.S. (HIPAA) and will undergo regular monitoring as detailed in 13.7. The Sponsor and representatives of the regulatory authorities are permitted to inspect the study documents (e.g., study protocol, CRFs, and original study-relevant medical records/files) as needed. All attempts will be made to preserve subject confidentiality.

##### 13.12 Final Report

A final report will be completed, even if the study is prematurely terminated. At the conclusion of the trial, an abstract reporting the results will be prepared and will be presented at a major meeting(s). A manuscript could also be prepared for publication in a reputable scientific journal as optional.

##### 13.13 Information Confidentiality

All information not previously published concerning the test device and research, including patent applications, manufacturing processes, basic scientific data, etc., is considered confidential and should remain the sole property of Vielight Inc. All information and data generated in association with this study will be held in strict confidence and remain the sole property of Vielight Inc. The Investigator agrees to use this information for the sole purpose of completing this study and for no other purpose without written consent from Vielight Inc.

##### 13.14 Trial Registration

The study will be registered in a publicly accessible trial database (clinicaltrials.gov) prior to study initiation.

#### 14 Study Close-out

##### 14.1 Timeline of Close-out

Study close-out will be conducted by the Sponsor's monitor within 45 days of receipt of the last case report form.

##### 14.2 Record Storage and Retention

The Sponsor shall retain copies of all study data and documentation for a minimum of 2 years after the last approval of a marketing application in an ICH region and until there are no pending

©2023 Vielight Inc.
All rights reserved

BRANY
IRB APPROVED
08/24/2023

or contemplated marketing applications in an ICH region or at least 2 years have elapsed since the formal discontinuation of clinical development.

##### 14.3 Discontinuation of Study

Vielight Inc. reserves the right to discontinue any study for business or ethical reasons at any time, such as but not limited to, a decision to discontinue further clinical investigations with the test article, improper conduct of the study by the investigator, inability to obtain the number of subjects required by the protocol, etc. Reimbursement for reasonable expenses will be made if such action is necessary.

#### 15 Definitions and Acronyms

**Adverse Event (AE)** - any untoward medical occurrence in a subject (ISO 14155). Note: This definition does not imply that there is a relationship between the adverse event and the device under investigation

An adverse event is any unfavourable and unintended sign (including an abnormal laboratory finding, symptom, or disease temporarily associated with the use of a medical treatment or procedure that may or may not be considered related to the medical treatment or procedure (CTCAE v 4.03).

**Serious Adverse Event (SAE)** - an adverse event that (ISO 14155):

- led to a death,
- led to a serious deterioration in the health of the subject,
- resulted in a life-threatening illness or injury,
- resulted in a permanent impairment of a body structure or a body function,
- required hospitalization or prolongation of existing hospitalization,
- resulted in medical or surgical intervention to prevent permanent impairment to body structure or function,
- led to fetal distress, fetal death, a congenital abnormality, or birth defect.

**Adverse Device Effect (ADE)** - any untoward and unintended response to a medical device, including that resulting from user error (ISO 14155)

**Serious Adverse Device Effect (SADE)** - an adverse device effect that has resulted in any of the consequences characteristic of a serious adverse event or that might have led to any of these consequences if suitable action had not been taken or intervention had not been made or if circumstances had been less opportune (ISO 14155).

**Unanticipated Adverse Device Effect (UADE)** - any serious adverse effect on health or safety or any life-threatening problem or death caused by, or associated with, a device, if that effect, problem, or death was not previously identified in nature, severity, or degree of incidence in the investigational plan or application (including a supplementary plan or application), or any other unanticipated serious problem associated with a device that relates to the rights, safety, or welfare of subjects (ISO 14155).

**Applicable Regulatory Requirement(s)-** Any law(s) and regulation(s) addressing the conduct of clinical trials of investigational products of the jurisdiction where trial is conducted (E6, GCP Guidance)

**Case Report Form (CRF)** - A printed document designed to record all protocol-required information to be reported to the sponsor on each subject (E6, GCP Guidance)

©2023 Vielight Inc.
All rights reserved
BRANY
IRB APPROVED
08/24/2023

**Contract Research Organization (CRO)** - A person or an organization (commercial, academic, or other) contracted by the sponsor to perform one or more of a sponsor's trial-related duties and functions (E6, GCP Guidance).

**Creyos** – is a cognitive assessment tool comprise of 12 tests.

**COVID-19** - Coronavirus Disease 2019

**Documentation** - All records, in any form (including, but not limited to, written electronic, magnetic, and optical records, and scans. X-rays, and electrocardiograms) that describe or record the methods, conduct, and/or results of a trial, the factors affecting a trial and the actions taken (E6, GCP Guidance).

**Electronic Data Capture (EDC)** – is a computerized system designed for the collection of clinical data in electronic format for use mainly in human clinical trials.

**EQ-5D-5L (Health Questionnaire)** – The descriptive system comprises five dimensions: mobility, self-care, usual activities, pain/discomfort, and anxiety/depression.

**Fatigue Assessment Scale (FAS)** - is a 10-item scale evaluating symptoms of chronic fatigue.

**Fitzpatrick scale** - is a numerical classification schema for human skin color.

**Good Clinical Practice (GCP)**- A standard for the design, conduct, performance, monitoring, auditing, recording, analyses, and reporting of clinical trials that provides assurance that the data and reported results are credible and accurate, and that the rights, integrity, and confidentiality of trial subjects are protected (E6, GCP Guidance)

**Informed Consent** - The process by which the subject voluntarily confirms his or her willingness to participate in a particular trial, after having been informed of all aspects of the trial that are relevant to the subject's decision to participate. Informed consent is documented by means of a written, signed, and dated informed consent form.

**Informed Consent Form (ICF)** - a document that describes:

a. the risks and anticipated benefits to his or her health arising from participation in the clinical trial; and
b. All other aspects of the clinical trial that are necessary for that person to make the decision to participate in the clinical trial.

**Institutional Review Board (IRB)**
IRB) - An independent body (a review board or a committee, institutional, regional, national or supranational), constituted of medical/scientific professionals and nonmedical/non-scientific members, whose responsibility is to ensure the protection of the rights, safety, and well-being of human subjects involved in a trial and to provide public assurance of that protection, by, among other things, reviewing and approving/providing favourable opinion on the trial protocol, the suitability of the investigator(s), facilities, and the methods and materials to be used in obtaining and documenting informed consent of the trial subjects (E6, GCP Guidance).

**Instructions for Use (IFU)** - Step by step directions for using the product.

**Investigator /Principal Investigator (PI)** - A person responsible for the conduct of the clinical trial. If a trial is conducted by a team of individuals, the investigator is the responsible leader of the team and may be called the principal investigator (E6, GCP Guidance).

**Investigator Site File (ISF)** - contains essential documents which shows that the clinical trial site and Investigator are following the regulatory requirements, from initiation to closeout.

©2023 Vielight Inc.
All rights reserved

BRANY
IRB APPROVED
08/24/2023

**Monitoring** - Monitor when used as a verb means to oversee an investigation (ISO 14155). Monitoring is the act of overseeing the progress of a clinical trial, and of ensuring that it is conducted, recorded, and reported in accordance with the protocol, standard operating procedures, GCP and the applicable regulatory requirements (E6, GCP Guidance).

**Neuropsychological Questionnaire** – is to provide a brief assessment of neuropsychological symptomatology.

**NIR** - Near Infrared

**PDQ-20 (Perceived Deficits Questionnaire)** - consists of 20 items and instructs participants to assess their cognitive functioning.

**PBM** – Photobiomodulation

**PHQ-9 (Patient Health Questionnaire - 9) -** objectifies and assesses degree of depression severity via questionnaire.

**Post COVID-19** -also known as Long COVID, long-haul COVID, post-acute COVID-19, post-acute sequelae of SARS CoV-2 infection (PASC), long-term effects of COVID, and chronic COVID. According to CDC, Long COVID is broadly defined as signs, symptoms, and conditions that continue or develop after initial COVID-19 or SARS-CoV-2 infection. The signs, symptoms, and conditions are present four weeks or more after the initial phase of infection; may be multisystemic; and may present with a relapsing– remitting pattern and progression or worsening over time, with the possibility of severe and life-threatening events even months or years after infection. Long COVID is not one condition. It represents many potentially overlapping entities, likely with different biological causes and different sets of risk factors and outcomes.

**SARS-CoV-2** - Severe Acute Respiratory Symptom Coronavirus 2.

**Source Data** - All information in original and identified records and certified copies of original records of clinical findings, observations, or other activities in a clinical investigation, necessary for the reconstruction and evaluation of the clinical investigation. Source data are contained in source documents (E6 GCP Guidance, ISO 14155).

**Source Documents** - Original documents, data, and records (ISO 14155).

*Note: This may be, for example, hospital records, laboratory notes, pharmacy dispensing records, copies or transcriptions certified after verification as being accurate copies, photographic negatives, radiographs, and records kept at the pharmacy, at the laboratories, and at medico-technical departments involved in the clinical investigation.*

**Standard Operating Procedure (SOP) -** are uniformly written procedures, with detailed instructions to record routine operations, processes and practices followed within a business organization.

**Symptom Burden Questionnaire (SBQ™-LC) -** for Long COVID system (SBQ™-LC) is a patient-reported outcome (PRO) measure and multi-domain item bank that has been developed according to international best-practice and regulatory guidance. A modified version of SBQ was utilized in this protocol.

**Termination** -Termination means a discontinuation, by sponsor or by withdrawal of IRB of an investigation before completion.

**Treatment** - Vielight Neuro RX Gamma advice and sham.

**Trial Master File (TMF)** – is a compilation of documents that prove that the clinical trial has been conducted following regulatory requirements (including Good Clinical Practice).

©2023 Vielight Inc.
All rights reserved

BRANY
IRB APPROVED
08/24/2023

©2023 Vielight Inc.
All rights reserved

BRANY
IRB APPROVED
08/24/2023

16 Park IS, Lee NJ, Rhyu IJ. Roles of the Declive, Folium, and Tuber Cerebellar Vermian Lobules in Sportspeople. Journal of clinical neurology (Seoul, Korea). 2018; 14(1):540 1-7. doi:10.3988/jcn.2018.14.1.1.

17 Fernández-Castañeda A, Lu P, Geraghty AC, et al. Mild respiratory COVID can cause multi-lineage neural cell and myelin dysregulation. Cell. June 12, 2022. doi:10.1016/j.cell.2022.06.008

18 de Matos BTL, Buchaim DV, Pomini KT, et al. Photobiomodulation Therapy as a Possible New Approach in COVID-19: A Systematic Review. Life (Basel). 2021;11(6):580. Published 2021 Jun 18. doi:10.3390/life11060580.

19 Macedo DB, Tim CR, Kido HW, et al. Influence of photobiomodulation therapy on the treatment of pulmonary inflammatory conditions and its impact on COVID-19. Lasers Med Sci. 2022;37(3):1921-1929. doi:10.1007/s10103-021-03452-5.

20 Iaccarino H, Singer A, Martorell A, et al. Gamma frequency entrainment attenuates amyloid load and modifies microglia. Nature. 2016;540:230–235. doi:10.1038/nature20587.

21 Ismail R, Hansen AK, Parbo P, et al. The Effect of 40-Hz Light Therapy on Amyloid Load in Patients with Prodromal and Clinical Alzheimer's Disease. Int J Alzheimers Dis. 2018; 6852303. doi:10.1155/2018/6852303.

22 de Freitas L. F and M. R. Hamblin (2016). "Proposed Mechanisms of Photobiomodulation or Low-Level Light Therapy." IEEE J Sel Top Quantum Electron 22(3):7000417. doi:10.1109/JSTQE.2016.2561201.

23 Fekrazad R, Fekrazad S. The Potential Role of Photobiomodulation in Long COVID-19 Patients Rehabilitation. Photobiomodul Photomed Laser Surg. 2021 Apr;39(4):229-231. doi: 10.1089/photob.2020.4984. Epub 2021 Jan 27. PMID: 33497594.

24 Dias, L. D., Blanco, K. C., de Faria, C. M., Dozza, C., Zanchin, E. M., Paolillo, F. R., ... & Bagnato, V. S. (2022). Perspectives on photobiomodulation and combined light-based therapies for rehabilitation of patients after COVID-19 recovery. Laser Physics Letters, 19(4), 045604.

25 de Matos BTL, Buchaim DV, Pomini KT, et al. Photobiomodulation Therapy as a Possible New Approach in COVID-19: A Systematic Review. Life (Basel). 2021;11(6):580. Published 2021 Jun 18. doi:10.3390/life11060580.

26 Macedo DB, Tim CR, Kido HW, et al. Influence of photobiomodulation therapy on the treatment of pulmonary inflammatory conditions and its impact on COVID-19. Lasers Med Sci. 2022;37(3):1921-1929. doi:10.1007/s10103-021-03452-5.

27 Iaccarino H, Singer A, Martorell A, et al. Gamma frequency entrainment attenuates amyloid load and modifies microglia. Nature. 2016;540:230–235. doi:10.1038/nature20587.

28 Ismail R, Hansen AK, Parbo P, et al. The Effect of 40-Hz Light Therapy on Amyloid Load in Patients with Prodromal and Clinical Alzheimer's Disease. Int J Alzheimers Dis. 2018; 6852303. doi:10.1155/2018/6852303.

29 Al Amir Dache Z., A. Otandault, R. Tanos, B. Pastor, R. Meddeb, C. Sanchez, G. Arena, L. Lasorsa, A. Bennett, T. Grange, S. El Messaoudi, T. Mazard, C. Prevostel and A. R. Thierry (2020). "Blood contains circulating cell-free respiratory competent mitochondria." FASEB J 34(3):3616-3630.

30 MgGuff, P. E., Deterling Jr, R. A., & Gottlieb, L. S. (1965). Tumoricidal effect of laser energy on experimental and human malignant tumors. New England Journal of Medicine, 273(9), 490-492.

©2023 Vielight Inc.
All rights reserved

BRANY
IRB APPROVED
08/24/2023

31 Mester, E., Spiry, T., & Szende, B. (1973). Effect of laser rays on wound healing. Bulletin de la Societe internationale de chirurgie, 32(2), 169-173.

32 Lane, N. (2006). Power games. Nature, 443, 901-903.

33 Mittal, M., Siddiqui, M. R., Tran, K., Reddy, S. P., & Malik, A. B. (2014). Reactive oxygen species in inflammation and tissue injury. Antioxidants & redox signaling, 20(7), 1126-1167.

34 Chen, A. C.-H., Huang, Y.-Y., Arany, P. R., & Hamblin, M. R. (2009). Role of reactive oxygen species in low level light therapy.

35 Bjordal, J. M., Couppé, C., Chow, R. T., Tunér, J., & Ljunggren, E. A. (2003). A systematic review of low level laser therapy with location-specific doses for pain from chronic joint disorders. Australian journal of physiotherapy, 49(2), 107-116.

36 Hamblin, M. R. (2017). Mechanisms and applications of the anti-inflammatory effects of photobiomodulation. AIMS biophysics, 4(3), 337.

37 Zomorrodi R., G. Loheswaran, A. Pushparaj and L. Lim (2019). “Pulsed Near Infrared Transcranial and Intranasal Photobiomodulation Significantly Modulates Neural Oscillations: a pilot exploratory study.” Sci Rep 9(1):6309.

38 Chao L.L. (2019). “Effects of Home Photobiomodulation Treatments on Cognitive and Behavioral Function, Cerebral Perfusion, and Resting-State Functional Connectivity in Patients with Dementia: A Pilot Trial”. Photobiomodul Photomed Laser Surg 37(3):133-141.

39 Saltmarche, A., M. Naeser, K. F. Ho, M. R. Hamblin, and L. Lim (2017). "Significant Improvement in Cognition in Mild to Moderately-Severe Dementia Cases Treated with Transcranial plus Intranasal Photobiomodulation: Pilot Study." Photomed Laser Surg 35(8):432-441.

40 Naeser M. A., P. I. Martin, M. D. Ho, M. H. Krengel, Y. Bogdanova, J. A. Knight, M. K. Yee, R. Zafonte, J. Frazier, M. R. Hamblin, B. B. Koo (2016). “Transcranial, Red/Near-Infrared Light-Emitting Diode Therapy to Improve Cognition in Chronic Traumatic Brain Injury.” Photomed Laser Surg 34(12):610-626.

41 EMEA European Medicines agency. ICH Topic E9, Statistical Principles for Clinical Trials, September 1998.

©2023 Vielight Inc.
All rights reserved

BRANY
IRB APPROVED
08/24/2023

**Appendix 1**

Neuropsychological Questionnaire

**Delusions**:

Do you have beliefs that you know are not true (for example, insisting that people are trying to harm you or steal from you)? Have you ever said that family members are not who they say they are or that your house is not your home? We are not asking about suspiciousness; we are interested if you are convinced that these things are happening to you.

Yes/No

**Hallucinations**:

Have you ever had hallucinations such as false visions or voices? Does he or she seem to hear or see things that are not present? By this question we do not mean just mistaken beliefs such as stating that someone who has died is still alive; rather we are asking if the patient has abnormal experiences of sounds or visions.

Yes/No

**Depression/Dysphoria**:

Over the past two weeks, have you been bothered by:

-Little interest or pleasure in doing things?

-Feeling down, depressed, or hopeless?

Yes/No

If yes, complete PHQ-9

**Bipolar disorder**:

Have you ever had a period of at least 1 week during which you felt any of the following: unusually happy; unusually outgoing; unusually energetic, more talkative than normal with thoughts racing in your head, or needed much less sleep than usual?

Yes/No

©2023 Vielight Inc.
All rights reserved

BRANY
IRB APPROVED
08/24/2023

**Neurological disorder screening:**

**Motor Disturbance**: Are you engaging in repetitive activities such as pacing around the house, handling buttons, wrapping string, or doing other things repeatedly?

**Do you have a history of Stroke?** Yes/No

**Do you have a history of Seizure?** Yes/No

**Do you have a diagnosis of chronic fatigue syndrome [CFS]?**Yes/No

**Suicidal thought questions:**

In the past week, have you been having thoughts about killing yourself or any other person?

Yes/No

#### Appendix 2

PHQ-9

©2023 Vielight Inc.
All rights reserved

BRANY
IRB APPROVED
08/24/2023

#### Patient Health Questionnaire (PHQ-9)

Name: ____________________________________________ Date: ________________________

| Over the last 2 weeks, how often have you been bothered by any of the following problems? | Not at all | Several days | More than half the days | Nearly every day |
|---|---|---|---|---|
| 1. Little interest or pleasure in doing things | 0 | 1 | 2 | 3 |
| 2. Feeling down, depressed, or hopeless | 0 | 1 | 2 | 3 |
| 3. Trouble falling or staying asleep, or sleeping too much | 0 | 1 | 2 | 3 |
| 4. Feeling tired or having little energy | 0 | 1 | 2 | 3 |
| 5. Poor appetite or overeating | 0 | 1 | 2 | 3 |
| 6. Feeling bad about yourself – or that you are a failure or have let yourself or your family down | 0 | 1 | 2 | 3 |
| 7. Trouble concentrating on things, such as reading the newspaper or watching television | 0 | 1 | 2 | 3 |
| 8. Moving or speaking so slowly that other people could have noticed? Or the opposite – being so fidgety or restless that you have been moving around a lot more than usual | 0 | 1 | 2 | 3 |
| 9. Thoughts that you would be better off dead or of hurting yourself in some way | 0 | 1 | 2 | 3 |

For office coding: Total Score _______ = ______ + ______ + ______

Total Score ______

If you checked off any problems, how difficult have these problems made it for you to do your work, take care of things at home, or get along with other people?

☐ Not difficult at all ☐ Somewhat difficult ☐ Very difficult ☐ Extremely difficult

##### Appendix 3

EQ-5D-5L

©2023 Vielight Inc.
All rights reserved

BRANY
IRB APPROVED
08/24/2023

Under each heading, please tick the ONE box that best describes your health TODAY.

**MOBILITY**

I have no problems in walking about ❑
I have slight problems in walking about ❑
I have moderate problems in walking about ❑
I have severe problems in walking about ❑
I am unable to walk about ❑

**SELF-CARE**

I have no problems washing or dressing myself ❑
I have slight problems washing or dressing myself ❑
I have moderate problems washing or dressing myself ❑
I have severe problems washing or dressing myself ❑
I am unable to wash or dress myself ❑

**USUAL ACTIVITIES** *(e.g. work, study, housework, family or leisure activities)*

I have no problems doing my usual activities ❑
I have slight problems doing my usual activities ❑
I have moderate problems doing my usual activities ❑
I have severe problems doing my usual activities ❑
I am unable to do my usual activities ❑

**PAIN / DISCOMFORT**

I have no pain or discomfort ❑
I have slight pain or discomfort ❑
I have moderate pain or discomfort ❑
I have severe pain or discomfort ❑
I have extreme pain or discomfort ❑

**ANXIETY / DEPRESSION**

I am not anxious or depressed ❑
I am slightly anxious or depressed ❑
I am moderately anxious or depressed ❑
I am severely anxious or depressed ❑
I am extremely anxious or depressed ❑

©2023 Vielight Inc.
All rights reserved

BRANY
IRB APPROVED
08/24/2023

- We would like to know how good or bad your health is TODAY.
- This scale is numbered from 0 to 100.
- 100 means the best health you can imagine.
  0 means the worst health you can imagine.
- Mark an X on the scale to indicate how your health is TODAY.
- Now, please write the number you marked on the scale in the box below.

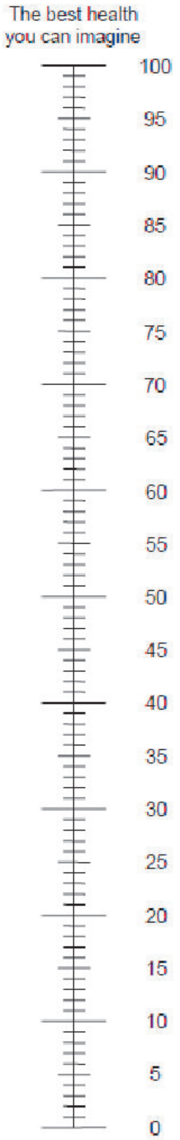

YOUR HEALTH TODAY = [ ]

**Appendix 4**

Fatigue Assessment Scale (FAS)

©2023 Vielight Inc.
All rights reserved

BRANY
IRB APPROVED
08/24/2023

#### Fatigue Assessment Scale (FAS)

The following ten statements refer to how you usually feel. Per statement you can choose one out of five answer categories, varying from Never to Always.
Please circle the answer to each question that is applicable to you. Please give an answer to each question, even if you do not have any complaints at the moment.

1. **Never**
2. **Sometimes** (about monthly or less)
3. **Regularly** (about a few times a month)
4. **Often** (about weekly)
5. **Always** (about every day)

| | | Never | Sometimes | Regularly | Often | Always |
|---|---|---|---|---|---|---|
| 1. | I am bothered by fatigue | ○ | ○ | ○ | ○ | ○ |
| 2. | I get tired very quickly | ○ | ○ | ○ | ○ | ○ |
| 3. | I don't do much during the day | ○ | ○ | ○ | ○ | ○ |
| 4. | I have enough energy for everyday life | ○ | ○ | ○ | ○ | ○ |
| 5. | Physically, I feel exhausted | ○ | ○ | ○ | ○ | ○ |
| 6. | I have problems to start things | ○ | ○ | ○ | ○ | ○ |
| 7. | I have problems to think clearly | ○ | ○ | ○ | ○ | ○ |
| 8. | I feel no desire to do anything | ○ | ○ | ○ | ○ | ○ |
| 9. | Mentally, I feel exhausted | ○ | ○ | ○ | ○ | ○ |
| 10. | When I am doing something, I can concentrate quite well | ○ | ○ | ○ | ○ | ○ |

#### Appendix 5

PDQ-20

©2023 Vielight Inc.
All rights reserved

BRANY
IRB APPROVED
08/24/2023

#### PERCEIVED DEFICIT QUESTIONNAIRE (PDQ)

**Instructions**: Everyone at some point experiences problems with memory, attention, or concentration. We are interested in examining how frequently individuals experience these types of problems. The following questions describe several situations in which a person may encounter problems with memory, attention and concentration. For each question, please circle a number from 0 to 4 to indicate how frequently you have experienced any of these matters during the **past week**. Please answer every question. If you are not sure which answer to select, please choose the one answer that comes closest to describing your experiences.

| During the past week, how often did you… | **Never** | **Rarely** | **Sometimes** | **Often** | **Almost always** |
|---|---|---|---|---|---|
| 1. lose your train of thought when speaking? | | | | | |
| 2. have difficulty remembering the names of people even if you have met them several times? | | | | | |
| 3. forget what you came into the room for? | | | | | |
| 4. have trouble getting things organized? | | | | | |
| 5. have trouble concentrating on what people are saying during a conversation? | | | | | |
| 6. forget if you had already done something? | | | | | |
| 7. miss appointments and meetings you had scheduled? | | | | | |
| 8. have difficulty planning what to do during the day? | | | | | |

©2023 Vielight Inc.
All rights reserved

BRANY
IRB APPROVED
08/24/2023

| During the past week, how often did you… | Never | Rarely | Sometimes | Often | Almost always |
|---|---|---|---|---|---|
| 9. have trouble concentrating on things like watching a television program or reading a book? | | | | | |
| 10. forget what you did the night before? | | | | | |
| 11. forget the date unless you looked it up? | | | | | |
| 12. have trouble getting started, even when you had a lot of things to do? | | | | | |
| 13. find your mind drifting? | | | | | |
| 14. forget what you talked about after a telephone conversation? | | | | | |
| 15. forget to do things like turn off the stove, or lock the door? | | | | | |
| 16. feel like your mind went totally blank? | | | | | |
| 17. have trouble holding phone numbers in your head, even for a few seconds? | | | | | |
| 18. forget what you did last weekend? | | | | | |
| 19. forget to take your medication? | | | | | |
| 20. have trouble making decisions? | | | | | |

©2023 Vielight Inc.
All rights reserved

BRANY
IRB APPROVED
08/24/2023

**Appendix 6**

Modified Symptom Burden Questionnaire

©2023 Vielight Inc.
All rights reserved

BRANY
IRB APPROVED
08/24/2023

### Symptom Burden Questionnaire™

LONG COVID

**Version 1.0**
**September 2021**

| |
|---|
| **Name/ID Number:** |
| **Date of Administration:** |
| **Name of person completing the SBQ™-LC: (if by interview):** |

The Symptom Burden Questionnaire™ for Long COVID (SBQ™-LC) asks for your views about your symptoms and their impact on daily life over **the last 7 days**.

It will take approximately 15-20 minutes to complete all the scales.

For each scale, please answer ALL the questions. Please rest and take breaks if needed.

Thank you for completing this questionnaire.

FUNDED BY
NIHR | National Institute for Health Research

UK Research and Innovation

© 2021 The University of Birmingham | All rights reserved

SBQ™ LONG COVID

### BREATHING

**These questions are about your BREATHING symptoms. For each question, please choose the response that best describes your experience over the last 7 days.**

In the last 7 days, how severe was your **shortness of breath (difficulty breathing) when sitting** at its worst?

- ☐ 0 - None
- ☐ 1 - Mild
- ☐ 2 - Moderate
- ☐ 3 - Severe

In the last 7 days, how severe was your **shortness of breath (difficulty breathing) when climbing a flight of stairs** at its worst?

- ☐ 0 - None
- ☐ 1 - Mild
- ☐ 2 - Moderate
- ☐ 3 - Severe

In the last 7 days, how severe was your **shortness of breath (difficulty breathing) when lying flat** at its worst?

- ☐ 0 - None
- ☐ 1 - Mild
- ☐ 2 - Moderate
- ☐ 3 - Severe

*Please go to the next page*

© 2021 The University of Birmingham | All rights reserved

©2023 Vielight Inc.
All rights reserved

BRANY
IRB APPROVED
08/24/2023

In the last 7 days, did you **wake up at night short of breath**?

- ☐ 0 - No
- ☐ 1 - Yes

In the last 7 days, was your **breathing faster than usual**?

- ☐ 0 - No
- ☐ 1 - Yes

In the last 7 days, how severe was the **tightness of your chest** at its worst?

- ☐ 0 - None
- ☐ 1 - Mild
- ☐ 2 - Moderate
- ☐ 3 - Severe

In the last 7 days, how severe was your **wheezing (noisy breathing)** at its worst?

- ☐ 0 - None
- ☐ 1 - Mild
- ☐ 2 - Moderate
- ☐ 3 - Severe

---

| **Breathing Scale Raw Score:** |
|---|

© 2021 The University of Birmingham | All rights reserved

©2023 Vielight Inc.
All rights reserved

BRANY
IRB APPROVED
08/24/2023

### PAIN

**These questions are about your PAIN symptoms. For each question, please choose the response that best describes your experience over the last 7 days.**

In the last 7 days, how severe was your **chest pain** at its worst?

- ☐ 0 - None
- ☐ 1 - Mild
- ☐ 2 - Moderate
- ☐ 3 - Severe

In the last 7 days, how severe was your **pain on breathing** at its worst?

- ☐ 0 - None
- ☐ 1 - Mild
- ☐ 2 - Moderate
- ☐ 3 - Severe

In the last 7 days, how severe was your **shooting or stabbing pain** in any place on your body at its worst?

- ☐ 0 - None
- ☐ 1 - Mild
- ☐ 2 - Moderate
- ☐ 3 - Severe

In the last 7 days, how severe was your **aching or burning pain** in any place on your body at its worst?

- ☐ 0 - None
- ☐ 1 - Mild
- ☐ 2 - Moderate
- ☐ 3 - Severe

---

| |
|---|
| **Pain Scale Raw Score:** |

© 2021 The University of Birmingham | All rights reserved

©2023 Vielight Inc.
All rights reserved
BRANY IRB APPROVED 08/24/2023

### FATIGUE

**These questions are about your FATIGUE symptoms. Please answer ALL the questions, thinking about your symptoms over the last 7 days.**

In the last 7 days, how severe was your **fatigue (feeling of physical or mental exhaustion that does not improve with rest)** at its worst?

- ☐ 0 - None
- ☐ 1 - Mild
- ☐ 2 - Moderate
- ☐ 3 - Severe

In the last 7 days, how severe was your **low energy (being interested and wanting to do things but not having the energy)**?

- ☐ 0 - None
- ☐ 1 - Mild
- ☐ 2 - Moderate
- ☐ 3 - Severe

In the last 7 days, how severe was your **tiredness (need for sleep)** at its worst?

- ☐ 0 - None
- ☐ 1 - Mild
- ☐ 2 - Moderate
- ☐ 3 - Severe

In the last 7 days, how severe was the **worsening of your symptoms following simple physical or mental activities** at its worst?

- ☐ 0 - None
- ☐ 1 - Mild
- ☐ 2 - Moderate
- ☐ 3 - Severe

---

| |
|---|
| **Fatigue Scale Raw Score:** |

© 2021 The University of Birmingham | All rights reserved

©2023 Vielight Inc.
All rights reserved

BRANY
IRB APPROVED
08/24/2023

#### MEMORY, THINKING AND COMMUNICATION

**These questions are about your MEMORY, THINKING, AND COMMUNCATION symptoms. Please answer ALL the questions, thinking about your symptoms over the last 7 days.**

In the last 7 days, how severe was your **difficulty remembering things** at its worst?

- ☐ 0 - None
- ☐ 1 - Mild
- ☐ 2 - Moderate
- ☐ 3 - Severe

In the last 7 days, how severe was your **memory loss** at its worst?

- ☐ 0 - None
- ☐ 1 - Mild
- ☐ 2 - Moderate
- ☐ 3 - Severe

In the last 7 days, how severe was your **brain fog (feeling sluggish, jet-lagged, or blanking out)** at its worst?

- ☐ 0 - None
- ☐ 1 - Mild
- ☐ 2 - Moderate
- ☐ 3 - Severe

In the last 7 days, how often did you **feel confused about what was happening around you**?

- ☐ 0 - Never
- ☐ 1 - Rarely
- ☐ 2 - Sometimes
- ☐ 3 - Always

In the last 7 days, how often did you have **difficulty concentrating**?

- ☐ 0 - Never
- ☐ 1 - Rarely
- ☐ 2 - Sometimes
- ☐ 3 - Always

*Please go to the next page*

© 2021 The University of Birmingham | All rights reserved

©2023 Vielight Inc.
All rights reserved

BRANY
IRB APPROVED
08/24/2023

In the last 7 days, how severe was your **difficulty planning** at its worst?

- ☐ 0 - None
- ☐ 1 - Mild
- ☐ 2 - Moderate
- ☐ 3 - Severe

In the last 7 days, how severe was your **word-finding difficulty (unable to think of the word you want to say or write)** at its worst?

- ☐ 0 - None
- ☐ 1 - Mild
- ☐ 2 - Moderate
- ☐ 3 - Severe

In the last 7 days, how severe was your **difficulty understanding what others were saying** at its worst?

- ☐ 0 - None
- ☐ 1 - Mild
- ☐ 2 - Moderate
- ☐ 3 - Severe

In the last 7 days, how severe was your **slurred speech** at its worst?

- ☐ 0 - None
- ☐ 1 - Mild
- ☐ 2 - Moderate
- ☐ 3 - Severe

In the last 7 days, how severe was your **reading difficulty (not related to dyslexia)**?

- ☐ 0 - None
- ☐ 1 - Mild
- ☐ 2 - Moderate
- ☐ 3 - Severe

---

| |
|---|
| **Memory, Thinking & Communication Scale Raw Score:** |

© 2021 The University of Birmingham | All rights reserved

©2023 Vielight Inc.
All rights reserved

BRANY
IRB APPROVED
08/24/2023

#### SLEEP

**These questions are about your SLEEP symptoms. Please answer ALL the questions, thinking about your symptoms over the last 7 days.**

In the last 7 days, how often did you have **problems falling asleep**?

- ☐ 0 - Never
- ☐ 1 - Rarely
- ☐ 2 - Sometimes
- ☐ 3 - Always

In the last 7 days, how often was your **sleep shorter than usual**?

- ☐ 0 - Never
- ☐ 1 - Rarely
- ☐ 2 - Sometimes
- ☐ 3 - Always

In the last 7 days, how often was your **sleep interrupted**?

- ☐ 0 - Never
- ☐ 1 - Rarely
- ☐ 2 - Sometimes
- ☐ 3 - Always

In the last 7 days, how often did you **sleep longer than usual**?

- ☐ 0 - Never
- ☐ 1 - Rarely
- ☐ 2 - Sometimes
- ☐ 3 - Always

| |
|---|
| **Sleep Scale Raw Score:** |

© 2021 The University of Birmingham | All rights reserved

©2023 Vielight Inc.
All rights reserved

BRANY
IRB APPROVED
08/24/2023

#### EARS, NOSE AND THROAT

**These questions are about your EAR, NOSE, AND THROAT symptoms. Please answer ALL the questions, thinking about your symptoms over the last 7 days.**

In the last 7 days, how severe was your **altered sense of smell (foods/objects smelling different to usual)** at its worst?

- ☐ 0 - None
- ☐ 1 - Mild
- ☐ 2 - Moderate
- ☐ 3 - Severe

In the last 7 days, how severe was your **altered sense of taste (foods tasting different to usual)** at its worst?

- ☐ 0 - None
- ☐ 1 - Mild
- ☐ 2 - Moderate
- ☐ 3 - Severe

In the last 7 days, how severe was your **sneezing** at its worst?

- ☐ 0 - None
- ☐ 1 - Mild
- ☐ 2 - Moderate
- ☐ 3 - Severe

In the last 7 days, how severe was your **stuffy or runny nose** at its worst?

- ☐ 0 - None
- ☐ 1 - Mild
- ☐ 2 - Moderate
- ☐ 3 - Severe

In the last 7 days, how severe was your **sinus congestion (discomfort or feeling of 'fullness' around nose, cheeks, forehead, or around the eyes)** at its worst?

- ☐ 0 - None
- ☐ 1 - Mild
- ☐ 2 - Moderate
- ☐ 3 - Severe

*Please go to the next page*

© 2021 The University of Birmingham | All rights reserved

©2023 Vielight Inc.
All rights reserved

BRANY
IRB APPROVED
08/24/2023

In the last 7 days, how severe was your **production of mucus (phlegm)** at its worst?

- ☐ 0 - None
- ☐ 1 - Mild
- ☐ 2 - Moderate
- ☐ 3 - Severe

In the last 7 days, how severe was your **cough** at its worst?

- ☐ 0 - None
- ☐ 1 - Mild
- ☐ 2 - Moderate
- ☐ 3 - Severe

In the last 7 days, how severe was your **sore throat** at its worst?

- ☐ 0 - None
- ☐ 1 - Mild
- ☐ 2 - Moderate
- ☐ 3 - Severe

In the last 7 days, how severe was your **hoarse voice (change in your voice quality)** at its worst?

- ☐ 0 - None
- ☐ 1 - Mild
- ☐ 2 - Moderate
- ☐ 3 - Severe

In the last 7 days, did you have **difficulty swallowing food or drink**?

- ☐ 0 - No
- ☐ 1 - Yes

In the last 7 days, how severe was your **earache (ear pain)** at its worst

- ☐ 0 - None
- ☐ 1 - Mild
- ☐ 2 - Moderate
- ☐ 3 - Severe

---

*Please go to the next page*

© 2021 The University of Birmingham | All rights reserved

©2023 Vielight Inc.
All rights reserved

BRANY
IRB APPROVED
08/24/2023

In the last 7 days, did you have ***new* hearing loss**?

☐ 0 - No
☐ 1 - Yes

In the last 7 days, how severe was your **tinnitus (noises or ringing sounds in your ears)** at its worst?

☐ 0 - None
☐ 1 - Mild
☐ 2 - Moderate
☐ 3 - Severe

In the last 7 days, how severe was your **sensitivity to sounds that were not a problem for others (everyday sounds were uncomfortably loud and/or painful) at its worst**?

☐ 0 - None
☐ 1 - Mild
☐ 2 - Moderate
☐ 3 - Severe

---

| |
|---|
| **Ears, Nose & Throat Scale Raw Score:** |

© 2021 The University of Birmingham | All rights reserved

©2023 Vielight Inc.
All rights reserved

BRANY
IRB APPROVED
08/24/2023

#### STOMACH AND DIGESTION

**These questions are about your STOMACH AND DIGESTION symptoms. Please answer ALL the questions, thinking about your symptoms over the last 7 days.**

In the last 7 days, how severe was your **belly/tummy pain** at its worst?

- ☐ 0 - None
- ☐ 1 - Mild
- ☐ 2 - Moderate
- ☐ 3 - Severe

In the last 7 days, how severe was the **bloating of your belly/tummy area** at its worst?

- ☐ 0 - None
- ☐ 1 - Mild
- ☐ 2 - Moderate
- ☐ 3 - Severe

In the last 7 days, how severe was your **nausea (urge to vomit)** at its worst.

- ☐ 0 - None
- ☐ 1 - Mild
- ☐ 2 - Moderate
- ☐ 3 - Severe

In the last 7 days, how severe was your **indigestion and/or heartburn** at its worst.

- ☐ 0 - None
- ☐ 1 - Mild
- ☐ 2 - Moderate
- ☐ 3 - Severe

*Please go to the next page*

© 2021 The University of Birmingham | All rights reserved

©2023 Vielight Inc.
All rights reserved

BRANY
IRB APPROVED
08/24/2023

In the last 7 days, have you been worried about your **unplanned weight loss**?

- ☐ 0 - No
- ☐ 1 - Yes

In the last 7 days, have you been worried about your **unplanned weight gain**?

- ☐ 0 - No
- ☐ 1 - Yes

In the last 7 days, how severe was your **diarrhoea** at its worst?

- ☐ 0 - None
- ☐ 1 - Mild
- ☐ 2 - Moderate
- ☐ 3 - Severe

In the last 7 days, how severe was your **constipation (bowel movements happen less often than normal)** at its worst?

- ☐ 0 - None
- ☐ 1 - Mild
- ☐ 2 - Moderate
- ☐ 3 - Severe

---

| |
|---|
| **Stomach & Digestion Scale Raw Score:** |

© 2021 The University of Birmingham | All rights reserved

©2023 Vielight Inc.
All rights reserved

BRANY
IRB APPROVED
08/24/2023

#### OTHER SYMPTOMS

**These questions are about your OTHER SYMPTOMS. Please answer ALL the questions thinking about your symptoms over the last 7 days.**

In the last 7 days, did you have a **fever**?

- ☐ 0 - No
- ☐ 1 - Yes

In the last 7 days, how often did you have **chills/shivering**?

- ☐ 0 - Never
- ☐ 1 - Rarely
- ☐ 2 - Sometimes
- ☐ 3 - Always

In the last 7 days, how severe was your **sweating problem** at its worst?

- ☐ 0 - None
- ☐ 1 - Mild
- ☐ 2 - Moderate
- ☐ 3 - Severe

In the last 7 days, how severe were your **hot flushes** at their worst?

- ☐ 0 - None
- ☐ 1 - Mild
- ☐ 2 - Moderate
- ☐ 3 - Severe

In the last 7 days, how severe was your **aching all over the body** at its worst?

- ☐ 0 - None
- ☐ 1 - Mild
- ☐ 2 - Moderate
- ☐ 3 - Severe

*Please go to the next page*

© 2021 The University of Birmingham | All rights reserved

©2023 Vielight Inc.
All rights reserved

BRANY
IRB APPROVED
08/24/2023

In the last 7 days, how severe was the **swelling of your glands (lymph nodes)** at its worst?

- ☐ 0 - None
- ☐ 1 - Mild
- ☐ 2 - Moderate
- ☐ 3 - Severe

In the last 7 days, how severe was your **vertigo (when everything around you was spinning enough to affect your balance)** at its worst?

- ☐ 0 - None
- ☐ 1 - Mild
- ☐ 2 - Moderate
- ☐ 3 - Severe

In the last 7 days, did you have **swelling of your face, lips, tongue, and/or throat**?

- ☐ 0 - No
- ☐ 1 - Yes

In the last 7 days, did you experience a **heightened reaction to known allergies**?

- ☐ 0 - No
- ☐ 1 - Yes

In the last 7 days, did you experience a **heightened reaction to new allergies**?

- ☐ 0 - No
- ☐ 1 - Yes

In the last 7 days, did you have **loss of control of urine (leakage)**?

- ☐ 0 - No
- ☐ 1 - Yes

In the last 7 days, did you have **difficulty passing urine**?

- ☐ 0 - No
- ☐ 1 - Yes

---

*Please go to the next page*

© 2021 The University of Birmingham | All rights reserved

©2023 Vielight Inc.
All rights reserved

BRANY
IRB APPROVED
08/24/2023

In the last 7 days, have you been **passing more urine than usual**?

- ☐ 0 - No
- ☐ 1 - Yes

In the last 7 days, how severe was your **increased thirst** at its worst?

- ☐ 0 - None
- ☐ 1 - Mild
- ☐ 2 - Moderate
- ☐ 3 - Severe

In the last 7 days, how severe were your **mouth ulcers** at their worst?

- ☐ 0 - None
- ☐ 1 - Mild
- ☐ 2 - Moderate
- ☐ 3 - Severe

In the last 7 days, did you experience a **worsening of known dental problems**?

- ☐ 0 - No
- ☐ 1 - Yes

In the last 7 days, how severe was your **dry mouth** at its worst?

- ☐ 0 - None
- ☐ 1 - Mild
- ☐ 2 - Moderate
- ☐ 3 - Severe

In the last 7 days, how severe was your **headache** at its worst?

- ☐ 0 - None
- ☐ 1 - Mild
- ☐ 2 - Moderate
- ☐ 3 - Severe

---

| |
|---|
| **Other Symptoms Scale Raw Score:** |

© 2021 The University of Birmingham | All rights reserved

©2023 Vielight Inc.
All rights reserved

BRANY
IRB APPROVED
08/24/2023
