## Supplementary 2: PBM Specifications for "Photobiomodulation for Cognitive Dysfunction (Brain Fog) in Post-COVID-19 Condition: A Randomized Sham-Controlled Pilot Trial"

| ***Specifications*** | |
| --- | --- |
| ***Manufacturer*** | ***Vielight (Toronto, Canada)*** |
| Model | Neuro RX Gamma v3 |
| Number of light sources | 6 |
| Light source type | Light-emitting diode (LED) |
| Primary wavelength | 810 nm |
| Spectral wavelength concentration | Within 20 nm of primary wavelength |
| Operating mode | Pulsed |
| Frequency | 40 Hz |
| Duty cycle | 50% |
| Pulse on duration | 25 ms |
| Aperture diameter | 1 $cm^2$ |
| Beam shape | Circular |
| Beam divergence | 0 degrees on contact |
| Exposure duration | 1200 s * 0.5 (duty cycle) = 600 s |
| Number and frequency of treatment sessions | Once a day for 6 days/week for 8 weeks |

***Light distribution, irradiance, and energy delivered***

| **LED location** | **Target** | **Irradiance** | **Dosimetry delivered** |
| --- | --- | --- | --- |
| Anterior head band | Midline, bilateral medial prefrontal cortex | 75 $mW/cm^2$ | 45 J |
| Posterior head band | Bilateral angular gyrus areas | 2 x100 $mW/cm^2$ | 2 x 60 J |
| Posterior head band | Precuneus | 100 $mW/cm^2$ | 60 J |
| Posterior head band | Cerebellum | 50 $mW/cm^2$ | 30 J |
| Nasal applicator | Orbitofrontal and ventromedial prefrontal cortices | 25 $mW/cm^2$ | 15 J |
| Energy per session | | | 270 J |
| Total energy over 48 days (6 days/week for 8 weeks) | | | 12, 960 J |
