## Supplementary 3: Detailed Results Dataset for "Photobiomodulation for Cognitive Dysfunction (Brain Fog) in Post-COVID-19 Condition: A Randomized Sham-Controlled Pilot Trial"

Table 1
Demography and Baseline Characteristics
(Safety Population)

| Variable | Statistic | Category | Active (N= 23) | Sham (N= 20) | Total (N= 43) | P-Value |
|---|---|---|---|---|---|---|
| | | | Treatment Device Group | | | |
| Gender | N (%) | Female | 17 (73.9) | 16 (80.0) | 33 (76.7) | 0.641 |
| | | Male | 6 (26.1) | 4 (20.0) | 10 (23.3) | |
| Age (years) | N | | 23 | 20 | 43 | 0.365 |
| | Mean (SD) | | 39.2 (10.92) | 42.2 (10.35) | 40.6 (10.64) | |
| | Median(IQR) | | 38 ( 17.0) | 38 ( 14.0) | 38 ( 13.0) | |
| | Min, Max | | ( 23, 61) | ( 29, 65) | ( 23, 65) | |
| Age >=45yr | N (%) | Strata Age < 45 | 17 (73.9) | 14 (70.0) | 31 (72.1) | 0.778 |
| | | Strata Age >=45 | 6 (26.1) | 6 (30.0) | 12 (27.9) | |
| Race | N (%) | White | 22 (95.7) | 18 (90.0) | 40 (93.0) | |
| | | Black | 1 ( 4.3) | 1 ( 5.0) | 2 ( 4.7) | 0.557 |
| | | Other | 0 | 1 ( 5.0) | 1 ( 2.3) | |
| Ethnicity | N (%) | Hispanic/Latino | 1 ( 4.3) | 2 (10.0) | 3 ( 7.0) | 0.473 |
| | | Non Hisp/Latino | 22 (95.7) | 18 (90.0) | 40 (93.0) | |
| Fitzpatrick Score | N (%) | 1 | 7 (30.4) | 6 (30.0) | 13 (30.2) | 0.438 |
| | | 2 | 11 (47.8) | 8 (40.0) | 19 (44.2) | |
| | | 3 | 5 (21.7) | 3 (15.0) | 8 (18.6) | |
| | | 4 | 0 | 2 (10.0) | 2 ( 4.7) | |
| | | 5 | 0 | 1 ( 5.0) | 1 ( 2.3) | |
| Current Smoker | N (%) | No | 23 ( 100) | 20 ( 100) | 43 ( 100) | |
| Former Smoker | N (%) | Yes | 8 (34.8) | 2 (10.0) | 10 (23.3) | |
| | | No | 14 (60.9) | 16 (80.0) | 30 (69.8) | 0.151 |
| | | Unknown | 1 ( 4.3) | 2 (10.0) | 3 ( 7.0) | |
| Height (ft) | N | | 23 | 20 | 43 | 0.767 |
| | Mean (SD) | | 5.5 ( 0.30) | 5.5 ( 0.29) | 5.5 ( 0.29) | |
| | Median(IQR) | | 5.5 ( 0.5) | 5.5 ( 0.3) | 5.5 ( 0.4) | |
| | Min, Max | | ( 5.1, 6.1) | ( 5.0, 6.2) | ( 5.0, 6.2) | |
| Weight (lbs) | N | | 23 | 20 | 43 | 0.984 |
| | Mean (SD) | | 158.9 (39.93) | 159.2 (51.89) | 159.0 (45.31) | |
| | Median(IQR) | | 148.0 ( 60.0) | 146.0 ( 48.5) | 148.0 ( 55.0) | |
| | Min, Max | | ( 105, 265.0) | (99.0, 335.0) | (99.0, 335.0) | |
| BMI (kg/m^2) | N | | 23 | 20 | 43 | 0.951 |
| | Mean (SD) | | 25.3 ( 5.65) | 25.4 ( 7.00) | 25.4 ( 6.23) | |
| | Median(IQR) | | 24.2 ( 9.2) | 24.2 ( 4.6) | 24.2 ( 6.7) | |
| | Min, Max | | (18.6, 41.5) | (17.5, 50.9) | (17.5, 50.9) | |

*p<0.05 by two-sample t-test (continuous) and Mantel-Haenszel Chi-Square test (categorical). Source: Listing 1. 31DEC24, T01_demo_mac

Table 1
Demography and Baseline Characteristics
Age < 45 years (Safety Population)

| Variable | Statistic | Category | Treatment Device Group Active (N= 17) | Sham (N= 14) | Total (N= 31) | P-Value |
|---|---|---|---|---|---|---|
| Gender | N (%) | Female | 14 (82.4) | 11 (78.6) | 25 (80.6) | 0.794 |
| | | Male | 3 (17.6) | 3 (21.4) | 6 (19.4) | |
| Age (years) | N | | 17 | 14 | 31 | 0.237 |
| | Mean (SD) | | 34.0 ( 6.26) | 36.4 ( 4.59) | 35.1 ( 5.62) | |
| | Median(IQR) | | 35 ( 11.0) | 36 ( 4.0) | 35 ( 10.0) | |
| | Min, Max | | ( 23, 44) | ( 29, 44) | ( 23, 44) | |
| Age >=45yr | N (%) | Strata Age < 45 | 17 ( 100) | 14 ( 100) | 31 ( 100) | |
| Race | N (%) | White | 17 ( 100) | 12 (85.7) | 29 (93.5) | |
| | | Black | 0 | 1 ( 7.1) | 1 ( 3.2) | 0.285 |
| | | Other | 0 | 1 ( 7.1) | 1 ( 3.2) | |
| Ethnicity | N (%) | Hispanic/Latino | 1 ( 5.9) | 2 (14.3) | 3 ( 9.7) | 0.438 |
| | | Non Hisp/Latino | 16 (94.1) | 12 (85.7) | 28 (90.3) | |
| Fitzpatrick Score | N (%) | 1 | 5 (29.4) | 6 (42.9) | 11 (35.5) | 0.148 |
| | | 2 | 9 (52.9) | 5 (35.7) | 14 (45.2) | |
| | | 3 | 3 (17.6) | 0 | 3 ( 9.7) | |
| | | 4 | 0 | 2 (14.3) | 2 ( 6.5) | |
| | | 5 | 0 | 1 ( 7.1) | 1 ( 3.2) | |
| Current Smoker | N (%) | No | 17 ( 100) | 14 ( 100) | 31 ( 100) | |
| Former Smoker | N (%) | Yes | 6 (35.3) | 1 ( 7.1) | 7 (22.6) | |
| | | No | 10 (58.8) | 11 (78.6) | 21 (67.7) | 0.167 |
| | | Unknown | 1 ( 5.9) | 2 (14.3) | 3 ( 9.7) | |
| Height (ft) | N | | 17 | 14 | 31 | 0.377 |
| | Mean (SD) | | 5.4 ( 0.24) | 5.5 ( 0.32) | 5.5 ( 0.28) | |
| | Median(IQR) | | 5.5 ( 0.3) | 5.5 ( 0.3) | 5.5 ( 0.3) | |
| | Min, Max | | ( 5.1, 5.9) | ( 5.0, 6.2) | ( 5.0, 6.2) | |
| Weight (lbs) | N | | 17 | 14 | 31 | 0.128 |
| | Mean (SD) | | 139.8 (20.39) | 164.6 (61.35) | 151.0 (44.83) | |
| | Median(IQR) | | 136.0 ( 26.0) | 145.0 ( 66.0) | 142.0 ( 46.0) | |
| | Min, Max | | ( 105, 173.0) | (99.0, 335.0) | (99.0, 335.0) | |
| BMI (kg/m^2) | N | | 17 | 14 | 31 | 0.234 |
| | Mean (SD) | | 23.2 ( 3.95) | 26.0 ( 8.34) | 24.5 ( 6.36) | |
| | Median(IQR) | | 21.3 ( 4.6) | 24.2 ( 6.1) | 23.0 ( 6.6) | |
| | Min, Max | | (18.6, 31.1) | (17.5, 50.9) | (17.5, 50.9) | |

*p<0.05 by two-sample t-test (continuous) and Mantel-Haenszel Chi-Square test (categorical). Source: Listing 1. 31DEC24, T01_demo_mac

Table 1
Demography and Baseline Characteristics
Age >=45 years (Safety Population)

| Variable | Statistic | Category | Active (N= 6) | Sham (N= 6) | Total (N= 12) | P-Value |
|---|---|---|---|---|---|---|
| | | | Treatment Device Group | | | |
| Gender | N (%) | Female | 3 (50.0) | 5 (83.3) | 8 (66.7) | 0.241 |
| | | Male | 3 (50.0) | 1 (16.7) | 4 (33.3) | |
| Age (years) | N | | 6 | 6 | 12 | 0.669 |
| | Mean (SD) | | 54.0 ( 6.66) | 55.7 ( 6.44) | 54.8 ( 6.31) | |
| | Median(IQR) | | 56 ( 14.0) | 56 ( 7.0) | 56 ( 10.5) | |
| | Min, Max | | ( 46, 61) | ( 46, 65) | ( 46, 65) | |
| Age >=45yr | N (%) | Strata Age >=45 | 6 ( 100) | 6 ( 100) | 12 ( 100) | |
| Race | N (%) | White | 5 (83.3) | 6 ( 100) | 11 (91.7) | |
| | | Black | 1 (16.7) | 0 | 1 ( 8.3) | 0.317 |
| Ethnicity | N (%) | Non Hisp/Latino | 6 ( 100) | 6 ( 100) | 12 ( 100) | |
| Fitzpatrick Score | N (%) | 1 | 2 (33.3) | 0 | 2 (16.7) | 0.333 |
| | | 2 | 2 (33.3) | 3 (50.0) | 5 (41.7) | |
| | | 3 | 2 (33.3) | 3 (50.0) | 5 (41.7) | |
| Current Smoker | N (%) | No | 6 ( 100) | 6 ( 100) | 12 ( 100) | |
| Former Smoker | N (%) | Yes | 2 (33.3) | 1 (16.7) | 3 (25.0) | |
| | | No | 4 (66.7) | 5 (83.3) | 9 (75.0) | 0.523 |
| Height (ft) | N | | 6 | 6 | 12 | 0.061 |
| | Mean (SD) | | 5.8 ( 0.33) | 5.4 ( 0.23) | 5.6 ( 0.33) | |
| | Median(IQR) | | 5.9 ( 0.5) | 5.4 ( 0.2) | 5.5 ( 0.6) | |
| | Min, Max | | ( 5.3, 6.1) | ( 5.2, 5.8) | ( 5.2, 6.1) | |
| Weight (lbs) | N | | 6 | 6 | 12 | <0.001* |
| | Mean (SD) | | 213.0 (30.56) | 146.7 (13.25) | 179.8 (41.28) | |
| | Median(IQR) | | 206.5 ( 45.0) | 146.0 ( 8.0) | 177.5 ( 60.5) | |
| | Min, Max | | ( 185, 265.0) | ( 130, 170.0) | ( 130, 265.0) | |
| BMI (kg/m^2) | N | | 6 | 6 | 12 | 0.015* |
| | Mean (SD) | | 31.3 ( 5.76) | 24.2 ( 1.56) | 27.7 ( 5.47) | |
| | Median(IQR) | | 30.7 ( 6.0) | 24.2 ( 2.8) | 25.9 ( 6.5) | |
| | Min, Max | | (25.1, 41.5) | (22.3, 26.3) | (22.3, 41.5) | |

*p<0.05 by two-sample t-test (continuous) and Mantel-Haenszel Chi-Square test (categorical). Source: Listing 1. 31DEC24, T01_demo_mac

Table 1
Demography and Baseline Characteristics
(Per-Protocol Population)

| Variable | Statistic | Category | Treatment Device Group Active (N= 21) | Sham (N= 20) | Total (N= 41) | P-Value |
|---|---|---|---|---|---|---|
| Gender | N (%) | Female | 16 (76.2) | 16 (80.0) | 32 (78.0) | 0.771 |
| | | Male | 5 (23.8) | 4 (20.0) | 9 (22.0) | |
| Age (years) | N | | 21 | 20 | 41 | 0.368 |
| | Mean (SD) | | 39.1 (11.42) | 42.2 (10.35) | 40.6 (10.89) | |
| | Median(IQR) | | 37 ( 17.0) | 38 ( 14.0) | 38 ( 13.0) | |
| | Min, Max | | ( 23, 61) | ( 29, 65) | ( 23, 65) | |
| Age >=45yr | N (%) | Strata Age < 45 | 15 (71.4) | 14 (70.0) | 29 (70.7) | 0.921 |
| | | Strata Age >=45 | 6 (28.6) | 6 (30.0) | 12 (29.3) | |
| Race | N (%) | White | 20 (95.2) | 18 (90.0) | 38 (92.7) | |
| | | Black | 1 ( 4.8) | 1 ( 5.0) | 2 ( 4.9) | 0.590 |
| | | Other | 0 | 1 ( 5.0) | 1 ( 2.4) | |
| Ethnicity | N (%) | Hispanic/Latino | 1 ( 4.8) | 2 (10.0) | 3 ( 7.3) | 0.525 |
| | | Non Hisp/Latino | 20 (95.2) | 18 (90.0) | 38 (92.7) | |
| Fitzpatrick Score | N (%) | 1 | 7 (33.3) | 6 (30.0) | 13 (31.7) | 0.474 |
| | | 2 | 9 (42.9) | 8 (40.0) | 17 (41.5) | |
| | | 3 | 5 (23.8) | 3 (15.0) | 8 (19.5) | |
| | | 4 | 0 | 2 (10.0) | 2 ( 4.9) | |
| | | 5 | 0 | 1 ( 5.0) | 1 ( 2.4) | |
| Current Smoker | N (%) | No | 21 ( 100) | 20 ( 100) | 41 ( 100) | |
| Former Smoker | N (%) | Yes | 6 (28.6) | 2 (10.0) | 8 (19.5) | |
| | | No | 14 (66.7) | 16 (80.0) | 30 (73.2) | 0.304 |
| | | Unknown | 1 ( 4.8) | 2 (10.0) | 3 ( 7.3) | |
| Height (ft) | N | | 21 | 20 | 41 | 0.949 |
| | Mean (SD) | | 5.5 ( 0.29) | 5.5 ( 0.29) | 5.5 ( 0.29) | |
| | Median(IQR) | | 5.5 ( 0.3) | 5.5 ( 0.3) | 5.5 ( 0.3) | |
| | Min, Max | | ( 5.1, 6.1) | ( 5.0, 6.2) | ( 5.0, 6.2) | |
| Weight (lbs) | N | | 21 | 20 | 41 | 0.997 |
| | Mean (SD) | | 159.1 (41.78) | 159.2 (51.89) | 159.2 (46.38) | |
| | Median(IQR) | | 148.0 ( 60.0) | 146.0 ( 48.5) | 148.0 ( 55.0) | |
| | Min, Max | | ( 105, 265.0) | (99.0, 335.0) | (99.0, 335.0) | |
| BMI (kg/m^2) | N | | 21 | 20 | 41 | 0.926 |
| | Mean (SD) | | 25.6 ( 5.82) | 25.4 ( 7.00) | 25.5 ( 6.34) | |
| | Median(IQR) | | 25.1 ( 9.2) | 24.2 ( 4.6) | 24.4 ( 6.6) | |
| | Min, Max | | (18.6, 41.5) | (17.5, 50.9) | (17.5, 50.9) | |

*p<0.05 by two-sample t-test (continuous) and Mantel-Haenszel Chi-Square test (categorical). Source: Listing 1. 31DEC24, T01_demo_mac

Table 1
Demography and Baseline Characteristics
Age < 45 years (Per-Protocol Population)

| Variable | Statistic | Category | Treatment Device Group: Active (N= 15) | Treatment Device Group: Sham (N= 14) | Treatment Device Group: Total (N= 29) | P-Value |
|---|---|---|---|---|---|---|
| Gender | N (%) | Female | 13 (86.7) | 11 (78.6) | 24 (82.8) | 0.571 |
| | | Male | 2 (13.3) | 3 (21.4) | 5 (17.2) | |
| Age (years) | N | | 15 | 14 | 29 | 0.113 |
| | Mean (SD) | | 33.1 ( 6.09) | 36.4 ( 4.59) | 34.7 ( 5.58) | |
| | Median(IQR) | | 33 ( 11.0) | 36 ( 4.0) | 35 ( 9.0) | |
| | Min, Max | | ( 23, 44) | ( 29, 44) | ( 23, 44) | |
| Age >=45yr | N (%) | Strata Age < 45 | 15 ( 100) | 14 ( 100) | 29 ( 100) | |
| Race | N (%) | White | 15 ( 100) | 12 (85.7) | 27 (93.1) | |
| | | Black | 0 | 1 ( 7.1) | 1 ( 3.4) | 0.329 |
| | | Other | 0 | 1 ( 7.1) | 1 ( 3.4) | |
| Ethnicity | N (%) | Hispanic/Latino | 1 ( 6.7) | 2 (14.3) | 3 (10.3) | 0.508 |
| | | Non Hisp/Latino | 14 (93.3) | 12 (85.7) | 26 (89.7) | |
| Fitzpatrick Score | N (%) | 1 | 5 (33.3) | 6 (42.9) | 11 (37.9) | 0.186 |
| | | 2 | 7 (46.7) | 5 (35.7) | 12 (41.4) | |
| | | 3 | 3 (20.0) | 0 | 3 (10.3) | |
| | | 4 | 0 | 2 (14.3) | 2 ( 6.9) | |
| | | 5 | 0 | 1 ( 7.1) | 1 ( 3.4) | |
| Current Smoker | N (%) | No | 15 ( 100) | 14 ( 100) | 29 ( 100) | |
| Former Smoker | N (%) | Yes | 4 (26.7) | 1 ( 7.1) | 5 (17.2) | |
| | | No | 10 (66.7) | 11 (78.6) | 21 (72.4) | 0.354 |
| | | Unknown | 1 ( 6.7) | 2 (14.3) | 3 (10.3) | |
| Height (ft) | N | | 15 | 14 | 29 | 0.132 |
| | Mean (SD) | | 5.4 ( 0.18) | 5.5 ( 0.32) | 5.4 ( 0.26) | |
| | Median(IQR) | | 5.4 ( 0.3) | 5.5 ( 0.3) | 5.4 ( 0.3) | |
| | Min, Max | | ( 5.1, 5.7) | ( 5.0, 6.2) | ( 5.0, 6.2) | |
| Weight (lbs) | N | | 15 | 14 | 29 | 0.119 |
| | Mean (SD) | | 137.6 (20.49) | 164.6 (61.35) | 150.6 (46.32) | |
| | Median(IQR) | | 135.0 ( 27.0) | 145.0 ( 66.0) | 136.0 ( 46.0) | |
| | Min, Max | | ( 105, 173.0) | (99.0, 335.0) | (99.0, 335.0) | |
| BMI (kg/m^2) | N | | 15 | 14 | 29 | 0.291 |
| | Mean (SD) | | 23.4 ( 4.19) | 26.0 ( 8.34) | 24.6 ( 6.54) | |
| | Median(IQR) | | 21.3 ( 7.6) | 24.2 ( 6.1) | 23.9 ( 6.6) | |
| | Min, Max | | (18.6, 31.1) | (17.5, 50.9) | (17.5, 50.9) | |

*p<0.05 by two-sample t-test (continuous) and Mantel-Haenszel Chi-Square test (categorical). Source: Listing 1. 31DEC24, T01_demo_mac

Table 1
Demography and Baseline Characteristics
Age >=45 years (Per-Protocol Population)

| Variable | Statistic | Category | Treatment Device Group: Active (N= 6) | Treatment Device Group: Sham (N= 6) | Treatment Device Group: Total (N= 12) | P-Value |
|---|---|---|---|---|---|---|
| Gender | N (%) | Female | 3 (50.0) | 5 (83.3) | 8 (66.7) | 0.241 |
| | | Male | 3 (50.0) | 1 (16.7) | 4 (33.3) | |
| Age (years) | N | | 6 | 6 | 12 | 0.669 |
| | Mean (SD) | | 54.0 ( 6.66) | 55.7 ( 6.44) | 54.8 ( 6.31) | |
| | Median(IQR) | | 56 ( 14.0) | 56 ( 7.0) | 56 ( 10.5) | |
| | Min, Max | | ( 46, 61) | ( 46, 65) | ( 46, 65) | |
| Age >=45yr | N (%) | Strata Age >=45 | 6 ( 100) | 6 ( 100) | 12 ( 100) | |
| Race | N (%) | White | 5 (83.3) | 6 ( 100) | 11 (91.7) | |
| | | Black | 1 (16.7) | 0 | 1 ( 8.3) | 0.317 |
| Ethnicity | N (%) | Non Hisp/Latino | 6 ( 100) | 6 ( 100) | 12 ( 100) | |
| Fitzpatrick Score | N (%) | 1 | 2 (33.3) | 0 | 2 (16.7) | 0.333 |
| | | 2 | 2 (33.3) | 3 (50.0) | 5 (41.7) | |
| | | 3 | 2 (33.3) | 3 (50.0) | 5 (41.7) | |
| Current Smoker | N (%) | No | 6 ( 100) | 6 ( 100) | 12 ( 100) | |
| Former Smoker | N (%) | Yes | 2 (33.3) | 1 (16.7) | 3 (25.0) | |
| | | No | 4 (66.7) | 5 (83.3) | 9 (75.0) | 0.523 |
| Height (ft) | N | | 6 | 6 | 12 | 0.061 |
| | Mean (SD) | | 5.8 ( 0.33) | 5.4 ( 0.23) | 5.6 ( 0.33) | |
| | Median(IQR) | | 5.9 ( 0.5) | 5.4 ( 0.2) | 5.5 ( 0.6) | |
| | Min, Max | | ( 5.3, 6.1) | ( 5.2, 5.8) | ( 5.2, 6.1) | |
| Weight (lbs) | N | | 6 | 6 | 12 | <0.001* |
| | Mean (SD) | | 213.0 (30.56) | 146.7 (13.25) | 179.8 (41.28) | |
| | Median(IQR) | | 206.5 ( 45.0) | 146.0 ( 8.0) | 177.5 ( 60.5) | |
| | Min, Max | | ( 185, 265.0) | ( 130, 170.0) | ( 130, 265.0) | |
| BMI (kg/m^2) | N | | 6 | 6 | 12 | 0.015* |
| | Mean (SD) | | 31.3 ( 5.76) | 24.2 ( 1.56) | 27.7 ( 5.47) | |
| | Median(IQR) | | 30.7 ( 6.0) | 24.2 ( 2.8) | 25.9 ( 6.5) | |
| | Min, Max | | (25.1, 41.5) | (22.3, 26.3) | (22.3, 41.5) | |

*p<0.05 by two-sample t-test (continuous) and Mantel-Haenszel Chi-Square test (categorical). Source: Listing 1. 31DEC24, T01_demo_mac

Table 2

Analysis of Variance
EQ5D Treatment Device Comparison in Change from Baseline by Parameter (Per-Protocol Population)

| | | Treatment Device Estimate | | | | | Treatment Device Comparison | | | | | |
|---|---|---|---|---|---|---|---|---|---|---|---|---|
| | | | | | | | Difference | | | | | |
| | | Treatment | | Std | 95% CI | | Comparator | | | 95% CI | | |
| Parameter | Visit | Device | LS Mean | Error | Lower | Upper | Device | LS Mean | Std Error | Lower | Upper | P-Value |
| We would like to know how good or bad your health is TODAY | BASELINE | Active | 51.74 | 4.669 | 47.07 | 56.41 | | | | | | |
| | | Sham | 65.75 | 3.350 | 62.40 | 69.10 | | | | | | |
| | | Active | | | | | Sham | -14.011 | 5.905 | -25.94 | -2.09 | 0.022* |
| | DAY 14 | Active | 1.68 | 2.860 | -4.10 | 7.46 | | | | | | |
| | | Sham | 2.18 | 2.991 | -3.87 | 8.23 | | | | | | |
| | | Active | | | | | Sham | -0.499 | 4.043 | -8.68 | 7.68 | 0.902 |
| | DAY 28 | Active | 4.66 | 3.327 | -2.06 | 11.38 | | | | | | |
| | | Sham | 0.18 | 3.434 | -6.76 | 7.12 | | | | | | |
| | | Active | | | | | Sham | 4.480 | 4.705 | -5.02 | 13.98 | 0.347 |
| | DAY 56 | Active | 3.79 | 3.454 | -3.18 | 10.76 | | | | | | |
| | | Sham | 4.93 | 3.580 | -2.30 | 12.16 | | | | | | |
| | | Active | | | | | Sham | -1.137 | 4.898 | -11.03 | 8.76 | 0.818 |
| | DAY 84 | Active | 4.24 | 3.337 | -2.49 | 10.98 | | | | | | |
| | | Sham | 5.43 | 3.427 | -1.50 | 12.36 | | | | | | |
| | | Active | | | | | Sham | -1.188 | 4.704 | -10.69 | 8.32 | 0.802 |
| Mobility | BASELINE | Active | 2.52 | 0.266 | 2.26 | 2.79 | | | | | | |
| | | Sham | 1.85 | 0.209 | 1.64 | 2.06 | | | | | | |
| | | Active | | | | | Sham | 0.672 | 0.345 | -0.03 | 1.37 | 0.059 |
| | DAY 14 | Active | 0.19 | 0.138 | -0.09 | 0.47 | | | | | | |
| | | Sham | -0.33 | 0.146 | -0.62 | -0.04 | | | | | | |
| | | Active | | | | | Sham | 0.517 | 0.197 | 0.12 | 0.91 | 0.012* |
| | DAY 28 | Active | -0.15 | 0.164 | -0.48 | 0.18 | | | | | | |
| | | Sham | -0.23 | 0.170 | -0.57 | 0.11 | | | | | | |
| | | Active | | | | | Sham | 0.082 | 0.233 | -0.39 | 0.55 | 0.725 |
| | DAY 56 | Active | 0.09 | 0.130 | -0.17 | 0.35 | | | | | | |
| | | Sham | -0.43 | 0.135 | -0.70 | -0.16 | | | | | | |
| | | Active | | | | | Sham | 0.520 | 0.183 | 0.15 | 0.89 | 0.007* |
| | DAY 84 | Active | -0.04 | 0.107 | -0.25 | 0.18 | | | | | | |
| | | Sham | -0.23 | 0.111 | -0.46 | -0.00 | | | | | | |
| | | Active | | | | | Sham | 0.192 | 0.150 | -0.11 | 0.50 | 0.209 |

*p<0.05 by analysis of covariance of the change from baseline to days 14, 28 and 56 in EQ5D parameters and day 84 (no device), with device visit, treatment by visit interaction, age >=45 years (yes, no) and baseline covariate. Confidence interval (CI), Least-squares (LS) mean. Integer scores from 1 (no problem) to 5 (extreme problem). A negative change is in the direction of improvement. VAS score is from 0 (worst) to 100 (best), and a positive change is in the direction of improvement. Source Listing 8. 31DEC24, T02_eq5d

Table 2

Analysis of Variance
EQ5D Treatment Device Comparison in Change from Baseline by Parameter (Per-Protocol Population)

| | | Treatment Device Estimate | | | | | Treatment Device Comparison | | | | | |
|---|---|---|---|---|---|---|---|---|---|---|---|---|
| | | | | | 95% CI | | | Difference | | 95% CI | | |
| Parameter | Visit | Treatment Device | LS Mean | Std Error | Lower | Upper | Comparator Device | LS Mean | Std Error | Lower | Upper | P-Value |
| Self Care | BASELINE | Active | 1.91 | 0.188 | 1.73 | 2.10 | | | | | | |
| | | Sham | 1.45 | 0.153 | 1.30 | 1.60 | | | | | | |
| | | Active | | | | | Sham | 0.463 | 0.247 | -0.04 | 0.96 | 0.068 |
| | DAY 14 | Active | 0.08 | 0.122 | -0.16 | 0.33 | | | | | | |
| | | Sham | -0.22 | 0.130 | -0.48 | 0.04 | | | | | | |
| | | Active | | | | | Sham | 0.303 | 0.172 | -0.05 | 0.65 | 0.087 |
| | DAY 28 | Active | 0.09 | 0.125 | -0.17 | 0.34 | | | | | | |
| | | Sham | -0.17 | 0.131 | -0.44 | 0.10 | | | | | | |
| | | Active | | | | | Sham | 0.256 | 0.177 | -0.10 | 0.61 | 0.155 |
| | DAY 56 | Active | 0.18 | 0.129 | -0.08 | 0.44 | | | | | | |
| | | Sham | -0.17 | 0.135 | -0.44 | 0.10 | | | | | | |
| | | Active | | | | | Sham | 0.351 | 0.182 | -0.02 | 0.72 | 0.061 |
| | DAY 84 | Active | 0.03 | 0.123 | -0.22 | 0.28 | | | | | | |
| | | Sham | -0.27 | 0.129 | -0.53 | -0.01 | | | | | | |
| | | Active | | | | | Sham | 0.304 | 0.173 | -0.05 | 0.65 | 0.087 |
| Usual Activities | BASELINE | Active | 3.17 | 0.257 | 2.92 | 3.43 | | | | | | |
| | | Sham | 2.50 | 0.246 | 2.25 | 2.75 | | | | | | |
| | | Active | | | | | Sham | 0.674 | 0.358 | -0.05 | 1.40 | 0.067 |
| | DAY 14 | Active | -0.20 | 0.124 | -0.45 | 0.05 | | | | | | |
| | | Sham | -0.40 | 0.132 | -0.66 | -0.13 | | | | | | |
| | | Active | | | | | Sham | 0.200 | 0.175 | -0.15 | 0.55 | 0.260 |
| | DAY 28 | Active | -0.20 | 0.188 | -0.58 | 0.18 | | | | | | |
| | | Sham | -0.40 | 0.194 | -0.79 | -0.00 | | | | | | |
| | | Active | | | | | Sham | 0.197 | 0.267 | -0.34 | 0.74 | 0.465 |
| | DAY 56 | Active | -0.20 | 0.143 | -0.49 | 0.09 | | | | | | |
| | | Sham | -0.55 | 0.149 | -0.85 | -0.24 | | | | | | |
| | | Active | | | | | Sham | 0.348 | 0.203 | -0.06 | 0.76 | 0.094 |
| | DAY 84 | Active | -0.25 | 0.151 | -0.55 | 0.06 | | | | | | |
| | | Sham | -0.55 | 0.155 | -0.86 | -0.23 | | | | | | |
| | | Active | | | | | Sham | 0.298 | 0.213 | -0.13 | 0.73 | 0.169 |
| Pain / Discomfort | BASELINE | Active | 2.70 | 0.230 | 2.47 | 2.93 | | | | | | |
| | | Sham | 2.25 | 0.190 | 2.06 | 2.44 | | | | | | |
| | | Active | | | | | Sham | 0.446 | 0.304 | -0.17 | 1.06 | 0.151 |

*p<0.05 by analysis of covariance of the change from baseline to days 14, 28 and 56 in EQ5D parameters and day 84 (no device), with device visit, treatment by visit interaction, age >=45 years (yes, no) and baseline covariate. Confidence interval (CI), Least-squares (LS) mean. Integer scores from 1 (no problem) to 5 (extreme problem). A negative change is in the direction of improvement. VAS score is from 0 (worst) to 100 (best), and a positive change is in the direction of improvement. Source Listing 8. 31DEC24, T02_eq5d

Table 2

Analysis of Variance
EQ5D Treatment Device Comparison in Change from Baseline by Parameter (Per-Protocol Population)

| | | Treatment Device Estimate | | | | | Treatment Device Comparison | | | | | |
|---|---|---|---|---|---|---|---|---|---|---|---|---|
| | | | | | 95% CI | | | Difference | | 95% CI | | |
| Parameter | Visit | Treatment Device | LS Mean | Std Error | Lower | Upper | Comparator Device | LS Mean | Std Error | Lower | Upper | P-Value |
| Pain / Discomfort | DAY 14 | Active | -0.15 | 0.119 | -0.39 | 0.09 | | | | | | |
| | | Sham | -0.17 | 0.124 | -0.42 | 0.08 | | | | | | |
| | | Active | | | | | Sham | 0.016 | 0.167 | -0.32 | 0.35 | 0.923 |
| | DAY 28 | Active | -0.16 | 0.125 | -0.41 | 0.09 | | | | | | |
| | | Sham | -0.22 | 0.128 | -0.48 | 0.04 | | | | | | |
| | | Active | | | | | Sham | 0.057 | 0.174 | -0.29 | 0.41 | 0.743 |
| | DAY 56 | Active | -0.26 | 0.157 | -0.57 | 0.06 | | | | | | |
| | | Sham | -0.07 | 0.161 | -0.39 | 0.26 | | | | | | |
| | | Active | | | | | Sham | -0.189 | 0.220 | -0.63 | 0.26 | 0.397 |
| | DAY 84 | Active | -0.25 | 0.147 | -0.55 | 0.05 | | | | | | |
| | | Sham | -0.02 | 0.148 | -0.32 | 0.28 | | | | | | |
| | | Active | | | | | Sham | -0.232 | 0.205 | -0.65 | 0.18 | 0.263 |
| Anxiety / Depression | BASELINE | Active | 2.00 | 0.189 | 1.81 | 2.19 | | | | | | |
| | | Sham | 1.85 | 0.182 | 1.67 | 2.03 | | | | | | |
| | | Active | | | | | Sham | 0.150 | 0.264 | -0.38 | 0.68 | 0.573 |
| | DAY 14 | Active | -0.21 | 0.103 | -0.41 | 0.00 | | | | | | |
| | | Sham | -0.02 | 0.107 | -0.23 | 0.20 | | | | | | |
| | | Active | | | | | Sham | -0.189 | 0.143 | -0.48 | 0.10 | 0.194 |
| | DAY 28 | Active | -0.14 | 0.123 | -0.39 | 0.11 | | | | | | |
| | | Sham | -0.07 | 0.126 | -0.32 | 0.19 | | | | | | |
| | | Active | | | | | Sham | -0.076 | 0.171 | -0.42 | 0.27 | 0.662 |
| | DAY 56 | Active | 0.01 | 0.126 | -0.25 | 0.26 | | | | | | |
| | | Sham | -0.22 | 0.128 | -0.48 | 0.04 | | | | | | |
| | | Active | | | | | Sham | 0.225 | 0.175 | -0.13 | 0.58 | 0.207 |
| | DAY 84 | Active | 0.00 | 0.161 | -0.33 | 0.33 | | | | | | |
| | | Sham | -0.02 | 0.163 | -0.35 | 0.31 | | | | | | |
| | | Active | | | | | Sham | 0.016 | 0.226 | -0.44 | 0.47 | 0.942 |
| Mean EQ-5D-5L Questions 1-5 | BASELINE | Active | 2.46 | 0.176 | 2.28 | 2.64 | | | | | | |
| | | Sham | 1.98 | 0.141 | 1.84 | 2.12 | | | | | | |
| | | Active | | | | | Sham | 0.481 | 0.230 | 0.02 | 0.95 | 0.043* |
| | DAY 14 | Active | -0.07 | 0.069 | -0.21 | 0.06 | | | | | | |
| | | Sham | -0.21 | 0.073 | -0.36 | -0.06 | | | | | | |

*p<0.05 by analysis of covariance of the change from baseline to days 14, 28 and 56 in EQ5D parameters and day 84 (no device), with device visit, treatment by visit interaction, age >=45 years (yes, no) and baseline covariate. Confidence interval (CI), Least-squares (LS) mean. Integer scores from 1 (no problem) to 5 (extreme problem). A negative change is in the direction of improvement. VAS score is from 0 (worst) to 100 (best), and a positive change is in the direction of improvement. Source Listing 8. 31DEC24, T02_eq5d

Table 2

Analysis of Variance
EQ5D Treatment Device Comparison in Change from Baseline by Parameter (Per-Protocol Population)

| | | Treatment Device Estimate | | | | | Treatment Device Comparison | | | | | |
|---|---|---|---|---|---|---|---|---|---|---|---|---|
| | | | | | 95% CI | | | Difference | | 95% CI | | |
| Parameter | Visit | Treatment Device | LS Mean | Std Error | Lower | Upper | Comparator Device | LS Mean | Std Error | Lower | Upper | P-Value |
| Mean EQ-5D-5L Questions 1-5 | DAY 14 | Active | | | | | Sham | 0.135 | 0.098 | -0.06 | 0.33 | 0.176 |
| | DAY 28 | Active | -0.13 | 0.090 | -0.31 | 0.05 | | | | | | |
| | | Sham | -0.20 | 0.093 | -0.39 | -0.01 | | | | | | |
| | | Active | | | | | Sham | 0.069 | 0.128 | -0.19 | 0.33 | 0.591 |
| | DAY 56 | Active | -0.05 | 0.087 | -0.23 | 0.12 | | | | | | |
| | | Sham | -0.27 | 0.090 | -0.45 | -0.09 | | | | | | |
| | | Active | | | | | Sham | 0.216 | 0.124 | -0.03 | 0.47 | 0.089 |
| | DAY 84 | Active | -0.12 | 0.087 | -0.29 | 0.06 | | | | | | |
| | | Sham | -0.20 | 0.089 | -0.38 | -0.02 | | | | | | |
| | | Active | | | | | Sham | 0.083 | 0.122 | -0.16 | 0.33 | 0.499 |

*p<0.05 by analysis of covariance of the change from baseline to days 14, 28 and 56 in EQ5D parameters and day 84 (no device), with device visit, treatment by visit interaction, age >=45 years (yes, no) and baseline covariate. Confidence interval (CI), Least-squares (LS) mean. Integer scores from 1 (no problem) to 5 (extreme problem). A negative change is in the direction of improvement. VAS score is from 0 (worst) to 100 (best), and a positive change is in the direction of improvement. Source Listing 8. 31DEC24, T02_eq5d

Table 2.1

Analysis of Variance
EQ5D Treatment Device Comparison in Change from Baseline by Parameter
Age < 45 Years (Per-Protocol Population)

| | | Treatment Device Estimate | | | | | Treatment Device Comparison — Difference | | | | | |
|---|---|---|---|---|---|---|---|---|---|---|---|---|
| Parameter | Visit | Treatment Device | LS Mean | Std Error | 95% CI Lower | 95% CI Upper | Comparator Device | LS Mean | Std Error | 95% CI Lower | 95% CI Upper | P-Value |
| We would like to know how good or bad your health is TODAY | BASELINE | Active | 52.33 | 6.150 | 46.18 | 58.48 | | | | | | |
| | | Sham | 63.57 | 4.304 | 59.27 | 67.88 | | | | | | |
| | | Active | | | | | Sham | -11.238 | 7.607 | -26.85 | 4.37 | 0.151 |
| | DAY 14 | Active | 2.42 | 2.755 | -3.24 | 8.09 | | | | | | |
| | | Sham | 3.86 | 2.861 | -2.02 | 9.74 | | | | | | |
| | | Active | | | | | Sham | -1.433 | 4.058 | -9.77 | 6.90 | 0.727 |
| | DAY 28 | Active | 6.76 | 4.016 | -1.48 | 15.00 | | | | | | |
| | | Sham | -0.79 | 4.164 | -9.33 | 7.76 | | | | | | |
| | | Active | | | | | Sham | 7.543 | 5.844 | -4.44 | 19.52 | 0.208 |
| | DAY 56 | Active | 4.42 | 4.165 | -4.12 | 12.96 | | | | | | |
| | | Sham | 4.93 | 4.317 | -3.92 | 13.78 | | | | | | |
| | | Active | | | | | Sham | -0.504 | 6.055 | -12.91 | 11.90 | 0.934 |
| | DAY 84 | Active | 6.52 | 3.633 | -0.94 | 13.98 | | | | | | |
| | | Sham | 5.64 | 3.696 | -1.96 | 13.25 | | | | | | |
| | | Active | | | | | Sham | 0.877 | 5.249 | -9.90 | 11.65 | 0.869 |
| Mobility | BASELINE | Active | 2.60 | 0.349 | 2.25 | 2.95 | | | | | | |
| | | Sham | 2.07 | 0.267 | 1.80 | 2.34 | | | | | | |
| | | Active | | | | | Sham | 0.529 | 0.444 | -0.38 | 1.44 | 0.244 |
| | DAY 14 | Active | 0.12 | 0.175 | -0.24 | 0.48 | | | | | | |
| | | Sham | -0.42 | 0.181 | -0.79 | -0.05 | | | | | | |
| | | Active | | | | | Sham | 0.535 | 0.254 | 0.02 | 1.06 | 0.044* |
| | DAY 28 | Active | -0.22 | 0.187 | -0.60 | 0.17 | | | | | | |
| | | Sham | -0.49 | 0.194 | -0.89 | -0.09 | | | | | | |
| | | Active | | | | | Sham | 0.273 | 0.271 | -0.28 | 0.83 | 0.323 |
| | DAY 56 | Active | 0.05 | 0.161 | -0.28 | 0.38 | | | | | | |
| | | Sham | -0.56 | 0.167 | -0.90 | -0.22 | | | | | | |
| | | Active | | | | | Sham | 0.611 | 0.234 | 0.13 | 1.09 | 0.014* |
| | DAY 84 | Active | -0.12 | 0.135 | -0.40 | 0.16 | | | | | | |
| | | Sham | -0.42 | 0.137 | -0.70 | -0.14 | | | | | | |
| | | Active | | | | | Sham | 0.299 | 0.195 | -0.10 | 0.70 | 0.138 |

*p<0.05 by analysis of covariance of the change from baseline to days 14, 28 and 56 in EQ5D parameters and day 84 (no device), with device visit, treatment by visit interaction, and baseline covariate. Confidence interval (CI), Least-squares (LS) mean.
Integer scores from 1 (no problem) to 5 (extreme problem). A negative change is in the direction of improvement. VAS score is from 0 (worst) to 100 (best), and a positive change is in the direction of Active improvement. Source Listing 8. 31DEC24, T02_eq5d_LT45

Table 2.1

Analysis of Variance
EQ5D Treatment Device Comparison in Change from Baseline by Parameter
Age < 45 Years (Per-Protocol Population)

| | | Treatment Device Estimate | | | | | Treatment Device Comparison | | | | | |
|---|---|---|---|---|---|---|---|---|---|---|---|---|
| | | | | | | | Difference | | | | | |
| | | | | | 95% CI | | | | | 95% CI | | |
| Parameter | Visit | Treatment Device | LS Mean | Std Error | Lower | Upper | Comparator Device | LS Mean | Std Error | Lower | Upper | P-Value |
| Self Care | BASELINE | Active | 2.07 | 0.228 | 1.84 | 2.29 | | | | | | |
| | | Sham | 1.57 | 0.202 | 1.37 | 1.77 | | | | | | |
| | | Active | | | | | Sham | 0.495 | 0.306 | -0.13 | 1.12 | 0.118 |
| | DAY 14 | Active | 0.09 | 0.152 | -0.22 | 0.40 | | | | | | |
| | | Sham | -0.24 | 0.158 | -0.56 | 0.09 | | | | | | |
| | | Active | | | | | Sham | 0.326 | 0.223 | -0.13 | 0.78 | 0.156 |
| | DAY 28 | Active | 0.02 | 0.149 | -0.28 | 0.33 | | | | | | |
| | | Sham | -0.24 | 0.155 | -0.56 | 0.08 | | | | | | |
| | | Active | | | | | Sham | 0.259 | 0.219 | -0.19 | 0.71 | 0.247 |
| | DAY 56 | Active | 0.09 | 0.153 | -0.23 | 0.40 | | | | | | |
| | | Sham | -0.24 | 0.159 | -0.56 | 0.09 | | | | | | |
| | | Active | | | | | Sham | 0.326 | 0.225 | -0.14 | 0.79 | 0.159 |
| | DAY 84 | Active | -0.05 | 0.146 | -0.35 | 0.25 | | | | | | |
| | | Sham | -0.31 | 0.149 | -0.61 | -0.00 | | | | | | |
| | | Active | | | | | Sham | 0.257 | 0.213 | -0.18 | 0.69 | 0.238 |
| Usual Activities | BASELINE | Active | 3.40 | 0.335 | 3.06 | 3.74 | | | | | | |
| | | Sham | 2.64 | 0.308 | 2.34 | 2.95 | | | | | | |
| | | Active | | | | | Sham | 0.757 | 0.457 | -0.18 | 1.69 | 0.109 |
| | DAY 14 | Active | -0.32 | 0.145 | -0.62 | -0.03 | | | | | | |
| | | Sham | -0.44 | 0.150 | -0.75 | -0.13 | | | | | | |
| | | Active | | | | | Sham | 0.120 | 0.213 | -0.32 | 0.56 | 0.576 |
| | DAY 28 | Active | -0.39 | 0.236 | -0.87 | 0.09 | | | | | | |
| | | Sham | -0.37 | 0.245 | -0.87 | 0.13 | | | | | | |
| | | Active | | | | | Sham | -0.018 | 0.343 | -0.72 | 0.68 | 0.959 |
| | DAY 56 | Active | -0.32 | 0.159 | -0.65 | 0.00 | | | | | | |
| | | Sham | -0.59 | 0.166 | -0.93 | -0.25 | | | | | | |
| | | Active | | | | | Sham | 0.263 | 0.234 | -0.22 | 0.74 | 0.270 |
| | DAY 84 | Active | -0.39 | 0.188 | -0.78 | -0.01 | | | | | | |
| | | Sham | -0.66 | 0.190 | -1.05 | -0.27 | | | | | | |
| | | Active | | | | | Sham | 0.263 | 0.270 | -0.29 | 0.82 | 0.339 |
| Pain / Discomfort | BASELINE | Active | 2.73 | 0.300 | 2.43 | 3.03 | | | | | | |
| | | Sham | 2.29 | 0.244 | 2.04 | 2.53 | | | | | | |

*p<0.05 by analysis of covariance of the change from baseline to days 14, 28 and 56 in EQ5D parameters and day 84 (no device), with device visit, treatment by visit interaction, and baseline covariate. Confidence interval (CI), Least-squares (LS) mean.
Integer scores from 1 (no problem) to 5 (extreme problem). A negative change is in the direction of improvement. VAS score is from 0 (worst) to 100 (best), and a positive change is in the direction of Active improvement. Source Listing 8. 31DEC24, T02_eq5d_LT45

Table 2.1

Analysis of Variance
EQ5D Treatment Device Comparison in Change from Baseline by Parameter
Age < 45 Years (Per-Protocol Population)

| | | Treatment Device Estimate | | | | | Treatment Device Comparison – Difference | | | | | |
|---|---|---|---|---|---|---|---|---|---|---|---|---|
| Parameter | Visit | Treatment Device | LS Mean | Std Error | 95% CI Lower | 95% CI Upper | Comparator Device | LS Mean | Std Error | 95% CI Lower | 95% CI Upper | P-Value |
| Pain / Discomfort | BASELINE | Active | | | | | Sham | 0.448 | 0.390 | -0.35 | 1.25 | 0.262 |
| | DAY 14 | Active | -0.27 | 0.140 | -0.56 | 0.01 | | | | | | |
| | | Sham | -0.21 | 0.145 | -0.51 | 0.09 | | | | | | |
| | | Active | | | | | Sham | -0.062 | 0.203 | -0.48 | 0.35 | 0.761 |
| | DAY 28 | Active | -0.27 | 0.142 | -0.56 | 0.02 | | | | | | |
| | | Sham | -0.28 | 0.147 | -0.58 | 0.02 | | | | | | |
| | | Active | | | | | Sham | 0.009 | 0.206 | -0.41 | 0.43 | 0.966 |
| | DAY 56 | Active | -0.41 | 0.170 | -0.76 | -0.06 | | | | | | |
| | | Sham | -0.07 | 0.176 | -0.43 | 0.29 | | | | | | |
| | | Active | | | | | Sham | -0.339 | 0.246 | -0.84 | 0.17 | 0.180 |
| | DAY 84 | Active | -0.40 | 0.187 | -0.78 | -0.01 | | | | | | |
| | | Sham | -0.07 | 0.188 | -0.45 | 0.32 | | | | | | |
| | | Active | | | | | Sham | -0.329 | 0.266 | -0.88 | 0.22 | 0.227 |
| Anxiety / Depression | BASELINE | Active | 1.80 | 0.223 | 1.58 | 2.02 | | | | | | |
| | | Sham | 1.86 | 0.254 | 1.60 | 2.11 | | | | | | |
| | | Active | | | | | Sham | -0.057 | 0.336 | -0.75 | 0.63 | 0.866 |
| | DAY 14 | Active | -0.34 | 0.109 | -0.56 | -0.12 | | | | | | |
| | | Sham | -0.13 | 0.113 | -0.37 | 0.10 | | | | | | |
| | | Active | | | | | Sham | -0.206 | 0.157 | -0.53 | 0.12 | 0.199 |
| | DAY 28 | Active | -0.21 | 0.144 | -0.50 | 0.09 | | | | | | |
| | | Sham | -0.13 | 0.149 | -0.44 | 0.17 | | | | | | |
| | | Active | | | | | Sham | -0.073 | 0.207 | -0.50 | 0.35 | 0.727 |
| | DAY 56 | Active | -0.07 | 0.120 | -0.32 | 0.17 | | | | | | |
| | | Sham | -0.28 | 0.124 | -0.53 | -0.02 | | | | | | |
| | | Active | | | | | Sham | 0.203 | 0.173 | -0.15 | 0.56 | 0.250 |
| | DAY 84 | Active | -0.20 | 0.160 | -0.53 | 0.13 | | | | | | |
| | | Sham | -0.13 | 0.163 | -0.47 | 0.20 | | | | | | |
| | | Active | | | | | Sham | -0.063 | 0.229 | -0.53 | 0.41 | 0.784 |
| Mean EQ-5D-5L Questions 1-5 | BASELINE | Active | 2.52 | 0.227 | 2.29 | 2.75 | | | | | | |
| | | Sham | 2.09 | 0.184 | 1.90 | 2.27 | | | | | | |
| | | Active | | | | | Sham | 0.434 | 0.294 | -0.17 | 1.04 | 0.151 |

*p<0.05 by analysis of covariance of the change from baseline to days 14, 28 and 56 in EQ5D parameters and day 84 (no device), with device visit, treatment by visit interaction, and baseline covariate. Confidence interval (CI), Least-squares (LS) mean.
Integer scores from 1 (no problem) to 5 (extreme problem). A negative change is in the direction of improvement. VAS score is from 0 (worst) to 100 (best), and a positive change is in the direction of Active improvement. Source Listing 8. 31DEC24, T02_eq5d_LT45

Table 2.1

Analysis of Variance
EQ5D Treatment Device Comparison in Change from Baseline by Parameter
Age < 45 Years (Per-Protocol Population)

| | | Treatment Device Estimate | | | | | Treatment Device Comparison | | | | | |
|---|---|---|---|---|---|---|---|---|---|---|---|---|
| | | | | | 95% CI | | | Difference | | 95% CI | | |
| Parameter | Visit | Treatment Device | LS Mean | Std Error | Lower | Upper | Comparator Device | LS Mean | Std Error | Lower | Upper | P-Value |
| Mean EQ-5D-5L Questions 1-5 | DAY 14 | Active | -0.16 | 0.080 | -0.32 | 0.01 | | | | | | |
| | | Sham | -0.28 | 0.083 | -0.45 | -0.11 | | | | | | |
| | | Active | | | | | Sham | 0.120 | 0.117 | -0.12 | 0.36 | 0.316 |
| | DAY 28 | Active | -0.22 | 0.118 | -0.47 | 0.02 | | | | | | |
| | | Sham | -0.29 | 0.122 | -0.54 | -0.04 | | | | | | |
| | | Active | | | | | Sham | 0.067 | 0.171 | -0.28 | 0.42 | 0.696 |
| | DAY 56 | Active | -0.14 | 0.099 | -0.35 | 0.06 | | | | | | |
| | | Sham | -0.33 | 0.103 | -0.54 | -0.12 | | | | | | |
| | | Active | | | | | Sham | 0.190 | 0.144 | -0.11 | 0.49 | 0.198 |
| | DAY 84 | Active | -0.25 | 0.102 | -0.46 | -0.04 | | | | | | |
| | | Sham | -0.31 | 0.104 | -0.52 | -0.09 | | | | | | |
| | | Active | | | | | Sham | 0.058 | 0.147 | -0.24 | 0.36 | 0.695 |

*p<0.05 by analysis of covariance of the change from baseline to days 14, 28 and 56 in EQ5D parameters and day 84 (no device), with device visit, treatment by visit interaction, and baseline covariate. Confidence interval (CI), Least-squares (LS) mean.
Integer scores from 1 (no problem) to 5 (extreme problem). A negative change is in the direction of improvement. VAS score is from 0 (worst) to 100 (best), and a positive change is in the direction of Active improvement. Source Listing 8. 31DEC24, T02_eq5d_LT45

Table 2.2

Analysis of Variance
EQ5D Treatment Device Comparison in Change from Baseline by Parameter
Age >= 45 Years (Per-Protocol Population)

| | | Treatment Device Estimate | | | | | Treatment Device Comparison: Difference | | | | | |
|---|---|---|---|---|---|---|---|---|---|---|---|---|
| Parameter | Visit | Treatment Device | LS Mean | Std Error | 95% CI Lower | 95% CI Upper | Comparator Device | LS Mean | Std Error | 95% CI Lower | 95% CI Upper | P-Value |
| We would like to know how good or bad your health is TODAY | BASELINE | Active | 51.67 | 9.545 | 42.12 | 61.21 | | | | | | |
| | | Sham | 70.83 | 4.729 | 66.10 | 75.56 | | | | | | |
| | | Active | | | | | Sham | -19.167 | 10.652 | -42.90 | 4.57 | 0.102 |
| | DAY 14 | Active | 3.94 | 8.390 | -15.58 | 23.47 | | | | | | |
| | | Sham | -3.94 | 8.390 | -23.47 | 15.58 | | | | | | |
| | | Active | | | | | Sham | 7.885 | 12.423 | -20.67 | 36.44 | 0.543 |
| | DAY 28 | Active | 3.94 | 6.006 | -9.93 | 17.81 | | | | | | |
| | | Sham | 0.22 | 6.006 | -13.65 | 14.09 | | | | | | |
| | | Active | | | | | Sham | 3.719 | 9.257 | -17.46 | 24.90 | 0.698 |
| | DAY 56 | Active | 6.44 | 6.609 | -8.74 | 21.62 | | | | | | |
| | | Sham | 2.72 | 6.609 | -12.46 | 17.91 | | | | | | |
| | | Active | | | | | Sham | 3.719 | 10.045 | -19.12 | 26.55 | 0.720 |
| | DAY 84 | Active | 3.11 | 8.098 | -15.22 | 21.44 | | | | | | |
| | | Sham | 2.72 | 8.098 | -15.61 | 21.06 | | | | | | |
| | | Active | | | | | Sham | 0.385 | 12.029 | -26.59 | 27.36 | 0.975 |
| Mobility | BASELINE | Active | 2.17 | 0.543 | 1.62 | 2.71 | | | | | | |
| | | Sham | 1.33 | 0.211 | 1.12 | 1.54 | | | | | | |
| | | Active | | | | | Sham | 0.833 | 0.582 | -0.46 | 2.13 | 0.183 |
| | DAY 14 | Active | 0.17 | 0.245 | -0.37 | 0.72 | | | | | | |
| | | Sham | -0.17 | 0.245 | -0.72 | 0.37 | | | | | | |
| | | Active | | | | | Sham | 0.349 | 0.347 | -0.42 | 1.12 | 0.338 |
| | DAY 28 | Active | -0.16 | 0.320 | -0.87 | 0.55 | | | | | | |
| | | Sham | 0.33 | 0.320 | -0.39 | 1.04 | | | | | | |
| | | Active | | | | | Sham | -0.485 | 0.453 | -1.49 | 0.52 | 0.310 |
| | DAY 56 | Active | 0.01 | 0.215 | -0.47 | 0.49 | | | | | | |
| | | Sham | -0.17 | 0.215 | -0.65 | 0.30 | | | | | | |
| | | Active | | | | | Sham | 0.182 | 0.304 | -0.50 | 0.86 | 0.563 |
| | DAY 84 | Active | 0.01 | 0.119 | -0.26 | 0.27 | | | | | | |
| | | Sham | 0.16 | 0.119 | -0.11 | 0.42 | | | | | | |
| | | Active | | | | | Sham | -0.151 | 0.170 | -0.53 | 0.23 | 0.394 |

*p<0.05 by analysis of covariance of the change from baseline to days 14, 28 and 56 in EQ5D parameters and day 84 (no device), with device visit, treatment by visit interaction, and baseline covariate. Confidence interval (CI), Least-squares (LS) mean.
Integer scores from 1 (no problem) to 5 (extreme problem). A negative change is in the direction of improvement. VAS score is from 0 (worst) to 100 (best), and a positive change is in the direction of Active improvement. Source Listing 8. 31DEC24, T02_eq5d_ge45

Table 2.2

Analysis of Variance
EQ5D Treatment Device Comparison in Change from Baseline by Parameter
Age >= 45 Years (Per-Protocol Population)

| | | Treatment Device Estimate | | | | | Treatment Device Comparison | | | | | |
|---|---|---|---|---|---|---|---|---|---|---|---|---|
| | | | | | | | | Difference | | | | |
| | | Treatment | | Std | 95% CI | | Comparator | | | 95% CI | | |
| Parameter | Visit | Device | LS Mean | Error | Lower | Upper | Device | LS Mean | Std Error | Lower | Upper | P-Value |
| Self Care | BASELINE | Active | 1.50 | 0.342 | 1.16 | 1.84 | | | | | | |
| | | Sham | 1.17 | 0.167 | 1.00 | 1.33 | | | | | | |
| | | Active | | | | | Sham | 0.333 | 0.380 | -0.51 | 1.18 | 0.401 |
| Usual Activities | BASELINE | Active | 2.67 | 0.422 | 2.25 | 3.09 | | | | | | |
| | | Sham | 2.17 | 0.401 | 1.77 | 2.57 | | | | | | |
| | | Active | | | | | Sham | 0.500 | 0.582 | -0.80 | 1.80 | 0.411 |
| Pain / Discomfort | BASELINE | Active | 2.50 | 0.500 | 2.00 | 3.00 | | | | | | |
| | | Sham | 2.17 | 0.307 | 1.86 | 2.47 | | | | | | |
| | | Active | | | | | Sham | 0.333 | 0.587 | -0.97 | 1.64 | 0.583 |
| | DAY 14 | Active | 0.01 | 0.223 | -0.49 | 0.51 | | | | | | |
| | | Sham | -0.18 | 0.223 | -0.68 | 0.32 | | | | | | |
| | | Active | | | | | Sham | 0.190 | 0.318 | -0.52 | 0.90 | 0.564 |
| | DAY 28 | Active | 0.01 | 0.228 | -0.50 | 0.52 | | | | | | |
| | | Sham | -0.18 | 0.228 | -0.69 | 0.33 | | | | | | |
| | | Active | | | | | Sham | 0.190 | 0.325 | -0.54 | 0.92 | 0.573 |
| | DAY 56 | Active | 0.01 | 0.296 | -0.65 | 0.68 | | | | | | |
| | | Sham | -0.18 | 0.296 | -0.84 | 0.49 | | | | | | |
| | | Active | | | | | Sham | 0.190 | 0.421 | -0.75 | 1.13 | 0.662 |
| | DAY 84 | Active | 0.01 | 0.189 | -0.41 | 0.44 | | | | | | |
| | | Sham | -0.01 | 0.189 | -0.44 | 0.41 | | | | | | |
| | | Active | | | | | Sham | 0.023 | 0.270 | -0.58 | 0.63 | 0.933 |
| Anxiety / Depression | BASELINE | Active | 2.33 | 0.422 | 1.91 | 2.75 | | | | | | |
| | | Sham | 1.83 | 0.167 | 1.67 | 2.00 | | | | | | |
| | | Active | | | | | Sham | 0.500 | 0.453 | -0.51 | 1.51 | 0.296 |
| | DAY 14 | Active | 0.04 | 0.228 | -0.47 | 0.55 | | | | | | |
| | | Sham | 0.13 | 0.228 | -0.38 | 0.64 | | | | | | |
| | | Active | | | | | Sham | -0.088 | 0.332 | -0.83 | 0.65 | 0.797 |
| | DAY 28 | Active | -0.13 | 0.230 | -0.64 | 0.39 | | | | | | |
| | | Sham | -0.04 | 0.230 | -0.55 | 0.47 | | | | | | |
| | | Active | | | | | Sham | -0.088 | 0.335 | -0.84 | 0.66 | 0.799 |
| | DAY 56 | Active | 0.04 | 0.334 | -0.71 | 0.78 | | | | | | |
| | | Sham | -0.21 | 0.334 | -0.95 | 0.54 | | | | | | |

*p<0.05 by analysis of covariance of the change from baseline to days 14, 28 and 56 in EQ5D parameters and day 84 (no device), with device visit, treatment by visit interaction, and baseline covariate. Confidence interval (CI), Least-squares (LS) mean.
Integer scores from 1 (no problem) to 5 (extreme problem). A negative change is in the direction of improvement. VAS score is from 0 (worst) to 100 (best), and a positive change is in the direction of Active improvement. Source Listing 8. 31DEC24, T02_eq5d_ge45

Table 2.2

Analysis of Variance
EQ5D Treatment Device Comparison in Change from Baseline by Parameter
Age >= 45 Years (Per-Protocol Population)

| | | Treatment Device Estimate | | | | | Treatment Device Comparison | | | | | |
|---|---|---|---|---|---|---|---|---|---|---|---|---|
| | | | | | | | | Difference | | | | |
| | | Treatment | | Std | 95% CI | | Comparator | | | 95% CI | | |
| Parameter | Visit | Device | LS Mean | Error | Lower | Upper | Device | LS Mean | Std Error | Lower | Upper | P-Value |
| Anxiety / Depression | DAY 56 | Active | | | | | Sham | 0.246 | 0.479 | -0.82 | 1.31 | 0.619 |
| | DAY 84 | Active | 0.37 | 0.400 | -0.52 | 1.27 | | | | | | |
| | | Sham | 0.13 | 0.400 | -0.77 | 1.02 | | | | | | |
| | | Active | | | | | Sham | 0.246 | 0.572 | -1.03 | 1.52 | 0.677 |
| Mean EQ-5D-5L Questions 1-5 | BASELINE | Active | 2.23 | 0.363 | 1.87 | 2.60 | | | | | | |
| | | Sham | 1.73 | 0.169 | 1.56 | 1.90 | | | | | | |
| | | Active | | | | | Sham | 0.500 | 0.400 | -0.39 | 1.39 | 0.240 |
| | DAY 14 | Active | 0.02 | 0.141 | -0.29 | 0.34 | | | | | | |
| | | Sham | -0.12 | 0.141 | -0.44 | 0.19 | | | | | | |
| | | Active | | | | | Sham | 0.146 | 0.205 | -0.31 | 0.60 | 0.491 |
| | DAY 28 | Active | -0.01 | 0.111 | -0.26 | 0.24 | | | | | | |
| | | Sham | -0.06 | 0.111 | -0.31 | 0.19 | | | | | | |
| | | Active | | | | | Sham | 0.046 | 0.164 | -0.32 | 0.41 | 0.784 |
| | DAY 56 | Active | 0.06 | 0.177 | -0.34 | 0.45 | | | | | | |
| | | Sham | -0.19 | 0.177 | -0.58 | 0.20 | | | | | | |
| | | Active | | | | | Sham | 0.246 | 0.255 | -0.32 | 0.81 | 0.357 |
| | DAY 84 | Active | 0.09 | 0.154 | -0.25 | 0.43 | | | | | | |
| | | Sham | -0.02 | 0.154 | -0.37 | 0.32 | | | | | | |
| | | Active | | | | | Sham | 0.113 | 0.223 | -0.38 | 0.61 | 0.623 |

*p<0.05 by analysis of covariance of the change from baseline to days 14, 28 and 56 in EQ5D parameters and day 84 (no device), with device visit, treatment by visit interaction, and baseline covariate. Confidence interval (CI), Least-squares (LS) mean.
Integer scores from 1 (no problem) to 5 (extreme problem). A negative change is in the direction of improvement. VAS score is from 0 (worst) to 100 (best), and a positive change is in the direction of Active improvement. Source Listing 8. 31DEC24, T02_eq5d_ge45

Table 3

Analysis of Covariance
FAS Treatment Device Comparison in Change from Baseline (Per-Protocol Population)

| | | Treatment Device Estimate | | | | | Treatment Device Comparison Difference | | | | | |
|---|---|---|---|---|---|---|---|---|---|---|---|---|
| Parameter | Visit | Treatment Device | LS Mean | Std Error | 95% CI Lower | 95% CI Upper | Treatment -Device | LS Mean | Std Error | 95% CI Lower | 95% CI Upper | P-Value |
| 1-I am bothered by fatigue | BASELINE | Active | 4.17 | 0.264 | 3.91 | 4.44 | | | | | | |
| | | Sham | 4.00 | 0.229 | 3.77 | 4.23 | | | | | | |
| | | Active | | | | | Sham | 0.17 | 0.355 | -0.54 | 0.89 | 0.627 |
| | DAY 56 | Active | -0.25 | 0.195 | -0.65 | 0.14 | | | | | | |
| | | Sham | -0.83 | 0.199 | -1.23 | -0.42 | | | | | | |
| | | Active | | | | | Sham | 0.57 | 0.266 | 0.03 | 1.11 | 0.038* |
| | DAY 84 | Active | -0.37 | 0.235 | -0.85 | 0.11 | | | | | | |
| | | Sham | -0.33 | 0.237 | -0.81 | 0.15 | | | | | | |
| | | Active | | | | | Sham | -0.04 | 0.325 | -0.70 | 0.61 | 0.894 |
| 2-I get tired very quickly | BASELINE | Active | 4.00 | 0.267 | 3.73 | 4.27 | | | | | | |
| | | Sham | 3.70 | 0.252 | 3.45 | 3.95 | | | | | | |
| | | Active | | | | | Sham | 0.30 | 0.370 | -0.45 | 1.05 | 0.423 |
| | DAY 56 | Active | 0.05 | 0.218 | -0.39 | 0.49 | | | | | | |
| | | Sham | -0.59 | 0.224 | -1.05 | -0.14 | | | | | | |
| | | Active | | | | | Sham | 0.64 | 0.298 | 0.04 | 1.25 | 0.037* |
| | DAY 84 | Active | -0.17 | 0.240 | -0.65 | 0.32 | | | | | | |
| | | Sham | -0.39 | 0.244 | -0.89 | 0.10 | | | | | | |
| | | Active | | | | | Sham | 0.23 | 0.329 | -0.44 | 0.90 | 0.491 |
| 3-I don't do much during the day | BASELINE | Active | 3.35 | 0.312 | 3.04 | 3.66 | | | | | | |
| | | Sham | 2.60 | 0.266 | 2.33 | 2.87 | | | | | | |
| | | Active | | | | | Sham | 0.75 | 0.416 | -0.09 | 1.59 | 0.080 |
| | DAY 56 | Active | -0.03 | 0.187 | -0.40 | 0.35 | | | | | | |
| | | Sham | -0.33 | 0.196 | -0.73 | 0.07 | | | | | | |
| | | Active | | | | | Sham | 0.30 | 0.264 | -0.23 | 0.84 | 0.256 |
| | DAY 84 | Active | -0.19 | 0.187 | -0.57 | 0.19 | | | | | | |
| | | Sham | -0.43 | 0.192 | -0.82 | -0.04 | | | | | | |
| | | Active | | | | | Sham | 0.24 | 0.262 | -0.29 | 0.77 | 0.363 |
| 4-I have enough energy for everyday life | BASELINE | Active | 3.00 | 0.302 | 2.70 | 3.30 | | | | | | |

*p<0.05 by analysis of covariance of the change from baseline to day 56 and day 84 in FAS parameters, with device, visit, treatment by visit interaction, age >=45 years(yes, no) and baseline covariate. Confidence interval (CI), Least-squares (LS) mean.
For Fatigue Assessment Scale (FAS) questions 1-3 and 5-9, a negative change is in the direction of improvement, and for questions 4 and 10, a positive change is in the direction of improvement. For the sum, a negative change is in the direction of improvement, with reverse score for questions 4 and 10 as (5 minus actual score). Source: Listing 10.

31DEC24, T03_fas

Table 3

Analysis of Covariance
FAS Treatment Device Comparison in Change from Baseline (Per-Protocol Population)

| | | Treatment Device Estimate | | | | | Treatment Device Comparison | | | | | |
|---|---|---|---|---|---|---|---|---|---|---|---|---|
| | | | | | | | Difference | | | | | |
| | | | | | 95% CI | | | | | 95% CI | | |
| Parameter | Visit | Treatment Device | LS Mean | Std Error | Lower | Upper | Treatment -Device | LS Mean | Std Error | Lower | Upper | P-Value |
| 4-I have enough energy for everyday life | BASELINE | Sham | 3.35 | 0.150 | 3.20 | 3.50 | | | | | | |
| | | Active | | | | | Sham | -0.35 | 0.353 | -1.06 | 0.36 | 0.327 |
| | DAY 56 | Active | -0.44 | 0.224 | -0.90 | 0.01 | | | | | | |
| | | Sham | -0.83 | 0.228 | -1.29 | -0.37 | | | | | | |
| | | Active | | | | | Sham | 0.38 | 0.311 | -0.25 | 1.01 | 0.224 |
| | DAY 84 | Active | -0.40 | 0.191 | -0.79 | -0.02 | | | | | | |
| | | Sham | -0.68 | 0.190 | -1.06 | -0.29 | | | | | | |
| | | Active | | | | | Sham | 0.27 | 0.260 | -0.25 | 0.80 | 0.300 |
| 5-Physically, I feel exhausted | BASELINE | Active | 3.91 | 0.281 | 3.63 | 4.19 | | | | | | |
| | | Sham | 3.50 | 0.256 | 3.24 | 3.76 | | | | | | |
| | | Active | | | | | Sham | 0.41 | 0.384 | -0.36 | 1.19 | 0.289 |
| | DAY 56 | Active | -0.20 | 0.231 | -0.66 | 0.27 | | | | | | |
| | | Sham | -0.62 | 0.238 | -1.10 | -0.14 | | | | | | |
| | | Active | | | | | Sham | 0.42 | 0.322 | -0.23 | 1.08 | 0.196 |
| | DAY 84 | Active | -0.18 | 0.203 | -0.59 | 0.23 | | | | | | |
| | | Sham | -0.42 | 0.207 | -0.84 | -0.00 | | | | | | |
| | | Active | | | | | Sham | 0.24 | 0.279 | -0.33 | 0.80 | 0.405 |
| 6-I have problems to start things | BASELINE | Active | 3.22 | 0.251 | 2.97 | 3.47 | | | | | | |
| | | Sham | 2.80 | 0.247 | 2.55 | 3.05 | | | | | | |
| | | Active | | | | | Sham | 0.42 | 0.354 | -0.30 | 1.13 | 0.246 |
| | DAY 56 | Active | -0.20 | 0.195 | -0.60 | 0.19 | | | | | | |
| | | Sham | -0.20 | 0.198 | -0.60 | 0.20 | | | | | | |
| | | Active | | | | | Sham | -0.00 | 0.268 | -0.55 | 0.54 | 0.993 |
| | DAY 84 | Active | -0.25 | 0.227 | -0.71 | 0.20 | | | | | | |
| | | Sham | -0.10 | 0.227 | -0.56 | 0.36 | | | | | | |
| | | Active | | | | | Sham | -0.16 | 0.313 | -0.79 | 0.48 | 0.621 |

*p<0.05 by analysis of covariance of the change from baseline to day 56 and day 84 in FAS parameters, with device, visit, treatment by visit interaction, age >=45 years(yes, no) and baseline covariate. Confidence interval (CI), Least-squares (LS) mean.
For Fatigue Assessment Scale (FAS) questions 1-3 and 5-9, a negative change is in the direction of improvement, and for questions 4 and 10, a positive change is in the direction of improvement. For the sum, a negative change is in the direction of improvement, with reverse score for questions 4 and 10 as (5 minus actual score). Source: Listing 10.

31DEC24, T03_fas

Table 3

Analysis of Covariance
FAS Treatment Device Comparison in Change from Baseline (Per-Protocol Population)

| Parameter | Visit | Treatment Device Estimate: Treatment Device | Treatment Device Estimate: LS Mean | Treatment Device Estimate: Std Error | Treatment Device Estimate: 95% CI Lower | Treatment Device Estimate: 95% CI Upper | Treatment Device Comparison: Difference: Treatment -Device | Treatment Device Comparison: Difference: LS Mean | Treatment Device Comparison: Std Error | Treatment Device Comparison: 95% CI Lower | Treatment Device Comparison: 95% CI Upper | Treatment Device Comparison: P-Value |
|---|---|---|---|---|---|---|---|---|---|---|---|---|
| 7-I have problems to think clearly | BASELINE | Active | 3.87 | 0.229 | 3.64 | 4.10 | | | | | | |
| | | Sham | 3.70 | 0.179 | 3.52 | 3.88 | | | | | | |
| | | Active | | | | | Sham | 0.17 | 0.297 | -0.43 | 0.77 | 0.572 |
| | DAY 56 | Active | -0.46 | 0.204 | -0.88 | -0.05 | | | | | | |
| | | Sham | -0.90 | 0.208 | -1.32 | -0.48 | | | | | | |
| | | Active | | | | | Sham | 0.44 | 0.279 | -0.13 | 1.00 | 0.123 |
| | DAY 84 | Active | -0.67 | 0.222 | -1.11 | -0.22 | | | | | | |
| | | Sham | -0.60 | 0.224 | -1.05 | -0.15 | | | | | | |
| | | Active | | | | | Sham | -0.06 | 0.304 | -0.68 | 0.55 | 0.833 |
| 8-I feel no desire to do anything | BASELINE | Active | 2.09 | 0.208 | 1.88 | 2.29 | | | | | | |
| | | Sham | 2.30 | 0.231 | 2.07 | 2.53 | | | | | | |
| | | Active | | | | | Sham | -0.21 | 0.310 | -0.84 | 0.41 | 0.495 |
| | DAY 56 | Active | -0.03 | 0.153 | -0.34 | 0.28 | | | | | | |
| | | Sham | -0.30 | 0.157 | -0.62 | 0.02 | | | | | | |
| | | Active | | | | | Sham | 0.26 | 0.210 | -0.16 | 0.69 | 0.215 |
| | DAY 84 | Active | -0.04 | 0.182 | -0.40 | 0.33 | | | | | | |
| | | Sham | -0.05 | 0.184 | -0.42 | 0.32 | | | | | | |
| | | Active | | | | | Sham | 0.01 | 0.251 | -0.49 | 0.52 | 0.958 |
| 9-Mentally, I feel exhausted | BASELINE | Active | 3.65 | 0.271 | 3.38 | 3.92 | | | | | | |
| | | Sham | 3.60 | 0.210 | 3.39 | 3.81 | | | | | | |
| | | Active | | | | | Sham | 0.05 | 0.351 | -0.66 | 0.76 | 0.883 |
| | DAY 56 | Active | -0.30 | 0.206 | -0.72 | 0.11 | | | | | | |
| | | Sham | -0.71 | 0.210 | -1.14 | -0.29 | | | | | | |
| | | Active | | | | | Sham | 0.41 | 0.281 | -0.16 | 0.98 | 0.153 |
| | DAY 84 | Active | -0.26 | 0.211 | -0.68 | 0.17 | | | | | | |
| | | Sham | -0.56 | 0.213 | -0.99 | -0.13 | | | | | | |
| | | Active | | | | | Sham | 0.31 | 0.288 | -0.28 | 0.89 | 0.294 |

*p<0.05 by analysis of covariance of the change from baseline to day 56 and day 84 in FAS parameters, with device, visit, treatment by visit interaction, age >=45 years(yes, no) and baseline covariate. Confidence interval (CI), Least-squares (LS) mean.
For Fatigue Assessment Scale (FAS) questions 1-3 and 5-9, a negative change is in the direction of improvement, and for questions 4 and 10, a positive change is in the direction of improvement. For the sum, a negative change is in the direction of improvement, with reverse score for questions 4 and 10 as (5 minus actual score). Source: Listing 10.

31DEC24, T03_fas

Table 3

Analysis of Covariance
FAS Treatment Device Comparison in Change from Baseline (Per-Protocol Population)

| Parameter | Visit | Treatment Device | LS Mean | Std Error | 95% CI Lower | 95% CI Upper | Treatment -Device | Difference LS Mean | Difference Std Error | 95% CI Lower | 95% CI Upper | P-Value |
|---|---|---|---|---|---|---|---|---|---|---|---|---|
| | | Treatment Device Estimate | | | | | Treatment Device Comparison | | | | | |
| 10-When I am doing something, I can concentrate quite well | BASELINE | Active | 2.74 | 0.180 | 2.56 | 2.92 | | | | | | |
| | | Sham | 2.85 | 0.182 | 2.67 | 3.03 | | | | | | |
| | | Active | | | | | Sham | -0.11 | 0.257 | -0.63 | 0.41 | 0.669 |
| | DAY 56 | Active | -0.51 | 0.220 | -0.96 | -0.07 | | | | | | |
| | | Sham | -0.36 | 0.224 | -0.81 | 0.10 | | | | | | |
| | | Active | | | | | Sham | -0.15 | 0.299 | -0.76 | 0.45 | 0.608 |
| | DAY 84 | Active | -0.47 | 0.231 | -0.94 | -0.00 | | | | | | |
| | | Sham | -0.21 | 0.233 | -0.68 | 0.27 | | | | | | |
| | | Active | | | | | Sham | -0.26 | 0.315 | -0.90 | 0.38 | 0.410 |
| Sum FAS Questions 1-10 | BASELINE | Active | 34.00 | 1.987 | 32.01 | 35.99 | | | | | | |
| | | Sham | 32.40 | 1.411 | 30.99 | 33.81 | | | | | | |
| | | Active | | | | | Sham | 1.60 | 2.506 | -3.46 | 6.66 | 0.527 |
| | DAY 56 | Active | -2.51 | 1.269 | -5.08 | 0.06 | | | | | | |
| | | Sham | -5.37 | 1.299 | -8.00 | -2.75 | | | | | | |
| | | Active | | | | | Sham | 2.87 | 1.738 | -0.66 | 6.39 | 0.108 |
| | DAY 84 | Active | -3.12 | 1.269 | -5.69 | -0.54 | | | | | | |
| | | Sham | -3.47 | 1.287 | -6.08 | -0.86 | | | | | | |
| | | Active | | | | | Sham | 0.36 | 1.733 | -3.15 | 3.87 | 0.837 |

*p<0.05 by analysis of covariance of the change from baseline to day 56 and day 84 in FAS parameters, with device, visit, treatment by visit interaction, age >=45 years(yes, no) and baseline covariate. Confidence interval (CI), Least-squares (LS) mean.
For Fatigue Assessment Scale (FAS) questions 1-3 and 5-9, a negative change is in the direction of improvement, and for questions 4 and 10, a positive change is in the direction of improvement. For the sum, a negative change is in the direction of improvement, with reverse score for questions 4 and 10 as (5 minus actual score). Source: Listing 10.

31DEC24, T03_fas

Table 3.1

Analysis of Covariance
FAS Treatment Device Comparison in Change from Baseline
Age < 45 Years (Per-Protocol Population)

| Parameter | Day | Treatment Device | LS Mean | Std Error | 95% CI Lower | 95% CI Upper | Treatment -Device | Difference LS Mean | Difference Std Error | Difference 95% CI Lower | Difference 95% CI Upper | P-Value |
|---|---|---|---|---|---|---|---|---|---|---|---|---|
| 1-I am bothered by fatigue | 0 | Active | 4.27 | 0.316 | 3.95 | 4.58 | | | | | | |
| | | Sham | 4.14 | 0.275 | 3.87 | 4.42 | | | | | | |
| | | Active | | | | | Sham | 0.12 | 0.421 | -0.74 | 0.99 | 0.771 |
| | 56 | Active | -0.46 | 0.236 | -0.94 | 0.03 | | | | | | |
| | | Sham | -1.12 | 0.245 | -1.62 | -0.61 | | | | | | |
| | | Active | | | | | Sham | 0.66 | 0.340 | -0.04 | 1.36 | 0.064 |
| | 84 | Active | -0.47 | 0.283 | -1.05 | 0.11 | | | | | | |
| | | Sham | -0.33 | 0.284 | -0.91 | 0.25 | | | | | | |
| | | Active | | | | | Sham | -0.14 | 0.401 | -0.97 | 0.68 | 0.721 |
| 2-I get tired very quickly | 0 | Active | 4.13 | 0.307 | 3.83 | 4.44 | | | | | | |
| | | Sham | 3.93 | 0.267 | 3.66 | 4.20 | | | | | | |
| | | Active | | | | | Sham | 0.20 | 0.409 | -0.63 | 1.04 | 0.620 |
| | 56 | Active | -0.18 | 0.269 | -0.73 | 0.38 | | | | | | |
| | | Sham | -0.84 | 0.280 | -1.41 | -0.26 | | | | | | |
| | | Active | | | | | Sham | 0.66 | 0.389 | -0.14 | 1.46 | 0.100 |
| | 84 | Active | -0.34 | 0.288 | -0.94 | 0.25 | | | | | | |
| | | Sham | -0.48 | 0.293 | -1.09 | 0.12 | | | | | | |
| | | Active | | | | | Sham | 0.14 | 0.412 | -0.71 | 0.99 | 0.739 |
| 3-I don't do much during the day | 0 | Active | 3.40 | 0.423 | 2.98 | 3.82 | | | | | | |
| | | Sham | 2.93 | 0.305 | 2.62 | 3.23 | | | | | | |
| | | Active | | | | | Sham | 0.47 | 0.528 | -0.61 | 1.56 | 0.380 |
| | 56 | Active | -0.16 | 0.229 | -0.63 | 0.32 | | | | | | |
| | | Sham | -0.50 | 0.238 | -0.98 | -0.01 | | | | | | |
| | | Active | | | | | Sham | 0.34 | 0.333 | -0.34 | 1.02 | 0.316 |
| | 84 | Active | -0.32 | 0.221 | -0.77 | 0.14 | | | | | | |
| | | Sham | -0.64 | 0.223 | -1.10 | -0.18 | | | | | | |
| | | Active | | | | | Sham | 0.32 | 0.317 | -0.33 | 0.97 | 0.321 |

*p<0.05 by analysis of covariance of the change from baseline to day 56 and day 84 in FAS parameters, with device, visit, treatment by visit interaction, and baseline covariate. Confidence interval (CI), Least-squares (LS) mean.
For Fatigue Assessment Scale (FAS) questions 1-3 and 5-9, a negative change is in the direction of improvement, and for questions 4 and 10, a positive change is in the direction of improvement. For the sum, a negative change is in the direction of improvement, with reverse score for questions 4 and 10 as (5 minus actual score). Source: Listing 10.

31DEC24, T03_fas_lt45

Table 3.1

Analysis of Covariance
FAS Treatment Device Comparison in Change from Baseline
Age < 45 Years (Per-Protocol Population)

| Parameter | Day | Treatment Device | LS Mean | Std Error | 95% CI Lower | 95% CI Upper | Treatment -Device | Difference LS Mean | Difference Std Error | 95% CI Lower | 95% CI Upper | P-Value |
|---|---|---|---|---|---|---|---|---|---|---|---|---|
| 4-I have enough energy for everyday life | 0 | Active | 3.20 | 0.368 | 2.83 | 3.57 | | | | | | |
| | | Sham | 3.43 | 0.173 | 3.26 | 3.60 | | | | | | |
| | | Active | | | | | Sham | -0.23 | 0.416 | -1.08 | 0.62 | 0.587 |
| | 56 | Active | -0.61 | 0.253 | -1.13 | -0.09 | | | | | | |
| | | Sham | -0.63 | 0.262 | -1.17 | -0.09 | | | | | | |
| | | Active | | | | | Sham | 0.02 | 0.364 | -0.73 | 0.77 | 0.964 |
| | 84 | Active | -0.28 | 0.184 | -0.66 | 0.10 | | | | | | |
| | | Sham | -0.49 | 0.185 | -0.87 | -0.11 | | | | | | |
| | | Active | | | | | Sham | 0.21 | 0.262 | -0.33 | 0.74 | 0.436 |
| 5-Physically, I feel exhausted | 0 | Active | 4.00 | 0.352 | 3.65 | 4.35 | | | | | | |
| | | Sham | 3.71 | 0.286 | 3.43 | 4.00 | | | | | | |
| | | Active | | | | | Sham | 0.29 | 0.457 | -0.65 | 1.22 | 0.537 |
| | 56 | Active | -0.35 | 0.284 | -0.94 | 0.23 | | | | | | |
| | | Sham | -0.72 | 0.295 | -1.33 | -0.12 | | | | | | |
| | | Active | | | | | Sham | 0.37 | 0.411 | -0.47 | 1.21 | 0.376 |
| | 84 | Active | -0.27 | 0.227 | -0.74 | 0.20 | | | | | | |
| | | Sham | -0.51 | 0.229 | -0.98 | -0.04 | | | | | | |
| | | Active | | | | | Sham | 0.24 | 0.325 | -0.43 | 0.91 | 0.469 |
| 6-I have problems to start things | 0 | Active | 3.00 | 0.309 | 2.69 | 3.31 | | | | | | |
| | | Sham | 2.93 | 0.286 | 2.64 | 3.21 | | | | | | |
| | | Active | | | | | Sham | 0.07 | 0.423 | -0.80 | 0.94 | 0.867 |
| | 56 | Active | -0.05 | 0.217 | -0.50 | 0.39 | | | | | | |
| | | Sham | -0.16 | 0.224 | -0.62 | 0.30 | | | | | | |
| | | Active | | | | | Sham | 0.10 | 0.312 | -0.54 | 0.74 | 0.744 |
| | 84 | Active | -0.13 | 0.284 | -0.71 | 0.46 | | | | | | |
| | | Sham | -0.16 | 0.284 | -0.74 | 0.43 | | | | | | |
| | | Active | | | | | Sham | 0.03 | 0.402 | -0.80 | 0.86 | 0.943 |

*p<0.05 by analysis of covariance of the change from baseline to day 56 and day 84 in FAS parameters, with device, visit, treatment by visit interaction, and baseline covariate. Confidence interval (CI), Least-squares (LS) mean.
For Fatigue Assessment Scale (FAS) questions 1-3 and 5-9, a negative change is in the direction of improvement, and for questions 4 and 10, a positive change is in the direction of improvement. For the sum, a negative change is in the direction of improvement, with reverse score for questions 4 and 10 as (5 minus actual score). Source: Listing 10.

31DEC24, T03_fas_lt45

Table 3.1

Analysis of Covariance
FAS Treatment Device Comparison in Change from Baseline
Age < 45 Years (Per-Protocol Population)

| | | Treatment Device Estimate | | | | | Treatment Device Comparison — Difference | | | | | |
|---|---|---|---|---|---|---|---|---|---|---|---|---|
| Parameter | Day | Treatment Device | LS Mean | Std Error | 95% CI Lower | 95% CI Upper | Treatment -Device | LS Mean | Std Error | 95% CI Lower | 95% CI Upper | P-Value |
| 7-I have problems to think clearly | 0 | Active | 4.00 | 0.276 | 3.72 | 4.28 | | | | | | |
| | | Sham | 3.71 | 0.244 | 3.47 | 3.96 | | | | | | |
| | | Active | | | | | Sham | 0.29 | 0.371 | -0.47 | 1.05 | 0.447 |
| | 56 | Active | -0.67 | 0.232 | -1.15 | -0.19 | | | | | | |
| | | Sham | -0.94 | 0.241 | -1.44 | -0.45 | | | | | | |
| | | Active | | | | | Sham | 0.27 | 0.336 | -0.42 | 0.96 | 0.428 |
| | 84 | Active | -0.89 | 0.255 | -1.41 | -0.37 | | | | | | |
| | | Sham | -0.58 | 0.260 | -1.12 | -0.05 | | | | | | |
| | | Active | | | | | Sham | -0.30 | 0.365 | -1.05 | 0.45 | 0.414 |
| 8-I feel no desire to do anything | 0 | Active | 1.73 | 0.206 | 1.53 | 1.94 | | | | | | |
| | | Sham | 2.43 | 0.309 | 2.12 | 2.74 | | | | | | |
| | | Active | | | | | Sham | -0.70 | 0.367 | -1.45 | 0.06 | 0.069 |
| | 56 | Active | -0.21 | 0.155 | -0.53 | 0.10 | | | | | | |
| | | Sham | -0.20 | 0.160 | -0.53 | 0.13 | | | | | | |
| | | Active | | | | | Sham | -0.01 | 0.229 | -0.48 | 0.46 | 0.952 |
| | 84 | Active | -0.08 | 0.184 | -0.45 | 0.30 | | | | | | |
| | | Sham | -0.20 | 0.186 | -0.58 | 0.18 | | | | | | |
| | | Active | | | | | Sham | 0.12 | 0.267 | -0.42 | 0.67 | 0.647 |
| 9-Mentally, I feel exhausted | 0 | Active | 3.80 | 0.296 | 3.50 | 4.10 | | | | | | |
| | | Sham | 3.79 | 0.214 | 3.57 | 4.00 | | | | | | |
| | | Active | | | | | Sham | 0.01 | 0.370 | -0.74 | 0.77 | 0.969 |
| | 56 | Active | -0.60 | 0.243 | -1.10 | -0.10 | | | | | | |
| | | Sham | -0.79 | 0.252 | -1.31 | -0.27 | | | | | | |
| | | Active | | | | | Sham | 0.19 | 0.350 | -0.53 | 0.91 | 0.592 |
| | 84 | Active | -0.54 | 0.250 | -1.05 | -0.03 | | | | | | |
| | | Sham | -0.65 | 0.254 | -1.17 | -0.13 | | | | | | |
| | | Active | | | | | Sham | 0.11 | 0.356 | -0.62 | 0.84 | 0.762 |

*p<0.05 by analysis of covariance of the change from baseline to day 56 and day 84 in FAS parameters, with device, visit, treatment by visit interaction, and baseline covariate. Confidence interval (CI), Least-squares (LS) mean.
For Fatigue Assessment Scale (FAS) questions 1-3 and 5-9, a negative change is in the direction of improvement, and for questions 4 and 10, a positive change is in the direction of improvement. For the sum, a negative change is in the direction of improvement, with reverse score for questions 4 and 10 as (5 minus actual score). Source: Listing 10.

31DEC24, T03_fas_lt45

Table 3.1

Analysis of Covariance
FAS Treatment Device Comparison in Change from Baseline
Age < 45 Years (Per-Protocol Population)

| | | Treatment Device Estimate | | | | | Treatment Device Comparison: Difference | | | | | |
|---|---|---|---|---|---|---|---|---|---|---|---|---|
| | | | | | 95% CI | | | | | 95% CI | | |
| Parameter | Day | Treatment Device | LS Mean | Std Error | Lower | Upper | Treatment -Device | LS Mean | Std Error | Lower | Upper | P-Value |
| 10-When I am doing something, I can concentrate quite well | 0 | Active | 2.67 | 0.232 | 2.43 | 2.90 | | | | | | |
| | | Sham | 2.79 | 0.261 | 2.53 | 3.05 | | | | | | |
| | | Active | | | | | Sham | -0.12 | 0.348 | -0.83 | 0.60 | 0.735 |
| | 56 | Active | -0.50 | 0.251 | -1.02 | 0.01 | | | | | | |
| | | Sham | -0.17 | 0.260 | -0.70 | 0.36 | | | | | | |
| | | Active | | | | | Sham | -0.33 | 0.361 | -1.08 | 0.41 | 0.365 |
| | 84 | Active | -0.52 | 0.261 | -1.05 | 0.02 | | | | | | |
| | | Sham | 0.19 | 0.265 | -0.36 | 0.73 | | | | | | |
| | | Active | | | | | Sham | -0.70 | 0.373 | -1.47 | 0.06 | 0.070 |
| Sum FAS Questions 1-10 | 0 | Active | 34.20 | 2.391 | 31.81 | 36.59 | | | | | | |
| | | Sham | 33.79 | 1.446 | 32.34 | 35.23 | | | | | | |
| | | Active | | | | | Sham | 0.41 | 2.843 | -5.42 | 6.25 | 0.885 |
| | 56 | Active | -3.86 | 1.518 | -6.98 | -0.74 | | | | | | |
| | | Sham | -5.93 | 1.572 | -9.16 | -2.70 | | | | | | |
| | | Active | | | | | Sham | 2.06 | 2.185 | -2.43 | 6.56 | 0.353 |
| | 84 | Active | -3.84 | 1.605 | -7.14 | -0.54 | | | | | | |
| | | Sham | -3.71 | 1.636 | -7.08 | -0.34 | | | | | | |
| | | Active | | | | | Sham | -0.13 | 2.293 | -4.84 | 4.59 | 0.957 |

*p<0.05 by analysis of covariance of the change from baseline to day 56 and day 84 in FAS parameters, with device, visit, treatment by visit interaction, and baseline covariate. Confidence interval (CI), Least-squares (LS) mean.
For Fatigue Assessment Scale (FAS) questions 1-3 and 5-9, a negative change is in the direction of improvement, and for questions 4 and 10, a positive change is in the direction of improvement. For the sum, a negative change is in the direction of improvement, with reverse score for questions 4 and 10 as (5 minus actual score). Source: Listing 10.

31DEC24, T03_fas_lt45

Table 3.2

Analysis of Covariance
FAS Treatment Device Comparison in Change from Baseline
Age >= 45 Years (Per-Protocol Population)

| Parameter | Visit | Treatment Device | LS Mean | Std Error | 95% CI Lower | 95% CI Upper | Treatment -Device | Difference LS Mean | Difference Std Error | Difference 95% CI Lower | Difference 95% CI Upper | P-Value |
|---|---|---|---|---|---|---|---|---|---|---|---|---|
| 1-I am bothered by fatigue | BASELINE | Active | 4.00 | 0.632 | 3.37 | 4.63 | | | | | | |
| | | Sham | 3.67 | 0.422 | 3.25 | 4.09 | | | | | | |
| | | Active | | | | | Sham | 0.33 | 0.760 | -1.36 | 2.03 | 0.670 |
| | DAY 56 | Active | 0.00 | 0.248 | -0.56 | 0.56 | | | | | | |
| | | Sham | -0.33 | 0.248 | -0.89 | 0.22 | | | | | | |
| | | Active | | | | | Sham | 0.34 | 0.352 | -0.46 | 1.13 | 0.365 |
| | DAY 84 | Active | -0.33 | 0.295 | -0.99 | 0.33 | | | | | | |
| | | Sham | -0.50 | 0.295 | -1.16 | 0.16 | | | | | | |
| | | Active | | | | | Sham | 0.17 | 0.418 | -0.77 | 1.11 | 0.695 |
| 2-I get tired very quickly | BASELINE | Active | 3.67 | 0.667 | 3.00 | 4.33 | | | | | | |
| | | Sham | 3.17 | 0.543 | 2.62 | 3.71 | | | | | | |
| | | Active | | | | | Sham | 0.50 | 0.860 | -1.42 | 2.42 | 0.574 |
| | DAY 56 | Active | 0.39 | 0.306 | -0.30 | 1.09 | | | | | | |
| | | Sham | -0.23 | 0.306 | -0.92 | 0.47 | | | | | | |
| | | Active | | | | | Sham | 0.62 | 0.438 | -0.37 | 1.61 | 0.191 |
| | DAY 84 | Active | 0.06 | 0.365 | -0.77 | 0.88 | | | | | | |
| | | Sham | -0.39 | 0.365 | -1.22 | 0.43 | | | | | | |
| | | Active | | | | | Sham | 0.45 | 0.520 | -0.72 | 1.63 | 0.406 |
| 3-I don't do much during the day | BASELINE | Active | 3.33 | 0.558 | 2.78 | 3.89 | | | | | | |
| | | Sham | 1.83 | 0.401 | 1.43 | 2.23 | | | | | | |
| | | Active | | | | | Sham | 1.50 | 0.687 | -0.03 | 3.03 | 0.054 |
| | DAY 56 | Active | 0.00 | 0.309 | -0.70 | 0.70 | | | | | | |
| | | Sham | -0.00 | 0.309 | -0.70 | 0.70 | | | | | | |
| | | Active | | | | | Sham | 0.00 | 0.483 | -1.09 | 1.09 | 1.000 |
| | DAY 84 | Active | -0.17 | 0.330 | -0.91 | 0.57 | | | | | | |
| | | Sham | -0.00 | 0.330 | -0.74 | 0.74 | | | | | | |
| | | Active | | | | | Sham | -0.17 | 0.511 | -1.31 | 0.97 | 0.751 |

*p<0.05 by analysis of covariance of the change from baseline to day 56 and day 84 in FAS parameters, with device, visit, treatment by visit interaction, and baseline covariate. Confidence interval (CI), Least-squares (LS) mean.
For Fatigue Assessment Scale (FAS) questions 1-3 and 5-9, a negative change is in the direction of improvement, and for questions 4 and 10, a positive change is in the direction of improvement. For the sum, a negative change is in the direction of improvement, with reverse score for questions 4 and 10 as (5 minus actual score). Source: Listing 10.

31DEC24, T03_fas_ge45

Table 3.2

Analysis of Covariance
FAS Treatment Device Comparison in Change from Baseline
Age >= 45 Years (Per-Protocol Population)

| | | Treatment Device Estimate | | | | | Treatment Device Comparison: Difference | | | | | |
|---|---|---|---|---|---|---|---|---|---|---|---|---|
| Parameter | Visit | Treatment Device | LS Mean | Std Error | 95% CI Lower | 95% CI Upper | Treatment -Device | LS Mean | Std Error | 95% CI Lower | 95% CI Upper | P-Value |
| 4-I have enough energy for everyday life | BASELINE | Active | 3.00 | 0.632 | 2.37 | 3.63 | | | | | | |
| | | Sham | 3.17 | 0.307 | 2.86 | 3.47 | | | | | | |
| | | Active | | | | | Sham | -0.17 | 0.703 | -1.73 | 1.40 | 0.817 |
| | DAY 56 | Active | 0.14 | 0.351 | -0.65 | 0.94 | | | | | | |
| | | Sham | -1.14 | 0.351 | -1.94 | -0.35 | | | | | | |
| | | Active | | | | | Sham | 1.29 | 0.496 | 0.16 | 2.41 | 0.029* |
| | DAY 84 | Active | -0.52 | 0.467 | -1.57 | 0.53 | | | | | | |
| | | Sham | -0.98 | 0.467 | -2.03 | 0.07 | | | | | | |
| | | Active | | | | | Sham | 0.45 | 0.661 | -1.03 | 1.94 | 0.510 |
| 5-Physically, I feel exhausted | BASELINE | Active | 3.83 | 0.654 | 3.18 | 4.49 | | | | | | |
| | | Sham | 3.00 | 0.516 | 2.48 | 3.52 | | | | | | |
| | | Active | | | | | Sham | 0.83 | 0.833 | -1.02 | 2.69 | 0.341 |
| | DAY 56 | Active | 0.02 | 0.323 | -0.71 | 0.75 | | | | | | |
| | | Sham | -0.35 | 0.323 | -1.08 | 0.38 | | | | | | |
| | | Active | | | | | Sham | 0.37 | 0.469 | -0.70 | 1.43 | 0.456 |
| | DAY 84 | Active | -0.15 | 0.413 | -1.09 | 0.79 | | | | | | |
| | | Sham | -0.18 | 0.413 | -1.12 | 0.76 | | | | | | |
| | | Active | | | | | Sham | 0.03 | 0.595 | -1.32 | 1.38 | 0.958 |
| 6-I have problems to start things | BASELINE | Active | 3.50 | 0.500 | 3.00 | 4.00 | | | | | | |
| | | Sham | 2.50 | 0.500 | 2.00 | 3.00 | | | | | | |
| | | Active | | | | | Sham | 1.00 | 0.707 | -0.58 | 2.58 | 0.188 |
| | DAY 56 | Active | -0.53 | 0.408 | -1.45 | 0.39 | | | | | | |
| | | Sham | -0.14 | 0.408 | -1.06 | 0.78 | | | | | | |
| | | Active | | | | | Sham | -0.39 | 0.599 | -1.74 | 0.96 | 0.531 |
| | DAY 84 | Active | -0.53 | 0.341 | -1.30 | 0.24 | | | | | | |
| | | Sham | 0.20 | 0.341 | -0.58 | 0.97 | | | | | | |
| | | Active | | | | | Sham | -0.72 | 0.509 | -1.87 | 0.43 | 0.189 |

*p<0.05 by analysis of covariance of the change from baseline to day 56 and day 84 in FAS parameters, with device, visit, treatment by visit interaction, and baseline covariate. Confidence interval (CI), Least-squares (LS) mean.
For Fatigue Assessment Scale (FAS) questions 1-3 and 5-9, a negative change is in the direction of improvement, and for questions 4 and 10, a positive change is in the direction of improvement. For the sum, a negative change is in the direction of improvement, with reverse score for questions 4 and 10 as (5 minus actual score). Source: Listing 10.

31DEC24, T03_fas_ge45

Table 3.2

Analysis of Covariance
FAS Treatment Device Comparison in Change from Baseline
Age >= 45 Years (Per-Protocol Population)

| | | Treatment Device Estimate | | | | | Treatment Device Comparison: Difference | | | | | |
|---|---|---|---|---|---|---|---|---|---|---|---|---|
| Parameter | Visit | Treatment Device | LS Mean | Std Error | 95% CI Lower | 95% CI Upper | Treatment -Device | LS Mean | Std Error | 95% CI Lower | 95% CI Upper | P-Value |
| 7-I have problems to think clearly | BASELINE | Active | 3.50 | 0.500 | 3.00 | 4.00 | | | | | | |
| | | Sham | 3.67 | 0.211 | 3.46 | 3.88 | | | | | | |
| | | Active | | | | | Sham | -0.17 | 0.543 | -1.38 | 1.04 | 0.765 |
| | DAY 56 | Active | 0.01 | 0.257 | -0.57 | 0.59 | | | | | | |
| | | Sham | -1.01 | 0.257 | -1.59 | -0.43 | | | | | | |
| | | Active | | | | | Sham | 1.02 | 0.364 | 0.20 | 1.84 | 0.020* |
| | DAY 84 | Active | -0.16 | 0.373 | -0.99 | 0.68 | | | | | | |
| | | Sham | -0.84 | 0.373 | -1.68 | -0.01 | | | | | | |
| | | Active | | | | | Sham | 0.69 | 0.528 | -0.50 | 1.87 | 0.225 |
| 8-I feel no desire to do anything | BASELINE | Active | 2.83 | 0.477 | 2.36 | 3.31 | | | | | | |
| | | Sham | 2.00 | 0.258 | 1.74 | 2.26 | | | | | | |
| | | Active | | | | | Sham | 0.83 | 0.543 | -0.38 | 2.04 | 0.156 |
| | DAY 56 | Active | 0.19 | 0.363 | -0.63 | 1.01 | | | | | | |
| | | Sham | -0.53 | 0.363 | -1.35 | 0.29 | | | | | | |
| | | Active | | | | | Sham | 0.72 | 0.544 | -0.51 | 1.95 | 0.219 |
| | DAY 84 | Active | -0.14 | 0.446 | -1.14 | 0.86 | | | | | | |
| | | Sham | 0.31 | 0.446 | -0.69 | 1.30 | | | | | | |
| | | Active | | | | | Sham | -0.45 | 0.656 | -1.91 | 1.01 | 0.509 |
| 9-Mentally, I feel exhausted | BASELINE | Active | 3.50 | 0.719 | 2.78 | 4.22 | | | | | | |
| | | Sham | 3.17 | 0.477 | 2.69 | 3.64 | | | | | | |
| | | Active | | | | | Sham | 0.33 | 0.863 | -1.59 | 2.26 | 0.707 |
| | DAY 56 | Active | 0.24 | 0.304 | -0.45 | 0.93 | | | | | | |
| | | Sham | -0.74 | 0.304 | -1.43 | -0.05 | | | | | | |
| | | Active | | | | | Sham | 0.98 | 0.432 | -0.00 | 1.96 | 0.051 |
| | DAY 84 | Active | 0.24 | 0.362 | -0.59 | 1.06 | | | | | | |
| | | Sham | -0.57 | 0.362 | -1.40 | 0.25 | | | | | | |
| | | Active | | | | | Sham | 0.81 | 0.513 | -0.36 | 1.98 | 0.151 |

*p<0.05 by analysis of covariance of the change from baseline to day 56 and day 84 in FAS parameters, with device, visit, treatment by visit interaction, and baseline covariate. Confidence interval (CI), Least-squares (LS) mean.
For Fatigue Assessment Scale (FAS) questions 1-3 and 5-9, a negative change is in the direction of improvement, and for questions 4 and 10, a positive change is in the direction of improvement. For the sum, a negative change is in the direction of improvement, with reverse score for questions 4 and 10 as (5 minus actual score). Source: Listing 10.

31DEC24, T03_fas_ge45

Table 3.2

Analysis of Covariance
FAS Treatment Device Comparison in Change from Baseline
Age >= 45 Years (Per-Protocol Population)

| Parameter | Visit | Treatment Device Estimate: Treatment Device | LS Mean | Std Error | 95% CI Lower | 95% CI Upper | Treatment Device Comparison: Treatment -Device | Difference: LS Mean | Std Error | 95% CI Lower | 95% CI Upper | P-Value |
|---|---|---|---|---|---|---|---|---|---|---|---|---|
| 10-When I am doing something, I can concentrate quite well | BASELINE | Active | 3.17 | 0.167 | 3.00 | 3.33 | | | | | | |
| | | Sham | 3.00 | 0.000 | 3.00 | 3.00 | | | | | | |
| | | Active | | | | | Sham | 0.17 | 0.167 | -0.20 | 0.54 | 0.341 |
| | DAY 56 | Active | -0.44 | 0.402 | -1.34 | 0.46 | | | | | | |
| | | Sham | -0.73 | 0.402 | -1.63 | 0.17 | | | | | | |
| | | Active | | | | | Sham | 0.29 | 0.581 | -1.01 | 1.59 | 0.629 |
| | DAY 84 | Active | -0.27 | 0.364 | -1.09 | 0.55 | | | | | | |
| | | Sham | -1.06 | 0.364 | -1.88 | -0.24 | | | | | | |
| | | Active | | | | | Sham | 0.79 | 0.529 | -0.40 | 1.98 | 0.169 |
| Sum FAS Questions 1-10 | BASELINE | Active | 34.33 | 4.828 | 29.51 | 39.16 | | | | | | |
| | | Sham | 29.17 | 3.092 | 26.07 | 32.26 | | | | | | |
| | | Active | | | | | Sham | 5.17 | 5.733 | -7.61 | 17.94 | 0.389 |
| | DAY 56 | Active | -0.55 | 1.928 | -4.89 | 3.78 | | | | | | |
| | | Sham | -4.61 | 1.928 | -8.95 | -0.28 | | | | | | |
| | | Active | | | | | Sham | 4.06 | 2.761 | -2.13 | 10.25 | 0.174 |
| | DAY 84 | Active | -2.55 | 1.404 | -5.73 | 0.62 | | | | | | |
| | | Sham | -3.45 | 1.404 | -6.62 | -0.27 | | | | | | |
| | | Active | | | | | Sham | 0.89 | 2.032 | -3.71 | 5.49 | 0.671 |

*p<0.05 by analysis of covariance of the change from baseline to day 56 and day 84 in FAS parameters, with device, visit, treatment by visit interaction, and baseline covariate. Confidence interval (CI), Least-squares (LS) mean.
For Fatigue Assessment Scale (FAS) questions 1-3 and 5-9, a negative change is in the direction of improvement, and for questions 4 and 10, a positive change is in the direction of improvement. For the sum, a negative change is in the direction of improvement, with reverse score for questions 4 and 10 as (5 minus actual score). Source: Listing 10.

31DEC24, T03_fas_ge45

Table 4

Primary Efficacy Analysis of Covariance
Creyos Treatment Device Comparison in Change from Baseline by Parameter (Per-Protocol Population)

| | | | Treatment Device Estimate | | | | Treatment Device Comparison | | | | | |
|---|---|---|---|---|---|---|---|---|---|---|---|---|
| | | | | Std | 95% CI | | | Difference | | 95% CI | | |
| Parameter | Visit | Treatment Device | LS Mean | Error | Lower | Upper | Comparator | LS Mean | Std Error | Lower | Upper | P-Value |
| PRIMARY ENDPOINT | BASELINE | Active | 0.722 | 0.020 | 0.702 | 0.743 | | | | | | |
| | | Sham | 0.716 | 0.018 | 0.698 | 0.735 | | | | | | |
| | | Active | | | | | Sham | 0.006 | 0.043 | -0.081 | 0.093 | 0.887 |
| | DAY 14 | Active | 0.002 | 0.024 | -0.046 | 0.050 | | | | | | |
| | | Sham | -0.040 | 0.024 | -0.089 | 0.009 | | | | | | |
| | | Active | | | | | Sham | 0.043 | 0.033 | -0.024 | 0.109 | 0.201 |
| | DAY 28 | Active | 0.030 | 0.022 | -0.015 | 0.074 | | | | | | |
| | | Sham | -0.005 | 0.023 | -0.052 | 0.041 | | | | | | |
| | | Active | | | | | Sham | 0.035 | 0.031 | -0.028 | 0.097 | 0.267 |
| | DAY 56 | Active | 0.050 | 0.018 | 0.013 | 0.086 | | | | | | |
| | | Sham | 0.007 | 0.018 | -0.030 | 0.044 | | | | | | |
| | | Active | | | | | Sham | 0.043 | 0.024 | -0.007 | 0.092 | 0.088 |
| | DAY 84 | Active | 0.051 | 0.025 | 0.001 | 0.100 | | | | | | |
| | | Sham | 0.012 | 0.026 | -0.040 | 0.064 | | | | | | |
| | | Active | | | | | Sham | 0.038 | 0.035 | -0.032 | 0.108 | 0.277 |

The primary efficacy endpoint is the mean of 7 test items including spatial planning, monkey ladder, rotations, feature match, paired associates, token search and polygons. Confidence interval (CI), Least-squares (LS) mean. Source: Listing 12.
*p<0.05 by analysis of covariance of the change from baseline to days 14, 28, 56 and 84 (no device) in Creyos parameters, with device, visit, treatment device by visit interaction, age >=45 years (yes, no) and baseline covariate.
A greater percentile is in the direction of improved function. 31DEC24, T04_creyos_prim

Table 4.1

Primary Efficacy Analysis of Covariance
Creyos Treatment Device Comparison in Change from Baseline by Parameter
Age < 45 Years (Per-Protocol Population)

| | | Treatment Device Estimate | | | | | Treatment Device Comparison | | | | | |
|---|---|---|---|---|---|---|---|---|---|---|---|---|
| | | | | | 95% CI | | | Difference | | 95% CI | | |
| Parameter | Visit | Treatment Device | LS Mean | Std Error | Lower | Upper | Comparator | LS Mean | Std Error | Lower | Upper | P-Value |
| PRIMARY ENDPOINT | BASELINE | Active | 0.755 | 0.023 | 0.732 | 0.779 | | | | | | |
| | | Sham | 0.737 | 0.021 | 0.715 | 0.758 | | | | | | |
| | | Active | | | | | Sham | 0.019 | 0.046 | -0.075 | 0.113 | 0.687 |
| | DAY 14 | Active | 0.035 | 0.024 | -0.014 | 0.084 | | | | | | |
| | | Sham | -0.036 | 0.025 | -0.086 | 0.015 | | | | | | |
| | | Active | | | | | Sham | 0.070 | 0.034 | -0.001 | 0.141 | 0.052 |
| | DAY 28 | Active | 0.051 | 0.025 | -0.001 | 0.104 | | | | | | |
| | | Sham | 0.011 | 0.027 | -0.045 | 0.067 | | | | | | |
| | | Active | | | | | Sham | 0.040 | 0.037 | -0.037 | 0.117 | 0.294 |
| | DAY 56 | Active | 0.082 | 0.018 | 0.046 | 0.118 | | | | | | |
| | | Sham | 0.023 | 0.018 | -0.014 | 0.060 | | | | | | |
| | | Active | | | | | Sham | 0.059 | 0.025 | 0.007 | 0.111 | 0.028* |
| | DAY 84 | Active | 0.071 | 0.024 | 0.021 | 0.120 | | | | | | |
| | | Sham | 0.034 | 0.026 | -0.019 | 0.087 | | | | | | |
| | | Active | | | | | Sham | 0.036 | 0.035 | -0.037 | 0.109 | 0.314 |

The primary efficacy endpoint is the mean of 7 test items including spatial planning, monkey ladder, rotations, feature match, paired associates, token search and polygons. Confidence interval (CI), Least-squares (LS) mean. Source: Listing 12.
*p<0.05 by analysis of covariance of the change from baseline to days 14, 28, 56 and 84 (no device) in Creyos parameters, with device, visit, treatment device by visit interaction, and baseline covariate.
A greater percentile is in the direction of improved function. 31DEC24, T04_creyos_prim_lt45

Table 4.2

Primary Efficacy Analysis of Covariance
Creyos Treatment Device Comparison in Change from Baseline by Parameter
Age >= 45 Years (Per-Protocol Population)

| | | | Treatment Device Estimate | | | | Treatment Device Comparison | | | | | |
|---|---|---|---|---|---|---|---|---|---|---|---|---|
| | | | | Std | 95% CI | | | Difference | | 95% CI | | |
| Parameter | Visit | Treatment Device | LS Mean | Error | Lower | Upper | Comparator | LS Mean | Std Error | Lower | Upper | P-Value |
| PRIMARY ENDPOINT | BASELINE | Active | 0.640 | 0.040 | 0.601 | 0.680 | | | | | | |
| | | Sham | 0.669 | 0.035 | 0.633 | 0.704 | | | | | | |
| | | Active | | | | | Sham | -0.029 | 0.092 | -0.233 | 0.176 | 0.762 |
| | DAY 14 | Active | -0.023 | 0.053 | -0.142 | 0.097 | | | | | | |
| | | Sham | -0.005 | 0.053 | -0.125 | 0.114 | | | | | | |
| | | Active | | | | | Sham | -0.017 | 0.075 | -0.186 | 0.152 | 0.825 |
| | DAY 28 | Active | 0.032 | 0.044 | -0.067 | 0.130 | | | | | | |
| | | Sham | 0.003 | 0.043 | -0.096 | 0.101 | | | | | | |
| | | Active | | | | | Sham | 0.029 | 0.062 | -0.111 | 0.169 | 0.652 |
| | DAY 56 | Active | 0.026 | 0.042 | -0.069 | 0.120 | | | | | | |
| | | Sham | 0.016 | 0.042 | -0.078 | 0.110 | | | | | | |
| | | Active | | | | | Sham | 0.010 | 0.060 | -0.124 | 0.144 | 0.873 |
| | DAY 84 | Active | 0.056 | 0.062 | -0.082 | 0.195 | | | | | | |
| | | Sham | -0.011 | 0.064 | -0.153 | 0.131 | | | | | | |
| | | Active | | | | | Sham | 0.067 | 0.089 | -0.131 | 0.266 | 0.465 |

The primary efficacy endpoint is the mean of 7 test items including spatial planning, monkey ladder, rotations, feature match, paired associates, token search and polygons. Confidence interval (CI), Least-squares (LS) mean. Source: Listing 12.
*p<0.05 by analysis of covariance of the change from baseline to days 14, 28, 56 and 84 (no device) in Creyos parameters, with device, visit, treatment device by visit interaction, and baseline covariate.
A greater percentile is in the direction of improved function.

31DEC24, T04_creyos_prim_ge45

Table 5

Analysis of Covariance
Creyos Treatment Device Comparison of Change from Baseline by Parameter (Per-Protocol Population)

| | | Treatment Device Estimate | | | | | Treatment Device Comparison | | | | | |
|---|---|---|---|---|---|---|---|---|---|---|---|---|
| | | Treatment | | Std | 95% CI | | | Difference | | 95% CI | | |
| Parameter | Visit | Device | LS Mean | Error | Lower | Upper | Comparator | LS Mean | Std Error | Lower | Upper | P-Value |
| Spatial Planning | BASELINE | Active | 0.716 | 0.046 | 0.67 | 0.76 | | | | | | |
| | | Sham | 0.750 | 0.038 | 0.71 | 0.79 | | | | | | |
| | | Active | | | | | Sham | -0.034 | 0.060 | -0.16 | 0.09 | 0.570 |
| | DAY 14 | Active | 0.037 | 0.043 | -0.05 | 0.12 | | | | | | |
| | | Sham | 0.016 | 0.044 | -0.07 | 0.10 | | | | | | |
| | | Active | | | | | Sham | 0.021 | 0.060 | -0.10 | 0.14 | 0.730 |
| | DAY 28 | Active | 0.053 | 0.036 | -0.02 | 0.13 | | | | | | |
| | | Sham | -0.011 | 0.037 | -0.09 | 0.06 | | | | | | |
| | | Active | | | | | Sham | 0.064 | 0.051 | -0.04 | 0.17 | 0.214 |
| | DAY 56 | Active | 0.150 | 0.026 | 0.10 | 0.20 | | | | | | |
| | | Sham | 0.126 | 0.026 | 0.07 | 0.18 | | | | | | |
| | | Active | | | | | Sham | 0.024 | 0.035 | -0.05 | 0.09 | 0.502 |
| | DAY 84 | Active | 0.135 | 0.032 | 0.07 | 0.20 | | | | | | |
| | | Sham | 0.093 | 0.034 | 0.02 | 0.16 | | | | | | |
| | | Active | | | | | Sham | 0.043 | 0.046 | -0.05 | 0.14 | 0.359 |
| Monkey Ladder | BASELINE | Active | 0.785 | 0.041 | 0.74 | 0.83 | | | | | | |
| | | Sham | 0.729 | 0.027 | 0.70 | 0.76 | | | | | | |
| | | Active | | | | | Sham | 0.056 | 0.050 | -0.04 | 0.16 | 0.266 |
| | DAY 14 | Active | -0.028 | 0.032 | -0.09 | 0.04 | | | | | | |
| | | Sham | -0.017 | 0.033 | -0.08 | 0.05 | | | | | | |
| | | Active | | | | | Sham | -0.010 | 0.044 | -0.10 | 0.08 | 0.816 |
| | DAY 28 | Active | -0.052 | 0.034 | -0.12 | 0.02 | | | | | | |
| | | Sham | 0.006 | 0.036 | -0.07 | 0.08 | | | | | | |
| | | Active | | | | | Sham | -0.058 | 0.048 | -0.16 | 0.04 | 0.238 |
| | DAY 56 | Active | 0.013 | 0.036 | -0.06 | 0.09 | | | | | | |
| | | Sham | -0.021 | 0.038 | -0.10 | 0.06 | | | | | | |
| | | Active | | | | | Sham | 0.034 | 0.051 | -0.07 | 0.14 | 0.505 |
| | DAY 84 | Active | -0.013 | 0.042 | -0.10 | 0.07 | | | | | | |
| | | Sham | -0.027 | 0.045 | -0.12 | 0.06 | | | | | | |
| | | Active | | | | | Sham | 0.013 | 0.061 | -0.11 | 0.14 | 0.827 |

*p<0.05 by analysis of covariance of the change from baseline to days 14, 28, 56 and 84 (no device) in Creyos parameters, with device, visit, treatment by visit interaction, age >=45 years (yes, no) and baseline covariate. Confidence interval (CI), Least-squares (LS) mean. A greater percentile is in the direction of improved function. Source: Listing 12. 31DEC24, T05_creyos

Table 5

Analysis of Covariance
Creyos Treatment Device Comparison of Change from Baseline by Parameter (Per-Protocol Population)

| | | Treatment Device Estimate | | | | | Treatment Device Comparison | | | | | |
|---|---|---|---|---|---|---|---|---|---|---|---|---|
| | | | | | 95% CI | | | Difference | | 95% CI | | |
| Parameter | Visit | Treatment Device | LS Mean | Std Error | Lower | Upper | Comparator | LS Mean | Std Error | Lower | Upper | P-Value |
| Rotations | BASELINE | Active | 0.675 | 0.070 | 0.60 | 0.74 | | | | | | |
| | | Sham | 0.710 | 0.059 | 0.65 | 0.77 | | | | | | |
| | | Active | | | | | Sham | -0.035 | 0.091 | -0.22 | 0.15 | 0.701 |
| | DAY 14 | Active | 0.068 | 0.053 | -0.04 | 0.18 | | | | | | |
| | | Sham | -0.033 | 0.055 | -0.14 | 0.08 | | | | | | |
| | | Active | | | | | Sham | 0.100 | 0.075 | -0.05 | 0.25 | 0.186 |
| | DAY 28 | Active | 0.005 | 0.051 | -0.10 | 0.11 | | | | | | |
| | | Sham | -0.001 | 0.053 | -0.11 | 0.11 | | | | | | |
| | | Active | | | | | Sham | 0.006 | 0.072 | -0.14 | 0.15 | 0.933 |
| | DAY 56 | Active | 0.085 | 0.055 | -0.03 | 0.20 | | | | | | |
| | | Sham | 0.005 | 0.056 | -0.11 | 0.12 | | | | | | |
| | | Active | | | | | Sham | 0.080 | 0.077 | -0.07 | 0.24 | 0.302 |
| | DAY 84 | Active | 0.081 | 0.055 | -0.03 | 0.19 | | | | | | |
| | | Sham | -0.030 | 0.058 | -0.15 | 0.09 | | | | | | |
| | | Active | | | | | Sham | 0.111 | 0.078 | -0.05 | 0.27 | 0.164 |
| Feature Match | BASELINE | Active | 0.676 | 0.061 | 0.62 | 0.74 | | | | | | |
| | | Sham | 0.767 | 0.051 | 0.72 | 0.82 | | | | | | |
| | | Active | | | | | Sham | -0.091 | 0.080 | -0.25 | 0.07 | 0.262 |
| | DAY 14 | Active | 0.003 | 0.056 | -0.11 | 0.12 | | | | | | |
| | | Sham | -0.110 | 0.057 | -0.22 | 0.00 | | | | | | |
| | | Active | | | | | Sham | 0.112 | 0.078 | -0.05 | 0.27 | 0.157 |
| | DAY 28 | Active | 0.076 | 0.049 | -0.02 | 0.17 | | | | | | |
| | | Sham | -0.091 | 0.050 | -0.19 | 0.01 | | | | | | |
| | | Active | | | | | Sham | 0.167 | 0.068 | 0.03 | 0.31 | 0.019* |
| | DAY 56 | Active | 0.046 | 0.051 | -0.06 | 0.15 | | | | | | |
| | | Sham | -0.064 | 0.052 | -0.17 | 0.04 | | | | | | |
| | | Active | | | | | Sham | 0.109 | 0.071 | -0.03 | 0.25 | 0.133 |
| | DAY 84 | Active | 0.075 | 0.050 | -0.03 | 0.18 | | | | | | |
| | | Sham | -0.100 | 0.052 | -0.21 | 0.01 | | | | | | |
| | | Active | | | | | Sham | 0.175 | 0.070 | 0.03 | 0.32 | 0.017* |

*p<0.05 by analysis of covariance of the change from baseline to days 14, 28, 56 and 84 (no device) in Creyos parameters, with device, visit, treatment by visit interaction, age >=45 years (yes, no) and baseline covariate. Confidence interval (CI), Least-squares (LS) mean. A greater percentile is in the direction of improved function. Source: Listing 12. 31DEC24, T05_creyos

Table 5

Analysis of Covariance
Creyos Treatment Device Comparison of Change from Baseline by Parameter (Per-Protocol Population)

| | | Treatment Device Estimate | | | | | Treatment Device Comparison | | | | | |
|---|---|---|---|---|---|---|---|---|---|---|---|---|
| | | | | | 95% CI | | | Difference | | 95% CI | | |
| Parameter | Visit | Treatment Device | LS Mean | Std Error | Lower | Upper | Comparator | LS Mean | Std Error | Lower | Upper | P-Value |
| Paired Associates | BASELINE | Active | 0.715 | 0.055 | 0.66 | 0.77 | | | | | | |
| | | Sham | 0.701 | 0.051 | 0.65 | 0.75 | | | | | | |
| | | Active | | | | | Sham | 0.014 | 0.075 | -0.14 | 0.17 | 0.852 |
| | DAY 14 | Active | -0.002 | 0.059 | -0.12 | 0.12 | | | | | | |
| | | Sham | -0.052 | 0.060 | -0.17 | 0.07 | | | | | | |
| | | Active | | | | | Sham | 0.050 | 0.082 | -0.12 | 0.22 | 0.547 |
| | DAY 28 | Active | -0.007 | 0.049 | -0.11 | 0.09 | | | | | | |
| | | Sham | -0.004 | 0.051 | -0.11 | 0.10 | | | | | | |
| | | Active | | | | | Sham | -0.004 | 0.069 | -0.14 | 0.14 | 0.957 |
| | DAY 56 | Active | -0.025 | 0.043 | -0.11 | 0.06 | | | | | | |
| | | Sham | -0.009 | 0.043 | -0.10 | 0.08 | | | | | | |
| | | Active | | | | | Sham | -0.016 | 0.059 | -0.13 | 0.10 | 0.789 |
| | DAY 84 | Active | 0.021 | 0.049 | -0.08 | 0.12 | | | | | | |
| | | Sham | -0.024 | 0.053 | -0.13 | 0.08 | | | | | | |
| | | Active | | | | | Sham | 0.045 | 0.070 | -0.10 | 0.19 | 0.527 |
| Token Search | BASELINE | Active | 0.829 | 0.026 | 0.80 | 0.86 | | | | | | |
| | | Sham | 0.775 | 0.034 | 0.74 | 0.81 | | | | | | |
| | | Active | | | | | Sham | 0.055 | 0.043 | -0.03 | 0.14 | 0.207 |
| | DAY 14 | Active | -0.038 | 0.025 | -0.09 | 0.01 | | | | | | |
| | | Sham | -0.006 | 0.026 | -0.06 | 0.05 | | | | | | |
| | | Active | | | | | Sham | -0.032 | 0.035 | -0.10 | 0.04 | 0.367 |
| | DAY 28 | Active | 0.004 | 0.025 | -0.05 | 0.06 | | | | | | |
| | | Sham | -0.006 | 0.026 | -0.06 | 0.05 | | | | | | |
| | | Active | | | | | Sham | 0.010 | 0.036 | -0.06 | 0.08 | 0.784 |
| | DAY 56 | Active | -0.005 | 0.022 | -0.05 | 0.04 | | | | | | |
| | | Sham | -0.012 | 0.022 | -0.06 | 0.03 | | | | | | |
| | | Active | | | | | Sham | 0.007 | 0.030 | -0.05 | 0.07 | 0.813 |
| | DAY 84 | Active | 0.016 | 0.021 | -0.03 | 0.06 | | | | | | |
| | | Sham | 0.029 | 0.022 | -0.02 | 0.07 | | | | | | |
| | | Active | | | | | Sham | -0.013 | 0.029 | -0.07 | 0.05 | 0.668 |

*p<0.05 by analysis of covariance of the change from baseline to days 14, 28, 56 and 84 (no device) in Creyos parameters, with device, visit, treatment by visit interaction, age >=45 years (yes, no) and baseline covariate. Confidence interval (CI), Least-squares (LS) mean. A greater percentile is in the direction of improved function. Source: Listing 12. 31DEC24, T05_creyos

Table 5

Analysis of Covariance
Creyos Treatment Device Comparison of Change from Baseline by Parameter (Per-Protocol Population)

| | | Treatment Device Estimate | | | | | Treatment Device Comparison | | | | | |
|---|---|---|---|---|---|---|---|---|---|---|---|---|
| | | Treatment | | Std | 95% CI | | | Difference | | 95% CI | | |
| Parameter | Visit | Device | LS Mean | Error | Lower | Upper | Comparator | LS Mean | Std Error | Lower | Upper | P-Value |
| Polygons | BASELINE | Active | 0.660 | 0.065 | 0.60 | 0.72 | | | | | | |
| | | Sham | 0.581 | 0.064 | 0.52 | 0.65 | | | | | | |
| | | Active | | | | | Sham | 0.079 | 0.091 | -0.11 | 0.26 | 0.390 |
| | DAY 14 | Active | -0.027 | 0.059 | -0.15 | 0.09 | | | | | | |
| | | Sham | -0.104 | 0.061 | -0.23 | 0.02 | | | | | | |
| | | Active | | | | | Sham | 0.077 | 0.083 | -0.09 | 0.25 | 0.360 |
| | DAY 28 | Active | 0.125 | 0.046 | 0.03 | 0.22 | | | | | | |
| | | Sham | 0.038 | 0.048 | -0.06 | 0.13 | | | | | | |
| | | Active | | | | | Sham | 0.087 | 0.064 | -0.04 | 0.22 | 0.185 |
| | DAY 56 | Active | 0.080 | 0.053 | -0.03 | 0.19 | | | | | | |
| | | Sham | 0.000 | 0.055 | -0.11 | 0.11 | | | | | | |
| | | Active | | | | | Sham | 0.080 | 0.075 | -0.07 | 0.23 | 0.291 |
| | DAY 84 | Active | 0.035 | 0.057 | -0.08 | 0.15 | | | | | | |
| | | Sham | 0.117 | 0.061 | -0.01 | 0.24 | | | | | | |
| | | Active | | | | | Sham | -0.082 | 0.082 | -0.25 | 0.08 | 0.322 |
| Grammatical Reasoning | BASELINE | Active | 0.631 | 0.067 | 0.56 | 0.70 | | | | | | |
| | | Sham | 0.774 | 0.057 | 0.72 | 0.83 | | | | | | |
| | | Active | | | | | Sham | -0.143 | 0.088 | -0.32 | 0.04 | 0.112 |
| | DAY 14 | Active | 0.008 | 0.042 | -0.08 | 0.09 | | | | | | |
| | | Sham | 0.013 | 0.043 | -0.07 | 0.10 | | | | | | |
| | | Active | | | | | Sham | -0.005 | 0.059 | -0.12 | 0.11 | 0.933 |
| | DAY 28 | Active | 0.051 | 0.042 | -0.03 | 0.13 | | | | | | |
| | | Sham | 0.040 | 0.043 | -0.05 | 0.13 | | | | | | |
| | | Active | | | | | Sham | 0.011 | 0.059 | -0.11 | 0.13 | 0.853 |
| | DAY 56 | Active | 0.097 | 0.037 | 0.02 | 0.17 | | | | | | |
| | | Sham | 0.036 | 0.037 | -0.04 | 0.11 | | | | | | |
| | | Active | | | | | Sham | 0.061 | 0.051 | -0.04 | 0.16 | 0.247 |
| | DAY 84 | Active | 0.070 | 0.039 | -0.01 | 0.15 | | | | | | |
| | | Sham | 0.057 | 0.041 | -0.03 | 0.14 | | | | | | |
| | | Active | | | | | Sham | 0.013 | 0.055 | -0.10 | 0.13 | 0.809 |

*p<0.05 by analysis of covariance of the change from baseline to days 14, 28, 56 and 84 (no device) in Creyos parameters, with device, visit, treatment by visit interaction, age >=45 years (yes, no) and baseline covariate. Confidence interval (CI), Least-squares (LS) mean. A greater percentile is in the direction of improved function. Source: Listing 12. 31DEC24, T05_creyos

Table 5

Analysis of Covariance
Creyos Treatment Device Comparison of Change from Baseline by Parameter (Per-Protocol Population)

| | | Treatment Device Estimate | | | | | Treatment Device Comparison | | | | | |
|---|---|---|---|---|---|---|---|---|---|---|---|---|
| | | Treatment | | Std | 95% CI | | | Difference | | 95% CI | | |
| Parameter | Visit | Device | LS Mean | Error | Lower | Upper | Comparator | LS Mean | Std Error | Lower | Upper | P-Value |
| Spatial Span | BASELINE | Active | 0.789 | 0.048 | 0.74 | 0.84 | | | | | | |
| | | Sham | 0.742 | 0.059 | 0.68 | 0.80 | | | | | | |
| | | Active | | | | | Sham | 0.047 | 0.075 | -0.11 | 0.20 | 0.534 |
| | DAY 14 | Active | -0.018 | 0.050 | -0.12 | 0.08 | | | | | | |
| | | Sham | 0.051 | 0.051 | -0.05 | 0.15 | | | | | | |
| | | Active | | | | | Sham | -0.069 | 0.070 | -0.21 | 0.07 | 0.330 |
| | DAY 28 | Active | 0.018 | 0.044 | -0.07 | 0.11 | | | | | | |
| | | Sham | -0.044 | 0.047 | -0.14 | 0.05 | | | | | | |
| | | Active | | | | | Sham | 0.062 | 0.063 | -0.06 | 0.19 | 0.327 |
| | DAY 56 | Active | -0.015 | 0.044 | -0.10 | 0.07 | | | | | | |
| | | Sham | -0.008 | 0.045 | -0.10 | 0.08 | | | | | | |
| | | Active | | | | | Sham | -0.007 | 0.062 | -0.13 | 0.12 | 0.912 |
| | DAY 84 | Active | 0.046 | 0.034 | -0.02 | 0.12 | | | | | | |
| | | Sham | 0.027 | 0.037 | -0.05 | 0.10 | | | | | | |
| | | Active | | | | | Sham | 0.019 | 0.049 | -0.08 | 0.12 | 0.700 |
| Digit Span | BASELINE | Active | 0.563 | 0.046 | 0.52 | 0.61 | | | | | | |
| | | Sham | 0.511 | 0.055 | 0.46 | 0.57 | | | | | | |
| | | Active | | | | | Sham | 0.052 | 0.072 | -0.09 | 0.20 | 0.472 |
| | DAY 14 | Active | 0.097 | 0.051 | -0.01 | 0.20 | | | | | | |
| | | Sham | 0.062 | 0.052 | -0.04 | 0.17 | | | | | | |
| | | Active | | | | | Sham | 0.035 | 0.071 | -0.11 | 0.18 | 0.628 |
| | DAY 28 | Active | 0.152 | 0.048 | 0.05 | 0.25 | | | | | | |
| | | Sham | 0.141 | 0.049 | 0.04 | 0.24 | | | | | | |
| | | Active | | | | | Sham | 0.011 | 0.066 | -0.12 | 0.14 | 0.872 |
| | DAY 56 | Active | 0.191 | 0.051 | 0.09 | 0.29 | | | | | | |
| | | Sham | 0.136 | 0.051 | 0.03 | 0.24 | | | | | | |
| | | Active | | | | | Sham | 0.055 | 0.070 | -0.09 | 0.20 | 0.433 |
| | DAY 84 | Active | 0.133 | 0.046 | 0.04 | 0.23 | | | | | | |
| | | Sham | 0.113 | 0.048 | 0.02 | 0.21 | | | | | | |
| | | Active | | | | | Sham | 0.020 | 0.064 | -0.11 | 0.15 | 0.760 |

*p<0.05 by analysis of covariance of the change from baseline to days 14, 28, 56 and 84 (no device) in Creyos parameters, with device, visit, treatment by visit interaction, age >=45 years (yes, no) and baseline covariate. Confidence interval (CI), Least-squares (LS) mean. A greater percentile is in the direction of improved function. Source: Listing 12. 31DEC24, T05_creyos

Table 5

Analysis of Covariance
Creyos Treatment Device Comparison of Change from Baseline by Parameter (Per-Protocol Population)

| | | Treatment Device Estimate | | | | | Treatment Device Comparison | | | | | |
|---|---|---|---|---|---|---|---|---|---|---|---|---|
| | | Treatment | | Std | 95% CI | | | Difference | | 95% CI | | |
| Parameter | Visit | Device | LS Mean | Error | Lower | Upper | Comparator | LS Mean | Std Error | Lower | Upper | P-Value |
| Odd One Out | BASELINE | Active | 0.635 | 0.056 | 0.58 | 0.69 | | | | | | |
| | | Sham | 0.682 | 0.044 | 0.64 | 0.73 | | | | | | |
| | | Active | | | | | Sham | -0.047 | 0.071 | -0.19 | 0.10 | 0.510 |
| | DAY 14 | Active | -0.092 | 0.061 | -0.22 | 0.03 | | | | | | |
| | | Sham | -0.051 | 0.062 | -0.18 | 0.08 | | | | | | |
| | | Active | | | | | Sham | -0.042 | 0.086 | -0.22 | 0.13 | 0.632 |
| | DAY 28 | Active | -0.064 | 0.043 | -0.15 | 0.02 | | | | | | |
| | | Sham | -0.055 | 0.045 | -0.15 | 0.04 | | | | | | |
| | | Active | | | | | Sham | -0.009 | 0.061 | -0.13 | 0.11 | 0.883 |
| | DAY 56 | Active | -0.039 | 0.058 | -0.16 | 0.08 | | | | | | |
| | | Sham | -0.075 | 0.059 | -0.19 | 0.04 | | | | | | |
| | | Active | | | | | Sham | 0.036 | 0.081 | -0.13 | 0.20 | 0.663 |
| | DAY 84 | Active | -0.097 | 0.057 | -0.21 | 0.02 | | | | | | |
| | | Sham | -0.054 | 0.062 | -0.18 | 0.07 | | | | | | |
| | | Active | | | | | Sham | -0.043 | 0.083 | -0.21 | 0.13 | 0.606 |
| Double Trouble | BASELINE | Active | 0.597 | 0.082 | 0.52 | 0.68 | | | | | | |
| | | Sham | 0.521 | 0.075 | 0.45 | 0.60 | | | | | | |
| | | Active | | | | | Sham | 0.075 | 0.111 | -0.15 | 0.30 | 0.501 |
| | DAY 14 | Active | 0.177 | 0.044 | 0.09 | 0.27 | | | | | | |
| | | Sham | 0.102 | 0.045 | 0.01 | 0.19 | | | | | | |
| | | Active | | | | | Sham | 0.075 | 0.061 | -0.05 | 0.20 | 0.221 |
| | DAY 28 | Active | 0.230 | 0.046 | 0.14 | 0.32 | | | | | | |
| | | Sham | 0.155 | 0.047 | 0.06 | 0.25 | | | | | | |
| | | Active | | | | | Sham | 0.074 | 0.063 | -0.05 | 0.20 | 0.246 |
| | DAY 56 | Active | 0.214 | 0.046 | 0.12 | 0.31 | | | | | | |
| | | Sham | 0.212 | 0.047 | 0.12 | 0.31 | | | | | | |
| | | Active | | | | | Sham | 0.002 | 0.064 | -0.13 | 0.13 | 0.975 |
| | DAY 84 | Active | 0.288 | 0.049 | 0.19 | 0.39 | | | | | | |
| | | Sham | 0.192 | 0.052 | 0.09 | 0.30 | | | | | | |
| | | Active | | | | | Sham | 0.095 | 0.069 | -0.05 | 0.24 | 0.177 |

*p<0.05 by analysis of covariance of the change from baseline to days 14, 28, 56 and 84 (no device) in Creyos parameters, with device, visit, treatment by visit interaction, age >=45 years (yes, no) and baseline covariate. Confidence interval (CI), Least-squares (LS) mean. A greater percentile is in the direction of improved function. Source: Listing 12. 31DEC24, T05_creyos

Table 5.1

Analysis of Covariance
Creyos Treatment Device Comparison of Change from Baseline by Parameter
Age < 45 Years (Per-Protocol Population)

| | | Treatment Device Estimate | | | | | Treatment Device Comparison | | | | | |
|---|---|---|---|---|---|---|---|---|---|---|---|---|
| | | | | | 95% CI | | | Difference | | 95% CI | | |
| Parameter | Visit | Treatment Device | LS Mean | Std Error | Lower | Upper | Comparator | LS Mean | Std Error | Lower | Upper | P-Value |
| Spatial Planning | BASELINE | Active | 0.698 | 0.053 | 0.64 | 0.75 | | | | | | |
| | | Sham | 0.784 | 0.045 | 0.74 | 0.83 | | | | | | |
| | | Active | | | | | Sham | -0.086 | 0.070 | -0.23 | 0.06 | 0.226 |
| | DAY 14 | Active | 0.092 | 0.053 | -0.02 | 0.20 | | | | | | |
| | | Sham | 0.092 | 0.055 | -0.02 | 0.20 | | | | | | |
| | | Active | | | | | Sham | 0.001 | 0.077 | -0.16 | 0.16 | 0.993 |
| | DAY 28 | Active | 0.082 | 0.037 | 0.01 | 0.16 | | | | | | |
| | | Sham | 0.093 | 0.039 | 0.01 | 0.17 | | | | | | |
| | | Active | | | | | Sham | -0.011 | 0.054 | -0.12 | 0.10 | 0.846 |
| | DAY 56 | Active | 0.228 | 0.019 | 0.19 | 0.27 | | | | | | |
| | | Sham | 0.197 | 0.019 | 0.16 | 0.24 | | | | | | |
| | | Active | | | | | Sham | 0.031 | 0.027 | -0.03 | 0.09 | 0.271 |
| | DAY 84 | Active | 0.182 | 0.032 | 0.12 | 0.25 | | | | | | |
| | | Sham | 0.143 | 0.035 | 0.07 | 0.21 | | | | | | |
| | | Active | | | | | Sham | 0.040 | 0.048 | -0.06 | 0.14 | 0.414 |
| Monkey Ladder | BASELINE | Active | 0.841 | 0.039 | 0.80 | 0.88 | | | | | | |
| | | Sham | 0.775 | 0.024 | 0.75 | 0.80 | | | | | | |
| | | Active | | | | | Sham | 0.065 | 0.047 | -0.03 | 0.16 | 0.175 |
| | DAY 14 | Active | -0.067 | 0.037 | -0.14 | 0.01 | | | | | | |
| | | Sham | -0.066 | 0.038 | -0.14 | 0.01 | | | | | | |
| | | Active | | | | | Sham | -0.001 | 0.054 | -0.11 | 0.11 | 0.986 |
| | DAY 28 | Active | -0.045 | 0.038 | -0.12 | 0.03 | | | | | | |
| | | Sham | -0.015 | 0.041 | -0.10 | 0.07 | | | | | | |
| | | Active | | | | | Sham | -0.030 | 0.057 | -0.15 | 0.09 | 0.597 |
| | DAY 56 | Active | -0.021 | 0.040 | -0.10 | 0.06 | | | | | | |
| | | Sham | -0.032 | 0.042 | -0.12 | 0.05 | | | | | | |
| | | Active | | | | | Sham | 0.010 | 0.059 | -0.11 | 0.13 | 0.861 |
| | DAY 84 | Active | -0.050 | 0.040 | -0.13 | 0.03 | | | | | | |
| | | Sham | 0.000 | 0.042 | -0.09 | 0.09 | | | | | | |
| | | Active | | | | | Sham | -0.050 | 0.059 | -0.17 | 0.07 | 0.403 |
| Rotations | BASELINE | Active | 0.715 | 0.082 | 0.63 | 0.80 | | | | | | |
| | | Sham | 0.679 | 0.072 | 0.61 | 0.75 | | | | | | |
| | | Active | | | | | Sham | 0.036 | 0.110 | -0.19 | 0.26 | 0.747 |

*p<0.05 by analysis of covariance of the change from baseline to days 14, 28, 56 and 84 (no device) in Creyos parameters, with device, visit, treatment by visit interaction, and baseline covariate. Confidence interval (CI), Least-squares (LS) mean.
A greater percentile is in the direction of improved function. Source: Listing 12. 31DEC24, T05_creyos_lt45

Table 5.1

Analysis of Covariance
Creyos Treatment Device Comparison of Change from Baseline by Parameter
Age < 45 Years (Per-Protocol Population)

| | | Treatment Device Estimate | | | | | Treatment Device Comparison | | | | | |
|---|---|---|---|---|---|---|---|---|---|---|---|---|
| | | | | | 95% CI | | | Difference | | 95% CI | | |
| Parameter | Visit | Treatment Device | LS Mean | Std Error | Lower | Upper | Comparator | LS Mean | Std Error | Lower | Upper | P-Value |
| Rotations | DAY 14 | Active | 0.130 | 0.061 | 0.00 | 0.26 | | | | | | |
| | | Sham | -0.013 | 0.063 | -0.14 | 0.12 | | | | | | |
| | | Active | | | | | Sham | 0.143 | 0.088 | -0.04 | 0.32 | 0.115 |
| | DAY 28 | Active | 0.050 | 0.065 | -0.08 | 0.18 | | | | | | |
| | | Sham | -0.002 | 0.069 | -0.14 | 0.14 | | | | | | |
| | | Active | | | | | Sham | 0.052 | 0.095 | -0.14 | 0.25 | 0.590 |
| | DAY 56 | Active | 0.123 | 0.058 | 0.00 | 0.24 | | | | | | |
| | | Sham | 0.053 | 0.060 | -0.07 | 0.18 | | | | | | |
| | | Active | | | | | Sham | 0.069 | 0.084 | -0.10 | 0.24 | 0.419 |
| | DAY 84 | Active | 0.117 | 0.061 | -0.01 | 0.24 | | | | | | |
| | | Sham | 0.036 | 0.065 | -0.10 | 0.17 | | | | | | |
| | | Active | | | | | Sham | 0.081 | 0.089 | -0.10 | 0.26 | 0.372 |
| Feature Match | BASELINE | Active | 0.770 | 0.059 | 0.71 | 0.83 | | | | | | |
| | | Sham | 0.743 | 0.066 | 0.68 | 0.81 | | | | | | |
| | | Active | | | | | Sham | 0.027 | 0.089 | -0.15 | 0.21 | 0.763 |
| | DAY 14 | Active | 0.059 | 0.055 | -0.05 | 0.17 | | | | | | |
| | | Sham | -0.048 | 0.057 | -0.16 | 0.07 | | | | | | |
| | | Active | | | | | Sham | 0.107 | 0.079 | -0.06 | 0.27 | 0.189 |
| | DAY 28 | Active | 0.134 | 0.053 | 0.02 | 0.24 | | | | | | |
| | | Sham | -0.012 | 0.058 | -0.13 | 0.11 | | | | | | |
| | | Active | | | | | Sham | 0.147 | 0.079 | -0.02 | 0.31 | 0.074 |
| | DAY 56 | Active | 0.107 | 0.061 | -0.02 | 0.23 | | | | | | |
| | | Sham | -0.016 | 0.063 | -0.15 | 0.11 | | | | | | |
| | | Active | | | | | Sham | 0.123 | 0.088 | -0.06 | 0.30 | 0.171 |
| | DAY 84 | Active | 0.137 | 0.049 | 0.04 | 0.24 | | | | | | |
| | | Sham | -0.029 | 0.052 | -0.14 | 0.08 | | | | | | |
| | | Active | | | | | Sham | 0.166 | 0.071 | 0.02 | 0.31 | 0.028* |
| Paired Associates | BASELINE | Active | 0.741 | 0.070 | 0.67 | 0.81 | | | | | | |
| | | Sham | 0.726 | 0.064 | 0.66 | 0.79 | | | | | | |
| | | Active | | | | | Sham | 0.015 | 0.096 | -0.18 | 0.21 | 0.878 |
| | DAY 14 | Active | 0.038 | 0.064 | -0.09 | 0.17 | | | | | | |
| | | Sham | -0.099 | 0.066 | -0.23 | 0.04 | | | | | | |
| | | Active | | | | | Sham | 0.137 | 0.092 | -0.05 | 0.33 | 0.148 |

*p<0.05 by analysis of covariance of the change from baseline to days 14, 28, 56 and 84 (no device) in Creyos parameters, with device, visit, treatment by visit interaction, and baseline covariate. Confidence interval (CI), Least-squares (LS) mean.
A greater percentile is in the direction of improved function. Source: Listing 12. 31DEC24, T05_creyos_lt45

Table 5.1

Analysis of Covariance
Creyos Treatment Device Comparison of Change from Baseline by Parameter
Age < 45 Years (Per-Protocol Population)

| | | Treatment Device Estimate | | | | | Treatment Device Comparison | | | | | |
|---|---|---|---|---|---|---|---|---|---|---|---|---|
| | | | | | 95% CI | | | Difference | | 95% CI | | |
| Parameter | Visit | Treatment Device | LS Mean | Std Error | Lower | Upper | Comparator | LS Mean | Std Error | Lower | Upper | P-Value |
| Paired Associates | DAY 28 | Active | -0.027 | 0.058 | -0.15 | 0.09 | | | | | | |
| | | Sham | -0.048 | 0.063 | -0.18 | 0.08 | | | | | | |
| | | Active | | | | | Sham | 0.020 | 0.086 | -0.16 | 0.20 | 0.813 |
| | DAY 56 | Active | -0.016 | 0.047 | -0.11 | 0.08 | | | | | | |
| | | Sham | -0.022 | 0.049 | -0.12 | 0.08 | | | | | | |
| | | Active | | | | | Sham | 0.006 | 0.068 | -0.13 | 0.14 | 0.931 |
| | DAY 84 | Active | 0.028 | 0.056 | -0.09 | 0.14 | | | | | | |
| | | Sham | -0.030 | 0.060 | -0.15 | 0.09 | | | | | | |
| | | Active | | | | | Sham | 0.057 | 0.082 | -0.11 | 0.23 | 0.492 |
| Token Search | BASELINE | Active | 0.840 | 0.035 | 0.80 | 0.87 | | | | | | |
| | | Sham | 0.788 | 0.033 | 0.76 | 0.82 | | | | | | |
| | | Active | | | | | Sham | 0.051 | 0.048 | -0.05 | 0.15 | 0.297 |
| | DAY 14 | Active | 0.043 | 0.023 | -0.00 | 0.09 | | | | | | |
| | | Sham | 0.012 | 0.024 | -0.04 | 0.06 | | | | | | |
| | | Active | | | | | Sham | 0.031 | 0.033 | -0.04 | 0.10 | 0.357 |
| | DAY 28 | Active | 0.023 | 0.030 | -0.04 | 0.09 | | | | | | |
| | | Sham | 0.026 | 0.032 | -0.04 | 0.09 | | | | | | |
| | | Active | | | | | Sham | -0.003 | 0.045 | -0.09 | 0.09 | 0.943 |
| | DAY 56 | Active | 0.037 | 0.017 | 0.00 | 0.07 | | | | | | |
| | | Sham | -0.006 | 0.017 | -0.04 | 0.03 | | | | | | |
| | | Active | | | | | Sham | 0.043 | 0.024 | -0.01 | 0.09 | 0.092 |
| | DAY 84 | Active | 0.047 | 0.022 | 0.00 | 0.09 | | | | | | |
| | | Sham | 0.036 | 0.024 | -0.01 | 0.08 | | | | | | |
| | | Active | | | | | Sham | 0.011 | 0.032 | -0.06 | 0.08 | 0.731 |
| Polygons | BASELINE | Active | 0.683 | 0.075 | 0.61 | 0.76 | | | | | | |
| | | Sham | 0.660 | 0.074 | 0.59 | 0.73 | | | | | | |
| | | Active | | | | | Sham | 0.023 | 0.105 | -0.19 | 0.24 | 0.831 |
| | DAY 14 | Active | -0.043 | 0.068 | -0.18 | 0.10 | | | | | | |
| | | Sham | -0.138 | 0.070 | -0.28 | 0.01 | | | | | | |
| | | Active | | | | | Sham | 0.095 | 0.097 | -0.11 | 0.29 | 0.340 |
| | DAY 28 | Active | 0.153 | 0.048 | 0.05 | 0.25 | | | | | | |
| | | Sham | -0.001 | 0.052 | -0.11 | 0.10 | | | | | | |
| | | Active | | | | | Sham | 0.154 | 0.071 | 0.01 | 0.30 | 0.039* |

*p<0.05 by analysis of covariance of the change from baseline to days 14, 28, 56 and 84 (no device) in Creyos parameters, with device, visit, treatment by visit interaction, and baseline covariate. Confidence interval (CI), Least-squares (LS) mean.
A greater percentile is in the direction of improved function. Source: Listing 12. 31DEC24, T05_creyos_lt45

Table 5.1

Analysis of Covariance
Creyos Treatment Device Comparison of Change from Baseline by Parameter
Age < 45 Years (Per-Protocol Population)

| | | Treatment Device Estimate | | | | | Treatment Device Comparison | | | | | |
|---|---|---|---|---|---|---|---|---|---|---|---|---|
| | | Treatment | | Std | 95% CI | | | Difference | | 95% CI | | |
| Parameter | Visit | Device | LS Mean | Error | Lower | Upper | Comparator | LS Mean | Std Error | Lower | Upper | P-Value |
| Polygons | DAY 56 | Active | 0.124 | 0.055 | 0.01 | 0.24 | | | | | | |
| | | Sham | -0.026 | 0.056 | -0.14 | 0.09 | | | | | | |
| | | Active | | | | | Sham | 0.150 | 0.079 | -0.01 | 0.31 | 0.066 |
| | DAY 84 | Active | 0.044 | 0.061 | -0.08 | 0.17 | | | | | | |
| | | Sham | 0.059 | 0.066 | -0.08 | 0.20 | | | | | | |
| | | Active | | | | | Sham | -0.016 | 0.090 | -0.20 | 0.17 | 0.864 |
| Grammatical Reasoning | BASELINE | Active | 0.693 | 0.077 | 0.62 | 0.77 | | | | | | |
| | | Sham | 0.763 | 0.067 | 0.70 | 0.83 | | | | | | |
| | | Active | | | | | Sham | -0.070 | 0.103 | -0.28 | 0.14 | 0.501 |
| | DAY 14 | Active | 0.032 | 0.043 | -0.06 | 0.12 | | | | | | |
| | | Sham | 0.059 | 0.044 | -0.03 | 0.15 | | | | | | |
| | | Active | | | | | Sham | -0.027 | 0.061 | -0.15 | 0.10 | 0.669 |
| | DAY 28 | Active | 0.104 | 0.045 | 0.01 | 0.20 | | | | | | |
| | | Sham | 0.085 | 0.048 | -0.01 | 0.18 | | | | | | |
| | | Active | | | | | Sham | 0.019 | 0.066 | -0.12 | 0.16 | 0.772 |
| | DAY 56 | Active | 0.127 | 0.038 | 0.05 | 0.20 | | | | | | |
| | | Sham | 0.094 | 0.039 | 0.01 | 0.17 | | | | | | |
| | | Active | | | | | Sham | 0.033 | 0.054 | -0.08 | 0.14 | 0.547 |
| | DAY 84 | Active | 0.093 | 0.038 | 0.01 | 0.17 | | | | | | |
| | | Sham | 0.117 | 0.041 | 0.03 | 0.20 | | | | | | |
| | | Active | | | | | Sham | -0.024 | 0.056 | -0.14 | 0.09 | 0.671 |
| Spatial Span | BASELINE | Active | 0.820 | 0.047 | 0.77 | 0.87 | | | | | | |
| | | Sham | 0.817 | 0.049 | 0.77 | 0.87 | | | | | | |
| | | Active | | | | | Sham | 0.004 | 0.068 | -0.14 | 0.14 | 0.958 |
| | DAY 14 | Active | -0.069 | 0.060 | -0.19 | 0.05 | | | | | | |
| | | Sham | 0.035 | 0.062 | -0.09 | 0.16 | | | | | | |
| | | Active | | | | | Sham | -0.104 | 0.087 | -0.28 | 0.07 | 0.242 |
| | DAY 28 | Active | -0.025 | 0.050 | -0.13 | 0.08 | | | | | | |
| | | Sham | -0.081 | 0.053 | -0.19 | 0.03 | | | | | | |
| | | Active | | | | | Sham | 0.056 | 0.073 | -0.09 | 0.21 | 0.446 |
| | DAY 56 | Active | -0.033 | 0.049 | -0.13 | 0.07 | | | | | | |
| | | Sham | -0.011 | 0.050 | -0.11 | 0.09 | | | | | | |

*p<0.05 by analysis of covariance of the change from baseline to days 14, 28, 56 and 84 (no device) in Creyos parameters, with device, visit, treatment by visit interaction, and baseline covariate. Confidence interval (CI), Least-squares (LS) mean.
A greater percentile is in the direction of improved function. Source: Listing 12. 31DEC24, T05_creyos_lt45

Table 5.1

Analysis of Covariance
Creyos Treatment Device Comparison of Change from Baseline by Parameter
Age < 45 Years (Per-Protocol Population)

| | | Treatment Device Estimate | | | | | Treatment Device Comparison | | | | | |
|---|---|---|---|---|---|---|---|---|---|---|---|---|
| | | | | | 95% CI | | | Difference | | 95% CI | | |
| Parameter | Visit | Treatment Device | LS Mean | Std Error | Lower | Upper | Comparator | LS Mean | Std Error | Lower | Upper | P-Value |
| Spatial Span | DAY 56 | Active | | | | | Sham | -0.022 | 0.070 | -0.17 | 0.12 | 0.758 |
| | DAY 84 | Active | 0.017 | 0.043 | -0.07 | 0.11 | | | | | | |
| | | Sham | 0.013 | 0.047 | -0.08 | 0.11 | | | | | | |
| | | Active | | | | | Sham | 0.004 | 0.064 | -0.13 | 0.14 | 0.947 |
| Digit Span | BASELINE | Active | 0.564 | 0.053 | 0.51 | 0.62 | | | | | | |
| | | Sham | 0.466 | 0.071 | 0.40 | 0.54 | | | | | | |
| | | Active | | | | | Sham | 0.098 | 0.087 | -0.08 | 0.28 | 0.271 |
| | DAY 14 | Active | 0.050 | 0.057 | -0.07 | 0.17 | | | | | | |
| | | Sham | 0.122 | 0.059 | 0.00 | 0.24 | | | | | | |
| | | Active | | | | | Sham | -0.072 | 0.083 | -0.24 | 0.10 | 0.390 |
| | DAY 28 | Active | 0.193 | 0.050 | 0.09 | 0.29 | | | | | | |
| | | Sham | 0.217 | 0.053 | 0.11 | 0.33 | | | | | | |
| | | Active | | | | | Sham | -0.024 | 0.073 | -0.17 | 0.13 | 0.747 |
| | DAY 56 | Active | 0.213 | 0.057 | 0.10 | 0.33 | | | | | | |
| | | Sham | 0.194 | 0.059 | 0.07 | 0.31 | | | | | | |
| | | Active | | | | | Sham | 0.019 | 0.082 | -0.15 | 0.19 | 0.816 |
| | DAY 84 | Active | 0.147 | 0.053 | 0.04 | 0.26 | | | | | | |
| | | Sham | 0.134 | 0.056 | 0.02 | 0.25 | | | | | | |
| | | Active | | | | | Sham | 0.014 | 0.077 | -0.15 | 0.17 | 0.862 |
| Odd One Out | BASELINE | Active | 0.675 | 0.066 | 0.61 | 0.74 | | | | | | |
| | | Sham | 0.694 | 0.057 | 0.64 | 0.75 | | | | | | |
| | | Active | | | | | Sham | -0.020 | 0.088 | -0.20 | 0.16 | 0.824 |
| | DAY 14 | Active | -0.144 | 0.066 | -0.28 | -0.01 | | | | | | |
| | | Sham | 0.010 | 0.069 | -0.13 | 0.15 | | | | | | |
| | | Active | | | | | Sham | -0.154 | 0.096 | -0.35 | 0.04 | 0.120 |
| | DAY 28 | Active | -0.032 | 0.044 | -0.12 | 0.06 | | | | | | |
| | | Sham | 0.049 | 0.047 | -0.05 | 0.15 | | | | | | |
| | | Active | | | | | Sham | -0.081 | 0.064 | -0.21 | 0.05 | 0.221 |
| | DAY 56 | Active | 0.028 | 0.057 | -0.09 | 0.15 | | | | | | |
| | | Sham | -0.045 | 0.059 | -0.17 | 0.08 | | | | | | |
| | | Active | | | | | Sham | 0.073 | 0.082 | -0.10 | 0.24 | 0.381 |
| | DAY 84 | Active | -0.068 | 0.068 | -0.21 | 0.07 | | | | | | |

*p<0.05 by analysis of covariance of the change from baseline to days 14, 28, 56 and 84 (no device) in Creyos parameters, with device, visit, treatment by visit interaction, and baseline covariate. Confidence interval (CI), Least-squares (LS) mean.
A greater percentile is in the direction of improved function. Source: Listing 12. 31DEC24, T05_creyos_lt45

Table 5.1

Analysis of Covariance
Creyos Treatment Device Comparison of Change from Baseline by Parameter
Age < 45 Years (Per-Protocol Population)

| | | Treatment Device Estimate | | | | | Treatment Device Comparison | | | | | |
|---|---|---|---|---|---|---|---|---|---|---|---|---|
| | | Treatment | | Std | 95% CI | | | Difference | | 95% CI | | |
| Parameter | Visit | Device | LS Mean | Error | Lower | Upper | Comparator | LS Mean | Std Error | Lower | Upper | P-Value |
| Odd One Out | DAY 84 | Sham | -0.056 | 0.073 | -0.21 | 0.09 | | | | | | |
| | | Active | | | | | Sham | -0.012 | 0.099 | -0.22 | 0.19 | 0.905 |
| Double Trouble | BASELINE | Active | 0.692 | 0.089 | 0.60 | 0.78 | | | | | | |
| | | Sham | 0.523 | 0.082 | 0.44 | 0.61 | | | | | | |
| | | Active | | | | | Sham | 0.169 | 0.122 | -0.08 | 0.42 | 0.176 |
| | DAY 14 | Active | 0.233 | 0.052 | 0.13 | 0.34 | | | | | | |
| | | Sham | 0.135 | 0.054 | 0.02 | 0.25 | | | | | | |
| | | Active | | | | | Sham | 0.098 | 0.076 | -0.06 | 0.25 | 0.207 |
| | DAY 28 | Active | 0.299 | 0.050 | 0.20 | 0.40 | | | | | | |
| | | Sham | 0.165 | 0.052 | 0.06 | 0.27 | | | | | | |
| | | Active | | | | | Sham | 0.134 | 0.073 | -0.02 | 0.28 | 0.079 |
| | DAY 56 | Active | 0.295 | 0.047 | 0.20 | 0.39 | | | | | | |
| | | Sham | 0.202 | 0.049 | 0.10 | 0.30 | | | | | | |
| | | Active | | | | | Sham | 0.093 | 0.069 | -0.05 | 0.23 | 0.187 |
| | DAY 84 | Active | 0.301 | 0.053 | 0.19 | 0.41 | | | | | | |
| | | Sham | 0.190 | 0.057 | 0.07 | 0.31 | | | | | | |
| | | Active | | | | | Sham | 0.111 | 0.079 | -0.05 | 0.27 | 0.170 |

*p<0.05 by analysis of covariance of the change from baseline to days 14, 28, 56 and 84 (no device) in Creyos parameters, with device, visit, treatment by visit interaction, and baseline covariate. Confidence interval (CI), Least-squares (LS) mean.
A greater percentile is in the direction of improved function. Source: Listing 12. 31DEC24, T05_creyos_lt45

Table 5.2

Analysis of Covariance
Creyos Treatment Device Comparison of Change from Baseline by Parameter
Age >= 45 Years (Per-Protocol Population)

| | | Treatment Device Estimate | | | | | Treatment Device Comparison | | | | | |
|---|---|---|---|---|---|---|---|---|---|---|---|---|
| | | | | | 95% CI | | | Difference | | 95% CI | | |
| Parameter | Visit | Treatment Device | LS Mean | Std Error | Lower | Upper | Comparator | LS Mean | Std Error | Lower | Upper | P-Value |
| Spatial Planning | BASELINE | Active | 0.762 | 0.101 | 0.66 | 0.86 | | | | | | |
| | | Sham | 0.672 | 0.063 | 0.61 | 0.73 | | | | | | |
| | | Active | | | | | Sham | 0.090 | 0.119 | -0.18 | 0.35 | 0.469 |
| | DAY 14 | Active | -0.007 | 0.077 | -0.18 | 0.17 | | | | | | |
| | | Sham | -0.055 | 0.078 | -0.23 | 0.12 | | | | | | |
| | | Active | | | | | Sham | 0.048 | 0.111 | -0.20 | 0.30 | 0.678 |
| | DAY 28 | Active | 0.073 | 0.080 | -0.11 | 0.26 | | | | | | |
| | | Sham | -0.145 | 0.081 | -0.33 | 0.04 | | | | | | |
| | | Active | | | | | Sham | 0.218 | 0.116 | -0.04 | 0.48 | 0.091 |
| | DAY 56 | Active | 0.049 | 0.074 | -0.12 | 0.22 | | | | | | |
| | | Sham | 0.066 | 0.074 | -0.10 | 0.23 | | | | | | |
| | | Active | | | | | Sham | -0.017 | 0.106 | -0.26 | 0.22 | 0.876 |
| | DAY 84 | Active | 0.112 | 0.079 | -0.07 | 0.29 | | | | | | |
| | | Sham | 0.101 | 0.082 | -0.08 | 0.28 | | | | | | |
| | | Active | | | | | Sham | 0.012 | 0.115 | -0.25 | 0.27 | 0.922 |
| Monkey Ladder | BASELINE | Active | 0.646 | 0.084 | 0.56 | 0.73 | | | | | | |
| | | Sham | 0.621 | 0.051 | 0.57 | 0.67 | | | | | | |
| | | Active | | | | | Sham | 0.025 | 0.098 | -0.19 | 0.24 | 0.806 |
| | DAY 14 | Active | 0.092 | 0.049 | -0.02 | 0.20 | | | | | | |
| | | Sham | 0.100 | 0.049 | -0.01 | 0.21 | | | | | | |
| | | Active | | | | | Sham | -0.008 | 0.070 | -0.16 | 0.15 | 0.914 |
| | DAY 28 | Active | -0.047 | 0.056 | -0.17 | 0.08 | | | | | | |
| | | Sham | 0.052 | 0.056 | -0.07 | 0.18 | | | | | | |
| | | Active | | | | | Sham | -0.099 | 0.079 | -0.28 | 0.08 | 0.240 |
| | DAY 56 | Active | 0.123 | 0.065 | -0.02 | 0.27 | | | | | | |
| | | Sham | 0.010 | 0.065 | -0.13 | 0.15 | | | | | | |
| | | Active | | | | | Sham | 0.113 | 0.092 | -0.09 | 0.32 | 0.244 |
| | DAY 84 | Active | 0.101 | 0.099 | -0.12 | 0.33 | | | | | | |
| | | Sham | -0.119 | 0.111 | -0.37 | 0.13 | | | | | | |
| | | Active | | | | | Sham | 0.220 | 0.149 | -0.11 | 0.55 | 0.172 |
| Rotations | BASELINE | Active | 0.574 | 0.135 | 0.44 | 0.71 | | | | | | |
| | | Sham | 0.782 | 0.104 | 0.68 | 0.89 | | | | | | |
| | | Active | | | | | Sham | -0.208 | 0.170 | -0.59 | 0.17 | 0.249 |

*p<0.05 by analysis of covariance of the change from baseline to days 14, 28, 56 and 84 (no device) in Creyos parameters, with device, visit, treatment by visit interaction, and baseline covariate. Confidence interval (CI), Least-squares (LS) mean.
A greater percentile is in the direction of improved function. Source: Listing 12. 31DEC24, T05_creyos_ge45

Table 5.2

Analysis of Covariance
Creyos Treatment Device Comparison of Change from Baseline by Parameter
Age >= 45 Years (Per-Protocol Population)

| | | Treatment Device Estimate | | | | | Treatment Device Comparison | | | | | |
|---|---|---|---|---|---|---|---|---|---|---|---|---|
| | | | | | 95% CI | | | Difference | | 95% CI | | |
| Parameter | Visit | Treatment Device | LS Mean | Std Error | Lower | Upper | Comparator | LS Mean | Std Error | Lower | Upper | P-Value |
| Rotations | DAY 14 | Active | -0.001 | 0.094 | -0.21 | 0.21 | | | | | | |
| | | Sham | -0.057 | 0.094 | -0.27 | 0.15 | | | | | | |
| | | Active | | | | | Sham | 0.056 | 0.134 | -0.24 | 0.35 | 0.688 |
| | DAY 28 | Active | -0.019 | 0.064 | -0.16 | 0.12 | | | | | | |
| | | Sham | 0.017 | 0.064 | -0.13 | 0.16 | | | | | | |
| | | Active | | | | | Sham | -0.035 | 0.091 | -0.24 | 0.17 | 0.709 |
| | DAY 56 | Active | 0.079 | 0.115 | -0.18 | 0.33 | | | | | | |
| | | Sham | -0.087 | 0.115 | -0.34 | 0.17 | | | | | | |
| | | Active | | | | | Sham | 0.166 | 0.163 | -0.20 | 0.53 | 0.333 |
| | DAY 84 | Active | 0.078 | 0.147 | -0.33 | 0.48 | | | | | | |
| | | Sham | -0.260 | 0.160 | -0.68 | 0.16 | | | | | | |
| | | Active | | | | | Sham | 0.338 | 0.218 | -0.25 | 0.92 | 0.189 |
| Feature Match | BASELINE | Active | 0.442 | 0.110 | 0.33 | 0.55 | | | | | | |
| | | Sham | 0.824 | 0.076 | 0.75 | 0.90 | | | | | | |
| | | Active | | | | | Sham | -0.382 | 0.134 | -0.68 | -0.08 | 0.017* |
| | DAY 14 | Active | 0.111 | 0.138 | -0.19 | 0.41 | | | | | | |
| | | Sham | -0.255 | 0.137 | -0.56 | 0.05 | | | | | | |
| | | Active | | | | | Sham | 0.366 | 0.206 | -0.08 | 0.81 | 0.100 |
| | DAY 28 | Active | 0.179 | 0.113 | -0.07 | 0.43 | | | | | | |
| | | Sham | -0.275 | 0.112 | -0.53 | -0.03 | | | | | | |
| | | Active | | | | | Sham | 0.455 | 0.173 | 0.07 | 0.83 | 0.023* |
| | DAY 56 | Active | 0.141 | 0.078 | -0.03 | 0.31 | | | | | | |
| | | Sham | -0.176 | 0.077 | -0.35 | -0.00 | | | | | | |
| | | Active | | | | | Sham | 0.317 | 0.129 | 0.03 | 0.60 | 0.034* |
| | DAY 84 | Active | 0.170 | 0.143 | -0.16 | 0.50 | | | | | | |
| | | Sham | -0.261 | 0.153 | -0.60 | 0.08 | | | | | | |
| | | Active | | | | | Sham | 0.431 | 0.219 | -0.05 | 0.92 | 0.076 |
| Paired Associates | BASELINE | Active | 0.651 | 0.077 | 0.57 | 0.73 | | | | | | |
| | | Sham | 0.642 | 0.087 | 0.56 | 0.73 | | | | | | |
| | | Active | | | | | Sham | 0.008 | 0.116 | -0.25 | 0.27 | 0.945 |
| | DAY 14 | Active | -0.091 | 0.120 | -0.36 | 0.18 | | | | | | |
| | | Sham | 0.070 | 0.121 | -0.20 | 0.34 | | | | | | |
| | | Active | | | | | Sham | -0.162 | 0.170 | -0.54 | 0.22 | 0.367 |

*p<0.05 by analysis of covariance of the change from baseline to days 14, 28, 56 and 84 (no device) in Creyos parameters, with device, visit, treatment by visit interaction, and baseline covariate. Confidence interval (CI), Least-squares (LS) mean.
A greater percentile is in the direction of improved function. Source: Listing 12. 31DEC24, T05_creyos_ge45

Table 5.2

Analysis of Covariance
Creyos Treatment Device Comparison of Change from Baseline by Parameter
Age >= 45 Years (Per-Protocol Population)

| | | Treatment Device Estimate | | | | | Treatment Device Comparison | | | | | |
|---|---|---|---|---|---|---|---|---|---|---|---|---|
| | | Treatment | | Std | 95% CI | | | Difference | | 95% CI | | |
| Parameter | Visit | Device | LS Mean | Error | Lower | Upper | Comparator | LS Mean | Std Error | Lower | Upper | P-Value |
| Paired Associates | DAY 28 | Active | 0.055 | 0.084 | -0.14 | 0.25 | | | | | | |
| | | Sham | 0.100 | 0.084 | -0.09 | 0.29 | | | | | | |
| | | Active | | | | | Sham | -0.045 | 0.119 | -0.31 | 0.23 | 0.717 |
| | DAY 56 | Active | -0.037 | 0.088 | -0.23 | 0.16 | | | | | | |
| | | Sham | 0.033 | 0.088 | -0.16 | 0.23 | | | | | | |
| | | Active | | | | | Sham | -0.069 | 0.124 | -0.35 | 0.21 | 0.589 |
| | DAY 84 | Active | 0.016 | 0.099 | -0.21 | 0.24 | | | | | | |
| | | Sham | 0.005 | 0.105 | -0.23 | 0.24 | | | | | | |
| | | Active | | | | | Sham | 0.011 | 0.144 | -0.31 | 0.34 | 0.940 |
| Token Search | BASELINE | Active | 0.803 | 0.029 | 0.77 | 0.83 | | | | | | |
| | | Sham | 0.743 | 0.087 | 0.66 | 0.83 | | | | | | |
| | | Active | | | | | Sham | 0.060 | 0.091 | -0.14 | 0.26 | 0.526 |
| | DAY 14 | Active | -0.195 | 0.044 | -0.29 | -0.10 | | | | | | |
| | | Sham | -0.002 | 0.044 | -0.10 | 0.10 | | | | | | |
| | | Active | | | | | Sham | -0.192 | 0.063 | -0.34 | -0.05 | 0.014* |
| | DAY 28 | Active | 0.003 | 0.045 | -0.10 | 0.10 | | | | | | |
| | | Sham | -0.036 | 0.045 | -0.14 | 0.07 | | | | | | |
| | | Active | | | | | Sham | 0.039 | 0.064 | -0.11 | 0.18 | 0.555 |
| | DAY 56 | Active | -0.064 | 0.057 | -0.19 | 0.06 | | | | | | |
| | | Sham | 0.019 | 0.058 | -0.11 | 0.15 | | | | | | |
| | | Active | | | | | Sham | -0.083 | 0.082 | -0.27 | 0.10 | 0.334 |
| | DAY 84 | Active | -0.018 | 0.042 | -0.11 | 0.08 | | | | | | |
| | | Sham | 0.057 | 0.045 | -0.04 | 0.16 | | | | | | |
| | | Active | | | | | Sham | -0.075 | 0.062 | -0.21 | 0.07 | 0.258 |
| Polygons | BASELINE | Active | 0.604 | 0.136 | 0.47 | 0.74 | | | | | | |
| | | Sham | 0.396 | 0.098 | 0.30 | 0.49 | | | | | | |
| | | Active | | | | | Sham | 0.208 | 0.168 | -0.17 | 0.58 | 0.245 |
| | DAY 14 | Active | 0.059 | 0.123 | -0.22 | 0.34 | | | | | | |
| | | Sham | 0.027 | 0.123 | -0.25 | 0.31 | | | | | | |
| | | Active | | | | | Sham | 0.032 | 0.178 | -0.37 | 0.43 | 0.861 |
| | DAY 28 | Active | 0.101 | 0.098 | -0.12 | 0.32 | | | | | | |
| | | Sham | 0.173 | 0.098 | -0.05 | 0.40 | | | | | | |
| | | Active | | | | | Sham | -0.073 | 0.144 | -0.40 | 0.25 | 0.625 |

*p<0.05 by analysis of covariance of the change from baseline to days 14, 28, 56 and 84 (no device) in Creyos parameters, with device, visit, treatment by visit interaction, and baseline covariate. Confidence interval (CI), Least-squares (LS) mean.
A greater percentile is in the direction of improved function. Source: Listing 12. 31DEC24, T05_creyos_ge45

Table 5.2

Analysis of Covariance
Creyos Treatment Device Comparison of Change from Baseline by Parameter
Age >= 45 Years (Per-Protocol Population)

| Parameter | Visit | Treatment Device | Treatment Device Estimate: LS Mean | Treatment Device Estimate: Std Error | Treatment Device Estimate: 95% CI Lower | Treatment Device Estimate: 95% CI Upper | Treatment Device Comparison: Comparator | Difference: LS Mean | Difference: Std Error | Difference: 95% CI Lower | Difference: 95% CI Upper | P-Value |
|---|---|---|---|---|---|---|---|---|---|---|---|---|
| Polygons | DAY 56 | Active | 0.016 | 0.121 | -0.25 | 0.28 | | | | | | |
| | | Sham | 0.115 | 0.120 | -0.15 | 0.38 | | | | | | |
| | | Active | | | | | Sham | -0.099 | 0.174 | -0.49 | 0.29 | 0.584 |
| | DAY 84 | Active | 0.060 | 0.124 | -0.22 | 0.34 | | | | | | |
| | | Sham | 0.341 | 0.134 | 0.04 | 0.64 | | | | | | |
| | | Active | | | | | Sham | -0.281 | 0.187 | -0.70 | 0.13 | 0.163 |
| Grammatical Reasoning | BASELINE | Active | 0.477 | 0.116 | 0.36 | 0.59 | | | | | | |
| | | Sham | 0.799 | 0.116 | 0.68 | 0.91 | | | | | | |
| | | Active | | | | | Sham | -0.322 | 0.164 | -0.69 | 0.04 | 0.077 |
| | DAY 14 | Active | 0.089 | 0.110 | -0.16 | 0.34 | | | | | | |
| | | Sham | -0.095 | 0.110 | -0.34 | 0.15 | | | | | | |
| | | Active | | | | | Sham | 0.185 | 0.161 | -0.18 | 0.54 | 0.279 |
| | DAY 28 | Active | 0.058 | 0.064 | -0.09 | 0.20 | | | | | | |
| | | Sham | -0.070 | 0.065 | -0.22 | 0.08 | | | | | | |
| | | Active | | | | | Sham | 0.129 | 0.100 | -0.10 | 0.35 | 0.229 |
| | DAY 56 | Active | 0.161 | 0.096 | -0.06 | 0.38 | | | | | | |
| | | Sham | -0.102 | 0.097 | -0.32 | 0.12 | | | | | | |
| | | Active | | | | | Sham | 0.263 | 0.143 | -0.06 | 0.58 | 0.098 |
| | DAY 84 | Active | 0.152 | 0.100 | -0.07 | 0.38 | | | | | | |
| | | Sham | -0.076 | 0.110 | -0.32 | 0.17 | | | | | | |
| | | Active | | | | | Sham | 0.228 | 0.154 | -0.12 | 0.57 | 0.172 |
| Spatial Span | BASELINE | Active | 0.712 | 0.121 | 0.59 | 0.83 | | | | | | |
| | | Sham | 0.568 | 0.144 | 0.42 | 0.71 | | | | | | |
| | | Active | | | | | Sham | 0.144 | 0.188 | -0.27 | 0.56 | 0.461 |
| | DAY 14 | Active | 0.120 | 0.084 | -0.07 | 0.31 | | | | | | |
| | | Sham | 0.107 | 0.084 | -0.08 | 0.29 | | | | | | |
| | | Active | | | | | Sham | 0.013 | 0.119 | -0.25 | 0.28 | 0.914 |
| | DAY 28 | Active | 0.137 | 0.091 | -0.07 | 0.34 | | | | | | |
| | | Sham | 0.069 | 0.091 | -0.13 | 0.27 | | | | | | |
| | | Active | | | | | Sham | 0.068 | 0.129 | -0.22 | 0.36 | 0.608 |
| | DAY 56 | Active | 0.043 | 0.093 | -0.16 | 0.25 | | | | | | |
| | | Sham | 0.019 | 0.093 | -0.19 | 0.23 | | | | | | |

*p<0.05 by analysis of covariance of the change from baseline to days 14, 28, 56 and 84 (no device) in Creyos parameters, with device, visit, treatment by visit interaction, and baseline covariate. Confidence interval (CI), Least-squares (LS) mean.
A greater percentile is in the direction of improved function. Source: Listing 12. 31DEC24, T05_creyos_ge45

Table 5.2

Analysis of Covariance
Creyos Treatment Device Comparison of Change from Baseline by Parameter
Age >= 45 Years (Per-Protocol Population)

| | | Treatment Device Estimate | | | | | Treatment Device Comparison | | | | | |
|---|---|---|---|---|---|---|---|---|---|---|---|---|
| | | | | | 95% CI | | | Difference | | 95% CI | | |
| Parameter | Visit | Treatment Device | LS Mean | Std Error | Lower | Upper | Comparator | LS Mean | Std Error | Lower | Upper | P-Value |
| Spatial Span | DAY 56 | Active | | | | | Sham | 0.024 | 0.132 | -0.27 | 0.32 | 0.859 |
| | DAY 84 | Active | 0.130 | 0.040 | 0.04 | 0.22 | | | | | | |
| | | Sham | 0.119 | 0.042 | 0.02 | 0.21 | | | | | | |
| | | Active | | | | | Sham | 0.011 | 0.058 | -0.12 | 0.14 | 0.849 |
| Digit Span | BASELINE | Active | 0.561 | 0.103 | 0.46 | 0.66 | | | | | | |
| | | Sham | 0.616 | 0.073 | 0.54 | 0.69 | | | | | | |
| | | Active | | | | | Sham | -0.055 | 0.127 | -0.34 | 0.23 | 0.673 |
| | DAY 14 | Active | 0.242 | 0.077 | 0.07 | 0.42 | | | | | | |
| | | Sham | -0.064 | 0.077 | -0.24 | 0.11 | | | | | | |
| | | Active | | | | | Sham | 0.306 | 0.109 | 0.06 | 0.55 | 0.021* |
| | DAY 28 | Active | 0.074 | 0.109 | -0.18 | 0.33 | | | | | | |
| | | Sham | -0.009 | 0.109 | -0.26 | 0.24 | | | | | | |
| | | Active | | | | | Sham | 0.083 | 0.155 | -0.28 | 0.44 | 0.606 |
| | DAY 56 | Active | 0.163 | 0.106 | -0.08 | 0.40 | | | | | | |
| | | Sham | 0.014 | 0.106 | -0.23 | 0.26 | | | | | | |
| | | Active | | | | | Sham | 0.149 | 0.150 | -0.19 | 0.49 | 0.349 |
| | DAY 84 | Active | 0.123 | 0.094 | -0.10 | 0.34 | | | | | | |
| | | Sham | 0.087 | 0.095 | -0.13 | 0.31 | | | | | | |
| | | Active | | | | | Sham | 0.037 | 0.134 | -0.27 | 0.35 | 0.791 |
| Odd One Out | BASELINE | Active | 0.536 | 0.100 | 0.44 | 0.64 | | | | | | |
| | | Sham | 0.655 | 0.069 | 0.59 | 0.72 | | | | | | |
| | | Active | | | | | Sham | -0.119 | 0.122 | -0.39 | 0.15 | 0.351 |
| | DAY 14 | Active | 0.150 | 0.107 | -0.09 | 0.39 | | | | | | |
| | | Sham | -0.127 | 0.106 | -0.37 | 0.11 | | | | | | |
| | | Active | | | | | Sham | 0.277 | 0.153 | -0.06 | 0.62 | 0.100 |
| | DAY 28 | Active | -0.034 | 0.079 | -0.21 | 0.15 | | | | | | |
| | | Sham | -0.222 | 0.078 | -0.40 | -0.04 | | | | | | |
| | | Active | | | | | Sham | 0.189 | 0.113 | -0.07 | 0.45 | 0.133 |
| | DAY 56 | Active | -0.096 | 0.148 | -0.43 | 0.24 | | | | | | |
| | | Sham | -0.080 | 0.148 | -0.41 | 0.25 | | | | | | |
| | | Active | | | | | Sham | -0.016 | 0.211 | -0.49 | 0.45 | 0.941 |
| | DAY 84 | Active | -0.059 | 0.101 | -0.28 | 0.17 | | | | | | |

*p<0.05 by analysis of covariance of the change from baseline to days 14, 28, 56 and 84 (no device) in Creyos parameters, with device, visit, treatment by visit interaction, and baseline covariate. Confidence interval (CI), Least-squares (LS) mean.
A greater percentile is in the direction of improved function. Source: Listing 12. 31DEC24, T05_creyos_ge45

Table 5.2

Analysis of Covariance
Creyos Treatment Device Comparison of Change from Baseline by Parameter
Age >= 45 Years (Per-Protocol Population)

| | | Treatment Device Estimate | | | | | Treatment Device Comparison | | | | | |
|---|---|---|---|---|---|---|---|---|---|---|---|---|
| | | | | | 95% CI | | | Difference | | 95% CI | | |
| Parameter | Visit | Treatment Device | LS Mean | Std Error | Lower | Upper | Comparator | LS Mean | Std Error | Lower | Upper | P-Value |
| Odd One Out | DAY 84 | Sham | 0.010 | 0.107 | -0.23 | 0.25 | | | | | | |
| | | Active | | | | | Sham | -0.069 | 0.149 | -0.40 | 0.26 | 0.653 |
| Double Trouble | BASELINE | Active | 0.359 | 0.147 | 0.21 | 0.51 | | | | | | |
| | | Sham | 0.518 | 0.174 | 0.34 | 0.69 | | | | | | |
| | | Active | | | | | Sham | -0.159 | 0.228 | -0.67 | 0.35 | 0.501 |
| | DAY 14 | Active | 0.140 | 0.082 | -0.04 | 0.32 | | | | | | |
| | | Sham | 0.087 | 0.083 | -0.10 | 0.27 | | | | | | |
| | | Active | | | | | Sham | 0.054 | 0.118 | -0.21 | 0.32 | 0.659 |
| | DAY 28 | Active | 0.159 | 0.090 | -0.04 | 0.36 | | | | | | |
| | | Sham | 0.202 | 0.091 | -0.00 | 0.40 | | | | | | |
| | | Active | | | | | Sham | -0.043 | 0.129 | -0.33 | 0.25 | 0.745 |
| | DAY 56 | Active | 0.114 | 0.097 | -0.10 | 0.33 | | | | | | |
| | | Sham | 0.298 | 0.097 | 0.08 | 0.52 | | | | | | |
| | | Active | | | | | Sham | -0.184 | 0.138 | -0.49 | 0.12 | 0.213 |
| | DAY 84 | Active | 0.357 | 0.104 | 0.12 | 0.59 | | | | | | |
| | | Sham | 0.271 | 0.112 | 0.02 | 0.52 | | | | | | |
| | | Active | | | | | Sham | 0.087 | 0.154 | -0.26 | 0.43 | 0.586 |

*p<0.05 by analysis of covariance of the change from baseline to days 14, 28, 56 and 84 (no device) in Creyos parameters, with device, visit, treatment by visit interaction, and baseline covariate. Confidence interval (CI), Least-squares (LS) mean.
A greater percentile is in the direction of improved function. Source: Listing 12. 31DEC24, T05_creyos_ge45

Table 6

Analysis of Covariance
Perceived Deficit Questionnaire (PDQ-20) Treatment Device Comparison in Change from Baseline to Day 56 (Per-Protocol Population)

| | | | Treatment Device Estimate | | | | Treatment Device Comparison | Difference | | | | |
|---|---|---|---|---|---|---|---|---|---|---|---|---|
| Parameter | Visit | Device | LS Mean | Std Error | 95% CI Lower | 95% CI Upper | vs. Device | LS Mean | Std Error | 95% CI Lower | 95% CI Upper | P-Value |
| 1-Lose train of thought when speaking | BASELINE | Active | 2.49 | 0.168 | 2.32 | 2.66 | | | | | | |
| | | Sham | 2.70 | 0.114 | 2.59 | 2.81 | | | | | | |
| | | Active | | | | | Sham | -0.224 | 0.289 | -0.81 | 0.36 | 0.444 |
| | DAY 56 | Active | -0.52 | 0.175 | -0.87 | -0.16 | | | | | | |
| | | Sham | -0.53 | 0.179 | -0.89 | -0.16 | | | | | | |
| | | Active | | | | | Sham | 0.012 | 0.241 | -0.48 | 0.50 | 0.961 |
| | DAY 84 | Active | -0.50 | 0.190 | -0.88 | -0.11 | | | | | | |
| | | Sham | -0.48 | 0.191 | -0.86 | -0.09 | | | | | | |
| | | Active | | | | | Sham | -0.020 | 0.261 | -0.55 | 0.51 | 0.939 |
| 2-Difficulty remembering names of people | BASELINE | Active | 2.59 | 0.156 | 2.43 | 2.74 | | | | | | |
| | | Sham | 2.90 | 0.151 | 2.75 | 3.05 | | | | | | |
| | | Active | | | | | Sham | -0.281 | 0.311 | -0.91 | 0.35 | 0.373 |
| | DAY 56 | Active | -0.72 | 0.200 | -1.12 | -0.31 | | | | | | |
| | | Sham | -0.65 | 0.204 | -1.07 | -0.24 | | | | | | |
| | | Active | | | | | Sham | -0.062 | 0.276 | -0.62 | 0.50 | 0.824 |
| | DAY 84 | Active | -0.63 | 0.178 | -0.99 | -0.27 | | | | | | |
| | | Sham | -0.35 | 0.179 | -0.72 | 0.01 | | | | | | |
| | | Active | | | | | Sham | -0.276 | 0.244 | -0.77 | 0.22 | 0.264 |
| 3-Forget what you came into the room for | BASELINE | Active | 2.37 | 0.194 | 2.17 | 2.56 | | | | | | |
| | | Sham | 2.70 | 0.114 | 2.59 | 2.81 | | | | | | |
| | | Active | | | | | Sham | -0.319 | 0.321 | -0.97 | 0.33 | 0.326 |
| | DAY 56 | Active | -0.36 | 0.219 | -0.81 | 0.08 | | | | | | |
| | | Sham | -0.47 | 0.222 | -0.92 | -0.02 | | | | | | |
| | | Active | | | | | Sham | 0.105 | 0.301 | -0.51 | 0.71 | 0.731 |
| | DAY 84 | Active | -0.23 | 0.204 | -0.64 | 0.18 | | | | | | |
| | | Sham | -0.32 | 0.204 | -0.73 | 0.09 | | | | | | |
| | | Active | | | | | Sham | 0.087 | 0.278 | -0.48 | 0.65 | 0.756 |

*p<0.05 by analysis of covariance of the change from baseline to day 56 and 84 (no device) in PDQ with device, age >=45 years (yes, no), and baseline covariate. Confidence interval (CI), Least-squares (LS) mean.
Perceived deficit questionnaire consists of 20 questions on 1 (never have deficit) to 5 (almost always have deficit) scale. A negative change is in the direction of improvement. Source: Listing 9. 31DEC24, T06_pdq

Table 6

Analysis of Covariance
Perceived Deficit Questionnaire (PDQ-20) Treatment Device Comparison in Change from Baseline to Day 56 (Per-Protocol Population)

| Parameter | Visit | Device | Treatment Device Estimate: LS Mean | Treatment Device Estimate: Std Error | Treatment Device Estimate: 95% CI Lower | Treatment Device Estimate: 95% CI Upper | Treatment Device Comparison: vs. Device | Treatment Device Comparison: Difference LS Mean | Treatment Device Comparison: Difference Std Error | Treatment Device Comparison: 95% CI Lower | Treatment Device Comparison: 95% CI Upper | P-Value |
|---|---|---|---|---|---|---|---|---|---|---|---|---|
| 4-Trouble getting things organized | BASELINE | Active | 2.39 | 0.209 | 2.18 | 2.60 | | | | | | |
| | | Sham | 2.80 | 0.120 | 2.68 | 2.92 | | | | | | |
| | | Active | | | | | Sham | -0.371 | 0.348 | -1.08 | 0.33 | 0.292 |
| | DAY 56 | Active | -0.27 | 0.145 | -0.56 | 0.02 | | | | | | |
| | | Sham | -0.34 | 0.148 | -0.64 | -0.04 | | | | | | |
| | | Active | | | | | Sham | 0.071 | 0.200 | -0.33 | 0.48 | 0.723 |
| | DAY 84 | Active | -0.07 | 0.165 | -0.40 | 0.27 | | | | | | |
| | | Sham | -0.24 | 0.166 | -0.58 | 0.09 | | | | | | |
| | | Active | | | | | Sham | 0.174 | 0.228 | -0.29 | 0.64 | 0.450 |
| 5-Trouble concentration during conversation | BASELINE | Active | 2.76 | 0.155 | 2.60 | 2.91 | | | | | | |
| | | Sham | 2.40 | 0.093 | 2.31 | 2.49 | | | | | | |
| | | Active | | | | | Sham | 0.362 | 0.258 | -0.16 | 0.88 | 0.169 |
| | DAY 56 | Active | -0.51 | 0.195 | -0.90 | -0.11 | | | | | | |
| | | Sham | -0.61 | 0.196 | -1.00 | -0.21 | | | | | | |
| | | Active | | | | | Sham | 0.099 | 0.267 | -0.44 | 0.64 | 0.713 |
| | DAY 84 | Active | -0.32 | 0.230 | -0.78 | 0.15 | | | | | | |
| | | Sham | -0.56 | 0.229 | -1.02 | -0.09 | | | | | | |
| | | Active | | | | | Sham | 0.238 | 0.316 | -0.40 | 0.88 | 0.457 |
| 6-Forget if you already done something | BASELINE | Active | 2.34 | 0.151 | 2.19 | 2.49 | | | | | | |
| | | Sham | 2.45 | 0.147 | 2.30 | 2.60 | | | | | | |
| | | Active | | | | | Sham | -0.117 | 0.299 | -0.72 | 0.49 | 0.698 |
| | DAY 56 | Active | -0.27 | 0.208 | -0.69 | 0.15 | | | | | | |
| | | Sham | -0.52 | 0.212 | -0.94 | -0.09 | | | | | | |
| | | Active | | | | | Sham | 0.244 | 0.285 | -0.33 | 0.82 | 0.398 |
| | DAY 84 | Active | -0.20 | 0.215 | -0.63 | 0.24 | | | | | | |
| | | Sham | -0.47 | 0.216 | -0.90 | -0.03 | | | | | | |
| | | Active | | | | | Sham | 0.269 | 0.294 | -0.33 | 0.87 | 0.366 |

*p<0.05 by analysis of covariance of the change from baseline to day 56 and 84 (no device) in PDQ with device, age >=45 years (yes, no), and baseline covariate. Confidence interval (CI), Least-squares (LS) mean.
Perceived deficit questionnaire consists of 20 questions on 1 (never have deficit) to 5 (almost always have deficit) scale. A negative change is in the direction of improvement. Source: Listing 9.

31DEC24, T06_pdq

Table 6

Analysis of Covariance
Perceived Deficit Questionnaire (PDQ-20) Treatment Device Comparison in Change from Baseline to Day 56 (Per-Protocol Population)

| | | | Treatment Device Estimate | | | | Treatment Device Comparison | | | | | |
|---|---|---|---|---|---|---|---|---|---|---|---|---|
| | | | | | | | | Difference | | | | |
| | | | | Std | 95% CI | | vs. | LS | Std | 95% CI | | |
| Parameter | Visit | Device | LS Mean | Error | Lower | Upper | Device | Mean | Error | Lower | Upper | P-Value |
| 7-Miss scheduled appointments and meetings | BASELINE | Active | 1.15 | 0.154 | 0.99 | 1.30 | | | | | | |
| | | Sham | 1.45 | 0.172 | 1.28 | 1.62 | | | | | | |
| | | Active | | | | | Sham | -0.260 | 0.333 | -0.93 | 0.41 | 0.440 |
| | DAY 56 | Active | -0.27 | 0.174 | -0.62 | 0.08 | | | | | | |
| | | Sham | -0.36 | 0.176 | -0.71 | -0.00 | | | | | | |
| | | Active | | | | | Sham | 0.087 | 0.238 | -0.40 | 0.57 | 0.718 |
| | DAY 84 | Active | 0.12 | 0.185 | -0.25 | 0.50 | | | | | | |
| | | Sham | -0.21 | 0.184 | -0.58 | 0.16 | | | | | | |
| | | Active | | | | | Sham | 0.331 | 0.251 | -0.18 | 0.84 | 0.196 |
| 8-Difficulty planning what to do during the day | BASELINE | Active | 2.22 | 0.162 | 2.06 | 2.38 | | | | | | |
| | | Sham | 2.25 | 0.112 | 2.14 | 2.36 | | | | | | |
| | | Active | | | | | Sham | -0.012 | 0.281 | -0.58 | 0.56 | 0.966 |
| | DAY 56 | Active | -0.41 | 0.189 | -0.80 | -0.03 | | | | | | |
| | | Sham | -0.39 | 0.192 | -0.78 | 0.00 | | | | | | |
| | | Active | | | | | Sham | -0.026 | 0.258 | -0.55 | 0.50 | 0.921 |
| | DAY 84 | Active | -0.11 | 0.210 | -0.54 | 0.31 | | | | | | |
| | | Sham | -0.49 | 0.210 | -0.91 | -0.06 | | | | | | |
| | | Active | | | | | Sham | 0.373 | 0.288 | -0.21 | 0.96 | 0.203 |
| 9-Trouble concentrating on things like TV or book | BASELINE | Active | 2.85 | 0.199 | 2.65 | 3.05 | | | | | | |
| | | Sham | 2.40 | 0.171 | 2.23 | 2.57 | | | | | | |
| | | Active | | | | | Sham | 0.457 | 0.372 | -0.30 | 1.21 | 0.227 |
| | DAY 56 | Active | -0.62 | 0.239 | -1.11 | -0.14 | | | | | | |
| | | Sham | -0.30 | 0.245 | -0.80 | 0.19 | | | | | | |
| | | Active | | | | | Sham | -0.316 | 0.335 | -1.00 | 0.36 | 0.351 |
| | DAY 84 | Active | -0.25 | 0.185 | -0.63 | 0.13 | | | | | | |
| | | Sham | -0.45 | 0.187 | -0.83 | -0.08 | | | | | | |
| | | Active | | | | | Sham | 0.205 | 0.254 | -0.31 | 0.72 | 0.424 |

*p<0.05 by analysis of covariance of the change from baseline to day 56 and 84 (no device) in PDQ with device, age >=45 years (yes, no), and baseline covariate. Confidence interval (CI), Least-squares (LS) mean.
Perceived deficit questionnaire consists of 20 questions on 1 (never have deficit) to 5 (almost always have deficit) scale. A negative change is in the direction of improvement. Source: Listing 9. 31DEC24, T06_pdq

Table 6

Analysis of Covariance
Perceived Deficit Questionnaire (PDQ-20) Treatment Device Comparison in Change from Baseline to Day 56 (Per-Protocol Population)

| | | | | | | | Treatment Device Comparison | | | | | |
|---|---|---|---|---|---|---|---|---|---|---|---|---|
| | | | Treatment Device Estimate | | | | | Difference | | | | |
| | | | | Std | 95% CI | | vs. | LS | Std | 95% CI | | |
| Parameter | Visit | Device | LS Mean | Error | Lower | Upper | Device | Mean | Error | Lower | Upper | P-Value |
| 10-Forget what you did the night before | BASELINE | Active | 2.05 | 0.200 | 1.85 | 2.25 | | | | | | |
| | | Sham | 1.95 | 0.164 | 1.79 | 2.11 | | | | | | |
| | | Active | | | | | Sham | 0.098 | 0.367 | -0.65 | 0.84 | 0.792 |
| | DAY 56 | Active | -0.27 | 0.195 | -0.66 | 0.13 | | | | | | |
| | | Sham | -0.37 | 0.199 | -0.77 | 0.04 | | | | | | |
| | | Active | | | | | Sham | 0.096 | 0.268 | -0.45 | 0.64 | 0.723 |
| | DAY 84 | Active | -0.20 | 0.186 | -0.58 | 0.18 | | | | | | |
| | | Sham | -0.47 | 0.186 | -0.84 | -0.09 | | | | | | |
| | | Active | | | | | Sham | 0.264 | 0.253 | -0.25 | 0.78 | 0.303 |
| 11-Forget the date unless you looked it up | BASELINE | Active | 2.80 | 0.168 | 2.64 | 2.97 | | | | | | |
| | | Sham | 2.45 | 0.138 | 2.31 | 2.59 | | | | | | |
| | | Active | | | | | Sham | 0.360 | 0.309 | -0.27 | 0.98 | 0.252 |
| | DAY 56 | Active | -0.39 | 0.197 | -0.78 | 0.01 | | | | | | |
| | | Sham | -0.10 | 0.199 | -0.50 | 0.30 | | | | | | |
| | | Active | | | | | Sham | -0.287 | 0.270 | -0.83 | 0.26 | 0.294 |
| | DAY 84 | Active | -0.18 | 0.204 | -0.59 | 0.23 | | | | | | |
| | | Sham | -0.15 | 0.204 | -0.56 | 0.26 | | | | | | |
| | | Active | | | | | Sham | -0.030 | 0.279 | -0.59 | 0.53 | 0.915 |
| 12-Trouble getting started even with things to do | BASELINE | Active | 2.68 | 0.183 | 2.50 | 2.87 | | | | | | |
| | | Sham | 2.60 | 0.138 | 2.46 | 2.74 | | | | | | |
| | | Active | | | | | Sham | 0.114 | 0.328 | -0.55 | 0.78 | 0.730 |
| | DAY 56 | Active | -0.57 | 0.166 | -0.90 | -0.23 | | | | | | |
| | | Sham | -0.48 | 0.170 | -0.83 | -0.14 | | | | | | |
| | | Active | | | | | Sham | -0.083 | 0.228 | -0.55 | 0.38 | 0.718 |
| | DAY 84 | Active | -0.29 | 0.174 | -0.65 | 0.06 | | | | | | |
| | | Sham | -0.43 | 0.175 | -0.79 | -0.08 | | | | | | |
| | | Active | | | | | Sham | 0.139 | 0.238 | -0.34 | 0.62 | 0.564 |
| 13-Find your mind drifting | BASELINE | Active | 2.90 | 0.139 | 2.76 | 3.04 | | | | | | |

*p<0.05 by analysis of covariance of the change from baseline to day 56 and 84 (no device) in PDQ with device, age >=45 years (yes, no), and baseline covariate. Confidence interval (CI), Least-squares (LS) mean.
Perceived deficit questionnaire consists of 20 questions on 1 (never have deficit) to 5 (almost always have deficit) scale. A negative change is in the direction of improvement. Source: Listing 9. 31DEC24, T06_pdq

Table 6

Analysis of Covariance
Perceived Deficit Questionnaire (PDQ-20) Treatment Device Comparison in Change from Baseline to Day 56 (Per-Protocol Population)

| | | Treatment Device Estimate | | | | Treatment Device Comparison Difference | | | | | |
|---|---|---|---|---|---|---|---|---|---|---|---|
| | | | | 95% CI | | | | | 95% CI | | |
| Parameter | Visit | Device | LS Mean | Std Error | Lower | Upper | vs. Device | LS Mean | Std Error | Lower | Upper | P-Value |
| 13-Find your mind drifting | BASELINE | Sham | 3.00 | 0.101 | 2.90 | 3.10 | | | | | | |
| | | Active | | | | | Sham | -0.095 | 0.244 | -0.59 | 0.40 | 0.699 |
| | DAY 56 | Active | -0.33 | 0.196 | -0.73 | 0.07 | | | | | | |
| | | Sham | -0.46 | 0.199 | -0.86 | -0.05 | | | | | | |
| | | Active | | | | | Sham | 0.126 | 0.268 | -0.42 | 0.67 | 0.640 |
| | DAY 84 | Active | -0.14 | 0.195 | -0.53 | 0.26 | | | | | | |
| | | Sham | -0.36 | 0.196 | -0.75 | 0.04 | | | | | | |
| | | Active | | | | | Sham | 0.218 | 0.266 | -0.32 | 0.76 | 0.419 |
| 14-Forget what talked about after call | BASELINE | Active | 2.07 | 0.196 | 1.88 | 2.27 | | | | | | |
| | | Sham | 2.00 | 0.175 | 1.82 | 2.18 | | | | | | |
| | | Active | | | | | Sham | 0.095 | 0.374 | -0.66 | 0.85 | 0.800 |
| | DAY 56 | Active | -0.57 | 0.212 | -1.00 | -0.14 | | | | | | |
| | | Sham | -0.50 | 0.216 | -0.93 | -0.06 | | | | | | |
| | | Active | | | | | Sham | -0.073 | 0.294 | -0.67 | 0.52 | 0.806 |
| | DAY 84 | Active | -0.32 | 0.175 | -0.68 | 0.03 | | | | | | |
| | | Sham | -0.55 | 0.175 | -0.90 | -0.19 | | | | | | |
| | | Active | | | | | Sham | 0.225 | 0.238 | -0.26 | 0.71 | 0.350 |
| 15-Forget to do things like turn off stove or lock door | BASELINE | Active | 1.56 | 0.156 | 1.40 | 1.72 | | | | | | |
| | | Sham | 1.60 | 0.163 | 1.44 | 1.76 | | | | | | |
| | | Active | | | | | Sham | 0.019 | 0.331 | -0.65 | 0.69 | 0.954 |
| | DAY 56 | Active | -0.54 | 0.144 | -0.83 | -0.25 | | | | | | |
| | | Sham | -0.49 | 0.146 | -0.78 | -0.19 | | | | | | |
| | | Active | | | | | Sham | -0.055 | 0.197 | -0.45 | 0.34 | 0.780 |
| | DAY 84 | Active | -0.24 | 0.161 | -0.57 | 0.08 | | | | | | |
| | | Sham | -0.44 | 0.161 | -0.76 | -0.11 | | | | | | |
| | | Active | | | | | Sham | 0.196 | 0.221 | -0.25 | 0.64 | 0.380 |
| 16-Feel like your mind went totally blank | BASELINE | Active | 2.61 | 0.160 | 2.45 | 2.77 | | | | | | |
| | | Sham | 2.10 | 0.133 | 1.97 | 2.23 | | | | | | |

*p<0.05 by analysis of covariance of the change from baseline to day 56 and 84 (no device) in PDQ with device, age >=45 years (yes, no), and baseline covariate. Confidence interval (CI), Least-squares (LS) mean.
Perceived deficit questionnaire consists of 20 questions on 1 (never have deficit) to 5 (almost always have deficit) scale. A negative change is in the direction of improvement. Source: Listing 9. 31DEC24, T06_pdq

Table 6

Analysis of Covariance
Perceived Deficit Questionnaire (PDQ-20) Treatment Device Comparison in Change from Baseline to Day 56 (Per-Protocol Population)

| | | | Treatment Device Estimate | | | | Treatment Device Comparison | | | | | |
|---|---|---|---|---|---|---|---|---|---|---|---|---|
| | | | | | 95% CI | | | Difference | | 95% CI | | |
| Parameter | Visit | Device | LS Mean | Std Error | Lower | Upper | vs. Device | LS Mean | Std Error | Lower | Upper | P-Value |
| 16-Feel like your mind went totally blank | BASELINE | Active | | | | | Sham | 0.519 | 0.295 | -0.08 | 1.12 | 0.086 |
| | DAY 56 | Active | -0.39 | 0.212 | -0.82 | 0.04 | | | | | | |
| | | Sham | -0.55 | 0.216 | -0.98 | -0.11 | | | | | | |
| | | Active | | | | | Sham | 0.156 | 0.295 | -0.44 | 0.75 | 0.600 |
| | DAY 84 | Active | -0.45 | 0.224 | -0.91 | -0.00 | | | | | | |
| | | Sham | -0.50 | 0.225 | -0.95 | -0.04 | | | | | | |
| | | Active | | | | | Sham | 0.040 | 0.311 | -0.59 | 0.67 | 0.898 |
| 17-Trouble holding phone numbers even for a few seconds | BASELINE | Active | 2.59 | 0.185 | 2.40 | 2.77 | | | | | | |
| | | Sham | 2.80 | 0.165 | 2.64 | 2.96 | | | | | | |
| | | Active | | | | | Sham | -0.181 | 0.354 | -0.90 | 0.54 | 0.612 |
| | DAY 56 | Active | -0.51 | 0.231 | -0.98 | -0.05 | | | | | | |
| | | Sham | -0.25 | 0.236 | -0.73 | 0.23 | | | | | | |
| | | Active | | | | | Sham | -0.261 | 0.318 | -0.90 | 0.38 | 0.417 |
| | DAY 84 | Active | -0.47 | 0.222 | -0.91 | -0.02 | | | | | | |
| | | Sham | -0.30 | 0.223 | -0.75 | 0.15 | | | | | | |
| | | Active | | | | | Sham | -0.164 | 0.303 | -0.78 | 0.45 | 0.592 |
| 18-Forgot what you did last week | BASELINE | Active | 2.22 | 0.196 | 2.02 | 2.42 | | | | | | |
| | | Sham | 2.25 | 0.174 | 2.08 | 2.42 | | | | | | |
| | | Active | | | | | Sham | -0.012 | 0.373 | -0.77 | 0.74 | 0.975 |
| | DAY 56 | Active | -0.25 | 0.200 | -0.66 | 0.15 | | | | | | |
| | | Sham | -0.37 | 0.203 | -0.78 | 0.04 | | | | | | |
| | | Active | | | | | Sham | 0.118 | 0.274 | -0.44 | 0.67 | 0.669 |
| | DAY 84 | Active | -0.22 | 0.226 | -0.68 | 0.23 | | | | | | |
| | | Sham | -0.47 | 0.226 | -0.93 | -0.02 | | | | | | |
| | | Active | | | | | Sham | 0.249 | 0.310 | -0.38 | 0.88 | 0.426 |
| 19-Forget to take your medication | BASELINE | Active | 1.71 | 0.157 | 1.55 | 1.86 | | | | | | |
| | | Sham | 1.45 | 0.129 | 1.32 | 1.58 | | | | | | |

*p<0.05 by analysis of covariance of the change from baseline to day 56 and 84 (no device) in PDQ with device, age >=45 years (yes, no), and baseline covariate. Confidence interval (CI), Least-squares (LS) mean.
Perceived deficit questionnaire consists of 20 questions on 1 (never have deficit) to 5 (almost always have deficit) scale. A negative change is in the direction of improvement. Source: Listing 9. 31DEC24, T06_pdq

Table 6

Analysis of Covariance
Perceived Deficit Questionnaire (PDQ-20) Treatment Device Comparison in Change from Baseline to Day 56 (Per-Protocol Population)

| | | | Treatment Device Estimate | | | | Treatment Device Comparison | | | | | |
|---|---|---|---|---|---|---|---|---|---|---|---|---|
| | | | | | | | | Difference | | | | |
| | | | | Std | 95% CI | | vs. | LS | Std | 95% CI | | |
| Parameter | Visit | Device | LS Mean | Error | Lower | Upper | Device | Mean | Error | Lower | Upper | P-Value |
| 19-Forget to take your medication | BASELINE | Active | | | | | Sham | 0.264 | 0.288 | -0.32 | 0.85 | 0.365 |
| | DAY 56 | Active | -0.22 | 0.200 | -0.63 | 0.18 | | | | | | |
| | | Sham | -0.41 | 0.205 | -0.83 | 0.00 | | | | | | |
| | | Active | | | | | Sham | 0.190 | 0.275 | -0.37 | 0.75 | 0.495 |
| | DAY 84 | Active | -0.05 | 0.206 | -0.47 | 0.37 | | | | | | |
| | | Sham | -0.46 | 0.209 | -0.88 | -0.04 | | | | | | |
| | | Active | | | | | Sham | 0.411 | 0.283 | -0.16 | 0.98 | 0.155 |
| 20-Have trouble making decisions | BASELINE | Active | 2.51 | 0.182 | 2.33 | 2.69 | | | | | | |
| | | Sham | 2.45 | 0.172 | 2.28 | 2.62 | | | | | | |
| | | Active | | | | | Sham | 0.074 | 0.354 | -0.64 | 0.79 | 0.836 |
| | DAY 56 | Active | -0.50 | 0.173 | -0.85 | -0.15 | | | | | | |
| | | Sham | -0.35 | 0.176 | -0.71 | 0.01 | | | | | | |
| | | Active | | | | | Sham | -0.146 | 0.236 | -0.62 | 0.33 | 0.541 |
| | DAY 84 | Active | -0.41 | 0.200 | -0.81 | -0.01 | | | | | | |
| | | Sham | -0.55 | 0.201 | -0.96 | -0.14 | | | | | | |
| | | Active | | | | | Sham | 0.140 | 0.274 | -0.42 | 0.69 | 0.613 |
| Mean PDQ Questions 1-20 | BASELINE | Active | 2.34 | 0.128 | 2.21 | 2.47 | | | | | | |
| | | Sham | 2.34 | 0.081 | 2.25 | 2.42 | | | | | | |
| | | Active | | | | | Sham | 0.025 | 0.217 | -0.41 | 0.46 | 0.911 |
| | DAY 56 | Active | -0.42 | 0.136 | -0.69 | -0.14 | | | | | | |
| | | Sham | -0.43 | 0.139 | -0.71 | -0.15 | | | | | | |
| | | Active | | | | | Sham | 0.010 | 0.187 | -0.37 | 0.39 | 0.958 |
| | DAY 84 | Active | -0.24 | 0.131 | -0.51 | 0.02 | | | | | | |
| | | Sham | -0.42 | 0.132 | -0.68 | -0.15 | | | | | | |
| | | Active | | | | | Sham | 0.171 | 0.177 | -0.19 | 0.53 | 0.340 |

*p<0.05 by analysis of covariance of the change from baseline to day 56 and 84 (no device) in PDQ with device, age >=45 years (yes, no), and baseline covariate. Confidence interval (CI), Least-squares (LS) mean.
Perceived deficit questionnaire consists of 20 questions on 1 (never have deficit) to 5 (almost always have deficit) scale. A negative change is in the direction of improvement. Source: Listing 9. 31DEC24, T06_pdq

Table 6.1

Analysis of Covariance
Perceived Deficit Questionnaire (PDQ-20) Treatment Device Comparison in Change from Baseline to Day 56
Age < 45 Years (Per-Protocol Population)

| | | | Treatment Device Estimate | | | | Treatment Device Comparison | | | | | |
|---|---|---|---|---|---|---|---|---|---|---|---|---|
| | | | | | | | | Difference | | | | |
| Parameter | Visit | Device | LS Mean | Std Error | 95% CI Lower | 95% CI Upper | vs. Device | LS Mean | Std Error | 95% CI Lower | 95% CI Upper | P-Value |
| 1-Lose train of thought when speaking | BASELINE | Active | 2.48 | 0.208 | 2.27 | 2.69 | | | | | | |
| | | Sham | 2.64 | 0.156 | 2.49 | 2.80 | | | | | | |
| | | Active | | | | | Sham | -0.176 | 0.371 | -0.94 | 0.59 | 0.639 |
| | DAY 56 | Active | -0.72 | 0.195 | -1.12 | -0.32 | | | | | | |
| | | Sham | -0.60 | 0.202 | -1.01 | -0.18 | | | | | | |
| | | Active | | | | | Sham | -0.117 | 0.281 | -0.70 | 0.46 | 0.680 |
| | DAY 84 | Active | -0.67 | 0.247 | -1.18 | -0.16 | | | | | | |
| | | Sham | -0.60 | 0.250 | -1.12 | -0.08 | | | | | | |
| | | Active | | | | | Sham | -0.070 | 0.352 | -0.80 | 0.66 | 0.843 |
| 2-Difficulty remembering names of people | BASELINE | Active | 2.69 | 0.211 | 2.48 | 2.90 | | | | | | |
| | | Sham | 2.79 | 0.195 | 2.59 | 2.98 | | | | | | |
| | | Active | | | | | Sham | -0.052 | 0.413 | -0.90 | 0.79 | 0.900 |
| | DAY 56 | Active | -1.14 | 0.174 | -1.49 | -0.78 | | | | | | |
| | | Sham | -0.62 | 0.180 | -0.99 | -0.25 | | | | | | |
| | | Active | | | | | Sham | -0.518 | 0.250 | -1.03 | -0.00 | 0.048* |
| | DAY 84 | Active | -0.87 | 0.214 | -1.30 | -0.43 | | | | | | |
| | | Sham | -0.33 | 0.216 | -0.78 | 0.11 | | | | | | |
| | | Active | | | | | Sham | -0.533 | 0.304 | -1.16 | 0.09 | 0.091 |
| 3-Forget what you came into the room for | BASELINE | Active | 2.38 | 0.250 | 2.13 | 2.63 | | | | | | |
| | | Sham | 2.79 | 0.149 | 2.64 | 2.93 | | | | | | |
| | | Active | | | | | Sham | -0.386 | 0.417 | -1.24 | 0.47 | 0.363 |
| | DAY 56 | Active | -0.68 | 0.271 | -1.23 | -0.12 | | | | | | |
| | | Sham | -0.62 | 0.281 | -1.20 | -0.04 | | | | | | |
| | | Active | | | | | Sham | -0.056 | 0.392 | -0.86 | 0.75 | 0.887 |
| | DAY 84 | Active | -0.49 | 0.222 | -0.95 | -0.04 | | | | | | |
| | | Sham | -0.41 | 0.225 | -0.87 | 0.06 | | | | | | |
| | | Active | | | | | Sham | -0.088 | 0.319 | -0.74 | 0.57 | 0.785 |

*p<0.05 by analysis of covariance of the change from baseline to day 56 and 84 (no device) with device, and baseline covariate.
Confidence interval (CI), Least-squares (LS) mean.
Perceived deficit questionnaire consists of 20 questions on 1 (never have deficit) to 5 (almost always have deficit) scale. A negative change is in the direction of improvement. Source: Listing 9.

Table 6.1

Analysis of Covariance
Perceived Deficit Questionnaire (PDQ-20) Treatment Device Comparison in Change from Baseline to Day 56
Age < 45 Years (Per-Protocol Population)

| | | | Treatment Device Estimate | | | | Treatment Device Comparison | | | | | |
|---|---|---|---|---|---|---|---|---|---|---|---|---|
| | | | | | | | | Difference | | | | |
| | | | | Std | 95% CI | | vs. | LS | Std | 95% CI | | |
| Parameter | Visit | Device | LS Mean | Error | Lower | Upper | Device | Mean | Error | Lower | Upper | P-Value |
| 4-Trouble getting things organized | BASELINE | Active | 2.34 | 0.273 | 2.07 | 2.62 | | | | | | |
| | | Sham | 2.79 | 0.149 | 2.64 | 2.93 | | | | | | |
| | | Active | | | | | Sham | -0.386 | 0.452 | -1.31 | 0.54 | 0.401 |
| | DAY 56 | Active | -0.27 | 0.179 | -0.64 | 0.10 | | | | | | |
| | | Sham | -0.40 | 0.186 | -0.78 | -0.02 | | | | | | |
| | | Active | | | | | Sham | 0.128 | 0.259 | -0.40 | 0.66 | 0.624 |
| | DAY 84 | Active | -0.20 | 0.187 | -0.58 | 0.18 | | | | | | |
| | | Sham | -0.33 | 0.189 | -0.72 | 0.06 | | | | | | |
| | | Active | | | | | Sham | 0.131 | 0.267 | -0.42 | 0.68 | 0.629 |
| 5-Trouble concentration during conversation | BASELINE | Active | 2.72 | 0.192 | 2.53 | 2.92 | | | | | | |
| | | Sham | 2.29 | 0.113 | 2.17 | 2.40 | | | | | | |
| | | Active | | | | | Sham | 0.448 | 0.318 | -0.21 | 1.10 | 0.171 |
| | DAY 56 | Active | -0.55 | 0.225 | -1.01 | -0.08 | | | | | | |
| | | Sham | -0.62 | 0.233 | -1.10 | -0.14 | | | | | | |
| | | Active | | | | | Sham | 0.071 | 0.330 | -0.61 | 0.75 | 0.832 |
| | DAY 84 | Active | -0.35 | 0.290 | -0.94 | 0.25 | | | | | | |
| | | Sham | -0.62 | 0.293 | -1.22 | -0.02 | | | | | | |
| | | Active | | | | | Sham | 0.272 | 0.417 | -0.58 | 1.13 | 0.520 |
| 6-Forget if you already done something | BASELINE | Active | 2.34 | 0.181 | 2.16 | 2.53 | | | | | | |
| | | Sham | 2.36 | 0.187 | 2.17 | 2.54 | | | | | | |
| | | Active | | | | | Sham | -0.024 | 0.368 | -0.78 | 0.73 | 0.949 |
| | DAY 56 | Active | -0.34 | 0.239 | -0.83 | 0.15 | | | | | | |
| | | Sham | -0.35 | 0.248 | -0.86 | 0.16 | | | | | | |
| | | Active | | | | | Sham | 0.011 | 0.345 | -0.70 | 0.72 | 0.975 |
| | DAY 84 | Active | -0.31 | 0.240 | -0.81 | 0.18 | | | | | | |
| | | Sham | -0.35 | 0.242 | -0.85 | 0.14 | | | | | | |
| | | Active | | | | | Sham | 0.042 | 0.341 | -0.66 | 0.74 | 0.904 |

*p<0.05 by analysis of covariance of the change from baseline to day 56 and 84 (no device) with device, and baseline covariate.
Confidence interval (CI), Least-squares (LS) mean.
Perceived deficit questionnaire consists of 20 questions on 1 (never have deficit) to 5 (almost always have deficit) scale. A negative change is in the direction of improvement. Source: Listing 9. 31DEC24, T06_pdq_lt45

Table 6.1

Analysis of Covariance
Perceived Deficit Questionnaire (PDQ-20) Treatment Device Comparison in Change from Baseline to Day 56
Age < 45 Years (Per-Protocol Population)

| | | | Treatment Device Estimate | | | | Treatment Device Comparison | | | | | |
|---|---|---|---|---|---|---|---|---|---|---|---|---|
| | | | | | | | | Difference | | | | |
| | | | | Std | 95% CI | | vs. | LS | Std | 95% CI | | |
| Parameter | Visit | Device | LS Mean | Error | Lower | Upper | Device | Mean | Error | Lower | Upper | P-Value |
| 7-Miss scheduled appointments and meetings | BASELINE | Active | 1.28 | 0.198 | 1.08 | 1.47 | | | | | | |
| | | Sham | 1.57 | 0.227 | 1.34 | 1.80 | | | | | | |
| | | Active | | | | | Sham | -0.238 | 0.434 | -1.13 | 0.65 | 0.587 |
| | DAY 56 | Active | -0.57 | 0.174 | -0.93 | -0.22 | | | | | | |
| | | Sham | -0.57 | 0.181 | -0.94 | -0.20 | | | | | | |
| | | Active | | | | | Sham | -0.002 | 0.252 | -0.52 | 0.51 | 0.994 |
| | DAY 84 | Active | -0.23 | 0.165 | -0.57 | 0.11 | | | | | | |
| | | Sham | -0.22 | 0.165 | -0.56 | 0.12 | | | | | | |
| | | Active | | | | | Sham | -0.015 | 0.235 | -0.50 | 0.47 | 0.949 |
| 8-Difficulty planning what to do during the day | BASELINE | Active | 2.31 | 0.205 | 2.11 | 2.52 | | | | | | |
| | | Sham | 2.43 | 0.140 | 2.29 | 2.57 | | | | | | |
| | | Active | | | | | Sham | -0.095 | 0.356 | -0.83 | 0.63 | 0.791 |
| | DAY 56 | Active | -0.68 | 0.240 | -1.17 | -0.19 | | | | | | |
| | | Sham | -0.55 | 0.248 | -1.06 | -0.04 | | | | | | |
| | | Active | | | | | Sham | -0.127 | 0.345 | -0.84 | 0.58 | 0.715 |
| | DAY 84 | Active | -0.40 | 0.212 | -0.84 | 0.03 | | | | | | |
| | | Sham | -0.48 | 0.213 | -0.92 | -0.04 | | | | | | |
| | | Active | | | | | Sham | 0.076 | 0.300 | -0.54 | 0.70 | 0.801 |
| 9-Trouble concentrating on things like TV or book | BASELINE | Active | 3.00 | 0.199 | 2.80 | 3.20 | | | | | | |
| | | Sham | 2.36 | 0.237 | 2.12 | 2.59 | | | | | | |
| | | Active | | | | | Sham | 0.643 | 0.436 | -0.25 | 1.54 | 0.152 |
| | DAY 56 | Active | -1.05 | 0.295 | -1.66 | -0.45 | | | | | | |
| | | Sham | -0.51 | 0.305 | -1.14 | 0.11 | | | | | | |
| | | Active | | | | | Sham | -0.540 | 0.428 | -1.42 | 0.34 | 0.217 |
| | DAY 84 | Active | -0.61 | 0.201 | -1.02 | -0.20 | | | | | | |
| | | Sham | -0.37 | 0.202 | -0.79 | 0.05 | | | | | | |
| | | Active | | | | | Sham | -0.239 | 0.291 | -0.84 | 0.36 | 0.418 |

*p<0.05 by analysis of covariance of the change from baseline to day 56 and 84 (no device) with device, and baseline covariate.
Confidence interval (CI), Least-squares (LS) mean.
Perceived deficit questionnaire consists of 20 questions on 1 (never have deficit) to 5 (almost always have deficit) scale. A negative change is in the direction of improvement. Source: Listing 9. 31DEC24, T06_pdq_lt45

Table 6.1

Analysis of Covariance
Perceived Deficit Questionnaire (PDQ-20) Treatment Device Comparison in Change from Baseline to Day 56
Age < 45 Years (Per-Protocol Population)

| | | | Treatment Device Estimate | | | | Treatment Device Comparison | | | | | |
|---|---|---|---|---|---|---|---|---|---|---|---|---|
| | | | | | | | Difference | | | | | |
| | | | | Std | 95% CI | | vs. | LS | Std | 95% CI | | |
| Parameter | Visit | Device | LS Mean | Error | Lower | Upper | Device | Mean | Error | Lower | Upper | P-Value |
| 10-Forget what you did the night before | BASELINE | Active | 2.07 | 0.238 | 1.83 | 2.31 | | | | | | |
| | | Sham | 1.86 | 0.204 | 1.65 | 2.06 | | | | | | |
| | | Active | | | | | Sham | 0.210 | 0.445 | -0.70 | 1.12 | 0.641 |
| | DAY 56 | Active | -0.57 | 0.230 | -1.05 | -0.10 | | | | | | |
| | | Sham | -0.39 | 0.238 | -0.87 | 0.10 | | | | | | |
| | | Active | | | | | Sham | -0.188 | 0.331 | -0.87 | 0.49 | 0.576 |
| | DAY 84 | Active | -0.40 | 0.237 | -0.89 | 0.08 | | | | | | |
| | | Sham | -0.39 | 0.240 | -0.88 | 0.11 | | | | | | |
| | | Active | | | | | Sham | -0.018 | 0.338 | -0.71 | 0.68 | 0.957 |
| 11-Forget the date unless you looked it up | BASELINE | Active | 2.66 | 0.206 | 2.45 | 2.86 | | | | | | |
| | | Sham | 2.50 | 0.189 | 2.31 | 2.69 | | | | | | |
| | | Active | | | | | Sham | 0.167 | 0.397 | -0.65 | 0.98 | 0.678 |
| | DAY 56 | Active | -0.56 | 0.244 | -1.06 | -0.06 | | | | | | |
| | | Sham | -0.04 | 0.253 | -0.56 | 0.48 | | | | | | |
| | | Active | | | | | Sham | -0.522 | 0.352 | -1.24 | 0.20 | 0.150 |
| | DAY 84 | Active | -0.34 | 0.236 | -0.82 | 0.15 | | | | | | |
| | | Sham | 0.03 | 0.239 | -0.46 | 0.53 | | | | | | |
| | | Active | | | | | Sham | -0.372 | 0.337 | -1.06 | 0.32 | 0.279 |
| 12-Trouble getting started even with things to do | BASELINE | Active | 2.76 | 0.231 | 2.53 | 2.99 | | | | | | |
| | | Sham | 2.86 | 0.160 | 2.70 | 3.02 | | | | | | |
| | | Active | | | | | Sham | -0.057 | 0.405 | -0.89 | 0.77 | 0.889 |
| | DAY 56 | Active | -0.60 | 0.196 | -1.01 | -0.20 | | | | | | |
| | | Sham | -0.63 | 0.203 | -1.05 | -0.21 | | | | | | |
| | | Active | | | | | Sham | 0.027 | 0.282 | -0.55 | 0.61 | 0.926 |
| | DAY 84 | Active | -0.49 | 0.207 | -0.91 | -0.06 | | | | | | |
| | | Sham | -0.56 | 0.209 | -0.99 | -0.13 | | | | | | |
| | | Active | | | | | Sham | 0.072 | 0.295 | -0.53 | 0.68 | 0.810 |

*p<0.05 by analysis of covariance of the change from baseline to day 56 and 84 (no device) with device, and baseline covariate.
Confidence interval (CI), Least-squares (LS) mean.
Perceived deficit questionnaire consists of 20 questions on 1 (never have deficit) to 5 (almost always have deficit) scale. A negative change is in the direction of improvement. Source: Listing 9. 31DEC24, T06_pdq_lt45

Table 6.1

Analysis of Covariance
Perceived Deficit Questionnaire (PDQ-20) Treatment Device Comparison in Change from Baseline to Day 56
Age < 45 Years (Per-Protocol Population)

| | | | Treatment Device Estimate | | | | Treatment Device Comparison Difference | | | | | |
|---|---|---|---|---|---|---|---|---|---|---|---|---|
| | | | | Std | 95% CI | | vs. | LS | Std | 95% CI | | |
| Parameter | Visit | Device | LS Mean | Error | Lower | Upper | Device | Mean | Error | Lower | Upper | P-Value |
| 13-Find your mind drifting | BASELINE | Active | 2.86 | 0.170 | 2.69 | 3.03 | | | | | | |
| | | Sham | 3.07 | 0.114 | 2.96 | 3.19 | | | | | | |
| | | Active | | | | | Sham | -0.205 | 0.292 | -0.80 | 0.39 | 0.489 |
| | DAY 56 | Active | -0.39 | 0.236 | -0.88 | 0.09 | | | | | | |
| | | Sham | -0.58 | 0.245 | -1.08 | -0.08 | | | | | | |
| | | Active | | | | | Sham | 0.187 | 0.341 | -0.51 | 0.89 | 0.588 |
| | DAY 84 | Active | -0.26 | 0.248 | -0.77 | 0.25 | | | | | | |
| | | Sham | -0.29 | 0.251 | -0.81 | 0.22 | | | | | | |
| | | Active | | | | | Sham | 0.034 | 0.354 | -0.69 | 0.76 | 0.925 |
| 14-Forget what talked about after call | BASELINE | Active | 2.03 | 0.255 | 1.78 | 2.29 | | | | | | |
| | | Sham | 1.93 | 0.235 | 1.69 | 2.16 | | | | | | |
| | | Active | | | | | Sham | 0.138 | 0.495 | -0.88 | 1.15 | 0.782 |
| | DAY 56 | Active | -0.69 | 0.232 | -1.17 | -0.22 | | | | | | |
| | | Sham | -0.53 | 0.240 | -1.02 | -0.03 | | | | | | |
| | | Active | | | | | Sham | -0.169 | 0.335 | -0.86 | 0.52 | 0.618 |
| | DAY 84 | Active | -0.42 | 0.213 | -0.86 | 0.02 | | | | | | |
| | | Sham | -0.45 | 0.214 | -0.89 | -0.01 | | | | | | |
| | | Active | | | | | Sham | 0.037 | 0.302 | -0.58 | 0.66 | 0.902 |
| 15-Forget to do things like turn off stove or lock door | BASELINE | Active | 1.72 | 0.198 | 1.53 | 1.92 | | | | | | |
| | | Sham | 1.64 | 0.225 | 1.42 | 1.87 | | | | | | |
| | | Active | | | | | Sham | 0.157 | 0.439 | -0.74 | 1.06 | 0.723 |
| | DAY 56 | Active | -0.81 | 0.169 | -1.15 | -0.46 | | | | | | |
| | | Sham | -0.59 | 0.174 | -0.95 | -0.23 | | | | | | |
| | | Active | | | | | Sham | -0.214 | 0.243 | -0.71 | 0.29 | 0.387 |
| | DAY 84 | Active | -0.45 | 0.195 | -0.85 | -0.05 | | | | | | |
| | | Sham | -0.59 | 0.197 | -1.00 | -0.19 | | | | | | |
| | | Active | | | | | Sham | 0.143 | 0.277 | -0.43 | 0.71 | 0.610 |

*p<0.05 by analysis of covariance of the change from baseline to day 56 and 84 (no device) with device, and baseline covariate.
Confidence interval (CI), Least-squares (LS) mean.
Perceived deficit questionnaire consists of 20 questions on 1 (never have deficit) to 5 (almost always have deficit) scale. A negative change is in the direction of improvement. Source: Listing 9. 31DEC24, T06_pdq_lt45

Table 6.1

Analysis of Covariance
Perceived Deficit Questionnaire (PDQ-20) Treatment Device Comparison in Change from Baseline to Day 56
Age < 45 Years (Per-Protocol Population)

| | | | Treatment Device Estimate | | | | Treatment Device Comparison | | | | | |
|---|---|---|---|---|---|---|---|---|---|---|---|---|
| | | | | Std | 95% CI | | vs. | Difference | | | | |
| Parameter | Visit | Device | LS Mean | Error | Lower | Upper | Device | LS Mean | Std Error | 95% CI Lower | Upper | P-Value |
| 16-Feel like your mind went totally blank | BASELINE | Active | 2.59 | 0.153 | 2.43 | 2.74 | | | | | | |
| | | Sham | 2.14 | 0.176 | 1.97 | 2.32 | | | | | | |
| | | Active | | | | | Sham | 0.457 | 0.330 | -0.22 | 1.13 | 0.178 |
| | DAY 56 | Active | -0.57 | 0.257 | -1.10 | -0.04 | | | | | | |
| | | Sham | -0.74 | 0.265 | -1.28 | -0.19 | | | | | | |
| | | Active | | | | | Sham | 0.164 | 0.375 | -0.61 | 0.93 | 0.666 |
| | DAY 84 | Active | -0.38 | 0.277 | -0.94 | 0.19 | | | | | | |
| | | Sham | -0.59 | 0.281 | -1.17 | -0.02 | | | | | | |
| | | Active | | | | | Sham | 0.218 | 0.400 | -0.60 | 1.04 | 0.591 |
| 17-Trouble holding phone numbers even for a few seconds | BASELINE | Active | 2.62 | 0.255 | 2.37 | 2.88 | | | | | | |
| | | Sham | 2.71 | 0.223 | 2.49 | 2.94 | | | | | | |
| | | Active | | | | | Sham | -0.048 | 0.486 | -1.04 | 0.95 | 0.923 |
| | DAY 56 | Active | -0.53 | 0.285 | -1.12 | 0.05 | | | | | | |
| | | Sham | -0.19 | 0.295 | -0.80 | 0.42 | | | | | | |
| | | Active | | | | | Sham | -0.343 | 0.410 | -1.19 | 0.50 | 0.410 |
| | DAY 84 | Active | -0.61 | 0.224 | -1.07 | -0.15 | | | | | | |
| | | Sham | -0.12 | 0.226 | -0.58 | 0.35 | | | | | | |
| | | Active | | | | | Sham | -0.494 | 0.319 | -1.15 | 0.16 | 0.133 |
| 18-Forgot what you did last week | BASELINE | Active | 2.17 | 0.253 | 1.92 | 2.43 | | | | | | |
| | | Sham | 2.29 | 0.223 | 2.06 | 2.51 | | | | | | |
| | | Active | | | | | Sham | -0.086 | 0.481 | -1.07 | 0.90 | 0.860 |
| | DAY 56 | Active | -0.48 | 0.236 | -0.96 | 0.01 | | | | | | |
| | | Sham | -0.55 | 0.244 | -1.05 | -0.05 | | | | | | |
| | | Active | | | | | Sham | 0.075 | 0.340 | -0.62 | 0.77 | 0.827 |
| | DAY 84 | Active | -0.51 | 0.266 | -1.06 | 0.04 | | | | | | |
| | | Sham | -0.48 | 0.266 | -1.03 | 0.07 | | | | | | |
| | | Active | | | | | Sham | -0.031 | 0.377 | -0.81 | 0.74 | 0.934 |

*p<0.05 by analysis of covariance of the change from baseline to day 56 and 84 (no device) with device, and baseline covariate.
Confidence interval (CI), Least-squares (LS) mean.
Perceived deficit questionnaire consists of 20 questions on 1 (never have deficit) to 5 (almost always have deficit) scale. A negative change is in the direction of improvement. Source: Listing 9. 31DEC24, T06_pdq_lt45

Table 6.1

Analysis of Covariance
Perceived Deficit Questionnaire (PDQ-20) Treatment Device Comparison in Change from Baseline to Day 56
Age < 45 Years (Per-Protocol Population)

| | | | Treatment Device Estimate | | | | Treatment Device Comparison | | | | | |
|---|---|---|---|---|---|---|---|---|---|---|---|---|
| | | | | | | | | Difference | | | | |
| | | | | Std | 95% CI | | vs. | LS | Std | 95% CI | | |
| Parameter | Visit | Device | LS Mean | Error | Lower | Upper | Device | Mean | Error | Lower | Upper | P-Value |
| 19-Forget to take your medication | BASELINE | Active | 1.86 | 0.197 | 1.67 | 2.06 | | | | | | |
| | | Sham | 1.50 | 0.174 | 1.33 | 1.67 | | | | | | |
| | | Active | | | | | Sham | 0.367 | 0.373 | -0.40 | 1.13 | 0.335 |
| | DAY 56 | Active | -0.31 | 0.246 | -0.81 | 0.20 | | | | | | |
| | | Sham | -0.38 | 0.254 | -0.90 | 0.14 | | | | | | |
| | | Active | | | | | Sham | 0.076 | 0.356 | -0.65 | 0.81 | 0.831 |
| | DAY 84 | Active | -0.36 | 0.229 | -0.83 | 0.11 | | | | | | |
| | | Sham | -0.31 | 0.231 | -0.79 | 0.17 | | | | | | |
| | | Active | | | | | Sham | -0.048 | 0.328 | -0.72 | 0.63 | 0.885 |
| 20-Have trouble making decisions | BASELINE | Active | 2.45 | 0.196 | 2.25 | 2.64 | | | | | | |
| | | Sham | 2.57 | 0.238 | 2.33 | 2.81 | | | | | | |
| | | Active | | | | | Sham | -0.105 | 0.436 | -1.00 | 0.79 | 0.812 |
| | DAY 56 | Active | -0.68 | 0.196 | -1.08 | -0.27 | | | | | | |
| | | Sham | -0.41 | 0.203 | -0.83 | 0.00 | | | | | | |
| | | Active | | | | | Sham | -0.263 | 0.283 | -0.84 | 0.32 | 0.361 |
| | DAY 84 | Active | -0.70 | 0.239 | -1.19 | -0.21 | | | | | | |
| | | Sham | -0.70 | 0.241 | -1.20 | -0.20 | | | | | | |
| | | Active | | | | | Sham | 0.002 | 0.340 | -0.70 | 0.70 | 0.994 |
| Mean PDQ Questions 1-20 | BASELINE | Active | 2.37 | 0.162 | 2.21 | 2.53 | | | | | | |
| | | Sham | 2.35 | 0.107 | 2.25 | 2.46 | | | | | | |
| | | Active | | | | | Sham | 0.036 | 0.279 | -0.54 | 0.61 | 0.897 |
| | DAY 56 | Active | -0.61 | 0.162 | -0.94 | -0.27 | | | | | | |
| | | Sham | -0.50 | 0.168 | -0.84 | -0.15 | | | | | | |
| | | Active | | | | | Sham | -0.108 | 0.233 | -0.59 | 0.37 | 0.646 |
| | DAY 84 | Active | -0.44 | 0.149 | -0.74 | -0.13 | | | | | | |
| | | Sham | -0.41 | 0.152 | -0.73 | -0.10 | | | | | | |
| | | Active | | | | | Sham | -0.025 | 0.213 | -0.46 | 0.41 | 0.908 |

*p<0.05 by analysis of covariance of the change from baseline to day 56 and 84 (no device) with device, and baseline covariate.
Confidence interval (CI), Least-squares (LS) mean.
Perceived deficit questionnaire consists of 20 questions on 1 (never have deficit) to 5 (almost always have deficit) scale. A negative change is in the direction of improvement. Source: Listing 9. 31DEC24, T06_pdq_lt45

Table 6.2

Analysis of Covariance
Perceived Deficit Questionnaire (PDQ-20) Treatment Device Comparison in Change from Baseline to Day 56
Age >= 45 Years (Per-Protocol Population)

| | | | Treatment Device Estimate | | | | Treatment Device Comparison | | | | | |
|---|---|---|---|---|---|---|---|---|---|---|---|---|
| | | | | Std | 95% CI | | vs. | Difference | | | | |
| Parameter | Visit | Device | LS Mean | Error | Lower | Upper | Device | LS Mean | Std Error | 95% CI Lower | 95% CI Upper | P-Value |
| 1-Lose train of thought when speaking | BASELINE | Active | 2.50 | 0.289 | 2.21 | 2.79 | | | | | | |
| | | Sham | 2.83 | 0.112 | 2.72 | 2.95 | | | | | | |
| | | Active | | | | | Sham | -0.333 | 0.459 | -1.36 | 0.69 | 0.485 |
| | DAY 56 | Active | -0.23 | 0.329 | -0.97 | 0.51 | | | | | | |
| | | Sham | -0.60 | 0.329 | -1.34 | 0.13 | | | | | | |
| | | Active | | | | | Sham | 0.373 | 0.468 | -0.67 | 1.42 | 0.445 |
| | DAY 84 | Active | -0.23 | 0.233 | -0.76 | 0.30 | | | | | | |
| | | Sham | -0.44 | 0.233 | -0.96 | 0.09 | | | | | | |
| | | Active | | | | | Sham | 0.206 | 0.334 | -0.54 | 0.96 | 0.552 |
| 2-Difficulty remembering names of people | BASELINE | Active | 2.33 | 0.142 | 2.19 | 2.48 | | | | | | |
| | | Sham | 3.17 | 0.207 | 2.96 | 3.37 | | | | | | |
| | | Active | | | | | Sham | -0.833 | 0.373 | -1.66 | -0.00 | 0.049* |
| | DAY 56 | Active | 0.21 | 0.461 | -0.80 | 1.23 | | | | | | |
| | | Sham | -1.05 | 0.461 | -2.06 | -0.03 | | | | | | |
| | | Active | | | | | Sham | 1.262 | 0.672 | -0.21 | 2.73 | 0.086 |
| | DAY 84 | Active | -0.12 | 0.245 | -0.67 | 0.43 | | | | | | |
| | | Sham | -0.71 | 0.245 | -1.27 | -0.16 | | | | | | |
| | | Active | | | | | Sham | 0.595 | 0.384 | -0.27 | 1.46 | 0.155 |
| 3-Forget what you came into the room for | BASELINE | Active | 2.33 | 0.284 | 2.05 | 2.62 | | | | | | |
| | | Sham | 2.50 | 0.151 | 2.35 | 2.65 | | | | | | |
| | | Active | | | | | Sham | -0.167 | 0.477 | -1.23 | 0.90 | 0.734 |
| | DAY 56 | Active | 0.14 | 0.326 | -0.61 | 0.88 | | | | | | |
| | | Sham | -0.30 | 0.326 | -1.05 | 0.45 | | | | | | |
| | | Active | | | | | Sham | 0.437 | 0.463 | -0.62 | 1.50 | 0.372 |
| | DAY 84 | Active | 0.14 | 0.352 | -0.67 | 0.94 | | | | | | |
| | | Sham | -0.30 | 0.352 | -1.11 | 0.51 | | | | | | |
| | | Active | | | | | Sham | 0.437 | 0.499 | -0.71 | 1.58 | 0.406 |

*p<0.05 by analysis of covariance of the change from baseline to day 56 and 84 (no device) with device, and baseline covariate.
Confidence interval (CI), Least-squares (LS) mean.
Perceived deficit questionnaire consists of 20 questions on 1 (never have deficit) to 5 (almost always have deficit) scale. A negative change is in the direction of improvement. Source: Listing 9. 31DEC24, T06_pdq_ge45

Table 6.2

Analysis of Covariance
Perceived Deficit Questionnaire (PDQ-20) Treatment Device Comparison in Change from Baseline to Day 56
Age >= 45 Years (Per-Protocol Population)

| | | | Treatment Device Estimate | | | | Treatment Device Comparison | | | | | |
|---|---|---|---|---|---|---|---|---|---|---|---|---|
| | | | | Std | 95% CI | | vs. | Difference LS | Std | 95% CI | | |
| Parameter | Visit | Device | LS Mean | Error | Lower | Upper | Device | Mean | Error | Lower | Upper | P-Value |
| 4-Trouble getting things organized | BASELINE | Active | 2.50 | 0.289 | 2.21 | 2.79 | | | | | | |
| | | Sham | 2.83 | 0.207 | 2.63 | 3.04 | | | | | | |
| | | Active | | | | | Sham | -0.333 | 0.527 | -1.51 | 0.84 | 0.541 |
| | DAY 56 | Active | -0.35 | 0.200 | -0.80 | 0.10 | | | | | | |
| | | Sham | -0.32 | 0.200 | -0.77 | 0.14 | | | | | | |
| | | Active | | | | | Sham | -0.036 | 0.284 | -0.68 | 0.61 | 0.903 |
| | DAY 84 | Active | 0.15 | 0.264 | -0.45 | 0.74 | | | | | | |
| | | Sham | -0.15 | 0.264 | -0.74 | 0.45 | | | | | | |
| | | Active | | | | | Sham | 0.298 | 0.375 | -0.55 | 1.14 | 0.447 |
| 5-Trouble concentration during conversation | BASELINE | Active | 2.83 | 0.271 | 2.56 | 3.10 | | | | | | |
| | | Sham | 2.67 | 0.142 | 2.52 | 2.81 | | | | | | |
| | | Active | | | | | Sham | 0.167 | 0.453 | -0.84 | 1.18 | 0.721 |
| | DAY 56 | Active | -0.49 | 0.340 | -1.25 | 0.28 | | | | | | |
| | | Sham | -0.68 | 0.340 | -1.44 | 0.09 | | | | | | |
| | | Active | | | | | Sham | 0.190 | 0.482 | -0.89 | 1.28 | 0.702 |
| | DAY 84 | Active | -0.32 | 0.299 | -1.00 | 0.35 | | | | | | |
| | | Sham | -0.51 | 0.299 | -1.19 | 0.16 | | | | | | |
| | | Active | | | | | Sham | 0.190 | 0.424 | -0.77 | 1.15 | 0.664 |
| 6-Forget if you already done something | BASELINE | Active | 2.33 | 0.284 | 2.05 | 2.62 | | | | | | |
| | | Sham | 2.67 | 0.225 | 2.44 | 2.89 | | | | | | |
| | | Active | | | | | Sham | -0.333 | 0.537 | -1.53 | 0.86 | 0.549 |
| | DAY 56 | Active | -0.10 | 0.363 | -0.92 | 0.71 | | | | | | |
| | | Sham | -0.90 | 0.363 | -1.71 | -0.08 | | | | | | |
| | | Active | | | | | Sham | 0.793 | 0.519 | -0.37 | 1.96 | 0.159 |
| | DAY 84 | Active | 0.06 | 0.415 | -0.87 | 0.99 | | | | | | |
| | | Sham | -0.73 | 0.415 | -1.66 | 0.20 | | | | | | |
| | | Active | | | | | Sham | 0.793 | 0.592 | -0.53 | 2.12 | 0.211 |

*p<0.05 by analysis of covariance of the change from baseline to day 56 and 84 (no device) with device, and baseline covariate.
Confidence interval (CI), Least-squares (LS) mean.
Perceived deficit questionnaire consists of 20 questions on 1 (never have deficit) to 5 (almost always have deficit) scale. A negative change is in the direction of improvement. Source: Listing 9. 31DEC24, T06_pdq_ge45

Table 6.2

Analysis of Covariance
Perceived Deficit Questionnaire (PDQ-20) Treatment Device Comparison in Change from Baseline to Day 56
Age >= 45 Years (Per-Protocol Population)

| Parameter | Visit | Device | Treatment Device Estimate LS Mean | Std Error | 95% CI Lower | 95% CI Upper | Treatment Device Comparison vs. Device | Difference LS Mean | Std Error | 95% CI Lower | 95% CI Upper | P-Value |
|---|---|---|---|---|---|---|---|---|---|---|---|---|
| 7-Miss scheduled appointments and meetings | BASELINE | Active | 0.83 | 0.207 | 0.63 | 1.04 | | | | | | |
| | | Sham | 1.17 | 0.207 | 0.96 | 1.37 | | | | | | |
| | | Active | | | | | Sham | -0.333 | 0.435 | -1.30 | 0.64 | 0.461 |
| | DAY 56 | Active | 0.21 | 0.404 | -0.70 | 1.12 | | | | | | |
| | | Sham | -0.04 | 0.404 | -0.96 | 0.87 | | | | | | |
| | | Active | | | | | Sham | 0.253 | 0.581 | -1.06 | 1.56 | 0.674 |
| | DAY 84 | Active | 0.71 | 0.432 | -0.26 | 1.68 | | | | | | |
| | | Sham | -0.38 | 0.432 | -1.35 | 0.60 | | | | | | |
| | | Active | | | | | Sham | 1.086 | 0.620 | -0.31 | 2.48 | 0.112 |
| 8-Difficulty planning what to do during the day | BASELINE | Active | 2.00 | 0.246 | 1.75 | 2.25 | | | | | | |
| | | Sham | 1.83 | 0.112 | 1.72 | 1.95 | | | | | | |
| | | Active | | | | | Sham | 0.167 | 0.401 | -0.73 | 1.06 | 0.687 |
| | DAY 56 | Active | 0.02 | 0.218 | -0.47 | 0.51 | | | | | | |
| | | Sham | -0.19 | 0.218 | -0.68 | 0.30 | | | | | | |
| | | Active | | | | | Sham | 0.209 | 0.309 | -0.49 | 0.91 | 0.516 |
| | DAY 84 | Active | 0.35 | 0.455 | -0.66 | 1.37 | | | | | | |
| | | Sham | -0.69 | 0.455 | -1.70 | 0.33 | | | | | | |
| | | Active | | | | | Sham | 1.042 | 0.644 | -0.39 | 2.48 | 0.137 |
| 9-Trouble concentrating on things like TV or book | BASELINE | Active | 2.50 | 0.485 | 2.02 | 2.98 | | | | | | |
| | | Sham | 2.50 | 0.151 | 2.35 | 2.65 | | | | | | |
| | | Active | | | | | Sham | 0.000 | 0.753 | -1.68 | 1.68 | 1.000 |
| | DAY 56 | Active | 0.17 | 0.334 | -0.59 | 0.92 | | | | | | |
| | | Sham | -0.00 | 0.334 | -0.75 | 0.75 | | | | | | |
| | | Active | | | | | Sham | 0.167 | 0.473 | -0.90 | 1.23 | 0.732 |
| | DAY 84 | Active | 0.33 | 0.285 | -0.31 | 0.98 | | | | | | |
| | | Sham | -0.83 | 0.285 | -1.48 | -0.19 | | | | | | |
| | | Active | | | | | Sham | 1.167 | 0.403 | 0.26 | 2.08 | 0.018* |

*p<0.05 by analysis of covariance of the change from baseline to day 56 and 84 (no device) with device, and baseline covariate.
Confidence interval (CI), Least-squares (LS) mean.
Perceived deficit questionnaire consists of 20 questions on 1 (never have deficit) to 5 (almost always have deficit) scale. A negative change is in the direction of improvement. Source: Listing 9. 31DEC24, T06_pdq_ge45

Table 6.2

Analysis of Covariance
Perceived Deficit Questionnaire (PDQ-20) Treatment Device Comparison in Change from Baseline to Day 56
Age >= 45 Years (Per-Protocol Population)

| | | | Treatment Device Estimate | | | | Treatment Device Comparison | | | | | |
|---|---|---|---|---|---|---|---|---|---|---|---|---|
| | | | | | | | | Difference | | | | |
| | | | | Std | 95% CI | | vs. | LS | Std | 95% CI | | |
| Parameter | Visit | Device | LS Mean | Error | Lower | Upper | Device | Mean | Error | Lower | Upper | P-Value |
| 10-Forget what you did the night before | BASELINE | Active | 2.00 | 0.389 | 1.61 | 2.39 | | | | | | |
| | | Sham | 2.17 | 0.271 | 1.90 | 2.44 | | | | | | |
| | | Active | | | | | Sham | -0.167 | 0.703 | -1.73 | 1.40 | 0.817 |
| | DAY 56 | Active | 0.32 | 0.275 | -0.30 | 0.93 | | | | | | |
| | | Sham | -0.48 | 0.275 | -1.10 | 0.13 | | | | | | |
| | | Active | | | | | Sham | 0.798 | 0.389 | -0.07 | 1.67 | 0.068 |
| | DAY 84 | Active | 0.15 | 0.135 | -0.16 | 0.46 | | | | | | |
| | | Sham | -0.82 | 0.135 | -1.12 | -0.51 | | | | | | |
| | | Active | | | | | Sham | 0.965 | 0.192 | 0.53 | 1.40 | <0.001* |
| 11-Forget the date unless you looked it up | BASELINE | Active | 3.17 | 0.271 | 2.90 | 3.44 | | | | | | |
| | | Sham | 2.33 | 0.142 | 2.19 | 2.48 | | | | | | |
| | | Active | | | | | Sham | 0.833 | 0.453 | -0.18 | 1.84 | 0.096 |
| | DAY 56 | Active | -0.09 | 0.168 | -0.47 | 0.29 | | | | | | |
| | | Sham | -0.25 | 0.168 | -0.63 | 0.13 | | | | | | |
| | | Active | | | | | Sham | 0.160 | 0.256 | -0.41 | 0.73 | 0.546 |
| | DAY 84 | Active | 0.08 | 0.262 | -0.50 | 0.66 | | | | | | |
| | | Sham | -0.58 | 0.262 | -1.16 | 0.00 | | | | | | |
| | | Active | | | | | Sham | 0.660 | 0.382 | -0.18 | 1.50 | 0.112 |
| 12-Trouble getting started even with things to do | BASELINE | Active | 2.50 | 0.289 | 2.21 | 2.79 | | | | | | |
| | | Sham | 2.00 | 0.174 | 1.83 | 2.17 | | | | | | |
| | | Active | | | | | Sham | 0.500 | 0.500 | -0.61 | 1.61 | 0.341 |
| | DAY 56 | Active | -0.58 | 0.280 | -1.21 | 0.05 | | | | | | |
| | | Sham | -0.25 | 0.280 | -0.88 | 0.38 | | | | | | |
| | | Active | | | | | Sham | -0.325 | 0.403 | -1.23 | 0.58 | 0.439 |
| | DAY 84 | Active | 0.09 | 0.302 | -0.59 | 0.77 | | | | | | |
| | | Sham | -0.25 | 0.302 | -0.93 | 0.43 | | | | | | |
| | | Active | | | | | Sham | 0.342 | 0.433 | -0.63 | 1.31 | 0.449 |

*p<0.05 by analysis of covariance of the change from baseline to day 56 and 84 (no device) with device, and baseline covariate.
Confidence interval (CI), Least-squares (LS) mean.
Perceived deficit questionnaire consists of 20 questions on 1 (never have deficit) to 5 (almost always have deficit) scale. A negative change is in the direction of improvement. Source: Listing 9. 31DEC24, T06_pdq_ge45

Table 6.2

Analysis of Covariance
Perceived Deficit Questionnaire (PDQ-20) Treatment Device Comparison in Change from Baseline to Day 56
Age >= 45 Years (Per-Protocol Population)

| | | | Treatment Device Estimate | | | | Treatment Device Comparison | | | | | |
|---|---|---|---|---|---|---|---|---|---|---|---|---|
| | | | | | | | | Difference | | | | |
| | | | | Std | 95% CI | | vs. | LS | Std | 95% CI | | |
| Parameter | Visit | Device | LS Mean | Error | Lower | Upper | Device | Mean | Error | Lower | Upper | P-Value |
| 13-Find your mind drifting | BASELINE | Active | 3.00 | 0.246 | 2.75 | 3.25 | | | | | | |
| | | Sham | 2.83 | 0.207 | 2.63 | 3.04 | | | | | | |
| | | Active | | | | | Sham | 0.167 | 0.477 | -0.90 | 1.23 | 0.734 |
| | DAY 56 | Active | -0.33 | 0.268 | -0.93 | 0.27 | | | | | | |
| | | Sham | -0.17 | 0.268 | -0.77 | 0.43 | | | | | | |
| | | Active | | | | | Sham | -0.156 | 0.380 | -1.00 | 0.69 | 0.690 |
| | DAY 84 | Active | 0.01 | 0.164 | -0.36 | 0.37 | | | | | | |
| | | Sham | -0.51 | 0.164 | -0.87 | -0.14 | | | | | | |
| | | Active | | | | | Sham | 0.510 | 0.233 | -0.01 | 1.03 | 0.055 |
| 14-Forget what talked about after call | BASELINE | Active | 2.17 | 0.271 | 1.90 | 2.44 | | | | | | |
| | | Sham | 2.17 | 0.207 | 1.96 | 2.37 | | | | | | |
| | | Active | | | | | Sham | 0.000 | 0.506 | -1.13 | 1.13 | 1.000 |
| | DAY 56 | Active | -0.33 | 0.470 | -1.39 | 0.73 | | | | | | |
| | | Sham | -0.50 | 0.470 | -1.56 | 0.56 | | | | | | |
| | | Active | | | | | Sham | 0.167 | 0.665 | -1.33 | 1.66 | 0.808 |
| | DAY 84 | Active | -0.17 | 0.242 | -0.71 | 0.38 | | | | | | |
| | | Sham | -0.83 | 0.242 | -1.38 | -0.29 | | | | | | |
| | | Active | | | | | Sham | 0.667 | 0.342 | -0.11 | 1.44 | 0.083 |
| 15-Forget to do things like turn off stove or lock door | BASELINE | Active | 1.17 | 0.207 | 0.96 | 1.37 | | | | | | |
| | | Sham | 1.50 | 0.151 | 1.35 | 1.65 | | | | | | |
| | | Active | | | | | Sham | -0.333 | 0.380 | -1.18 | 0.51 | 0.401 |
| | DAY 56 | Active | -0.09 | 0.237 | -0.62 | 0.45 | | | | | | |
| | | Sham | -0.41 | 0.237 | -0.95 | 0.12 | | | | | | |
| | | Active | | | | | Sham | 0.328 | 0.339 | -0.43 | 1.09 | 0.357 |
| | DAY 84 | Active | 0.08 | 0.268 | -0.52 | 0.68 | | | | | | |
| | | Sham | -0.25 | 0.268 | -0.85 | 0.35 | | | | | | |
| | | Active | | | | | Sham | 0.328 | 0.383 | -0.53 | 1.19 | 0.412 |

*p<0.05 by analysis of covariance of the change from baseline to day 56 and 84 (no device) with device, and baseline covariate.
Confidence interval (CI), Least-squares (LS) mean.
Perceived deficit questionnaire consists of 20 questions on 1 (never have deficit) to 5 (almost always have deficit) scale. A negative change is in the direction of improvement. Source: Listing 9. 31DEC24, T06_pdq_ge45

Table 6.2

Analysis of Covariance
Perceived Deficit Questionnaire (PDQ-20) Treatment Device Comparison in Change from Baseline to Day 56
Age >= 45 Years (Per-Protocol Population)

| | | | Treatment Device Estimate | | | | Treatment Device Comparison | | | | | |
|---|---|---|---|---|---|---|---|---|---|---|---|---|
| | | | | | | | | Difference | | | | |
| | | | | Std | 95% CI | | vs. | LS | Std | 95% CI | | |
| Parameter | Visit | Device | LS Mean | Error | Lower | Upper | Device | Mean | Error | Lower | Upper | P-Value |
| 16-Feel like your mind went totally blank | BASELINE | Active | 2.67 | 0.414 | 2.25 | 3.08 | | | | | | |
| | | Sham | 2.00 | 0.174 | 1.83 | 2.17 | | | | | | |
| | | Active | | | | | Sham | 0.667 | 0.667 | -0.82 | 2.15 | 0.341 |
| | DAY 56 | Active | -0.09 | 0.310 | -0.78 | 0.60 | | | | | | |
| | | Sham | -0.24 | 0.310 | -0.93 | 0.45 | | | | | | |
| | | Active | | | | | Sham | 0.150 | 0.445 | -0.84 | 1.14 | 0.743 |
| | DAY 84 | Active | -0.76 | 0.307 | -1.44 | -0.07 | | | | | | |
| | | Sham | -0.41 | 0.307 | -1.10 | 0.28 | | | | | | |
| | | Active | | | | | Sham | -0.350 | 0.442 | -1.33 | 0.63 | 0.447 |
| 17-Trouble holding phone numbers even for a few seconds | BASELINE | Active | 2.50 | 0.151 | 2.35 | 2.65 | | | | | | |
| | | Sham | 3.00 | 0.174 | 2.83 | 3.17 | | | | | | |
| | | Active | | | | | Sham | -0.500 | 0.342 | -1.26 | 0.26 | 0.174 |
| | DAY 56 | Active | -0.40 | 0.369 | -1.24 | 0.43 | | | | | | |
| | | Sham | -0.43 | 0.369 | -1.26 | 0.40 | | | | | | |
| | | Active | | | | | Sham | 0.024 | 0.550 | -1.22 | 1.26 | 0.966 |
| | DAY 84 | Active | -0.07 | 0.486 | -1.15 | 1.01 | | | | | | |
| | | Sham | -0.76 | 0.486 | -1.84 | 0.32 | | | | | | |
| | | Active | | | | | Sham | 0.690 | 0.709 | -0.88 | 2.26 | 0.352 |
| 18-Forgot what you did last week | BASELINE | Active | 2.33 | 0.284 | 2.05 | 2.62 | | | | | | |
| | | Sham | 2.17 | 0.271 | 1.90 | 2.44 | | | | | | |
| | | Active | | | | | Sham | 0.167 | 0.582 | -1.13 | 1.46 | 0.780 |
| | DAY 56 | Active | 0.01 | 0.345 | -0.76 | 0.79 | | | | | | |
| | | Sham | -0.18 | 0.345 | -0.96 | 0.60 | | | | | | |
| | | Active | | | | | Sham | 0.196 | 0.489 | -0.91 | 1.30 | 0.698 |
| | DAY 84 | Active | 0.18 | 0.377 | -0.67 | 1.03 | | | | | | |
| | | Sham | -0.68 | 0.377 | -1.53 | 0.17 | | | | | | |
| | | Active | | | | | Sham | 0.862 | 0.534 | -0.34 | 2.06 | 0.139 |

*p<0.05 by analysis of covariance of the change from baseline to day 56 and 84 (no device) with device, and baseline covariate.
Confidence interval (CI), Least-squares (LS) mean.
Perceived deficit questionnaire consists of 20 questions on 1 (never have deficit) to 5 (almost always have deficit) scale. A negative change is in the direction of improvement. Source: Listing 9. 31DEC24, T06_pdq_ge45

Table 6.2

Analysis of Covariance
Perceived Deficit Questionnaire (PDQ-20) Treatment Device Comparison in Change from Baseline to Day 56
Age >= 45 Years (Per-Protocol Population)

| | | | Treatment Device Estimate | | | | Treatment Device Comparison | | | | | |
|---|---|---|---|---|---|---|---|---|---|---|---|---|
| | | | | | | | | Difference | | | | |
| | | | | Std | 95% CI | | vs. | LS | Std | 95% CI | | |
| Parameter | Visit | Device | LS Mean | Error | Lower | Upper | Device | Mean | Error | Lower | Upper | P-Value |
| 19-Forget to take your medication | BASELINE | Active | 1.33 | 0.225 | 1.11 | 1.56 | | | | | | |
| | | Sham | 1.33 | 0.142 | 1.19 | 1.48 | | | | | | |
| | | Active | | | | | Sham | 0.000 | 0.394 | -0.88 | 0.88 | 1.000 |
| | DAY 56 | Active | -0.00 | 0.254 | -0.57 | 0.57 | | | | | | |
| | | Sham | -0.50 | 0.254 | -1.07 | 0.07 | | | | | | |
| | | Active | | | | | Sham | 0.500 | 0.359 | -0.31 | 1.31 | 0.196 |
| | DAY 84 | Active | 0.67 | 0.272 | 0.05 | 1.28 | | | | | | |
| | | Sham | -0.83 | 0.272 | -1.45 | -0.22 | | | | | | |
| | | Active | | | | | Sham | 1.500 | 0.385 | 0.63 | 2.37 | 0.003* |
| 20-Have trouble making decisions | BASELINE | Active | 2.67 | 0.414 | 2.25 | 3.08 | | | | | | |
| | | Sham | 2.17 | 0.112 | 2.05 | 2.28 | | | | | | |
| | | Active | | | | | Sham | 0.500 | 0.637 | -0.92 | 1.92 | 0.451 |
| | DAY 56 | Active | -0.20 | 0.287 | -0.85 | 0.45 | | | | | | |
| | | Sham | -0.46 | 0.287 | -1.11 | 0.19 | | | | | | |
| | | Active | | | | | Sham | 0.261 | 0.412 | -0.67 | 1.20 | 0.542 |
| | DAY 84 | Active | 0.13 | 0.352 | -0.67 | 0.93 | | | | | | |
| | | Sham | -0.46 | 0.352 | -1.26 | 0.34 | | | | | | |
| | | Active | | | | | Sham | 0.595 | 0.503 | -0.55 | 1.74 | 0.268 |
| Mean PDQ Questions 1-20 | BASELINE | Active | 2.28 | 0.205 | 2.08 | 2.49 | | | | | | |
| | | Sham | 2.29 | 0.108 | 2.18 | 2.40 | | | | | | |
| | | Active | | | | | Sham | -0.008 | 0.344 | -0.77 | 0.76 | 0.981 |
| | DAY 56 | Active | -0.10 | 0.202 | -0.55 | 0.35 | | | | | | |
| | | Sham | -0.41 | 0.202 | -0.86 | 0.04 | | | | | | |
| | | Active | | | | | Sham | 0.309 | 0.285 | -0.33 | 0.95 | 0.304 |
| | DAY 84 | Active | 0.08 | 0.172 | -0.30 | 0.47 | | | | | | |
| | | Sham | -0.57 | 0.172 | -0.95 | -0.18 | | | | | | |
| | | Active | | | | | Sham | 0.651 | 0.244 | 0.10 | 1.20 | 0.025* |

*p<0.05 by analysis of covariance of the change from baseline to day 56 and 84 (no device) with device, and baseline covariate.
Confidence interval (CI), Least-squares (LS) mean.
Perceived deficit questionnaire consists of 20 questions on 1 (never have deficit) to 5 (almost always have deficit) scale. A negative change is in the direction of improvement. Source: Listing 9. 31DEC24, T06_pdq_ge45

Table 7

Analysis of Covariance
MSBQ Treatment Device Comparison in Change from Baseline to Day 56 (Per-Protocol Population)

| | | | Treatment Estimate | | | | Treatment Comparison | | | | | |
|---|---|---|---|---|---|---|---|---|---|---|---|---|
| | | | | | 95% CI | | | Difference | | 95% CI | | |
| Parameter | Visit | Device | LS Mean | Std Error | Lower | Upper | vs. Device | LS Mean | Std Error | Lower | Upper | P-Value |
| Breathing | BASELINE | Active | 4.10 | 0.733 | 3.36 | 4.83 | | | | | | |
| | | Sham | 3.25 | 0.774 | 2.48 | 4.02 | | | | | | |
| | | Active | | | | | Sham | 0.845 | 1.065 | -1.31 | 3.00 | 0.432 |
| | DAY 56 | Active | 0.12 | 0.419 | -0.73 | 0.97 | | | | | | |
| | | Sham | -0.33 | 0.426 | -1.20 | 0.53 | | | | | | |
| | | Active | | | | | Sham | 0.451 | 0.572 | -0.71 | 1.61 | 0.435 |
| Ears/Nose/Throat | BASELINE | Active | 10.19 | 1.609 | 8.58 | 11.80 | | | | | | |
| | | Sham | 6.60 | 0.944 | 5.66 | 7.54 | | | | | | |
| | | Active | | | | | Sham | 3.590 | 1.889 | -0.23 | 7.41 | 0.065 |
| | DAY 56 | Active | -0.57 | 1.164 | -2.93 | 1.79 | | | | | | |
| | | Sham | -1.38 | 1.179 | -3.77 | 1.01 | | | | | | |
| | | Active | | | | | Sham | 0.808 | 1.615 | -2.46 | 4.08 | 0.620 |
| Fatigue | BASELINE | Active | 8.81 | 0.783 | 8.03 | 9.59 | | | | | | |
| | | Sham | 8.25 | 0.464 | 7.79 | 8.71 | | | | | | |
| | | Active | | | | | Sham | 0.560 | 0.921 | -1.30 | 2.42 | 0.547 |
| | DAY 56 | Active | -0.72 | 0.535 | -1.80 | 0.37 | | | | | | |
| | | Sham | -1.70 | 0.547 | -2.81 | -0.60 | | | | | | |
| | | Active | | | | | Sham | 0.988 | 0.729 | -0.49 | 2.47 | 0.184 |
| Memory | BASELINE | Active | 17.00 | 1.510 | 15.49 | 18.51 | | | | | | |
| | | Sham | 15.80 | 1.111 | 14.69 | 16.91 | | | | | | |
| | | Active | | | | | Sham | 1.200 | 1.890 | -2.62 | 5.02 | 0.529 |
| | DAY 56 | Active | -2.94 | 1.026 | -5.02 | -0.86 | | | | | | |
| | | Sham | -4.74 | 1.043 | -6.85 | -2.62 | | | | | | |
| | | Active | | | | | Sham | 1.795 | 1.397 | -1.04 | 4.62 | 0.207 |
| Other | BASELINE | Active | 10.57 | 1.722 | 8.85 | 12.29 | | | | | | |
| | | Sham | 8.60 | 0.930 | 7.67 | 9.53 | | | | | | |
| | | Active | | | | | Sham | 1.971 | 1.985 | -2.04 | 5.99 | 0.327 |
| | DAY 56 | Active | -1.52 | 0.989 | -3.52 | 0.49 | | | | | | |
| | | Sham | -2.21 | 1.001 | -4.24 | -0.18 | | | | | | |
| | | Active | | | | | Sham | 0.693 | 1.349 | -2.04 | 3.43 | 0.611 |

*p<0.05 by analysis of covariance of the change from baseline to day 56 with terms for device, age >=45 years (yes,no) and baseline covariate. Confidence interval (CI), Least-squares (LS) mean. A lower score is in the direction of improvement. Source: Listing 11.

31DEC24, T07_msbq2

Table 7

Analysis of Covariance
MSBQ Treatment Device Comparison in Change from Baseline to Day 56 (Per-Protocol Population)

| | | | Treatment Estimate | | | | Treatment Comparison | Difference | | | | |
|---|---|---|---|---|---|---|---|---|---|---|---|---|
| | | | | Std | 95% CI | | vs. | LS | Std | 95% CI | | |
| Parameter | Visit | Device | LS Mean | Error | Lower | Upper | Device | Mean | Error | Lower | Upper | P-Value |
| Pain | BASELINE | Active | 4.05 | 0.567 | 3.48 | 4.61 | | | | | | |
| | | Sham | 3.85 | 0.666 | 3.18 | 4.52 | | | | | | |
| | | Active | | | | | Sham | 0.198 | 0.872 | -1.57 | 1.96 | 0.822 |
| | DAY 56 | Active | -0.59 | 0.372 | -1.35 | 0.16 | | | | | | |
| | | Sham | -0.72 | 0.379 | -1.49 | 0.05 | | | | | | |
| | | Active | | | | | Sham | 0.127 | 0.505 | -0.90 | 1.15 | 0.802 |
| Sleep | BASELINE | Active | 6.38 | 0.455 | 5.93 | 6.84 | | | | | | |
| | | Sham | 6.25 | 0.575 | 5.67 | 6.83 | | | | | | |
| | | Active | | | | | Sham | 0.131 | 0.730 | -1.34 | 1.61 | 0.858 |
| | DAY 56 | Active | -1.08 | 0.497 | -2.08 | -0.07 | | | | | | |
| | | Sham | -1.11 | 0.505 | -2.13 | -0.08 | | | | | | |
| | | Active | | | | | Sham | 0.030 | 0.675 | -1.34 | 1.40 | 0.965 |
| Stomach | BASELINE | Active | 5.57 | 0.977 | 4.59 | 6.55 | | | | | | |
| | | Sham | 4.20 | 0.866 | 3.33 | 5.07 | | | | | | |
| | | Active | | | | | Sham | 1.371 | 1.311 | -1.28 | 4.02 | 0.302 |
| | DAY 56 | Active | -0.66 | 0.498 | -1.67 | 0.35 | | | | | | |
| | | Sham | -0.48 | 0.506 | -1.50 | 0.55 | | | | | | |
| | | Active | | | | | Sham | -0.181 | 0.681 | -1.56 | 1.20 | 0.791 |

*p<0.05 by analysis of covariance of the change from baseline to day 56 with terms for device, age >=45 years (yes,no) and baseline covariate. Confidence interval (CI), Least-squares (LS) mean. A lower score is in the direction of improvement. Source: Listing 11.

31DEC24, T07_msbq2

Table 7.1

Analysis of Covariance
MSBQ Treatment Device Comparison in Change from Baseline to Day 56
Age < 45 Years (Per-Protocol Population)

| | | | Treatment Estimate | | | | Treatment Comparison | | | | | |
|---|---|---|---|---|---|---|---|---|---|---|---|---|
| | | | | Std | 95% CI | | vs. | Difference LS | Difference Std | 95% CI | | |
| Parameter | Visit | Device | LS Mean | Error | Lower | Upper | Device | Mean | Error | Lower | Upper | P-Value |
| Breathing | BASELINE | Active | 4.00 | 0.910 | 3.09 | 4.91 | | | | | | |
| | | Sham | 3.36 | 0.987 | 2.37 | 4.34 | | | | | | |
| | | Active | | | | | Sham | 0.643 | 1.340 | -2.11 | 3.39 | 0.635 |
| | DAY 56 | Active | 0.01 | 0.509 | -1.04 | 1.05 | | | | | | |
| | | Sham | -0.44 | 0.526 | -1.52 | 0.65 | | | | | | |
| | | Active | | | | | Sham | 0.443 | 0.733 | -1.06 | 1.95 | 0.551 |
| Ears/Nose/Throat | BASELINE | Active | 10.60 | 2.093 | 8.51 | 12.69 | | | | | | |
| | | Sham | 5.86 | 0.876 | 4.98 | 6.73 | | | | | | |
| | | Active | | | | | Sham | 4.743 | 2.328 | -0.03 | 9.52 | 0.051 |
| | DAY 56 | Active | -1.63 | 1.395 | -4.50 | 1.24 | | | | | | |
| | | Sham | -1.97 | 1.447 | -4.94 | 1.01 | | | | | | |
| | | Active | | | | | Sham | 0.338 | 2.080 | -3.94 | 4.61 | 0.872 |
| Fatigue | BASELINE | Active | 9.00 | 0.926 | 8.07 | 9.93 | | | | | | |
| | | Sham | 8.71 | 0.559 | 8.16 | 9.27 | | | | | | |
| | | Active | | | | | Sham | 0.286 | 1.101 | -1.97 | 2.54 | 0.797 |
| | DAY 56 | Active | -1.36 | 0.636 | -2.67 | -0.05 | | | | | | |
| | | Sham | -1.83 | 0.658 | -3.18 | -0.47 | | | | | | |
| | | Active | | | | | Sham | 0.466 | 0.916 | -1.42 | 2.35 | 0.616 |
| Memory | BASELINE | Active | 17.07 | 1.896 | 15.17 | 18.96 | | | | | | |
| | | Sham | 15.79 | 1.597 | 14.19 | 17.38 | | | | | | |
| | | Active | | | | | Sham | 1.281 | 2.497 | -3.84 | 6.40 | 0.612 |
| | DAY 56 | Active | -4.35 | 1.145 | -6.70 | -2.00 | | | | | | |
| | | Sham | -4.77 | 1.185 | -7.20 | -2.33 | | | | | | |
| | | Active | | | | | Sham | 0.417 | 1.652 | -2.98 | 3.81 | 0.803 |
| Other | BASELINE | Active | 10.67 | 2.067 | 8.60 | 12.73 | | | | | | |
| | | Sham | 8.00 | 1.195 | 6.80 | 9.20 | | | | | | |
| | | Active | | | | | Sham | 2.667 | 2.433 | -2.33 | 7.66 | 0.283 |
| | DAY 56 | Active | -2.61 | 1.088 | -4.85 | -0.37 | | | | | | |
| | | Sham | -1.70 | 1.127 | -4.02 | 0.61 | | | | | | |
| | | Active | | | | | Sham | -0.906 | 1.584 | -4.16 | 2.35 | 0.572 |

*p<0.05 by analysis of covariance of the change from baseline to day 56 with terms for device, and baseline covariate.
Confidence interval (CI), Least-squares (LS) mean. A lower score is in the direction of improvement. Source: Listing 11.

31DEC24, T07_msbq2_lt45

Table 7.1

Analysis of Covariance
MSBQ Treatment Device Comparison in Change from Baseline to Day 56
Age < 45 Years (Per-Protocol Population)

| | | | Treatment Estimate | | | | Treatment Comparison | | | | | |
|---|---|---|---|---|---|---|---|---|---|---|---|---|
| | | | | Std | 95% CI | | vs. | Difference | | | | |
| | | | | | | | | LS | Std | 95% CI | | |
| Parameter | Visit | Device | LS Mean | Error | Lower | Upper | Device | Mean | Error | Lower | Upper | P-Value |
| Pain | BASELINE | Active | 4.00 | 0.697 | 3.30 | 4.70 | | | | | | |
| | | Sham | 4.36 | 0.899 | 3.46 | 5.26 | | | | | | |
| | | Active | | | | | Sham | -0.357 | 1.129 | -2.67 | 1.96 | 0.754 |
| | DAY 56 | Active | -0.78 | 0.364 | -1.53 | -0.04 | | | | | | |
| | | Sham | -0.87 | 0.377 | -1.65 | -0.10 | | | | | | |
| | | Active | | | | | Sham | 0.089 | 0.525 | -0.99 | 1.17 | 0.867 |
| Sleep | BASELINE | Active | 6.07 | 0.621 | 5.45 | 6.69 | | | | | | |
| | | Sham | 6.57 | 0.661 | 5.91 | 7.23 | | | | | | |
| | | Active | | | | | Sham | -0.505 | 0.906 | -2.36 | 1.35 | 0.582 |
| | DAY 56 | Active | -0.49 | 0.500 | -1.52 | 0.54 | | | | | | |
| | | Sham | -0.55 | 0.517 | -1.61 | 0.52 | | | | | | |
| | | Active | | | | | Sham | 0.056 | 0.721 | -1.43 | 1.54 | 0.939 |
| Stomach | BASELINE | Active | 5.80 | 1.101 | 4.70 | 6.90 | | | | | | |
| | | Sham | 3.93 | 1.160 | 2.77 | 5.09 | | | | | | |
| | | Active | | | | | Sham | 1.871 | 1.598 | -1.41 | 5.15 | 0.252 |
| | DAY 56 | Active | -0.96 | 0.604 | -2.20 | 0.28 | | | | | | |
| | | Sham | -0.47 | 0.626 | -1.76 | 0.81 | | | | | | |
| | | Active | | | | | Sham | -0.487 | 0.880 | -2.30 | 1.32 | 0.585 |

*p<0.05 by analysis of covariance of the change from baseline to day 56 with terms for device, and baseline covariate.
Confidence interval (CI), Least-squares (LS) mean. A lower score is in the direction of improvement. Source: Listing 11.

31DEC24, T07_msbq2_lt45

Table 7.2

Analysis of Covariance
MSBQ Treatment Device Comparison in Change from Baseline to Day 56
Age >= 45 Years (Per-Protocol Population)

| | | | Treatment Estimate | | | | Treatment Comparison | | | | | |
|---|---|---|---|---|---|---|---|---|---|---|---|---|
| | | | | Std | 95% CI | | vs. | Difference | | | | |
| | | | | | | | | LS | Std | 95% CI | | |
| Parameter | Visit | Device | LS Mean | Error | Lower | Upper | Device | Mean | Error | Lower | Upper | P-Value |
| Breathing | BASELINE | Active | 4.33 | 1.308 | 3.03 | 5.64 | | | | | | |
| | | Sham | 3.00 | 1.291 | 1.71 | 4.29 | | | | | | |
| | | Active | | | | | Sham | 1.333 | 1.838 | -2.76 | 5.43 | 0.485 |
| | DAY 56 | Active | 0.14 | 0.543 | -1.09 | 1.37 | | | | | | |
| | | Sham | -0.14 | 0.543 | -1.37 | 1.09 | | | | | | |
| | | Active | | | | | Sham | 0.285 | 0.778 | -1.48 | 2.05 | 0.723 |
| Ears/Nose/Throat | BASELINE | Active | 9.17 | 2.301 | 6.87 | 11.47 | | | | | | |
| | | Sham | 8.33 | 2.404 | 5.93 | 10.74 | | | | | | |
| | | Active | | | | | Sham | 0.833 | 3.327 | -6.58 | 8.25 | 0.807 |
| | DAY 56 | Active | 0.97 | 1.704 | -2.88 | 4.82 | | | | | | |
| | | Sham | -1.47 | 1.704 | -5.32 | 2.38 | | | | | | |
| | | Active | | | | | Sham | 2.441 | 2.413 | -3.02 | 7.90 | 0.338 |
| Fatigue | BASELINE | Active | 8.33 | 1.585 | 6.75 | 9.92 | | | | | | |
| | | Sham | 7.17 | 0.703 | 6.46 | 7.87 | | | | | | |
| | | Active | | | | | Sham | 1.167 | 1.734 | -2.70 | 5.03 | 0.516 |
| | DAY 56 | Active | 0.24 | 0.645 | -1.22 | 1.70 | | | | | | |
| | | Sham | -1.74 | 0.645 | -3.20 | -0.28 | | | | | | |
| | | Active | | | | | Sham | 1.983 | 0.922 | -0.10 | 4.07 | 0.060 |
| Memory | BASELINE | Active | 16.83 | 2.600 | 14.23 | 19.43 | | | | | | |
| | | Sham | 15.83 | 0.401 | 15.43 | 16.23 | | | | | | |
| | | Active | | | | | Sham | 1.000 | 2.631 | -4.86 | 6.86 | 0.712 |
| | DAY 56 | Active | -0.68 | 1.068 | -3.10 | 1.73 | | | | | | |
| | | Sham | -5.48 | 1.068 | -7.90 | -3.07 | | | | | | |
| | | Active | | | | | Sham | 4.801 | 1.516 | 1.37 | 8.23 | 0.011* |
| Other | BASELINE | Active | 10.33 | 3.403 | 6.93 | 13.74 | | | | | | |
| | | Sham | 10.00 | 1.317 | 8.68 | 11.32 | | | | | | |
| | | Active | | | | | Sham | 0.333 | 3.648 | -7.80 | 8.46 | 0.929 |
| | DAY 56 | Active | 0.70 | 1.580 | -2.88 | 4.27 | | | | | | |
| | | Sham | -4.03 | 1.580 | -7.61 | -0.46 | | | | | | |
| | | Active | | | | | Sham | 4.730 | 2.235 | -0.33 | 9.79 | 0.063 |

*p<0.05 by analysis of covariance of the change from baseline to day 56 with terms for device, and baseline covariate.
Confidence interval (CI), Least-squares (LS) mean. A lower score is in the direction of improvement. Source: Listing 11.

31DEC24, T07_msbq2_ge45

Table 7.2

Analysis of Covariance
MSBQ Treatment Device Comparison in Change from Baseline to Day 56
Age >= 45 Years (Per-Protocol Population)

| | | Treatment Estimate | | | | Treatment Comparison | | | | | |
|---|---|---|---|---|---|---|---|---|---|---|---|
| | | | | | | | Difference | | | | |
| | | | | Std | 95% CI | | vs. | LS | Std | 95% CI | | |
| Parameter | Visit | Device | LS Mean | Error | Lower | Upper | Device | Mean | Error | Lower | Upper | P-Value |

| Parameter | Visit | Device | LS Mean | Std Error | 95% CI Lower | 95% CI Upper | vs. Device | Difference LS Mean | Difference Std Error | 95% CI Lower | 95% CI Upper | P-Value |
|---|---|---|---|---|---|---|---|---|---|---|---|---|
| Pain | BASELINE | Active | 4.17 | 1.046 | 3.12 | 5.21 | | | | | | |
| | | Sham | 2.67 | 0.558 | 2.11 | 3.22 | | | | | | |
| | | Active | | | | | Sham | 1.500 | 1.186 | -1.14 | 4.14 | 0.234 |
| | DAY 56 | Active | -0.76 | 0.770 | -2.50 | 0.98 | | | | | | |
| | | Sham | -0.07 | 0.770 | -1.81 | 1.67 | | | | | | |
| | | Active | | | | | Sham | -0.694 | 1.128 | -3.25 | 1.86 | 0.553 |
| Sleep | BASELINE | Active | 7.17 | 0.167 | 7.00 | 7.33 | | | | | | |
| | | Sham | 5.50 | 1.176 | 4.32 | 6.68 | | | | | | |
| | | Active | | | | | Sham | 1.667 | 1.188 | -0.98 | 4.31 | 0.191 |
| | DAY 56 | Active | -1.55 | 1.234 | -4.34 | 1.24 | | | | | | |
| | | Sham | -1.79 | 1.234 | -4.58 | 1.00 | | | | | | |
| | | Active | | | | | Sham | 0.239 | 1.821 | -3.88 | 4.36 | 0.899 |
| Stomach | BASELINE | Active | 5.00 | 2.191 | 2.81 | 7.19 | | | | | | |
| | | Sham | 4.83 | 1.108 | 3.73 | 5.94 | | | | | | |
| | | Active | | | | | Sham | 0.167 | 2.455 | -5.30 | 5.64 | 0.947 |
| | DAY 56 | Active | -0.14 | 0.743 | -1.82 | 1.54 | | | | | | |
| | | Sham | -0.69 | 0.743 | -2.37 | 0.99 | | | | | | |
| | | Active | | | | | Sham | 0.553 | 1.050 | -1.82 | 2.93 | 0.611 |

*p<0.05 by analysis of covariance of the change from baseline to day 56 with terms for device, and baseline covariate.
Confidence interval (CI), Least-squares (LS) mean. A lower score is in the direction of improvement. Source: Listing 11.

31DEC24, T07_msbq2_ge45

Table 8

Compliance
(Safety Population)

| Parameter | Statistic | Treatment Device: Active (N= 23) | Treatment Device: Sham (N= 20) | Treatment Device: Total (N= 43) | P-Value |
|---|---|---|---|---|---|
| Duration (days) | N | 23 | 20 | 43 | 0.263 |
| | Mean (SD) | 51.0 (13.59) | 54.5 ( 2.28) | 52.6 (10.11) | |
| | Median (IQR) | 55.0 ( 0.0) | 55.0 ( 0.0) | 55.0 ( 0.0) | |
| | Min, Max | ( 1.0, 55.0) | (45.0, 56.0) | ( 1.0, 56.0) | |
| Actual (days) | N | 23 | 20 | 43 | 0.217 |
| | Mean (SD) | 49.7 (13.61) | 53.7 ( 3.13) | 51.6 (10.26) | |
| | Median (IQR) | 55.0 ( 3.0) | 55.0 ( 1.0) | 55.0 ( 2.0) | |
| | Min, Max | ( 1.0, 55.0) | (45.0, 56.0) | ( 1.0, 56.0) | |

Duration is the number of days from first to last device use, plus 1. Actual days equals duration minus days not used.
Source: Listings 4, 5. 31DEC24, T08_comply

Table 9
Treatment-Emergent Adverse Events by System Organ Class and Preferred Term (Safety Population)

| MedDRA System Organ Class | MedDRA Preferred Term | Treatment Device Group Active (N= 23) | Treatment Device Group Sham (N= 20) | Treatment Device Group Total (N= 43) | P-Value |
|---|---|---|---|---|---|
| === Total Patients === | === Total Patients === | 15 ( 65.2) | 9 ( 45.0) | 24 ( 55.8) | 0.228 |
| Ear And Labyrinth Disorders | === Total Patients === | 1 ( 4.3) | 1 ( 5.0) | 2 ( 4.7) | 1.000 |
| | Ear Discomfort | 1 ( 4.3) | 0 ( 0.0) | 1 ( 2.3) | 1.000 |
| | Vertigo | 1 ( 4.3) | 1 ( 5.0) | 2 ( 4.7) | 1.000 |
| Gastrointestinal Disorders | === Total Patients === | 2 ( 8.7) | 1 ( 5.0) | 3 ( 7.0) | 1.000 |
| | Abdominal Pain | 1 ( 4.3) | 0 ( 0.0) | 1 ( 2.3) | 1.000 |
| | Emesis | 1 ( 4.3) | 0 ( 0.0) | 1 ( 2.3) | 1.000 |
| | Nausea | 0 ( 0.0) | 1 ( 5.0) | 1 ( 2.3) | 0.465 |
| General Disorders and Administrative | === Total Patients === | 2 ( 8.7) | 2 ( 10.0) | 4 ( 9.3) | 1.000 |
| | Fatigue | 1 ( 4.3) | 2 ( 10.0) | 3 ( 7.0) | 0.590 |
| | Malaise | 1 ( 4.3) | 0 ( 0.0) | 1 ( 2.3) | 1.000 |
| Infections And Infestations | === Total Patients === | 4 ( 17.4) | 4 ( 20.0) | 8 ( 18.6) | 1.000 |
| | Conjunctivitis | 1 ( 4.3) | 0 ( 0.0) | 1 ( 2.3) | 1.000 |
| | Covid-19 | 1 ( 4.3) | 0 ( 0.0) | 1 ( 2.3) | 1.000 |
| | Gastroenteritis | 0 ( 0.0) | 1 ( 5.0) | 1 ( 2.3) | 0.465 |
| | Pharyngitis | 0 ( 0.0) | 1 ( 5.0) | 1 ( 2.3) | 0.465 |
| | Post Viral Fatigue Syndrome | 0 ( 0.0) | 1 ( 5.0) | 1 ( 2.3) | 0.465 |
| | Rhinitis | 2 ( 8.7) | 0 ( 0.0) | 2 ( 4.7) | 0.491 |
| | Sinusitis | 2 ( 8.7) | 0 ( 0.0) | 2 ( 4.7) | 0.491 |
| | Upper Respiratory Tract Infectio | 1 ( 4.3) | 2 ( 10.0) | 3 ( 7.0) | 0.590 |
| | Urinary Tract Infection | 1 ( 4.3) | 0 ( 0.0) | 1 ( 2.3) | 1.000 |
| Injury, Poisoning And Procedural | === Total Patients === | 1 ( 4.3) | 0 ( 0.0) | 1 ( 2.3) | 1.000 |
| | Reaction To Previous Exposure To | 1 ( 4.3) | 0 ( 0.0) | 1 ( 2.3) | 1.000 |
| Musculoskeletal and Connective Tissue Disorders | === Total Patients === | 1 ( 4.3) | 1 ( 5.0) | 2 ( 4.7) | 1.000 |
| | Muscle Spasms | 1 ( 4.3) | 1 ( 5.0) | 2 ( 4.7) | 1.000 |
| Nervous System Disorders | === Total Patients === | 6 ( 26.1) | 3 ( 15.0) | 9 ( 20.9) | 0.467 |
| | Headache | 5 ( 21.7) | 2 ( 10.0) | 7 ( 16.3) | 0.420 |
| | Migraine | 1 ( 4.3) | 0 ( 0.0) | 1 ( 2.3) | 1.000 |
| | Presyncope | 0 ( 0.0) | 1 ( 5.0) | 1 ( 2.3) | 0.465 |
| Reproductive System and Breast Disorders | === Total Patients === | 0 ( 0.0) | 2 ( 10.0) | 2 ( 4.7) | 0.210 |
| | Endometriosis | 0 ( 0.0) | 1 ( 5.0) | 1 ( 2.3) | 0.465 |
| | Mastitis | 0 ( 0.0) | 1 ( 5.0) | 1 ( 2.3) | 0.465 |
| Respiratory, Thoracic and Mediastinal Disorders | === Total Patients === | 3 ( 13.0) | 4 ( 20.0) | 7 ( 16.3) | 0.687 |
| | Epistaxis | 1 ( 4.3) | 2 ( 10.0) | 3 ( 7.0) | 0.590 |
| | Nasal Discomfort | 1 ( 4.3) | 2 ( 10.0) | 3 ( 7.0) | 0.590 |
| | Nasal Mucosal Disorder | 1 ( 4.3) | 0 ( 0.0) | 1 ( 2.3) | 1.000 |
| Skin And Subcutaneous Tissue Dis | === Total Patients === | 5 ( 21.7) | 1 ( 5.0) | 6 ( 14.0) | 0.192 |

For summaries by preferred term (and SOC term), subjects with more than one AE are counted once. Source: Listings 6, 7.
P-value by Fisher's Exact Test. 31DEC24, T09_ae

Table 9
Treatment-Emergent Adverse Events by System Organ Class and Preferred Term (Safety Population)

| MedDRA | | Treatment Device Group | | | |
|---|---|---|---|---|---|
| System Organ Class | Preferred Term | Active (N= 23) | Sham (N= 20) | Total (N= 43) | P-Value |
| Skin And Subcutaneous Tissue Dis | Skin Abrasion | 1 ( 4.3) | 0 ( 0.0) | 1 ( 2.3) | 1.000 |
| | Skin Irritation | 4 ( 17.4) | 1 ( 5.0) | 5 ( 11.6) | 0.351 |

For summaries by preferred term (and SOC term), subjects with more than one AE are counted once. Source: Listings 6, 7.
P-value by Fisher's Exact Test. 31DEC24, T09_ae
